# Separating the effects of early and later life adiposity on colorectal cancer risk: a Mendelian randomization study

**DOI:** 10.1101/2022.07.01.22277131

**Authors:** Nikos Papadimitriou, Caroline J Bull, Mazda Jenab, David J. Hughes, Joshua A Bell, Eleanor Sanderson, Nicholas J Timpson, George Davey Smith, Demetrius Albanes, Peter T Campbell, Sébastien Küry, Loic Le Marchand, Cornelia M Ulrich, Kala Visvanathan, Jane C Figueiredo, Polly A Newcomb, Rish K Pai, Ulrike Peters, Kostas K Tsilidis, Jolanda M.A. Boer, Emma E Vincent, Daniela Mariosa, Marc J Gunter, Tom G Richardson, Neil Murphy

**Affiliations:** Nutrition and Metabolism Branch, International Agency for Research on Cancer, Lyon, France; MRC Integrative Epidemiology Unit (IEU), University of Bristol, Bristol, UK; Population Health Sciences, Bristol Medical School, University of Bristol, UK; School of Translational Health Sciences, University of Bristol, Bristol, UK; Cancer Biology and Therapeutics Group, UCD Conway Institute, School of Biomolecular and Biomedical Science, University College Dublin, Dublin, Ireland; Division of Cancer Epidemiology and Genetics, National Cancer Institute, National Institute of Health, Bethesda, Maryland, USA; Department of Epidemiology and Population Health, Albert Einstein College of Medicine, Bronx, New York, USA; Behavioural and Epidemiology Research Group, American Cancer Society, Atlanta, Georgia, USA; Service de Génétique Médicale, Centre Hospitalier Universitaire (CHU) Nantes, Nantes, France; University of Hawaii Cancer Center, Honolulu, Hawaii, USA; Huntsman Cancer Institute and Department of Population Health Sciences, University of Utah, Salt Lake City, Utah, USA; Department of Epidemiology, Johns Hopkins Bloomberg School of Public Health, Baltimore, Maryland, USA; Department of Medicine, Samuel Oschin Comprehensive Cancer Institute, Cedars-Sinai Medical Center, Los Angeles, CA, USA; Public Health Sciences Division, Fred Hutchinson Cancer Research Center, Seattle, Washington, USA; Department of Epidemiology, University of Washington, Seattle, Washington, USA; Department of Laboratory Medicine and Pathology, Mayo Clinic Arizona, Scottsdale, Arizona, USA; Department of Epidemiology and Biostatistics, School of Public Health, Imperial College London, London, UK; Department of Hygiene and Epidemiology, University of Ioannina School of Medicine, Ioannina, Greece; National Institute for Public Health and the Environment, The Netherlands; Section of Genomic Epidemiology, International Agency for Research on Cancer, Lyon, France; Novo Nordisk Research Centre, Headington, Oxford, UK

## Abstract

**Background:** Observational studies have linked childhood obesity with elevated risk of colorectal cancer, however, it is unclear if this association is causal or independent from the effects of obesity in adulthood on colorectal cancer risk.

**Methods:** We conducted Mendelian randomization (MR) analyses to investigate potential causal relationships between self-perceived body size in early life (age 10) and measured body mass index in adulthood (mean age 56.5) with risk of colorectal cancer. The total and independent effects of body size exposures were estimated using univariable and multivariable MR, respectively. Summary data were obtained from a genome-wide association study of 453,169 participants in UK Biobank for body size and from a genome-wide association study meta-analysis of three colorectal cancer consortia of 125,478 participants.

**Results:** Genetically predicted early life body size was estimated to increase odds of colorectal cancer (odds ratio [OR] per category change: 1.12, 95% confidence interval [CI]: 0.98-1.27), with stronger results for colon cancer (OR: 1.16, 95% CI: 1.00-1.35), and distal colon cancer (OR: 1.25, 95% CI: 1.04-1.51). After accounting for adult body size using multivariable MR, effect estimates for early life body size were attenuated towards the null for colorectal cancer (OR: 0.97, 95% CI: 0.77-1.22) and colon cancer (OR: 0.97, 95% CI: 0.76-1.25), while the estimate for distal colon cancer was of similar magnitude but more imprecise (OR: 1.27, 95% CI: 0.90-1.77).

**Conclusions:** Our findings suggest that the positive association between early life body size and colorectal cancer risk is likely due to large body size retainment into adulthood.

## Introduction

Childhood obesity is a major global public health challenge with increasing prevalence observed in most geographic regions over the past three decades (1). The rising prevalence of childhood obesity may have important consequences for population health, irrespective of adiposity later in life, given evidence from observational studies linking early life adiposity with elevated risks of chronic diseases, including cancer (1-4).

Two recent meta-analyses of observational studies have reported positive associations between early life body size measures (in adolescence and early adulthood) and later life colorectal cancer risk in both men and women (5, 6). Despite these associations, causal inference of the health effects of early life adiposity on later life disease risk can be challenging, as individuals who are obese in childhood often remain so during adulthood (7). Consequently, it is currently unknown if the prior positive associations between early life adiposity and colorectal cancer risk are a direct effect of early life obesity or to what extent they are mediated by later life adiposity.

Mendelian randomization (MR) uses germline genetic variants as proxies to allow causal inference between a given exposure and outcome (8). Compared to traditional observational analyses, MR analyses should be less susceptible to conventional confounding and reverse causation, given the randomly allocated and fixed nature of genetic variants (9). Additionally, multivariable MR allows the estimation of independent effects of multiple exposures (e.g., early life and adult body size) on disease outcomes (10-12). Univariable MR can be used to estimate the total effect of early life body size on colorectal cancer (Figure 1A). Whereas multivariable MR can be used to estimate the effect of childhood obesity specifically on later life chronic disease risk, independently of adult body size (13, 14). Under the multivariable framework, we hypothesize three scenarios in which early life body size affects colorectal cancer risk after considering adult body size (Figures 1B-D): Firstly, early life body size solely has direct effects on colorectal cancer, which are not influenced by adult body size (Figure 1B); secondly, early life body size has only indirect effects on colorectal cancer through adult body size (Figure 1C); thirdly, early life body size can have both direct and indirect effects on colorectal cancer risk (Figure 1D).

**Figure 1:**
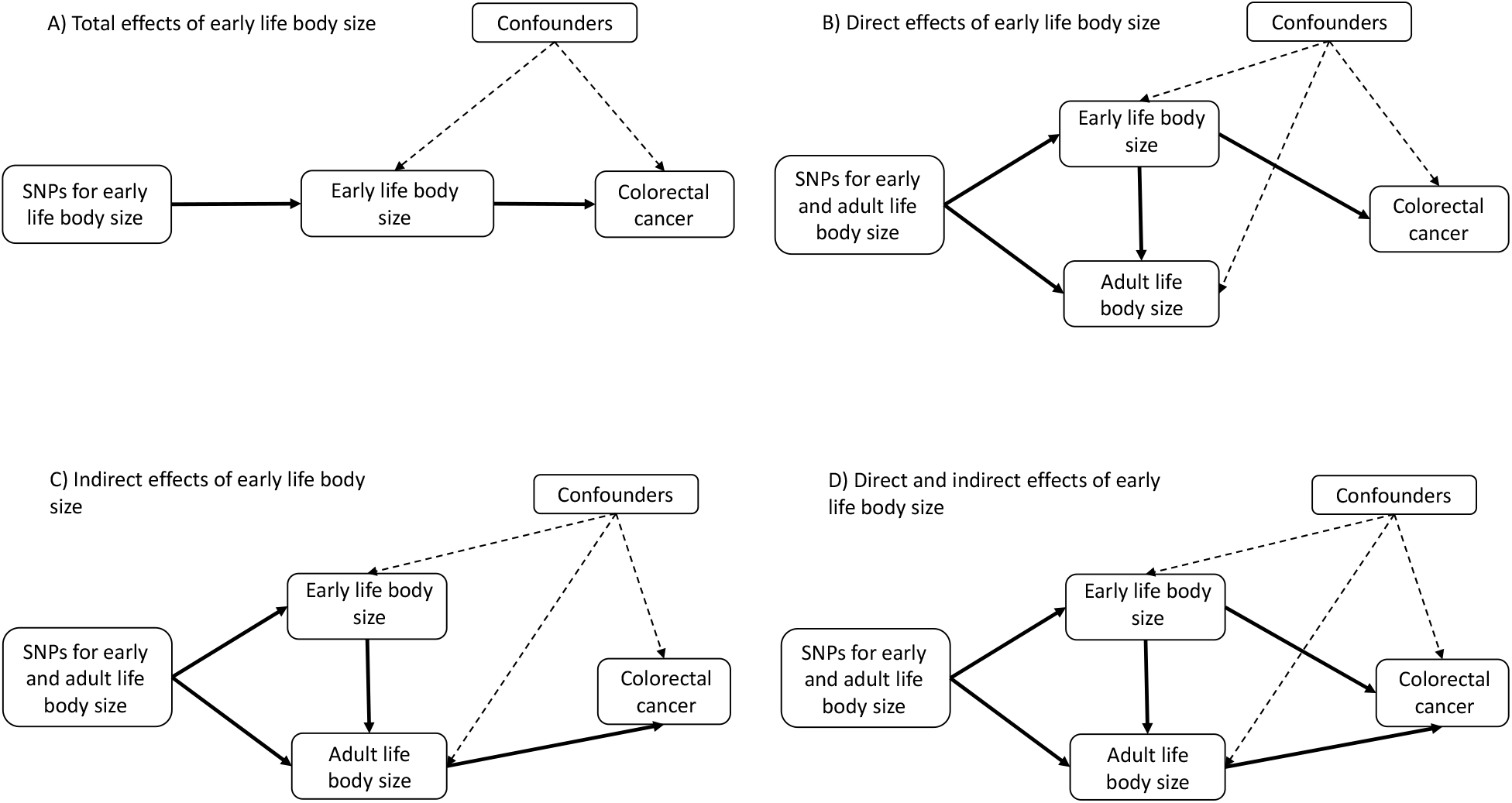
Directed acyclic graphs displaying four possible scenarios that could explain a causal effect between body size at age 10 years and colorectal cancer within the univariable and multivariable Mendelian randomisation analysis. (Top left) The total effect of early life body size on colorectal cancer risk, (top right) early life body size has a direct effect on colorectal cancer risk independent of adult body size, (bottom left) early life body size has an indirect effect on colorectal cancer risk only through adult body size, and (bottom right) early life body size has both direct and indirect effects on colorectal cancer risk.

We used a two-sample multivariable MR framework to examine potential causal associations between early life body size and colorectal cancer risk, independent of adult body size. We combined genetic variants associated with recalled early life and measured adult body size from a recent genome-wide association study (GWAS) of 453,169 adults (13), and then examined the association of these variants with colorectal cancer risk in a large consortium of up to 125,478 adults (58,131 cases and 67,347 controls) (15, 16).

## Methods

### Data on early life and adult body size

Genetic variants associated with early life and adult body size were identified from a recent GWAS of 453,169 participants of European descent from the UK Biobank (13). For early life body size, participants were asked at baseline: “*When you were 10 years old, compared to average would you describe yourself as thinner, plumper, or about average*?”. For adult body size, body mass index (BMI) was derived using height (centimeters) and weight (kilograms) measured at baseline. To improve comparability between the different body size traits, BMI in adults was converted into a categorical variable with three groups like early life body size. A linear regression model was applied, assuming similar effects of a given single nucleotide polymorphism (SNP) on moving from the lowest to the middle and from the middle to the highest category of the body size variables. The genome-wide significant (P<5×10^−8^) variants identified in this GWAS were pruned based on a linkage disequilibrium (LD) level of R^2^<0.001 using genotype data from European individuals from phase 3 (version 5) enrolled in the 1000 genomes project as a reference panel (13). The resulting instruments for early life body size (305 SNPs) and adult body size (557 SNPs) explained 4.5% and 6.4% of variability in these traits, respectively. Early life and adult body size genetic instruments were comprised of 138 and 215 SNPs for women and 68 and 159 SNPs for men, respectively (Supplementary Table 1). These genetic instruments have been used in prior MR studies and validated in external cohorts (13, 14, 17, 18). The genetic correlation between early life and adult body size in UK Biobank was found to be 0.61(13).

### Data on colorectal cancer

Summary data for the associations of the early life and adult body size related genetic variants with colorectal cancer (overall and by site: colon, proximal colon, distal colon, rectum) were obtained from a GWAS of 125,478 adults (58,131 cancer cases and 67,347 controls) within the ColoRectal Transdisciplinary Study (CORECT), the Colon Cancer Family Registry (CCFR), and the Genetics and Epidemiology of Colorectal Cancer (GECCO) consortium (15).

Imputation was performed using the Haplotype Reference Consortium (HRC) r1.0 reference panel and regression models were further adjusted for age, sex, genotyping platform and genomic principal components as detailed here (15). Subsite specific estimates by sex were obtained from a GWAS of 112,373 adults (48,214 cancer cases and 64,159 controls) within the same consortia (16). Colorectal cancer estimates for each SNP are presented in Supplementary Table 2. Summary statistics were calculated after excluding participants from UK Biobank to avoid potential overlap between the two datasets. Supplementary Table 3 presents the final number of cancer cases by sex and subsite. Ethics were approved by respective institutional review boards.

### Statistical analysis

#### Power calculations

The a priori statistical power was calculated using an online tool at http://cnsgenomics.com/shiny/mRnd/ (19). For early life body size, given a type 1 error of 5%, there was sufficient power (> 80%) to detect an OR ≥1.10 for overall colorectal cancer per increase in odds conferred for each category change for both sexes combined while, an OR ≥1.15 was needed for the cancer sub-site analyses. In sex specific analyses, there was 80% power to detect an OR ≥1.21 and 1.16 or men and women, respectively. For adult body size, given a type 1 error of 5%, there was > 80% power to detect an OR ≥ 1.09 for overall colorectal cancer increase in odds conferred for each category change and an OR ≥ 1.17 in sex specific analyses. Supplementary Table 4 presents in more detail the statistical power for the two exposures and all outcomes in sex combined and specific analyses.

#### Univariable MR analysis to estimate the total effect of early and adult body size on CRC

A two-sample MR approach using summary data and the fixed-effect inverse-variance weighted method was implemented. Where Cochran’s Q statistics identified heterogeneity across the individual SNPs in the early life and adult body size instruments, random-effect inverse-variance weighted analyses were conducted (20, 21). Univariable MR analyses for men and women combined were conducted to estimate the effect of both early life and adult body size independently on colorectal cancer risk. Analyses according to sex and tumor anatomical subsite were also conducted. Heterogeneity of associations according to sex and colorectal anatomical subsites was assessed by calculating the *χ*^2^ statistic (22). The strength of each instrument was measured by calculating the F-statistic using the following formula: *F* =*R*^2^ ∗ (*N* − 2)⁄(1 −*R*^2^), where R^2^ is the proportion of the variability of the phenotype explained by each instrument and N the sample size of the GWAS for the exposures, with values below 10 denoting the presence of weak instrument bias (23, 24).

#### Multivariable MR analysis to estimate the independent (direct) effects of early and adult body size on CRC

Due to the correlation between genetic determinants of early life and adult body size, any difference between the total and independent (direct) effects of later life body size is likely driven by pleiotropy if there is also a direct effect of early body size on colorectal cancer (13). Consequently, the focus of our analyses was on the total effect of early body size and the direct effects of early and adult body size. We therefore conducted multivariable MR analyses to estimate the direct effects of early and later life body size on colorectal cancer risk. For multivariable MR, we calculated three quantities: the conditional *F*_*early life body size*_, *F*_*adult body size*_ and Q_a_. *F*_*early life body size*_ and *F*_*adult body size*_ to examine the variance explained by the genetic variants on the main (i.e., early life body size) and secondary exposures (i.e., adult body size); again values over 10 were interpreted to suggest little evidence of weak instrument bias (11). Q_a_ is a generalisation of the Q statistic for the multivariable scenario, where high values based again on a 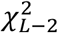 distribution denote heterogeneity and potential pleiotropy even when corrected for adult body size (11).

#### Sensitivity analyses

Sensitivity analyses were used to check and correct for the presence of pleiotropy in the estimates. To evaluate the extent to which directional pleiotropy may have affected the causal estimates for the early and adult body size and colorectal cancer association, we used MR-Egger regression (25, 26). We also computed OR estimates using the complementary weighted-median method which can give valid MR estimates under the presence of horizontal pleiotropy when up to 50% of the included instruments are invalid (27).

Odds ratio (OR) from MR analyses reflect the increase in odds conferred for each category change (i.e. thinner to average and average to plumper) in the early and adult life body size phenotypes. All analyses were undertaken using R (version 3.6.3) using the MendelianRandomisation package (28, 29). LD clumping between early life and adult body size SNPs in the multivariable MR analyses was done using the ieugwasr R package (based on linkage disequilibrium R^2^ = 0.001) and the plots were created using the forestplot R package (30, 31). The list of SNPs included in the multivariable MR analyses is given in Supplementary Table 5.

## Results

### Early life body size

Genetically-predicted early life body size was estimated to increase risk of colorectal cancer (OR per category change: 1.12, 95% confidence interval [CI]: 0.98-1.27), colon cancer (OR: 1.16, 95% CI: 1.00-1.35), and distal colon cancer (OR: 1.25, 95% CI: 1.04-1.51). While proximal colon and rectal cancer were minimally influenced by early life body size (OR: 1.11, 95% CI: 0.93, 1.32 and OR: 1.14 95% CI: 0.93, 1.38, respectively) (Figure 2, Supplementary Table 7). In the multivariable models, the direct effect estimates of early life body size were attenuated to null for colorectal (OR: 0.97, 95% CI: 0.77-1.22) and colon cancer (OR: 0.97, 95% CI: 0.76-1.25), while an estimate of similar magnitude with more imprecision was observed for distal colon cancer (OR: 1.27, 95% CI: 0.90-1.77) (Figure 2). In multivariable models for proximal colon and rectal cancer, there was little evidence for a direct effect of genetically predicted early life body size (OR: 0.82, 95% CI: 0.61, 1.09 and OR: 1.05 95% CI: 0.76, 1.45, respectively).

**Figure 2:**
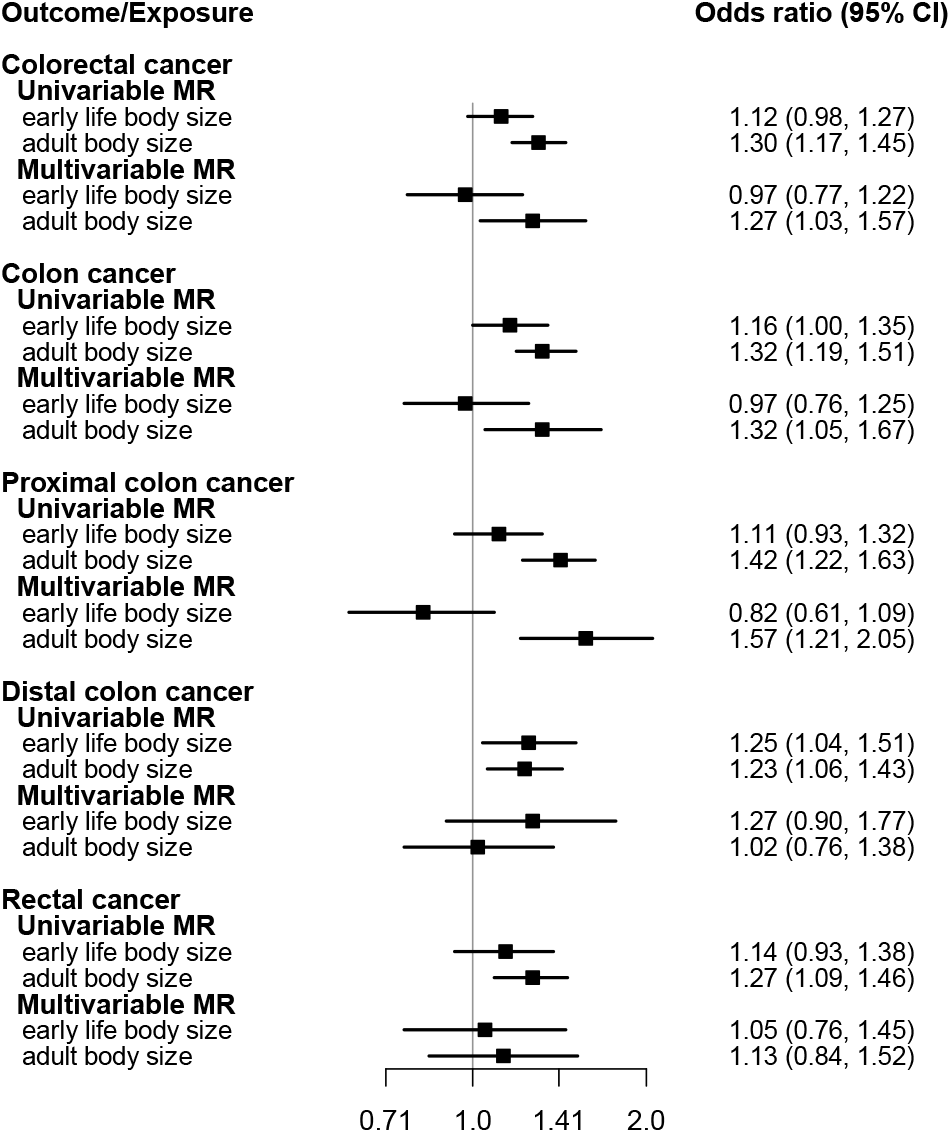
Forest plot showing the estimated direct and indirect effects for genetically predicted early (age 10 years) and adult body size in on colorectal cancer both overall and by cancer sub-site. The error bars correspond to the odds ratios with 95% confidence intervals and those within the grey frameworks represent the results of the univariable Mendelian randomisation (MR) analysis.

For women, early life body size was estimated to increase colorectal cancer (OR: 1.20, 95% CI: 0.97-1.48) and colon cancer (OR 1.20, 95% CI: 0.95-1.51) (Figure 3, Supplementary Table 7). These genetically predicted effects attenuated towards the null in the multivariable model (colorectal cancer, OR: 1.08, 95% CI: 0.81-1.45; colon cancer, OR: 0.99, 95% CI: 0.71-1.36) with similar patterns for the other subsite models. For men, little evidence for an effect of early life body size on colorectal cancer risk was observed in the univariable model (OR: 0.96, 95% CI: 0.73-1.26) with an inverse point estimate observed in the multivariable model (OR: 0.74, 95% CI: 0.49-1.12) (Figure 3).

**Figure 3:**
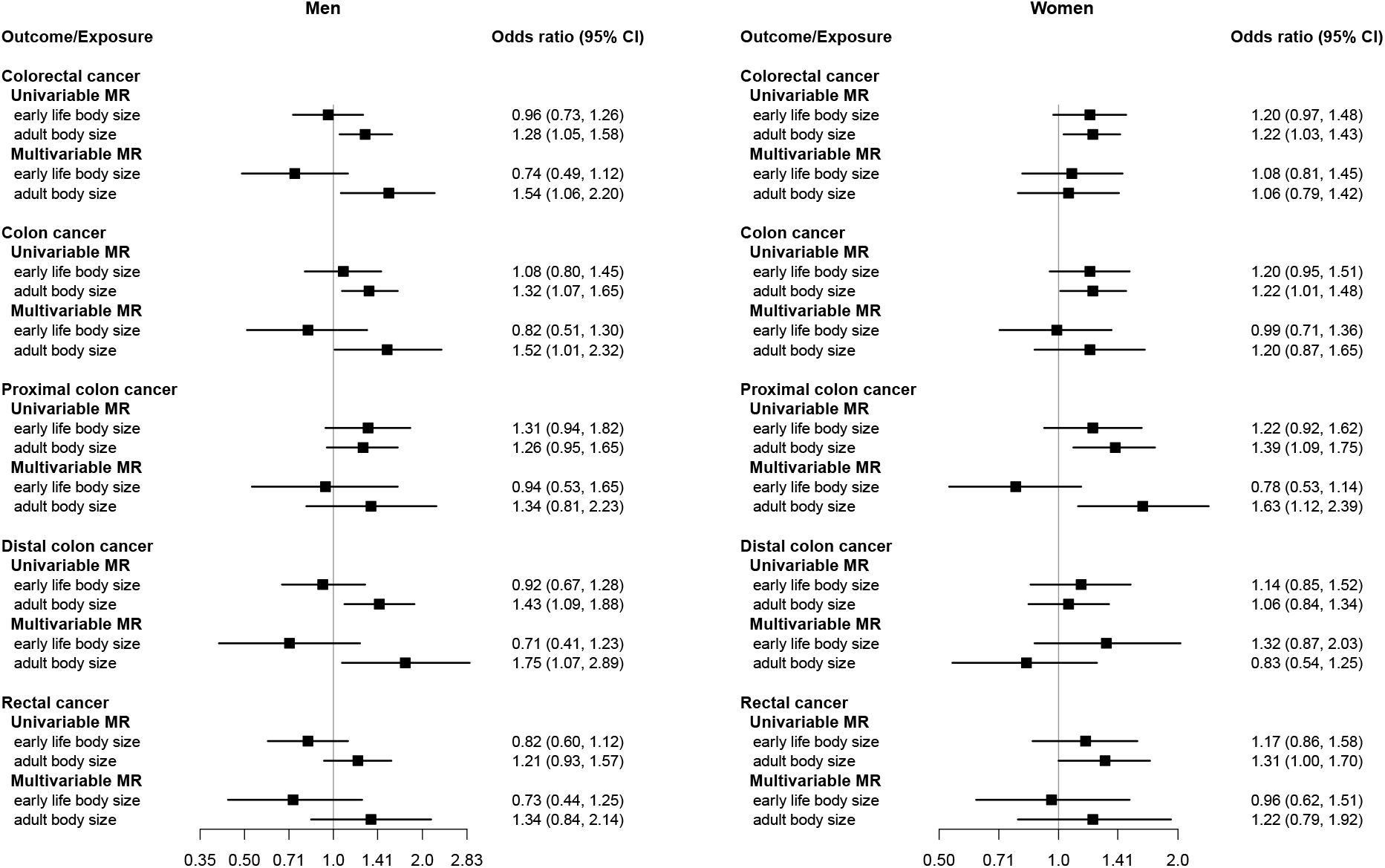
Forest plot showing the estimated direct and indirect effects for genetically predicted early (age 10 years) and adult body size in on colorectal cancer both overall and by cancer sub-site in men and women separately. The error bars correspond to the odds ratios with 95% confidence intervals and those within the grey frameworks represent the results of the univariable Mendelian randomisation (MR) analysis.

The effect estimates were similar across cancer subsite (P-heterogeneity≥ 0.14) and by sex (P-heterogeneity≥ 0.10) in both the univariable and multivariable analyses. For distal colon cancer opposing direct effect estimates of early life body size with wide confidence intervals were observed for men (OR: 0.71, 95% CI: 0.41, 1.23) and women (OR: 1.32, 95% CI: 0.87, 2.03); P-heterogeneity = 0.08.

### Adult life body size

In the sex-combined multivariable model, adult body size was estimated to directly increase the risk of colorectal cancer (OR per category change: 1.27, 95% CI: 1.03, 1.57), colon cancer (OR: 1.32, 95% CI: 1.05, 1.67), and proximal colon cancer (OR: 1.57, 95% CI: 1.21, 2.05) whereas estimates for distal colon (OR: 1.02, 95% CI: 0.76, 1.38) and rectal (OR: 1.13, 95% CI: 0.84, 1.52) cancers were of smaller magnitude (Figure 2).

For women, adult body size was estimated to directly increase, albeit imprecisely, the risk of colon (OR: 1.20, 95% CI: 0.87, 1.65), proximal colon (OR: 1.63, 95% CI: 1.12, 2.39), and rectal cancer (OR: 1.22, 95% CI: 0.79, 1.92); whereas null estimates were observed for colorectal cancer (OR: 1.06, 95% CI: 0.79, 1.42) (Figure 3). For men, adult body size was estimated to directly increase the risk of colorectal cancer (OR: 1.54, 95% CI: 1.06, 2.20), colon cancer (OR: 1.52, 95% CI: 1.01, 2.32), and distal colon cancer (OR: 1.75, 95% CI: 1.07, 2.89) with similar positive effect estimates found for proximal colon (OR: 1.34, 95% CI: 0.81, 2.23) and rectal cancer (OR: 1.34, 95% CI: 0.84, 2.14) (Figure 3).

Similarly to the early life body size analysis, opposing multivariable analysis estimates were observed for the direct effect of adult body size on distal colon cancer in men and women (OR: 1.75, 95% CI: 1.07, 2.89 and OR: 0.83, 95% CI: 0.54, 1.25, respectively, P-heterogeneity=0.02).

### Sensitivity analyses

Regarding the MR assumptions, we found little evidence of weak instrument bias under the univariable framework (F statistics were > 10 for all SNPs included in the analysis). In multivariable MR *F*_*early life body size*_, *F*_*adult body size*_ statistics were all over 10 with the exception of the analysis in men where *F*_*early life body size*_ was equal to 8, and *F*_*adult body size*_ was equal to 10 suggesting that weak instrument bias may influence the analyses for men (Supplementary Tables 1, 7). There was also evidence of heterogeneity in most of the analyses as denoted by the Q statistics (Supplementary Tables 6, 7). Under the univariable framework, based on Egger’s intercept test, evidence of directional pleiotropy was found for the effect of early life body size on colorectal cancer (overall, women only) and proximal colon cancer (overall), with stronger positive effect estimates observed for the MR Egger regression models (overall colorectal cancer, OR 1.46, 95% CI: 1.09, 1.93; colorectal cancer women only, OR 1.84, 95% CI: 1.16, 2.92; and overall proximal colon cancer, OR 1.60, 95% CI: 1.09, 2.34) (Supplementary Table 7). Finally, the multivariable MR Egger’s intercept test identified some evidence of pleiotropy in the analyses of adult body size on colorectal, colon and proximal colon risk, but the related effect estimates replicated the positive direct effects of adult body size observed in the main analysis (Supplementary Table 8).

## Discussion

In the current study we investigated the effect of early life body size (at age 10 years) on risk of later life colorectal cancer, and whether this effect remained robust after accounting for adult body size. Mendelian randomization estimates of the total effect of early life body size on colorectal cancer risk revealed positive genetically predicted effects of early life body size and colorectal cancer risk which were strongest for colon and distal colon cancer. However, these associations were generally attenuated following adjustment for adult body size, suggesting no direct effects of early life body size on colorectal cancer risk. One exception was distal colon cancer, where the point estimate for the effect of early life body size on cancer risk remained in the multivariable scenario, though confidence intervals spanned the null. A similar pattern of results was reported in the sex-specific analyses, except for opposing effect estimates for both early life and adult body size on distal colon cancer under the multivariable framework, where greater early life body size appeared protective against distal colon cancer in male and detrimental in females and later life body size protective in females and detrimental in males, though both sets of estimates spanned the null.

Relatively few observational studies have examined the relationship between early life body size and risk of later life colorectal cancer. A joint Nurses’ Health Study (NHS) and Health Professionals Follow-up Study (HPFS) found that higher body fatness in early life (childhood and adolescence) was associated with greater colorectal cancer risk for women, but not men with similar results across cancer sub-sites (32); with these associations largely unchanged after multivariable adjustment for adult BMI (32). A Swedish study of more than 200,000 men who had their height and weight measured in adolescence, reported a more than twofold higher risk for later life colorectal cancer for the obese group (BMI ≥30 kg/m^2^) when compared with the normal weight group (BMI 18.5-<25 kg/m^2^) (33). A large Israeli study of almost 1.8 million men and women showed that being overweight and obese at adolescence was associated with higher colon cancer risk for both men (HR for overweight, 1.53; 95% CI, 1.28-1.84; HR for obesity, 1.54; 95% CI, 1.15-2.06; statistically significant from a BMI of 23.4 kg/m^2^) and women (HR for overweight, 1.54; 95% CI, 1.22-1.93; HR for obesity, 1.51; 95% CI, 0.89-2.57; significant from a BMI of 23.6 kg/m^2^) (34). However, the Swedish and Israeli studies lacked data on information on important potential confounders including adult body size.

A recent MR study examined the associations between childhood obesity and cancer risk without taking into account adult body size, using a genetic instrument of 15 SNPs from a GWAS of 47,541 children from the Early Growth Genetics (EGG) consortium (35). Effect estimates from this study did not support a positive relationship between childhood BMI and overall colorectal cancer (OR, 1.11; 95% CI, 0.93-1.32) (35). Results from our analyses, that crucially adjusted for adult body size, also reflect little evidence of a positive relationship between early life body size and later life colorectal cancer risk. An additional MR study following a similar approach but using a different dataset for colorectal cancer also did not find any positive relationship between childhood BMI and overall colorectal cancer after adjusting for adult body size (18). However, this study did not investigate further associations stratified by sex or cancer sub-site. Our findings suggest that positive effect estimates found between early life body size and colorectal cancer may be attributable to participants who were overweight or obese in childhood remaining this weight during adulthood, rather than any distinct harm from childhood adiposity itself. There was weak evidence for a direct effect of early life body size on distal colon cancer risk which warrants further investigation in new and larger studies with measured early life body size.

We observed divergent effect estimates for early life body size and distal colon cancer risk for men and women in the multivariable models (P-heterogeneity = 0.08). However, caution is needed when interpreting the results for men as the estimates for early life body size might suffer from weak instrument bias based on the low conditional F statistic.

Strengths of the current study include our use of novel multivariable MR methods to disentangle the effects of early life body size from adult body size on colorectal cancer risk. Importantly, both early life and adult body size data were sourced from the identical participants within UK Biobank belonging to the same generation of individuals contributing data to the colorectal cancer GWAS used in this analysis, this should reduce bias that can be introduced to MR estimates when two samples do not represent the same underlying population (13). We obtained summary stats for CRC from the Huyghe et al. GWAS excluding UK biobank participants, thus independent datasets for the exposure and outcome phenotypes were used, avoiding potential bias due to sample overlap (36). Finally, the large cancer sample size allowed us to conduct a more detailed analysis by sex and cancer sub-site.

A limitation of our study is that the early life body size phenotype was based on a self-reported questionnaire data than on direct measurements. Therefore, we cannot exclude potential recall bias in the measurements. However, the early life body size SNPs were successfully validated in two independent studies with direct measures of childhood BMI, demonstrating their validity as instruments for early life body size (13, 14, 17). As exposures investigated were categorical variables with three broad categories this may limit power to detect small effects, this did not appear to limit site and sex combined analyses, but power was lower in analyses according to sex and subsite, as evidenced by the imprecision of these effect estimates. Moreover, analysing two correlated phenotypes together, like early and adult body size may have introduced collinearity which leads to greater imprecision and possible bias in the multivariable MR effect estimates especially for early life body size, however the same set of SNPs were strong enough to show independent effects of early life body size on breast cancer (13). Lastly, given the large number of comparisons, we cannot exclude that some of the observed effects might be due to chance

In conclusion, we found that the suggested effects of early life body size on later life colorectal cancer risk are likely a consequence of individuals who are overweight or obese during childhood remaining this way into adulthood. Our results suggest that being overweight in childhood may not be detrimental in terms of colorectal carcinogenesis providing individuals reach and then maintain normal weight during adulthood. Further research is required to examine any possible role of early life body size on distal colon cancer development.

## Data Availability

The summary statistics used in this study are contained in the supplementary materials. The summary-level GWAS data on outcomes used in this study are available following an application to the Genetics and Epidemiology of Colorectal Cancer Consortium (GECCO).

https://www.fredhutch.org/en/research/divisions/public-health-sciences-division/research/cancer-prevention/genetics-epidemiology-colorectal-cancer-consortium-gecco.html

