## Supplementary Tables for "Separating the effects of early and later life adiposity on colorectal cancer risk: a Mendelian randomization study"

**Legends of supplementary tables**

**Supplementary Table 1**: Beta estimates for genome-wide significant SNPs for early/adult life body size in UK Biobank (overall, men, women)

**Supplementary Table 2**: Summary information on colorectal cancer risk for the SNPs used in the analysis

**Supplementary Table 3**: Number of cancer cases by sex and subsite

**Supplementary Table 4**: Sample size and power calculations for each phenotype and group in the Mendelian randomization study of early and adult life body size and risk of colorectal cancer

**Supplementary Table 5**: Beta estimates for early/adult life body size in UK Biobank for the SNPs included in the MVMR analysis (overall, men, women)

**Supplementary Table 6**: $F_{early\ life\ body\ size}$, $F_{adult\ body\ size}$ and $Q_a$ in the multivariable MR between early, adult body size and colorectal cancer

**Supplementary Table 7**: Univariable Mendelian randomization estimates between early and adult body size and colorectal cancer risk

**Supplementary Table 8**: Multivariable MR Egger analysis to assess the effect of both predicted early life and adult body size on colorectal cancer

| **Supplementary table 1: Beta estimates for genome-wide significant SNPs for early/adult life body size in UK Biobank (overall, men, women)** | | | | | | | | | | | |
|---|---|---|---|---|---|---|---|---|---|---|---|
| Genome-wide significant SNPs for early life body size in UK Biobank (overall) | | | | | | | | | | | |
| SNP | Chr | Position | Closest gene | EA | OA | EAF | Beta | SE | pvalue | $R^2$ | F statistic |
| rs2229330 | 1 | 6649228 | ZBTB48 | T | G | 0.93 | -0.020 | 0.003 | 2.20E-13 | 1.19E-04 | 54 |
| rs2175171 | 1 | 7028842 | CAMTA1 | G | C | 0.44 | -0.008 | 0.001 | 3.90E-08 | 6.66E-05 | 30 |
| rs6577497 | 1 | 8605667 | RERE | A | T | 0.61 | 0.008 | 0.001 | 1.90E-08 | 6.97E-05 | 32 |
| rs12045879 | 1 | 15817090 | CELA2B | C | T | 0.69 | 0.010 | 0.002 | 5.30E-11 | 9.51E-05 | 43 |
| rs212517 | 1 | 21577159 | ECE1 | T | A | 0.40 | 0.009 | 0.001 | 8.90E-10 | 8.29E-05 | 38 |
| rs2356864 | 1 | 50839740 | DMRTA2 | G | A | 0.48 | -0.009 | 0.001 | 8.00E-10 | 8.33E-05 | 38 |
| rs630602 | 1 | 54728864 | SSBP3 | G | C | 0.39 | -0.010 | 0.001 | 1.60E-12 | 1.10E-04 | 50 |
| rs12140153 | 1 | 62579891 | INADL | G | T | 0.91 | 0.022 | 0.002 | 6.20E-19 | 1.74E-04 | 79 |
| rs2767486 | 1 | 65991203 | LEPR | A | G | 0.80 | -0.015 | 0.002 | 1.10E-18 | 1.72E-04 | 78 |
| rs7522014 | 1 | 66551759 | PDE4B | A | G | 0.67 | 0.009 | 0.002 | 8.00E-10 | 8.33E-05 | 38 |
| rs2755253 | 1 | 67470843 | SLC35D1 | C | T | 0.29 | 0.009 | 0.002 | 1.60E-08 | 7.06E-05 | 32 |
| rs11209943 | 1 | 72750500 | NEGR1 | A | G | 0.40 | -0.018 | 0.001 | 9.70E-38 | 3.64E-04 | 165 |
| rs12042908 | 1 | 74997762 | TNNI3K | A | G | 0.44 | 0.027 | 0.001 | 2.30E-84 | 8.35E-04 | 379 |
| rs34517439 | 1 | 78450517 | DNAJB4 | C | A | 0.88 | -0.014 | 0.002 | 2.20E-10 | 8.89E-05 | 40 |
| rs11165687 | 1 | 97099767 | PTBP2 | C | T | 0.59 | -0.008 | 0.001 | 9.40E-09 | 7.27E-05 | 33 |
| rs7550711 | 1 | 110082886 | GPR61 | C | T | 0.97 | -0.048 | 0.004 | 4.40E-28 | 2.66E-04 | 121 |
| rs3013431 | 1 | 113242488 | MOV10 | C | T | 0.39 | 0.009 | 0.001 | 1.40E-09 | 8.10E-05 | 37 |
| rs12132598 | 1 | 115235785 | AMPD1 | A | G | 0.67 | -0.008 | 0.001 | 2.10E-08 | 6.94E-05 | 31 |
| rs7536458 | 1 | 118864602 | SPAG17 | T | G | 0.74 | -0.010 | 0.002 | 1.80E-10 | 8.98E-05 | 41 |
| rs11205303 | 1 | 149906413 | MTMR11 | T | C | 0.59 | 0.010 | 0.001 | 1.70E-11 | 1.00E-04 | 45 |
| rs35588936 | 1 | 155950963 | ARHGEF2 | C | T | 0.94 | 0.018 | 0.003 | 3.00E-10 | 8.76E-05 | 40 |
| rs12748436 | 1 | 177761109 | SEC16B | C | G | 0.92 | -0.016 | 0.003 | 3.50E-09 | 7.69E-05 | 35 |
| rs543874 | 1 | 177889480 | SEC16B | A | G | 0.79 | -0.047 | 0.002 | 8.80E-163 | 1.63E-03 | 739 |
| rs78444298 | 1 | 184672098 | EDEM3 | G | A | 0.98 | 0.042 | 0.005 | 1.70E-16 | 1.50E-04 | 68 |
| rs4074404 | 1 | 187683956 | PLA2G4A | T | A | 0.84 | -0.013 | 0.002 | 4.30E-11 | 9.59E-05 | 43 |
| rs16839832 | 1 | 196349909 | KCNT2 | G | T | 0.92 | -0.014 | 0.003 | 3.30E-08 | 6.74E-05 | 31 |
| rs9438393 | 1 | 205782718 | SLC41A1 | A | G | 0.59 | 0.010 | 0.001 | 3.10E-13 | 1.17E-04 | 53 |
| rs7354849 | 1 | 232765308 | SIPA1L2 | A | G | 0.54 | -0.008 | 0.001 | 7.40E-09 | 7.38E-05 | 33 |
| rs62106258 | 2 | 417167 | FAM150B | T | C | 0.95 | 0.080 | 0.003 | 2.40E-134 | 1.34E-03 | 609 |
| rs12992672 | 2 | 632592 | TMEM18 | G | A | 0.17 | -0.043 | 0.002 | 5.50E-121 | 1.21E-03 | 547 |
| rs2867116 | 2 | 682363 | TMEM18 | C | A | 0.86 | -0.012 | 0.002 | 1.40E-09 | 8.09E-05 | 37 |
| rs2141004 | 2 | 6194359 | DKFZP761K2322 | A | C | 0.74 | 0.010 | 0.002 | 2.60E-10 | 8.82E-05 | 40 |
| rs10182458 | 2 | 25150641 | ADCY3 | A | G | 0.51 | -0.036 | 0.001 | 3.10E-145 | 1.45E-03 | 659 |
| rs146910503 | 2 | 25446473 | DNMT3A | G | A | 0.98 | 0.033 | 0.005 | 2.70E-11 | 9.79E-05 | 44 |
| rs6719507 | 2 | 29733801 | ALK | G | A | 0.56 | 0.008 | 0.001 | 2.90E-08 | 6.79E-05 | 31 |
| rs62134189 | 2 | 45046339 | CAMKMT | A | G | 0.90 | 0.014 | 0.002 | 1.00E-09 | 8.22E-05 | 37 |
| rs2902142 | 2 | 58849805 | FANCL | C | T | 0.63 | 0.009 | 0.001 | 1.80E-09 | 7.99E-05 | 36 |
| rs2539692 | 2 | 59841988 | BCL11A | T | A | 0.39 | 0.008 | 0.001 | 1.80E-08 | 7.01E-05 | 32 |
| rs1177279 | 2 | 61295122 | KIAA1841 | A | G | 0.28 | 0.009 | 0.002 | 1.40E-08 | 7.10E-05 | 32 |
| rs7565437 | 2 | 65646966 | SPRED2 | T | C | 0.59 | 0.008 | 0.001 | 9.50E-09 | 7.27E-05 | 33 |

| | | | | | | | | | | | |
|---|---|---|---|---|---|---|---|---|---|---|---|
| rs12713889 | 2 | 77225361 | LRRTM4 | T | C | 0.67 | 0.012 | 0.001 | 3.70E-15 | 1.36E-04 | 62 |
| rs772175 | 2 | 96944553 | SNRNP200 | G | A | 0.66 | 0.008 | 0.001 | 5.00E-08 | 6.56E-05 | 30 |
| rs1384660 | 2 | 142299735 | LRP1B | G | A | 0.81 | 0.016 | 0.002 | 1.90E-18 | 1.69E-04 | 77 |
| rs62175963 | 2 | 161339964 | RBMS1 | T | C | 0.58 | 0.009 | 0.001 | 2.20E-10 | 8.88E-05 | 40 |
| rs115319174 | 2 | 207066474 | GPR1 | G | C | 0.94 | -0.042 | 0.003 | 4.00E-44 | 4.28E-04 | 194 |
| rs11891707 | 2 | 207120604 | ZDBF2 | T | C | 0.86 | 0.012 | 0.002 | 3.50E-09 | 7.70E-05 | 35 |
| rs3791478 | 2 | 240064139 | HDAC4 | T | C | 0.89 | 0.012 | 0.002 | 3.50E-08 | 6.71E-05 | 30 |
| rs1476698 | 2 | 242296449 | FARP2 | A | G | 0.63 | 0.008 | 0.001 | 1.50E-08 | 7.08E-05 | 32 |
| rs2594994 | 3 | 11339960 | ATG7 | T | A | 0.18 | 0.015 | 0.002 | 2.50E-17 | 1.58E-04 | 72 |
| rs7619139 | 3 | 25110415 | RARB | T | A | 0.41 | -0.010 | 0.001 | 6.20E-12 | 1.04E-04 | 47 |
| rs1402989 | 3 | 27056851 | NEK10 | C | T | 0.51 | -0.008 | 0.001 | 2.60E-08 | 6.84E-05 | 31 |
| rs2268762 | 3 | 38516075 | ACVR2B | A | G | 0.39 | -0.008 | 0.001 | 2.50E-08 | 6.85E-05 | 31 |
| rs754635 | 3 | 42305131 | CCK | C | G | 0.11 | -0.016 | 0.002 | 5.50E-13 | 1.15E-04 | 52 |
| rs2034963 | 3 | 48170802 | CDC25A | G | C | 0.35 | 0.011 | 0.001 | 6.50E-14 | 1.24E-04 | 56 |
| rs35926495 | 3 | 50255663 | GNAI2 | C | T | 0.62 | -0.009 | 0.001 | 4.80E-10 | 8.55E-05 | 39 |
| rs3774604 | 3 | 53824136 | CACNA1D | C | T | 0.63 | 0.008 | 0.001 | 4.60E-08 | 6.59E-05 | 30 |
| rs2629881 | 3 | 59778271 | FHIT | C | T | 0.22 | -0.011 | 0.002 | 3.50E-10 | 8.69E-05 | 39 |
| rs538579 | 3 | 62711674 | CADPS | G | C | 0.69 | -0.008 | 0.002 | 2.50E-08 | 6.86E-05 | 31 |
| rs115903965 | 3 | 66009529 | MAGI1 | G | A | 0.98 | -0.026 | 0.005 | 1.60E-08 | 7.04E-05 | 32 |
| rs4677156 | 3 | 72417857 | RYBP | A | T | 0.23 | 0.009 | 0.002 | 3.70E-08 | 6.68E-05 | 30 |
| rs1666132 | 3 | 77649190 | ROBO2 | C | T | 0.42 | 0.008 | 0.001 | 6.90E-09 | 7.41E-05 | 34 |
| rs1357798 | 3 | 83780583 | CADM2 | C | T | 0.21 | -0.009 | 0.002 | 4.20E-08 | 6.63E-05 | 30 |
| rs6783281 | 3 | 84791851 | CADM2 | A | G | 0.71 | -0.010 | 0.002 | 5.70E-10 | 8.47E-05 | 38 |
| rs7355953 | 3 | 85792137 | CADM2 | T | C | 0.78 | -0.015 | 0.002 | 6.60E-19 | 1.74E-04 | 79 |
| rs2735556 | 3 | 88105360 | CGGBP1 | T | C | 0.89 | 0.017 | 0.002 | 4.80E-15 | 1.35E-04 | 61 |
| rs11925138 | 3 | 131626048 | CPNE4 | G | A | 0.90 | 0.013 | 0.002 | 3.10E-08 | 6.76E-05 | 31 |
| rs7625768 | 3 | 131774642 | CPNE4 | G | A | 0.68 | -0.010 | 0.002 | 1.30E-11 | 1.01E-04 | 46 |
| rs1199333 | 3 | 138091701 | MRAS | G | T | 0.18 | 0.014 | 0.002 | 6.10E-14 | 1.24E-04 | 56 |
| rs59714050 | 3 | 141267294 | RASA2 | T | A | 0.93 | -0.023 | 0.003 | 5.20E-16 | 1.45E-04 | 66 |
| rs355748 | 3 | 153964496 | ARHGEF26 | G | T | 0.62 | -0.010 | 0.001 | 7.30E-12 | 1.04E-04 | 47 |
| rs7633995 | 3 | 180369750 | CCDC39 | A | G | 0.90 | -0.014 | 0.002 | 4.70E-09 | 7.57E-05 | 34 |
| rs10937241 | 3 | 185822774 | ETV5 | A | G | 0.16 | -0.011 | 0.002 | 1.10E-08 | 7.21E-05 | 33 |
| rs34811474 | 4 | 25408838 | ANAPC4 | G | A | 0.77 | 0.010 | 0.002 | 8.80E-10 | 8.29E-05 | 38 |
| rs7656673 | 4 | 30840331 | PCDH7 | A | G | 0.60 | -0.012 | 0.001 | 6.80E-16 | 1.44E-04 | 65 |
| rs34722008 | 4 | 38659594 | AC021860.1 | G | A | 0.65 | 0.008 | 0.001 | 1.60E-08 | 7.04E-05 | 32 |
| rs7439324 | 4 | 44501351 | KCTD8 | C | T | 0.84 | 0.011 | 0.002 | 1.40E-08 | 7.10E-05 | 32 |
| rs12641981 | 4 | 45179883 | GNPDA2 | C | T | 0.57 | -0.022 | 0.001 | 2.20E-55 | 5.42E-04 | 246 |
| rs788858 | 4 | 82138300 | PRKG2 | A | G | 0.71 | 0.012 | 0.002 | 1.50E-15 | 1.40E-04 | 64 |
| rs7377083 | 4 | 102708997 | BANK1 | C | A | 0.57 | -0.014 | 0.001 | 6.40E-22 | 2.04E-04 | 93 |
| rs72675820 | 4 | 130746149 | C4orf33 | A | C | 0.36 | 0.010 | 0.001 | 1.00E-11 | 1.02E-04 | 46 |
| rs35189091 | 4 | 137147696 | PCDH18 | A | G | 0.63 | 0.009 | 0.001 | 5.50E-10 | 8.49E-05 | 38 |
| rs11727676 | 4 | 145659064 | HHIP | T | C | 0.90 | 0.013 | 0.002 | 3.20E-08 | 6.75E-05 | 31 |
| rs3936511 | 5 | 55860781 | AC022431.2 | A | G | 0.81 | 0.011 | 0.002 | 1.10E-09 | 8.19E-05 | 37 |
| rs6449532 | 5 | 60715446 | ZSWIM6 | C | T | 0.64 | 0.011 | 0.001 | 3.30E-14 | 1.27E-04 | 58 |
| rs9291816 | 5 | 63932508 | RGS7BP | C | T | 0.68 | 0.013 | 0.001 | 3.20E-19 | 1.77E-04 | 80 |

| | | | | | | | | | | | |
|---|---|---|---|---|---|---|---|---|---|---|---|
| rs39862 | 5 | 66185151 | MAST4 | T | C | 0.72 | 0.012 | 0.002 | 7.40E-14 | 1.23E-04 | 56 |
| rs2307111 | 5 | 75003678 | POC5 | T | C | 0.60 | 0.009 | 0.001 | 1.10E-09 | 8.20E-05 | 37 |
| rs1422067 | 5 | 77424836 | AP3B1 | C | T | 0.76 | 0.011 | 0.002 | 4.60E-11 | 9.56E-05 | 43 |
| rs2115885 | 5 | 87598818 | TMEM161B | G | A | 0.79 | 0.011 | 0.002 | 1.00E-09 | 8.23E-05 | 37 |
| rs77960 | 5 | 103964585 | NUDT12 | G | A | 0.67 | 0.010 | 0.001 | 2.20E-12 | 1.09E-04 | 49 |
| rs4958568 | 5 | 152016093 | NMUR2 | G | A | 0.72 | 0.010 | 0.002 | 2.20E-10 | 8.88E-05 | 40 |
| rs7719067 | 5 | 153538241 | MFAP3 | A | G | 0.43 | 0.013 | 0.001 | 1.40E-21 | 2.01E-04 | 91 |
| rs7711823 | 5 | 158489315 | EBF1 | A | G | 0.64 | 0.008 | 0.001 | 9.00E-09 | 7.29E-05 | 33 |
| rs918472 | 5 | 170738836 | TLX3 | G | A | 0.28 | -0.010 | 0.002 | 2.60E-10 | 8.82E-05 | 40 |
| rs12214497 | 6 | 10015908 | OFCC1 | G | T | 0.65 | 0.011 | 0.001 | 9.70E-13 | 1.12E-04 | 51 |
| rs10498713 | 6 | 22729300 | HDGFL1 | G | T | 0.85 | -0.012 | 0.002 | 2.10E-09 | 7.91E-05 | 36 |
| rs35162296 | 6 | 26318262 | HIST1H4H | C | T | 0.89 | -0.020 | 0.002 | 1.20E-18 | 1.72E-04 | 78 |
| rs34196306 | 6 | 27425644 | ZNF184 | G | C | 0.89 | -0.020 | 0.002 | 2.60E-18 | 1.68E-04 | 76 |
| rs3131336 | 6 | 28831611 | TRIM27 | C | T | 0.88 | -0.019 | 0.002 | 2.10E-18 | 1.69E-04 | 77 |
| rs3129942 | 6 | 32338283 | C6orf10 | G | T | 0.74 | -0.014 | 0.002 | 2.90E-18 | 1.68E-04 | 76 |
| rs9366803 | 6 | 32621132 | HLA-DQB1 | T | C | 0.68 | -0.011 | 0.002 | 1.10E-10 | 9.17E-05 | 42 |
| rs686431 | 6 | 35974217 | SLC26A8 | C | T | 0.98 | -0.030 | 0.005 | 6.70E-09 | 7.42E-05 | 34 |
| rs73422097 | 6 | 41727740 | PGC | A | G | 0.70 | -0.011 | 0.002 | 3.10E-12 | 1.07E-04 | 49 |
| rs76187039 | 6 | 43233990 | TTBK1 | G | T | 0.87 | -0.011 | 0.002 | 3.40E-08 | 6.72E-05 | 30 |
| rs3798519 | 6 | 50788778 | TFAP2B | A | C | 0.82 | -0.025 | 0.002 | 9.70E-43 | 4.14E-04 | 188 |
| rs1775255 | 6 | 51243035 | PKHD1 | G | T | 0.52 | -0.013 | 0.001 | 2.30E-21 | 1.99E-04 | 90 |
| rs1342831 | 6 | 54096151 | MLIP | T | C | 0.94 | -0.022 | 0.003 | 8.70E-14 | 1.23E-04 | 56 |
| rs12110721 | 6 | 55190480 | GFRAL | G | A | 0.83 | -0.019 | 0.002 | 1.60E-23 | 2.20E-04 | 100 |
| rs9370527 | 6 | 56245812 | COL21A1 | G | A | 0.76 | -0.009 | 0.002 | 6.90E-09 | 7.41E-05 | 34 |
| rs435775 | 6 | 97237609 | GPR63 | A | G | 0.25 | -0.010 | 0.002 | 5.20E-10 | 8.51E-05 | 39 |
| rs6931604 | 6 | 98578215 | POU3F2 | C | T | 0.40 | -0.008 | 0.001 | 6.20E-09 | 7.45E-05 | 34 |
| rs34260097 | 6 | 100727703 | SIM1 | T | G | 0.78 | -0.018 | 0.002 | 2.50E-26 | 2.49E-04 | 113 |
| rs7759938 | 6 | 105378954 | LIN28B | C | T | 0.32 | -0.010 | 0.001 | 3.20E-11 | 9.71E-05 | 44 |
| rs7753558 | 6 | 117523471 | VGLL2 | C | A | 0.37 | 0.009 | 0.001 | 5.10E-10 | 8.53E-05 | 39 |
| rs1452991 | 6 | 141473363 | NMBR | G | A | 0.63 | -0.010 | 0.001 | 5.40E-12 | 1.05E-04 | 48 |
| rs796915 | 6 | 154304628 | OPRM1 | C | G | 0.31 | -0.013 | 0.002 | 2.60E-17 | 1.58E-04 | 72 |
| rs62425398 | 6 | 166416028 | PDE10A | C | A | 0.89 | -0.015 | 0.002 | 1.90E-11 | 9.94E-05 | 45 |
| rs2349179 | 7 | 8404389 | NXPH1 | T | A | 0.62 | -0.008 | 0.001 | 5.00E-08 | 6.56E-05 | 30 |
| rs2722406 | 7 | 24306762 | NPY | C | T | 0.72 | -0.012 | 0.002 | 4.60E-15 | 1.36E-04 | 61 |
| rs4723263 | 7 | 33194826 | BBS9 | G | C | 0.57 | -0.008 | 0.001 | 3.60E-09 | 7.69E-05 | 35 |
| rs10234366 | 7 | 46743746 | AC011294.3 | G | A | 0.90 | -0.014 | 0.002 | 1.10E-09 | 8.18E-05 | 37 |
| rs1852006 | 7 | 77829768 | MAGI2 | G | A | 0.64 | 0.009 | 0.001 | 1.70E-09 | 8.00E-05 | 36 |
| rs6974282 | 7 | 100098295 | NYAP1 | C | T | 0.80 | 0.010 | 0.002 | 1.50E-08 | 7.06E-05 | 32 |
| rs7808296 | 7 | 103127620 | RELN | C | T | 0.68 | -0.010 | 0.002 | 2.80E-11 | 9.78E-05 | 44 |
| rs262338 | 7 | 103417134 | RELN | G | T | 0.55 | -0.009 | 0.001 | 4.80E-10 | 8.55E-05 | 39 |
| rs10953577 | 7 | 108263540 | DNAJB9 | T | C | 0.62 | -0.009 | 0.001 | 3.40E-09 | 7.71E-05 | 35 |
| rs67679818 | 7 | 110672704 | IMMP2L | C | T | 0.42 | 0.008 | 0.001 | 4.70E-08 | 6.58E-05 | 30 |
| rs6979832 | 7 | 127856276 | LEP | A | G | 0.50 | -0.009 | 0.001 | 2.70E-11 | 9.79E-05 | 44 |
| rs11525873 | 7 | 138817193 | TTC26 | T | C | 0.90 | 0.017 | 0.002 | 1.60E-13 | 1.20E-04 | 54 |
| rs10503246 | 8 | 4130363 | CSMD1 | A | G | 0.71 | -0.010 | 0.002 | 4.50E-10 | 8.58E-05 | 39 |

| | | | | | | | | | | | |
|---|---|---|---|---|---|---|---|---|---|---|---|
| rs77976727 | 8 | 4300554 | CSMD1 | C | T | 0.91 | -0.015 | 0.002 | 3.10E-10 | 8.74E-05 | 40 |
| rs7814267 | 8 | 5545084 | CSMD1 | A | G | 0.81 | -0.011 | 0.002 | 2.30E-09 | 7.87E-05 | 36 |
| rs11777719 | 8 | 8581038 | CLDN23 | A | G | 0.72 | -0.012 | 0.002 | 8.50E-14 | 1.23E-04 | 56 |
| rs13256357 | 8 | 9979535 | MSRA | C | T | 0.80 | -0.014 | 0.002 | 6.30E-15 | 1.34E-04 | 61 |
| rs2409743 | 8 | 11070360 | XKR6 | C | G | 0.50 | 0.011 | 0.001 | 3.00E-15 | 1.37E-04 | 62 |
| rs10503555 | 8 | 15763818 | TUSC3 | A | G | 0.56 | 0.008 | 0.001 | 3.90E-08 | 6.66E-05 | 30 |
| rs884152 | 8 | 25770557 | EBF2 | G | T | 0.36 | -0.009 | 0.001 | 6.00E-09 | 7.47E-05 | 34 |
| rs7012648 | 8 | 28091482 | ELP3 | G | A | 0.41 | -0.010 | 0.001 | 8.40E-12 | 1.03E-04 | 47 |
| rs4739558 | 8 | 38337264 | FGFR1 | A | G | 0.40 | 0.008 | 0.001 | 6.60E-09 | 7.43E-05 | 34 |
| rs10095724 | 8 | 53739232 | RB1CC1 | G | A | 0.64 | 0.009 | 0.001 | 1.40E-10 | 9.10E-05 | 41 |
| rs10111937 | 8 | 54160092 | OPRK1 | C | T | 0.70 | -0.008 | 0.002 | 4.20E-08 | 6.63E-05 | 30 |
| rs7840305 | 8 | 57168101 | CHCHD7 | A | G | 0.63 | 0.008 | 0.001 | 2.20E-08 | 6.92E-05 | 31 |
| rs13254613 | 8 | 64804804 | YTHDF3 | A | C | 0.65 | -0.013 | 0.001 | 1.60E-17 | 1.60E-04 | 73 |
| rs7817581 | 8 | 143136114 | TSNARE1 | G | A | 0.67 | 0.009 | 0.001 | 6.80E-09 | 7.41E-05 | 34 |
| rs10962279 | 9 | 16103030 | CCDC171 | T | C | 0.78 | -0.010 | 0.002 | 1.70E-09 | 8.00E-05 | 36 |
| rs3118252 | 9 | 25115154 | TUSC1 | G | C | 0.42 | -0.008 | 0.001 | 6.10E-09 | 7.46E-05 | 34 |
| rs1935354 | 9 | 27822531 | LINGO2 | T | C | 0.52 | -0.010 | 0.001 | 5.60E-13 | 1.15E-04 | 52 |
| rs1619120 | 9 | 87302196 | NTRK2 | A | G | 0.40 | -0.008 | 0.001 | 1.80E-08 | 7.00E-05 | 32 |
| rs4744246 | 9 | 96254464 | FAM120A | A | G | 0.66 | -0.016 | 0.001 | 1.70E-26 | 2.50E-04 | 113 |
| rs7020564 | 9 | 109670016 | ZNF462 | A | T | 0.70 | 0.010 | 0.002 | 1.40E-10 | 9.10E-05 | 41 |
| rs957512 | 9 | 120405705 | TLR4 | T | C | 0.67 | 0.010 | 0.001 | 4.70E-12 | 1.05E-04 | 48 |
| rs10116891 | 9 | 122651993 | RP11-295D22.1 | G | A | 0.90 | -0.014 | 0.002 | 5.60E-09 | 7.49E-05 | 34 |
| rs2275241 | 9 | 129370576 | LMX1B | G | A | 0.63 | -0.010 | 0.001 | 6.30E-13 | 1.14E-04 | 52 |
| rs117911387 | 9 | 130446836 | STXBP1 | G | A | 0.95 | -0.025 | 0.003 | 1.60E-13 | 1.20E-04 | 54 |
| rs7084503 | 10 | 2666859 | PFKP | T | C | 0.49 | 0.013 | 0.001 | 5.70E-20 | 1.85E-04 | 84 |
| rs11256627 | 10 | 10535954 | CELF2 | G | A | 0.30 | -0.009 | 0.002 | 9.30E-09 | 7.28E-05 | 33 |
| rs4572029 | 10 | 70889053 | VPS26A | A | G | 0.79 | 0.011 | 0.002 | 2.00E-10 | 8.93E-05 | 40 |
| rs10823504 | 10 | 72034062 | NPFFR1 | G | A | 0.94 | 0.016 | 0.003 | 3.70E-08 | 6.68E-05 | 30 |
| rs2242258 | 10 | 75607168 | CAMK2G | T | C | 0.74 | -0.009 | 0.002 | 8.80E-09 | 7.30E-05 | 33 |
| rs17399739 | 10 | 87490850 | GRID1 | A | G | 0.93 | -0.021 | 0.003 | 3.80E-14 | 1.26E-04 | 57 |
| rs10887571 | 10 | 88030441 | GRID1 | C | T | 0.55 | -0.008 | 0.001 | 1.80E-08 | 6.99E-05 | 32 |
| rs41310284 | 10 | 102447647 | PAX2 | C | A | 0.90 | 0.021 | 0.002 | 1.40E-18 | 1.71E-04 | 77 |
| rs75387636 | 10 | 120278394 | PRLHR | G | A | 0.96 | -0.021 | 0.004 | 1.50E-09 | 8.07E-05 | 37 |
| rs2939931 | 10 | 121636406 | MCMBP | T | C | 0.52 | -0.008 | 0.001 | 1.90E-08 | 6.96E-05 | 32 |
| rs1061072 | 10 | 126684796 | CTBP2 | G | A | 0.89 | 0.013 | 0.002 | 2.40E-08 | 6.88E-05 | 31 |
| rs56133711 | 11 | 27723334 | BDNF | G | A | 0.74 | -0.016 | 0.002 | 3.30E-24 | 2.27E-04 | 103 |
| rs4267058 | 11 | 28645584 | METTL15 | T | C | 0.63 | 0.009 | 0.001 | 1.30E-09 | 8.12E-05 | 37 |
| rs661878 | 11 | 29188691 | METTL15 | A | G | 0.87 | 0.014 | 0.002 | 2.80E-11 | 9.78E-05 | 44 |
| rs3181269 | 11 | 33755956 | CD59 | C | T | 0.75 | 0.009 | 0.002 | 5.30E-09 | 7.52E-05 | 34 |
| rs7951870 | 11 | 46373311 | DGKZ | T | C | 0.83 | -0.012 | 0.002 | 5.30E-10 | 8.51E-05 | 39 |
| rs12798028 | 11 | 47604639 | NDUFS3 | C | T | 0.59 | -0.015 | 0.001 | 1.10E-24 | 2.32E-04 | 105 |
| rs3862342 | 11 | 49357043 | FOLH1 | C | T | 0.29 | -0.009 | 0.002 | 3.00E-08 | 6.77E-05 | 31 |
| rs2958542 | 11 | 62181882 | SCGB1A1 | C | T | 0.63 | 0.008 | 0.001 | 1.80E-08 | 7.00E-05 | 32 |
| rs10791902 | 11 | 67093360 | SSH3 | C | T | 0.59 | -0.008 | 0.001 | 3.60E-08 | 6.70E-05 | 30 |
| rs10896348 | 11 | 68357368 | PPP6R3 | T | C | 0.72 | 0.012 | 0.002 | 3.40E-14 | 1.27E-04 | 57 |

| | | | | | | | | | | | |
|---|---|---|---|---|---|---|---|---|---|---|---|
| rs10796828 | 11 | 69490346 | ORAOV1 | T | G | 0.37 | -0.011 | 0.001 | 7.10E-15 | 1.34E-04 | 61 |
| rs11215403 | 11 | 115058585 | CADM1 | G | A | 0.76 | 0.013 | 0.002 | 5.10E-16 | 1.45E-04 | 66 |
| rs7123283 | 11 | 122809055 | C11orf63 | C | T | 0.53 | 0.008 | 0.001 | 3.70E-09 | 7.67E-05 | 35 |
| rs10790809 | 11 | 126372550 | KIRREL3 | A | G | 0.44 | -0.010 | 0.001 | 7.20E-12 | 1.04E-04 | 47 |
| rs55726687 | 12 | 991306 | WNK1 | G | A | 0.79 | -0.015 | 0.002 | 2.70E-17 | 1.58E-04 | 72 |
| rs2187642 | 12 | 11855624 | ETV6 | A | C | 0.38 | -0.011 | 0.001 | 1.10E-14 | 1.32E-04 | 60 |
| rs10841379 | 12 | 19992303 | AEBP2 | A | G | 0.32 | -0.008 | 0.002 | 2.90E-08 | 6.79E-05 | 31 |
| rs10842356 | 12 | 24621348 | BCAT1 | A | T | 0.48 | 0.008 | 0.001 | 4.10E-09 | 7.63E-05 | 35 |
| rs61937656 | 12 | 39483502 | CPNE8 | G | A | 0.77 | 0.012 | 0.002 | 2.70E-12 | 1.08E-04 | 49 |
| rs7958241 | 12 | 49509262 | LMBR1L | A | G | 0.66 | -0.013 | 0.001 | 7.90E-19 | 1.73E-04 | 79 |
| rs7132908 | 12 | 50263148 | FAIM2 | G | A | 0.62 | -0.031 | 0.001 | 1.70E-104 | 1.04E-03 | 471 |
| rs836179 | 12 | 50503082 | GPD1 | A | G | 0.63 | 0.010 | 0.001 | 1.20E-11 | 1.01E-04 | 46 |
| rs78607331 | 12 | 57648644 | R3HDM2 | C | T | 0.96 | -0.024 | 0.003 | 1.70E-12 | 1.10E-04 | 50 |
| rs7306710 | 12 | 66376091 | HMGA2 | T | C | 0.48 | 0.010 | 0.001 | 1.60E-12 | 1.10E-04 | 50 |
| rs10860295 | 12 | 98542699 | RP11-181C3.1 | T | C | 0.55 | -0.008 | 0.001 | 1.80E-09 | 7.98E-05 | 36 |
| rs1552759 | 12 | 99640557 | ANKS1B | T | C | 0.34 | 0.009 | 0.001 | 6.30E-10 | 8.43E-05 | 38 |
| rs12817542 | 12 | 103736499 | C12orf42 | C | T | 0.94 | -0.018 | 0.003 | 1.70E-09 | 8.02E-05 | 36 |
| rs61936936 | 12 | 116391685 | MED13L | A | T | 0.90 | -0.014 | 0.002 | 7.80E-09 | 7.35E-05 | 33 |
| rs7305424 | 12 | 118399491 | KSR2 | A | T | 0.66 | -0.010 | 0.001 | 2.30E-12 | 1.09E-04 | 49 |
| rs12308065 | 12 | 120624085 | GCN1L1 | A | G | 0.37 | -0.008 | 0.001 | 1.10E-08 | 7.19E-05 | 33 |
| rs28629903 | 12 | 122497001 | BCL7A | T | C | 0.44 | 0.009 | 0.001 | 1.50E-10 | 9.06E-05 | 41 |
| rs7989098 | 13 | 27925496 | GTF3A | T | C | 0.26 | -0.012 | 0.002 | 1.00E-13 | 1.22E-04 | 55 |
| rs9652090 | 13 | 27983367 | GTF3A | G | T | 0.54 | -0.008 | 0.001 | 4.10E-09 | 7.63E-05 | 35 |
| rs1933437 | 13 | 28624294 | FLT3 | G | A | 0.37 | 0.014 | 0.001 | 1.60E-22 | 2.10E-04 | 95 |
| rs9603697 | 13 | 40783323 | AL133318.1 | C | T | 0.67 | -0.013 | 0.001 | 5.00E-18 | 1.65E-04 | 75 |
| rs9594686 | 13 | 42723197 | DGKH | C | T | 0.82 | 0.011 | 0.002 | 6.40E-09 | 7.44E-05 | 34 |
| rs12429545 | 13 | 54102206 | OLFM4 | G | A | 0.87 | -0.021 | 0.002 | 1.20E-22 | 2.12E-04 | 96 |
| rs9538141 | 13 | 59178258 | PCDH17 | G | A | 0.49 | -0.012 | 0.001 | 2.20E-16 | 1.49E-04 | 67 |
| rs1333010 | 13 | 66205228 | PCDH9 | G | A | 0.39 | 0.012 | 0.001 | 5.20E-17 | 1.55E-04 | 70 |
| rs1576655 | 13 | 79587841 | RBM26 | A | C | 0.40 | -0.012 | 0.001 | 4.30E-16 | 1.46E-04 | 66 |
| rs61978655 | 14 | 30491807 | PRKD1 | G | A | 0.96 | -0.034 | 0.004 | 1.30E-20 | 1.91E-04 | 87 |
| rs7161424 | 14 | 33309274 | AKAP6 | G | A | 0.52 | -0.010 | 0.001 | 4.50E-13 | 1.16E-04 | 52 |
| rs1865719 | 14 | 79923667 | NRXN3 | A | G | 0.37 | -0.011 | 0.001 | 8.40E-14 | 1.23E-04 | 56 |
| rs10133279 | 14 | 82702712 | SEL1L | C | T | 0.56 | -0.009 | 0.001 | 3.00E-10 | 8.75E-05 | 40 |
| rs7145052 | 14 | 92461192 | TRIP11 | C | T | 0.55 | -0.008 | 0.001 | 3.70E-09 | 7.67E-05 | 35 |
| rs7159126 | 14 | 93783176 | BTBD7 | T | C | 0.72 | 0.009 | 0.002 | 2.50E-08 | 6.86E-05 | 31 |
| rs78420139 | 14 | 101201687 | DLK1 | G | A | 0.94 | 0.017 | 0.003 | 3.60E-08 | 6.70E-05 | 30 |
| rs12436513 | 14 | 103269755 | TRAF3 | C | A | 0.36 | 0.009 | 0.001 | 4.20E-09 | 7.62E-05 | 35 |
| rs824207 | 15 | 24007729 | NDN | A | G | 0.47 | -0.009 | 0.001 | 3.70E-11 | 9.66E-05 | 44 |
| rs62048187 | 15 | 38117049 | TMCO5A | G | C | 0.68 | -0.008 | 0.002 | 4.00E-08 | 6.66E-05 | 30 |
| rs10519136 | 15 | 47933163 | SEMA6D | C | T | 0.60 | 0.008 | 0.001 | 1.60E-08 | 7.04E-05 | 32 |
| rs8030456 | 15 | 68076856 | MAP2K5 | C | T | 0.77 | 0.021 | 0.002 | 2.60E-37 | 3.59E-04 | 163 |
| rs7162542 | 15 | 84514290 | ADAMTSL3 | C | G | 0.44 | 0.009 | 0.001 | 3.80E-11 | 9.65E-05 | 44 |
| rs3817428 | 15 | 89415247 | ACAN | C | G | 0.74 | -0.011 | 0.002 | 1.30E-11 | 1.01E-04 | 46 |
| rs1000471 | 15 | 89986583 | RHCG | C | T | 0.78 | -0.009 | 0.002 | 3.70E-08 | 6.68E-05 | 30 |

| | | | | | | | | | | | |
|---|---|---|---|---|---|---|---|---|---|---|---|
| rs2970356 | 15 | 90623540 | ZNF710 | C | G | 0.68 | -0.010 | 0.002 | 4.50E-11 | 9.57E-05 | 43 |
| rs72755233 | 15 | 100692953 | ADAMTS17 | G | A | 0.89 | -0.018 | 0.002 | 2.10E-15 | 1.39E-04 | 63 |
| rs2238435 | 16 | 4014282 | ADCY9 | C | G | 0.39 | -0.016 | 0.001 | 2.90E-30 | 2.88E-04 | 131 |
| rs55880046 | 16 | 19941557 | GPRC5B | T | G | 0.86 | 0.030 | 0.002 | 3.70E-50 | 4.89E-04 | 222 |
| rs4432271 | 16 | 20245283 | GP2 | C | T | 0.13 | -0.017 | 0.002 | 1.00E-16 | 1.52E-04 | 69 |
| rs9922288 | 16 | 24550930 | RBBP6 | A | G | 0.24 | 0.010 | 0.002 | 3.40E-10 | 8.69E-05 | 39 |
| rs62037365 | 16 | 28868962 | SH2B1 | C | G | 0.60 | -0.014 | 0.001 | 9.90E-22 | 2.02E-04 | 92 |
| rs4889630 | 16 | 30877544 | BCL7C | T | C | 0.20 | 0.012 | 0.002 | 1.90E-12 | 1.09E-04 | 50 |
| rs4783789 | 16 | 51446707 | SALL1 | T | C | 0.78 | 0.010 | 0.002 | 1.40E-08 | 7.10E-05 | 32 |
| rs1421085 | 16 | 53800954 | FTO | T | C | 0.60 | -0.047 | 0.001 | 1.10E-241 | 2.43E-03 | 1102 |
| rs594585 | 16 | 65939803 | CDH5 | T | G | 0.41 | 0.008 | 0.001 | 3.00E-08 | 6.78E-05 | 31 |
| rs117903946 | 16 | 67449639 | ZDHHC1 | G | A | 0.97 | -0.032 | 0.004 | 3.20E-16 | 1.47E-04 | 67 |
| rs7672 | 16 | 68294800 | PLA2G15 | C | G | 0.29 | 0.009 | 0.002 | 7.90E-09 | 7.35E-05 | 33 |
| rs4985555 | 16 | 70690166 | IL34 | A | G | 0.50 | 0.009 | 0.001 | 4.60E-10 | 8.57E-05 | 39 |
| rs11642090 | 16 | 81730582 | CMIP | T | C | 0.63 | -0.012 | 0.001 | 9.20E-16 | 1.43E-04 | 65 |
| rs72819571 | 17 | 2170501 | SMG6 | G | T | 0.65 | 0.012 | 0.001 | 6.70E-16 | 1.44E-04 | 65 |
| rs67603370 | 17 | 7524504 | SHBG | G | A | 0.92 | -0.016 | 0.003 | 3.40E-09 | 7.71E-05 | 35 |
| rs3815156 | 17 | 29685150 | NF1 | A | G | 0.82 | -0.010 | 0.002 | 2.20E-08 | 6.91E-05 | 31 |
| rs12601380 | 17 | 34904985 | GGNBP2 | A | C | 0.59 | 0.008 | 0.001 | 4.30E-08 | 6.62E-05 | 30 |
| rs9299 | 17 | 46669430 | HOXB3 | C | T | 0.34 | -0.010 | 0.001 | 7.30E-11 | 9.36E-05 | 42 |
| rs17637472 | 17 | 47461433 | RP11-81K2.1 | G | A | 0.60 | -0.011 | 0.001 | 4.10E-14 | 1.26E-04 | 57 |
| rs7217460 | 17 | 65393344 | PITPNC1 | G | A | 0.77 | 0.009 | 0.002 | 2.80E-08 | 6.81E-05 | 31 |
| rs12941038 | 17 | 66509143 | PRKAR1A | C | T | 0.77 | -0.010 | 0.002 | 1.00E-08 | 7.24E-05 | 33 |
| rs2246623 | 17 | 74084449 | EXOC7 | C | T | 0.55 | 0.009 | 0.001 | 2.20E-10 | 8.88E-05 | 40 |
| rs11150745 | 17 | 78757626 | RPTOR | A | G | 0.68 | 0.012 | 0.002 | 7.00E-16 | 1.44E-04 | 65 |
| rs7503580 | 17 | 79087036 | BAIAP2 | C | T | 0.84 | -0.011 | 0.002 | 8.70E-09 | 7.31E-05 | 33 |
| rs1013737 | 18 | 937050 | ADCYAP1 | G | C | 0.51 | -0.011 | 0.001 | 3.30E-14 | 1.27E-04 | 58 |
| rs1808579 | 18 | 21104888 | NPC1 | C | T | 0.52 | 0.009 | 0.001 | 7.70E-11 | 9.34E-05 | 42 |
| rs7237444 | 18 | 39468982 | PIK3C3 | G | A | 0.33 | 0.010 | 0.002 | 3.40E-11 | 9.69E-05 | 44 |
| rs7239114 | 18 | 45921214 | ZBTB7C | G | A | 0.46 | -0.013 | 0.001 | 1.80E-21 | 2.00E-04 | 91 |
| rs68015088 | 18 | 51484010 | MBD2 | G | A | 0.65 | 0.008 | 0.001 | 4.30E-08 | 6.62E-05 | 30 |
| rs12606230 | 18 | 52492252 | RAB27B | T | C | 0.77 | -0.013 | 0.002 | 1.50E-14 | 1.31E-04 | 59 |
| rs663129 | 18 | 57838401 | MC4R | G | A | 0.77 | -0.033 | 0.002 | 8.80E-90 | 8.90E-04 | 404 |
| rs113728099 | 18 | 58157767 | MC4R | G | A | 0.98 | 0.044 | 0.005 | 4.30E-22 | 2.06E-04 | 93 |
| rs8096658 | 18 | 77156537 | NFATC1 | C | G | 0.51 | 0.009 | 0.001 | 2.30E-10 | 8.87E-05 | 40 |
| rs62621197 | 19 | 8670147 | ADAMTS10 | C | T | 0.96 | -0.023 | 0.004 | 1.00E-09 | 8.22E-05 | 37 |
| rs4545941 | 19 | 16534207 | EPS15L1 | T | C | 0.83 | -0.011 | 0.002 | 1.10E-08 | 7.22E-05 | 33 |
| rs116399833 | 19 | 18502374 | LRRC25 | C | A | 0.77 | -0.012 | 0.002 | 4.60E-13 | 1.16E-04 | 52 |
| rs4808961 | 19 | 19577215 | GATAD2A | C | G | 0.63 | 0.009 | 0.001 | 1.20E-09 | 8.16E-05 | 37 |
| rs3810304 | 19 | 30861683 | ZNF536 | A | G | 0.23 | 0.010 | 0.002 | 8.50E-10 | 8.30E-05 | 38 |
| rs1800437 | 19 | 46181392 | GIPR | G | C | 0.81 | 0.012 | 0.002 | 2.90E-11 | 9.76E-05 | 44 |
| rs3810291 | 19 | 47569003 | ZC3H4 | G | A | 0.32 | -0.015 | 0.001 | 5.60E-23 | 2.15E-04 | 97 |
| rs601338 | 19 | 49206674 | FUT2 | G | A | 0.49 | 0.009 | 0.001 | 2.10E-11 | 9.90E-05 | 45 |
| rs16996644 | 20 | 15813475 | MACROD2 | C | G | 0.87 | -0.019 | 0.002 | 4.80E-20 | 1.85E-04 | 84 |
| rs947088 | 20 | 17171373 | PCSK2 | G | T | 0.28 | -0.011 | 0.002 | 9.50E-12 | 1.02E-04 | 46 |

| | | | | | | | | | | | |
|---|---|---|---|---|---|---|---|---|---|---|---|
| rs8117463 | 20 | 17231063 | PCSK2 | G | A | 0.68 | 0.009 | 0.001 | 3.30E-09 | 7.72E-05 | 35 |
| rs73085586 | 20 | 22430241 | FOXA2 | G | A | 0.80 | -0.010 | 0.002 | 3.00E-08 | 6.78E-05 | 31 |
| rs2281148 | 20 | 36433288 | CTNNBL1 | T | C | 0.75 | -0.010 | 0.002 | 1.50E-09 | 8.06E-05 | 37 |
| rs2207894 | 20 | 54387343 | CBLN4 | C | T | 0.81 | 0.014 | 0.002 | 8.00E-16 | 1.43E-04 | 65 |
| rs117455294 | 20 | 57427951 | GNAS | C | A | 0.95 | 0.019 | 0.003 | 1.20E-09 | 8.17E-05 | 37 |
| rs8130408 | 21 | 39237138 | KCNJ6 | A | C | 0.26 | -0.009 | 0.002 | 2.30E-08 | 6.89E-05 | 31 |
| rs13047416 | 21 | 40309436 | ETS2 | C | G | 0.62 | 0.012 | 0.001 | 1.40E-17 | 1.61E-04 | 73 |
| rs78907487 | 22 | 22151939 | MAPK1 | A | C | 0.85 | -0.012 | 0.002 | 4.10E-10 | 8.62E-05 | 39 |
| rs9610387 | 22 | 36476762 | RBFOX2 | G | A | 0.91 | 0.014 | 0.003 | 3.90E-08 | 6.66E-05 | 30 |
| rs6001872 | 22 | 40703245 | TNRC6B | A | G | 0.65 | 0.013 | 0.001 | 1.50E-17 | 1.60E-04 | 73 |
| rs9611560 | 22 | 41750622 | ZC3H7B | T | C | 0.26 | 0.009 | 0.002 | 6.50E-09 | 7.43E-05 | 34 |
| Genome-wide significant SNPs for early life body size in UK Biobank (men only) | | | | | | | | | | | |
| SNP | Chr | Position | Closest gene | EA | OA | EAF | Beta | SE | pvalue | $R^2$ | F statistic |
| rs12140153 | 1 | 62579891 | INADL | G | T | 0.905 | 0.023 | 0.004 | 6.4E-11 | 2.07E-04 | 43 |
| rs2012697 | 1 | 72819612 | NEGR1 | T | C | 0.401 | -0.019 | 0.002 | 4.5E-20 | 4.07E-04 | 84 |
| rs4650277 | 1 | 74993721 | TNNI3K | A | G | 0.435 | 0.023 | 0.002 | 1.4E-27 | 5.73E-04 | 118 |
| rs7550711 | 1 | 110082886 | GPR61 | C | T | 0.974 | -0.051 | 0.006 | 5.8E-15 | 2.95E-04 | 61 |
| rs539515 | 1 | 177889025 | SEC16B | A | C | 0.795 | -0.039 | 0.003 | 1.3E-52 | 1.13E-03 | 233 |
| rs78444298 | 1 | 184672098 | EDEM3 | G | A | 0.981 | 0.047 | 0.008 | 3.5E-10 | 1.90E-04 | 39 |
| rs1772143 | 1 | 205799987 | PM20D1 | T | A | 0.587 | 0.015 | 0.002 | 2.2E-12 | 2.38E-04 | 49 |
| rs77165542 | 2 | 430975 | FAM150B | C | T | 0.965 | 0.073 | 0.006 | 1.7E-38 | 8.14E-04 | 168 |
| rs6749422 | 2 | 25150011 | ADCY3 | C | G | 0.514 | -0.031 | 0.002 | 1.4E-52 | 1.13E-03 | 233 |
| rs2862874 | 2 | 58990485 | FANCL | G | T | 0.388 | -0.012 | 0.002 | 1E-08 | 1.59E-04 | 33 |
| rs10496885 | 2 | 142285767 | LRP1B | G | A | 0.814 | 0.017 | 0.003 | 3.3E-10 | 1.91E-04 | 39 |
| rs115319174 | 2 | 207066474 | GPR1 | G | C | 0.942 | -0.043 | 0.004 | 1.4E-22 | 4.63E-04 | 96 |
| rs9880272 | 3 | 85687785 | CADM2 | T | G | 0.759 | -0.014 | 0.002 | 7.8E-09 | 1.61E-04 | 33 |
| rs34722008 | 4 | 38659594 | AC021860.1 | G | A | 0.646 | 0.012 | 0.002 | 4E-08 | 1.46E-04 | 30 |
| rs10938398 | 4 | 45186139 | GNPDA2 | G | A | 0.565 | -0.021 | 0.002 | 1.7E-23 | 4.83E-04 | 100 |
| rs13107325 | 4 | 103188709 | SLC39A8 | C | T | 0.924 | -0.023 | 0.004 | 5.2E-09 | 1.65E-04 | 34 |
| rs3212519 | 5 | 52351182 | ITGA2 | A | G | 0.930 | 0.022 | 0.004 | 2.9E-08 | 1.49E-04 | 31 |
| rs75577466 | 5 | 60605507 | ZSWIM6 | G | C | 0.823 | 0.017 | 0.003 | 3.6E-10 | 1.90E-04 | 39 |
| rs25842 | 5 | 66163135 | MAST4 | C | T | 0.723 | 0.013 | 0.002 | 5.4E-09 | 1.65E-04 | 34 |
| rs55654862 | 5 | 77428253 | AP3B1 | A | G | 0.763 | 0.014 | 0.002 | 1.6E-08 | 1.54E-04 | 32 |
| rs4958361 | 5 | 153545587 | MFAP3 | C | G | 0.472 | 0.012 | 0.002 | 3.8E-09 | 1.68E-04 | 35 |
| rs2240071 | 6 | 30070932 | TRIM31 | C | G | 0.747 | -0.013 | 0.002 | 3E-08 | 1.49E-04 | 31 |
| rs62405422 | 6 | 50796905 | TFAP2B | T | C | 0.820 | -0.022 | 0.003 | 2.5E-16 | 3.25E-04 | 67 |
| rs1775255 | 6 | 51243035 | PKHD1 | G | T | 0.524 | -0.013 | 0.002 | 1.1E-10 | 2.02E-04 | 42 |
| rs115597956 | 6 | 54042871 | MLIP | G | A | 0.945 | -0.030 | 0.005 | 3.5E-11 | 2.12E-04 | 44 |
| rs12110721 | 6 | 55190480 | GFRAL | G | A | 0.826 | -0.018 | 0.003 | 5.3E-11 | 2.08E-04 | 43 |
| rs2693560 | 6 | 117523671 | VGLL2 | A | G | 0.371 | 0.012 | 0.002 | 4.9E-08 | 1.44E-04 | 30 |
| rs1452991 | 6 | 141473363 | NMBR | G | A | 0.627 | -0.012 | 0.002 | 1.5E-08 | 1.55E-04 | 32 |
| rs16120 | 7 | 24334724 | NPY | A | G | 0.480 | 0.012 | 0.002 | 1.4E-08 | 1.56E-04 | 32 |
| rs7796922 | 7 | 33171110 | BBS9 | A | G | 0.906 | 0.020 | 0.004 | 2.9E-08 | 1.49E-04 | 31 |
| rs2979139 | 8 | 8268313 | SGK223 | A | G | 0.507 | -0.012 | 0.002 | 4.2E-09 | 1.67E-04 | 35 |

| | | | | | | | | | | | |
|---|---|---|---|---|---|---|---|---|---|---|---|
| rs12674871 | 8 | 9979144 | MSRA | C | T | 0.799 | -0.014 | 0.003 | 2.1E-08 | 1.52E-04 | 31 |
| rs10504620 | 8 | 77288303 | ZFHX4 | T | C | 0.301 | -0.015 | 0.002 | 1.3E-11 | 2.22E-04 | 46 |
| rs10968101 | 9 | 27772542 | LINGO2 | G | A | 0.522 | -0.014 | 0.002 | 2.2E-11 | 2.17E-04 | 45 |
| rs11790060 | 9 | 96202932 | FAM120AOS | T | C | 0.669 | -0.013 | 0.002 | 4.2E-09 | 1.67E-04 | 35 |
| rs41310284 | 10 | 102447647 | PAX2 | C | A | 0.899 | 0.024 | 0.003 | 1.9E-12 | 2.40E-04 | 50 |
| rs11030102 | 11 | 27681596 | BDNF | C | G | 0.740 | -0.018 | 0.002 | 4.1E-14 | 2.76E-04 | 57 |
| rs3817334 | 11 | 47650993 | MTCH2 | C | T | 0.593 | -0.014 | 0.002 | 6E-11 | 2.07E-04 | 43 |
| rs10896348 | 11 | 68357368 | PPP6R3 | T | C | 0.723 | 0.013 | 0.002 | 6E-09 | 1.64E-04 | 34 |
| rs11218734 | 11 | 122523246 | UBASH3B | A | G | 0.743 | -0.013 | 0.002 | 3.6E-08 | 1.47E-04 | 30 |
| rs7978659 | 12 | 49507127 | LMBR1L | G | T | 0.658 | -0.013 | 0.002 | 7E-10 | 1.84E-04 | 38 |
| rs7132908 | 12 | 50263148 | FAIM2 | G | A | 0.615 | -0.032 | 0.002 | 3.6E-51 | 1.09E-03 | 226 |
| rs7306710 | 12 | 66376091 | HMGA2 | T | C | 0.479 | 0.012 | 0.002 | 2E-09 | 1.74E-04 | 36 |
| rs7316962 | 13 | 28613993 | FLT3 | A | G | 0.453 | 0.013 | 0.002 | 4E-10 | 1.89E-04 | 39 |
| rs9568868 | 13 | 54107583 | OLFM4 | G | T | 0.870 | -0.025 | 0.003 | 7.6E-16 | 3.14E-04 | 65 |
| rs1576655 | 13 | 79587841 | RBM26 | A | C | 0.404 | -0.012 | 0.002 | 3.7E-08 | 1.47E-04 | 30 |
| rs61978655 | 14 | 30491807 | PRKD1 | G | A | 0.961 | -0.030 | 0.005 | 1.2E-08 | 1.57E-04 | 32 |
| rs2143975 | 14 | 33297398 | AKAP6 | C | G | 0.466 | 0.012 | 0.002 | 3.2E-09 | 1.70E-04 | 35 |
| rs7159126 | 14 | 93783176 | BTBD7 | T | C | 0.718 | 0.013 | 0.002 | 3.6E-08 | 1.47E-04 | 30 |
| rs3784710 | 15 | 68072458 | MAP2K5 | T | C | 0.774 | 0.019 | 0.002 | 5.1E-15 | 2.96E-04 | 61 |
| rs7190603 | 16 | 19928662 | GPRC5B | T | C | 0.858 | 0.030 | 0.003 | 1.2E-23 | 4.86E-04 | 101 |
| rs56094641 | 16 | 53806453 | FTO | A | G | 0.593 | -0.046 | 0.002 | 1.2E-108 | 2.37E-03 | 490 |
| rs17637472 | 17 | 47461433 | RP11-81K2.1 | G | A | 0.602 | -0.012 | 0.002 | 5.2E-09 | 1.65E-04 | 34 |
| rs2250081 | 17 | 74090700 | EXOC7 | G | A | 0.552 | 0.011 | 0.002 | 2.7E-08 | 1.50E-04 | 31 |
| rs7239114 | 18 | 45921214 | ZBTB7C | G | A | 0.459 | -0.013 | 0.002 | 8.1E-10 | 1.83E-04 | 38 |
| rs3764516 | 18 | 52494374 | RAB27B | A | C | 0.235 | 0.013 | 0.002 | 3.3E-08 | 1.48E-04 | 31 |
| rs663129 | 18 | 57838401 | MC4R | G | A | 0.766 | -0.031 | 0.002 | 2.5E-36 | 7.66E-04 | 158 |
| rs1532127 | 19 | 47571938 | ZC3H4 | G | A | 0.318 | -0.016 | 0.002 | 1.2E-12 | 2.45E-04 | 51 |
| rs1321434 | 20 | 6624443 | BMP2 | A | G | 0.540 | -0.013 | 0.002 | 5.4E-10 | 1.86E-04 | 39 |
| rs73898513 | 20 | 15806210 | MACROD2 | C | T | 0.878 | -0.019 | 0.003 | 3.7E-09 | 1.68E-04 | 35 |
| Genome-wide significant SNPs for early life body size in UK Biobank (women only) | | | | | | | | | | | |
| SNP | Chr | Position | Closest gene | EA | OA | EAF | Beta | SE | pvalue | $R^2$ | F statistic |
| rs212540 | 1 | 21593117 | ECE1 | C | T | 0.393 | 0.013 | 0.002 | 2.3E-10 | 1.63E-04 | 40 |
| rs582220 | 1 | 54724762 | SSBP3 | A | G | 0.433 | -0.011 | 0.002 | 4.5E-09 | 1.39E-04 | 34 |
| rs12140153 | 1 | 62579891 | INADL | G | T | 0.906 | 0.020 | 0.003 | 5.5E-09 | 1.38E-04 | 34 |
| rs2767486 | 1 | 65991203 | LEPR | A | G | 0.798 | -0.020 | 0.002 | 3.5E-17 | 2.88E-04 | 71 |
| rs7522014 | 1 | 66551759 | PDE4B | A | G | 0.673 | 0.012 | 0.002 | 4.2E-08 | 1.22E-04 | 30 |
| rs11209943 | 1 | 72750500 | NEGR1 | A | G | 0.397 | -0.017 | 0.002 | 8.1E-19 | 3.18E-04 | 78 |
| rs12042908 | 1 | 74997762 | TNNI3K | A | G | 0.438 | 0.032 | 0.002 | 2.1E-59 | 1.07E-03 | 264 |
| rs41279738 | 1 | 110082551 | GPR61 | T | G | 0.974 | -0.046 | 0.006 | 2.2E-14 | 2.36E-04 | 58 |
| rs543874 | 1 | 177889480 | SEC16B | A | G | 0.795 | -0.055 | 0.002 | 1.6E-115 | 2.11E-03 | 522 |
| rs10798139 | 1 | 187714179 | PLA2G4A | C | T | 0.783 | -0.014 | 0.002 | 5.9E-09 | 1.37E-04 | 34 |
| rs815339 | 1 | 190116575 | BRINP3 | T | A | 0.495 | -0.011 | 0.002 | 3.8E-08 | 1.23E-04 | 30 |
| rs4971239 | 1 | 203491150 | OPTC | G | A | 0.831 | -0.015 | 0.003 | 9.9E-09 | 1.33E-04 | 33 |
| rs62106258 | 2 | 417167 | FAM150B | T | C | 0.951 | 0.087 | 0.004 | 3.4E-84 | 1.53E-03 | 378 |
| rs12992672 | 2 | 632592 | TMEM18 | G | A | 0.172 | -0.044 | 0.003 | 1.1E-66 | 1.21E-03 | 298 |

| | | | | | | | | | | | |
|---|---|---|---|---|---|---|---|---|---|---|---|
| rs6738433 | 2 | 25159501 | DNAJC27 | G | C | 0.521 | -0.040 | 0.002 | 7.6E-97 | 1.77E-03 | 436 |
| rs146910503 | 2 | 25446473 | DNMT3A | G | A | 0.979 | 0.047 | 0.007 | 1.3E-11 | 1.86E-04 | 46 |
| rs1446725 | 2 | 77222421 | LRRTM4 | T | G | 0.587 | 0.014 | 0.002 | 4.8E-12 | 1.94E-04 | 48 |
| rs1483153 | 2 | 142358477 | LRP1B | C | T | 0.219 | -0.015 | 0.002 | 5.3E-10 | 1.56E-04 | 39 |
| rs55959207 | 2 | 161016981 | ITGB6 | A | C | 0.574 | 0.011 | 0.002 | 2E-08 | 1.28E-04 | 31 |
| rs17464221 | 2 | 188278203 | CALCRL | C | T | 0.711 | 0.012 | 0.002 | 8.7E-09 | 1.34E-04 | 33 |
| rs115319174 | 2 | 207066474 | GPR1 | G | C | 0.943 | -0.041 | 0.004 | 1.3E-22 | 3.89E-04 | 96 |
| rs2594989 | 3 | 11316143 | ATG7 | C | T | 0.178 | 0.019 | 0.003 | 1.8E-13 | 2.20E-04 | 54 |
| rs754635 | 3 | 42305131 | CCK | C | G | 0.113 | -0.017 | 0.003 | 4.8E-08 | 1.21E-04 | 30 |
| rs2034963 | 3 | 48170802 | CDC25A | G | C | 0.351 | 0.013 | 0.002 | 5.1E-11 | 1.75E-04 | 43 |
| rs2629881 | 3 | 59778271 | FHIT | C | T | 0.221 | -0.013 | 0.002 | 1.6E-08 | 1.29E-04 | 32 |
| rs79569013 | 3 | 61218295 | FHIT | T | G | 0.847 | 0.020 | 0.003 | 8.6E-14 | 2.26E-04 | 56 |
| rs818219 | 3 | 85374589 | CADM2 | T | C | 0.541 | -0.013 | 0.002 | 6.5E-12 | 1.91E-04 | 47 |
| rs2735556 | 3 | 88105360 | CGGBP1 | T | C | 0.885 | 0.018 | 0.003 | 1.9E-09 | 1.46E-04 | 36 |
| rs1199328 | 3 | 138112107 | MRAS | G | A | 0.191 | 0.015 | 0.002 | 3.2E-10 | 1.60E-04 | 40 |
| rs76152047 | 3 | 141183792 | ZBTB38 | A | G | 0.934 | -0.025 | 0.004 | 1.3E-10 | 1.67E-04 | 41 |
| rs7656673 | 4 | 30840331 | PCDH7 | A | G | 0.598 | -0.014 | 0.002 | 3.5E-12 | 1.96E-04 | 48 |
| rs12641981 | 4 | 45179883 | GNPDA2 | C | T | 0.566 | -0.023 | 0.002 | 1.2E-32 | 5.74E-04 | 142 |
| rs1349641 | 4 | 82212652 | PRKG2 | T | G | 0.603 | 0.012 | 0.002 | 2.9E-10 | 1.61E-04 | 40 |
| rs7377083 | 4 | 102708997 | BANK1 | C | A | 0.570 | -0.015 | 0.002 | 4.4E-15 | 2.49E-04 | 62 |
| rs3936511 | 5 | 55860781 | AC022431.2 | A | G | 0.808 | 0.015 | 0.002 | 4E-10 | 1.59E-04 | 39 |
| rs10050620 | 5 | 63927239 | RGS7BP | C | T | 0.675 | 0.016 | 0.002 | 1.3E-14 | 2.41E-04 | 59 |
| rs13190020 | 5 | 65012526 | SGTB | G | A | 0.654 | -0.011 | 0.002 | 1.7E-08 | 1.29E-04 | 32 |
| rs9293494 | 5 | 87204121 | TMEM161B | T | G | 0.764 | 0.015 | 0.002 | 3.3E-11 | 1.78E-04 | 44 |
| rs4235642 | 5 | 103818412 | NUDT12 | A | G | 0.621 | 0.012 | 0.002 | 5.8E-10 | 1.56E-04 | 38 |
| rs6860760 | 5 | 142868379 | NR3C1 | A | G | 0.434 | 0.011 | 0.002 | 3.6E-08 | 1.23E-04 | 30 |
| rs815610 | 5 | 153517178 | MFAP3 | C | G | 0.443 | 0.015 | 0.002 | 4.7E-15 | 2.49E-04 | 61 |
| rs12214497 | 6 | 10015908 | OFCC1 | G | T | 0.656 | 0.014 | 0.002 | 2.7E-11 | 1.80E-04 | 44 |
| rs75782365 | 6 | 26408551 | BTN3A1 | T | G | 0.893 | -0.025 | 0.003 | 7.6E-16 | 2.63E-04 | 65 |
| rs34196306 | 6 | 27425644 | ZNF184 | G | C | 0.893 | -0.026 | 0.003 | 9.6E-17 | 2.80E-04 | 69 |
| rs3749971 | 6 | 29342775 | OR5V1 | G | A | 0.878 | -0.024 | 0.003 | 7.9E-16 | 2.63E-04 | 65 |
| rs3131934 | 6 | 30931844 | DPCR1 | T | C | 0.831 | -0.023 | 0.003 | 1.4E-18 | 3.14E-04 | 77 |
| rs9268235 | 6 | 32290208 | C6orf10 | C | T | 0.869 | -0.021 | 0.003 | 1.7E-13 | 2.20E-04 | 54 |
| rs141127771 | 6 | 32632200 | HLA-DQB1 | G | A | 0.741 | -0.016 | 0.003 | 1.8E-09 | 1.47E-04 | 36 |
| rs3798544 | 6 | 34520267 | SPDEF | G | A | 0.867 | -0.016 | 0.003 | 1.7E-08 | 1.29E-04 | 32 |
| rs2206277 | 6 | 50798526 | TFAP2B | C | T | 0.820 | -0.027 | 0.003 | 1.4E-26 | 4.61E-04 | 114 |
| rs1775255 | 6 | 51243035 | PKHD1 | G | T | 0.523 | -0.014 | 0.002 | 1.9E-12 | 2.01E-04 | 50 |
| rs12110721 | 6 | 55190480 | GFRAL | G | A | 0.825 | -0.019 | 0.003 | 1.5E-13 | 2.21E-04 | 55 |
| rs34260097 | 6 | 100727703 | SIM1 | T | G | 0.776 | -0.025 | 0.002 | 1.3E-27 | 4.80E-04 | 119 |
| rs7759938 | 6 | 105378954 | LIN28B | C | T | 0.322 | -0.012 | 0.002 | 2.2E-08 | 1.27E-04 | 31 |
| rs796915 | 6 | 154304628 | OPRM1 | C | G | 0.306 | -0.016 | 0.002 | 2.6E-14 | 2.35E-04 | 58 |
| rs62425122 | 6 | 166311987 | PDE10A | G | A | 0.703 | 0.012 | 0.002 | 4.5E-09 | 1.39E-04 | 34 |
| rs983949 | 7 | 24299013 | NPY | T | G | 0.715 | -0.013 | 0.002 | 1.3E-09 | 1.49E-04 | 37 |
| rs7808296 | 7 | 103127620 | RELN | C | T | 0.685 | -0.012 | 0.002 | 2E-08 | 1.28E-04 | 31 |
| rs6979832 | 7 | 127856276 | LEP | A | G | 0.503 | -0.011 | 0.002 | 5.7E-09 | 1.38E-04 | 34 |

| | | | | | | | | | | | |
|---|---|---|---|---|---|---|---|---|---|---|---|
| rs13233916 | 7 | 138874416 | TTC26 | C | G | 0.911 | 0.021 | 0.003 | 5.8E-10 | 1.56E-04 | 38 |
| rs7005216 | 8 | 8547110 | CLDN23 | G | C | 0.522 | 0.011 | 0.002 | 8.4E-09 | 1.35E-04 | 33 |
| rs351776 | 8 | 28191306 | PNOC | A | C | 0.452 | -0.012 | 0.002 | 2E-09 | 1.46E-04 | 36 |
| rs62515439 | 8 | 57165417 | CHCHD7 | C | T | 0.633 | 0.013 | 0.002 | 2.5E-10 | 1.62E-04 | 40 |
| rs13254613 | 8 | 64804804 | YTHDF3 | A | C | 0.654 | -0.017 | 0.002 | 3E-16 | 2.71E-04 | 67 |
| rs2126474 | 8 | 76878957 | HNF4G | G | T | 0.586 | 0.020 | 0.002 | 1.9E-23 | 4.04E-04 | 100 |
| rs10821163 | 9 | 96343060 | PHF2 | G | C | 0.659 | -0.018 | 0.002 | 2.8E-19 | 3.27E-04 | 81 |
| rs957512 | 9 | 120405705 | TLR4 | T | C | 0.669 | 0.012 | 0.002 | 2.2E-08 | 1.27E-04 | 31 |
| rs2275241 | 9 | 129370576 | LMX1B | G | A | 0.628 | -0.014 | 0.002 | 5.5E-12 | 1.93E-04 | 48 |
| rs7084503 | 10 | 2666859 | PFKP | T | C | 0.492 | 0.015 | 0.002 | 2E-15 | 2.56E-04 | 63 |
| rs76971642 | 10 | 87575243 | GRID1 | T | C | 0.953 | -0.031 | 0.005 | 1.6E-11 | 1.84E-04 | 45 |
| rs962369 | 11 | 27734420 | BDNF | T | C | 0.694 | -0.015 | 0.002 | 3.5E-13 | 2.15E-04 | 53 |
| rs661878 | 11 | 29188691 | METTL15 | A | G | 0.867 | 0.019 | 0.003 | 3.4E-11 | 1.78E-04 | 44 |
| rs11039307 | 11 | 47611152 | C1QTNF4 | C | T | 0.591 | -0.016 | 0.002 | 1.7E-15 | 2.57E-04 | 63 |
| rs678653 | 11 | 69466737 | CCND1 | C | G | 0.364 | -0.012 | 0.002 | 1.3E-09 | 1.49E-04 | 37 |
| rs11215403 | 11 | 115058585 | CADM1 | G | A | 0.757 | 0.016 | 0.002 | 7.6E-13 | 2.08E-04 | 51 |
| rs11611246 | 12 | 939480 | WNK1 | G | T | 0.787 | -0.017 | 0.002 | 2.3E-12 | 2.00E-04 | 49 |
| rs2187642 | 12 | 11855624 | ETV6 | A | C | 0.376 | -0.012 | 0.002 | 2.6E-09 | 1.44E-04 | 35 |
| rs10876457 | 12 | 39453689 | CPNE8 | G | A | 0.775 | 0.013 | 0.002 | 3.2E-08 | 1.24E-04 | 31 |
| rs10783302 | 12 | 49498130 | LMBR1L | G | T | 0.635 | -0.013 | 0.002 | 4.7E-11 | 1.76E-04 | 43 |
| rs7132908 | 12 | 50263148 | FAIM2 | G | A | 0.616 | -0.031 | 0.002 | 8E-56 | 1.00E-03 | 248 |
| rs76919525 | 12 | 50541733 | CERS5 | T | A | 0.342 | -0.011 | 0.002 | 4.6E-08 | 1.21E-04 | 30 |
| rs78607331 | 12 | 57648644 | R3HDM2 | C | T | 0.955 | -0.029 | 0.005 | 6.2E-10 | 1.55E-04 | 38 |
| rs10784514 | 12 | 66452879 | LLPH | C | T | 0.320 | -0.012 | 0.002 | 2.4E-08 | 1.26E-04 | 31 |
| rs2364232 | 12 | 93994827 | SOCS2 | A | C | 0.741 | 0.012 | 0.002 | 2.8E-08 | 1.25E-04 | 31 |
| rs12309017 | 12 | 99677307 | ANKS1B | G | T | 0.747 | -0.012 | 0.002 | 2.9E-08 | 1.25E-04 | 31 |
| rs11111647 | 12 | 103945224 | STAB2 | G | A | 0.792 | 0.013 | 0.002 | 4.6E-08 | 1.21E-04 | 30 |
| rs7305424 | 12 | 118399491 | KSR2 | A | T | 0.658 | -0.012 | 0.002 | 3.9E-09 | 1.41E-04 | 35 |
| rs35202265 | 13 | 27986069 | GTF3A | C | T | 0.378 | -0.012 | 0.002 | 7.1E-09 | 1.36E-04 | 33 |
| rs9551428 | 13 | 28618462 | FLT3 | C | T | 0.374 | 0.015 | 0.002 | 2.1E-14 | 2.37E-04 | 58 |
| rs1336486 | 13 | 40784814 | AL133318.1 | T | G | 0.671 | -0.015 | 0.002 | 9.3E-13 | 2.07E-04 | 51 |
| rs4477562 | 13 | 54104968 | OLFM4 | C | T | 0.872 | -0.017 | 0.003 | 3.7E-09 | 1.41E-04 | 35 |
| rs9317002 | 13 | 59175727 | PCDH17 | C | A | 0.485 | -0.014 | 0.002 | 1.2E-12 | 2.04E-04 | 50 |
| rs58681688 | 13 | 62467001 | PCDH20 | C | G | 0.813 | -0.014 | 0.002 | 4.7E-08 | 1.21E-04 | 30 |
| rs9540493 | 13 | 66205704 | PCDH9 | A | G | 0.454 | 0.011 | 0.002 | 2E-08 | 1.28E-04 | 31 |
| rs1576655 | 13 | 79587841 | RBM26 | A | C | 0.404 | -0.012 | 0.002 | 5.8E-09 | 1.37E-04 | 34 |
| rs61980008 | 14 | 30464716 | PRKD1 | G | A | 0.961 | -0.037 | 0.005 | 2.9E-13 | 2.16E-04 | 53 |
| rs1865719 | 14 | 79923667 | NRXN3 | A | G | 0.367 | -0.013 | 0.002 | 4.5E-10 | 1.58E-04 | 39 |
| rs8030456 | 15 | 68076856 | MAP2K5 | C | T | 0.773 | 0.023 | 0.002 | 4.2E-24 | 4.16E-04 | 103 |
| rs4932430 | 15 | 89363866 | ACAN | A | C | 0.521 | 0.012 | 0.002 | 1.5E-09 | 1.48E-04 | 37 |
| rs2970356 | 15 | 90623540 | ZNF710 | C | G | 0.679 | -0.013 | 0.002 | 1.2E-09 | 1.50E-04 | 37 |
| rs72755233 | 15 | 100692953 | ADAMTS17 | G | A | 0.888 | -0.018 | 0.003 | 7.2E-09 | 1.36E-04 | 33 |
| rs2531991 | 16 | 4023553 | ADCY9 | G | A | 0.253 | -0.020 | 0.002 | 2.1E-19 | 3.29E-04 | 81 |
| rs148965598 | 16 | 19975731 | GPR139 | A | G | 0.861 | 0.032 | 0.003 | 3.1E-30 | 5.29E-04 | 131 |
| rs4432271 | 16 | 20245283 | GP2 | C | T | 0.131 | -0.022 | 0.003 | 1.4E-13 | 2.22E-04 | 55 |

| | | | | | | | | | | | |
|---|---|---|---|---|---|---|---|---|---|---|---|
| rs7189927 | 16 | 28913787 | ATP2A1 | T | C | 0.352 | 0.016 | 0.002 | 2.1E-15 | 2.56E-04 | 63 |
| rs4889630 | 16 | 30877544 | BCL7C | T | C | 0.200 | 0.016 | 0.002 | 2.1E-11 | 1.82E-04 | 45 |
| rs1421085 | 16 | 53800954 | FTO | T | C | 0.599 | -0.049 | 0.002 | 1.9E-133 | 2.45E-03 | 604 |
| rs11863799 | 16 | 61933401 | CDH8 | C | T | 0.675 | 0.011 | 0.002 | 3.4E-08 | 1.23E-04 | 30 |
| rs34229857 | 16 | 67434917 | ZDHHC1 | C | T | 0.971 | -0.045 | 0.006 | 1E-14 | 2.43E-04 | 60 |
| rs11642090 | 16 | 81730582 | CMIP | T | C | 0.627 | -0.013 | 0.002 | 3.4E-11 | 1.78E-04 | 44 |
| rs242922 | 17 | 43946370 | SPPL2C | A | C | 0.406 | -0.011 | 0.002 | 3.6E-08 | 1.23E-04 | 30 |
| rs999493 | 17 | 46625519 | HOXB3 | G | A | 0.378 | -0.011 | 0.002 | 2.2E-08 | 1.27E-04 | 31 |
| rs12185242 | 17 | 47407071 | ZNF652 | A | C | 0.544 | -0.011 | 0.002 | 1.5E-08 | 1.30E-04 | 32 |
| rs11150745 | 17 | 78757626 | RPTOR | A | G | 0.682 | 0.012 | 0.002 | 7.3E-09 | 1.36E-04 | 33 |
| rs1013737 | 18 | 937050 | ADCYAP1 | G | C | 0.507 | -0.011 | 0.002 | 2.2E-08 | 1.27E-04 | 31 |
| rs303753 | 18 | 21074922 | RIOK3 | G | A | 0.655 | -0.012 | 0.002 | 1.9E-08 | 1.28E-04 | 32 |
| rs7239114 | 18 | 45921214 | ZBTB7C | G | A | 0.458 | -0.014 | 0.002 | 5E-13 | 2.12E-04 | 52 |
| rs12606230 | 18 | 52492252 | RAB27B | T | C | 0.767 | -0.013 | 0.002 | 3.5E-08 | 1.23E-04 | 30 |
| rs2168711 | 18 | 57848531 | MC4R | T | C | 0.767 | -0.035 | 0.002 | 3.6E-54 | 9.73E-04 | 240 |
| rs17066856 | 18 | 58049656 | MC4R | T | C | 0.909 | 0.028 | 0.003 | 6.8E-17 | 2.83E-04 | 70 |
| rs16982345 | 19 | 18500722 | GDF15 | G | A | 0.750 | -0.012 | 0.002 | 2.7E-08 | 1.25E-04 | 31 |
| rs3810304 | 19 | 30861683 | ZNF536 | A | G | 0.225 | 0.014 | 0.002 | 6.7E-10 | 1.55E-04 | 38 |
| rs4805881 | 19 | 33896432 | PEPD | A | C | 0.334 | -0.012 | 0.002 | 1.2E-08 | 1.32E-04 | 32 |
| rs3810291 | 19 | 47569003 | ZC3H4 | G | A | 0.324 | -0.015 | 0.002 | 1E-12 | 2.06E-04 | 51 |
| rs633372 | 19 | 49209226 | FUT2 | G | A | 0.466 | 0.011 | 0.002 | 1.1E-08 | 1.33E-04 | 33 |
| rs994308 | 20 | 6603622 | BMP2 | C | T | 0.596 | -0.012 | 0.002 | 3.8E-10 | 1.59E-04 | 39 |
| rs7268466 | 20 | 15810676 | MACROD2 | C | T | 0.856 | -0.020 | 0.003 | 4.9E-13 | 2.12E-04 | 52 |
| rs947088 | 20 | 17171373 | PCSK2 | G | T | 0.282 | -0.013 | 0.002 | 6.7E-09 | 1.36E-04 | 34 |
| rs66469746 | 20 | 54374063 | CBLN4 | C | A | 0.714 | 0.014 | 0.002 | 2.6E-11 | 1.80E-04 | 44 |
| rs4817973 | 21 | 40309592 | ETS2 | G | A | 0.640 | 0.014 | 0.002 | 1.8E-11 | 1.83E-04 | 45 |
| rs6001872 | 22 | 40703245 | TNRC6B | A | G | 0.651 | 0.014 | 0.002 | 8.3E-12 | 1.89E-04 | 47 |

Genome-wide significant SNPs for adult body size in UK Biobank (overall)

| SNP | Chr | Position | Closest gene | EA | OA | EAF | Beta | SE | pvalue | $R^2$ | F statistic |
|---|---|---|---|---|---|---|---|---|---|---|---|
| rs4648450 | 1 | 2723214 | TTC34 | C | A | 0.533 | 0.010 | 0.001 | 1.2E-12 | 1.11E-04 | 50 |
| rs2076363 | 1 | 6684906 | THAP3 | C | G | 0.663 | -0.008 | 0.001 | 3.1E-08 | 6.76E-05 | 31 |
| rs4908677 | 1 | 7738180 | CAMTA1 | C | T | 0.520 | -0.008 | 0.001 | 2.7E-08 | 6.83E-05 | 31 |
| rs78886584 | 1 | 16859325 | FAM231B | A | G | 0.509 | -0.008 | 0.001 | 8.7E-10 | 8.30E-05 | 38 |
| rs10799778 | 1 | 23313353 | LACTBL1 | T | G | 0.166 | 0.013 | 0.002 | 7.8E-12 | 1.03E-04 | 47 |
| rs7511698 | 1 | 25015638 | SRRM1 | C | T | 0.694 | 0.008 | 0.001 | 3.2E-08 | 6.74E-05 | 31 |
| rs945211 | 1 | 32191798 | BAI2 | G | C | 0.384 | -0.008 | 0.001 | 1.6E-09 | 8.03E-05 | 36 |
| rs3737992 | 1 | 33234128 | KIAA1522 | G | A | 0.831 | 0.014 | 0.002 | 5.3E-14 | 1.25E-04 | 57 |
| rs12031634 | 1 | 34584393 | CSMD2 | G | A | 0.703 | 0.009 | 0.002 | 4.8E-09 | 7.56E-05 | 34 |
| rs116195355 | 1 | 39941508 | MACF1 | C | A | 0.969 | 0.026 | 0.004 | 5.9E-11 | 9.46E-05 | 43 |
| rs2744801 | 1 | 41155486 | NFYC | C | T | 0.663 | 0.008 | 0.001 | 4.3E-08 | 6.62E-05 | 30 |
| rs4660586 | 1 | 42407229 | HIVEP3 | C | T | 0.261 | 0.009 | 0.002 | 1E-08 | 7.25E-05 | 33 |
| rs6669341 | 1 | 47678458 | TAL1 | A | G | 0.417 | 0.011 | 0.001 | 1.3E-14 | 1.31E-04 | 59 |
| rs1167311 | 1 | 49996959 | AGBL4 | G | A | 0.319 | 0.012 | 0.001 | 1.2E-16 | 1.51E-04 | 69 |
| rs630602 | 1 | 54728864 | SSBP3 | G | C | 0.393 | -0.008 | 0.001 | 4.1E-08 | 6.64E-05 | 30 |

| | | | | | | | | | | | |
|---|---|---|---|---|---|---|---|---|---|---|---|
| rs12140153 | 1 | 62579891 | INADL | G | T | 0.906 | 0.021 | 0.002 | 4.2E-19 | 1.76E-04 | 80 |
| rs11208659 | 1 | 65979280 | LEPR | T | C | 0.917 | -0.014 | 0.002 | 2.1E-08 | 6.93E-05 | 31 |
| rs7519259 | 1 | 66434743 | PDE4B | G | A | 0.472 | -0.009 | 0.001 | 3.2E-11 | 9.73E-05 | 44 |
| rs2613499 | 1 | 72751552 | NEGR1 | A | G | 0.809 | 0.019 | 0.002 | 1.2E-26 | 2.52E-04 | 114 |
| rs7553158 | 1 | 75005238 | TNNI3K | G | A | 0.438 | 0.011 | 0.001 | 2E-14 | 1.29E-04 | 58 |
| rs115778101 | 1 | 78198554 | USP33 | T | C | 0.952 | 0.018 | 0.003 | 3.4E-08 | 6.72E-05 | 30 |
| rs34517439 | 1 | 78450517 | DNAJB4 | C | A | 0.878 | -0.025 | 0.002 | 1.9E-31 | 3.00E-04 | 136 |
| rs651533 | 1 | 82375561 | LPHN2 | T | A | 0.244 | 0.009 | 0.002 | 5.6E-09 | 7.50E-05 | 34 |
| rs28726372 | 1 | 84353839 | TTLL7 | T | C | 0.693 | -0.009 | 0.001 | 9.4E-10 | 8.26E-05 | 37 |
| rs7548936 | 1 | 91207757 | BARHL2 | G | C | 0.627 | -0.008 | 0.001 | 2.1E-09 | 7.92E-05 | 36 |
| rs6679458 | 1 | 96946253 | PTBP2 | G | T | 0.409 | -0.012 | 0.001 | 9.1E-17 | 1.53E-04 | 69 |
| rs12072739 | 1 | 98315893 | DPYD | A | G | 0.776 | -0.012 | 0.002 | 1.8E-12 | 1.10E-04 | 50 |
| rs41279738 | 1 | 110082551 | GPR61 | T | G | 0.974 | -0.043 | 0.004 | 3.9E-23 | 2.16E-04 | 98 |
| rs12033257 | 1 | 112318484 | KCND3 | A | G | 0.618 | 0.010 | 0.001 | 4.9E-12 | 1.05E-04 | 48 |
| rs7549358 | 1 | 115252609 | NRAS | G | C | 0.356 | 0.009 | 0.001 | 6.5E-10 | 8.42E-05 | 38 |
| rs1409158 | 1 | 119538890 | TBX15 | C | T | 0.237 | 0.009 | 0.002 | 1E-08 | 7.25E-05 | 33 |
| rs142315514 | 1 | 147050816 | BCL9 | C | A | 0.965 | -0.021 | 0.004 | 1.7E-08 | 7.03E-05 | 32 |
| rs10749659 | 1 | 151033979 | MLLT11 | C | T | 0.228 | 0.010 | 0.002 | 4.4E-10 | 8.59E-05 | 39 |
| rs3753639 | 1 | 154986091 | ZBTB7B | T | C | 0.756 | -0.011 | 0.002 | 7.3E-13 | 1.14E-04 | 51 |
| rs61813324 | 1 | 156049877 | MEX3A | C | T | 0.864 | -0.018 | 0.002 | 1.8E-19 | 1.80E-04 | 81 |
| rs1778830 | 1 | 156489974 | IQGAP3 | G | A | 0.638 | -0.010 | 0.001 | 3.8E-12 | 1.06E-04 | 48 |
| rs4916229 | 1 | 171443368 | PRRC2C | C | G | 0.904 | -0.015 | 0.002 | 8.9E-11 | 9.28E-05 | 42 |
| rs148137538 | 1 | 173399677 | PRDX6 | A | G | 0.977 | 0.025 | 0.005 | 4.4E-08 | 6.61E-05 | 30 |
| rs77560793 | 1 | 175001179 | MRPS14 | G | A | 0.969 | 0.025 | 0.004 | 7.8E-10 | 8.34E-05 | 38 |
| rs539515 | 1 | 177889025 | SEC16B | A | C | 0.795 | -0.030 | 0.002 | 8.4E-71 | 6.98E-04 | 316 |
| rs9425634 | 1 | 184663581 | EDEM3 | T | C | 0.542 | -0.008 | 0.001 | 5.7E-09 | 7.49E-05 | 34 |
| rs815163 | 1 | 190294726 | BRINP3 | T | C | 0.437 | 0.011 | 0.001 | 6.1E-15 | 1.34E-04 | 61 |
| rs76702514 | 1 | 195148296 | KCNT2 | C | G | 0.789 | 0.010 | 0.002 | 2E-09 | 7.95E-05 | 36 |
| rs2678204 | 1 | 201800511 | IPO9 | T | G | 0.660 | -0.015 | 0.001 | 2.7E-25 | 2.38E-04 | 108 |
| rs4971239 | 1 | 203491150 | OPTC | G | A | 0.831 | -0.011 | 0.002 | 4.5E-10 | 8.58E-05 | 39 |
| rs7539903 | 1 | 209208033 | CAMK1G | T | A | 0.385 | 0.008 | 0.001 | 4.6E-08 | 6.59E-05 | 30 |
| rs78508049 | 1 | 210344884 | SYT14 | T | C | 0.812 | -0.011 | 0.002 | 2.1E-10 | 8.91E-05 | 40 |
| rs12037905 | 1 | 219628036 | SLC30A10 | C | T | 0.581 | 0.008 | 0.001 | 1.1E-08 | 7.20E-05 | 33 |
| rs7518221 | 1 | 225561346 | DNAH14 | T | C | 0.353 | 0.008 | 0.001 | 2.8E-08 | 6.80E-05 | 31 |
| rs10779835 | 1 | 230299949 | GALNT2 | T | C | 0.387 | 0.008 | 0.001 | 1.2E-08 | 7.16E-05 | 32 |
| rs10927006 | 1 | 243557659 | SDCCAG8 | T | C | 0.856 | 0.012 | 0.002 | 8.3E-10 | 8.32E-05 | 38 |
| rs4658403 | 1 | 243832560 | AKT3 | C | T | 0.166 | 0.014 | 0.002 | 1.2E-13 | 1.21E-04 | 55 |
| rs6752378 | 2 | 25150116 | ADCY3 | C | A | 0.514 | -0.020 | 0.001 | 1.8E-48 | 4.72E-04 | 214 |
| rs1631026 | 2 | 26953850 | KCNK3 | C | T | 0.527 | -0.011 | 0.001 | 5.9E-15 | 1.34E-04 | 61 |
| rs10204994 | 2 | 35443726 | CRIM1 | G | A | 0.771 | 0.010 | 0.002 | 5.3E-10 | 8.51E-05 | 39 |
| rs10185199 | 2 | 40282202 | SLC8A1 | G | A | 0.719 | 0.010 | 0.002 | 1.6E-10 | 9.01E-05 | 41 |
| rs10169594 | 2 | 41637688 | C2orf91 | T | C | 0.637 | -0.008 | 0.001 | 3.3E-08 | 6.74E-05 | 31 |
| rs35809007 | 2 | 47019521 | LINC01118 | G | A | 0.637 | 0.011 | 0.001 | 1.2E-13 | 1.22E-04 | 55 |
| rs72618637 | 2 | 48953979 | GTF2A1L | T | A | 0.811 | 0.010 | 0.002 | 1.8E-08 | 6.99E-05 | 32 |
| rs6761463 | 2 | 50201547 | NRXN1 | G | C | 0.162 | 0.013 | 0.002 | 1.3E-11 | 1.01E-04 | 46 |

| | | | | | | | | | | | |
|---|---|---|---|---|---|---|---|---|---|---|---|
| rs59428052 | 2 | 53861389 | GPR75-ASB3 | A | G | 0.851 | 0.011 | 0.002 | 9.8E-09 | 7.25E-05 | 33 |
| rs7601895 | 2 | 55281901 | RTN4 | C | G | 0.692 | 0.010 | 0.001 | 4.1E-12 | 1.06E-04 | 48 |
| rs4671328 | 2 | 58935282 | FANCL | T | G | 0.449 | 0.013 | 0.001 | 2.1E-21 | 1.99E-04 | 90 |
| rs4672338 | 2 | 60217457 | BCL11A | C | T | 0.664 | -0.008 | 0.001 | 8E-09 | 7.34E-05 | 33 |
| rs12477088 | 2 | 67841326 | ETAA1 | T | C | 0.590 | 0.010 | 0.001 | 4.7E-14 | 1.25E-04 | 57 |
| rs6752979 | 2 | 81741750 | CTNNA2 | G | A | 0.683 | -0.009 | 0.001 | 1.1E-09 | 8.19E-05 | 37 |
| rs396354 | 2 | 86850022 | CHMP3 | T | C | 0.284 | 0.010 | 0.002 | 8E-12 | 1.03E-04 | 47 |
| rs11691869 | 2 | 100805996 | AFF3 | C | A | 0.638 | 0.011 | 0.001 | 3.5E-15 | 1.37E-04 | 62 |
| rs1451533 | 2 | 105466005 | POU3F3 | G | A | 0.726 | -0.011 | 0.002 | 1.3E-12 | 1.11E-04 | 50 |
| rs72851476 | 2 | 142874787 | LRP1B | A | C | 0.844 | 0.011 | 0.002 | 2.8E-08 | 6.80E-05 | 31 |
| rs62171698 | 2 | 143959096 | ARHGAP15 | C | A | 0.859 | -0.012 | 0.002 | 8.8E-10 | 8.29E-05 | 38 |
| rs75706763 | 2 | 145669168 | ZEB2 | A | G | 0.948 | -0.018 | 0.003 | 1.7E-08 | 7.03E-05 | 32 |
| rs409696 | 2 | 147900651 | ACVR2A | G | A | 0.424 | 0.011 | 0.001 | 1.8E-16 | 1.50E-04 | 68 |
| rs12692596 | 2 | 161265910 | RBMS1 | C | T | 0.628 | -0.008 | 0.001 | 8.1E-09 | 7.34E-05 | 33 |
| rs12477385 | 2 | 166144850 | SCN2A | G | T | 0.775 | 0.009 | 0.002 | 1.8E-08 | 7.00E-05 | 32 |
| rs788163 | 2 | 172931559 | METAP1D | A | C | 0.724 | -0.010 | 0.002 | 5.9E-11 | 9.45E-05 | 43 |
| rs72917533 | 2 | 175238924 | CIR1 | T | C | 0.814 | 0.011 | 0.002 | 8.2E-11 | 9.31E-05 | 42 |
| rs7570446 | 2 | 193801010 | TMEFF2 | C | A | 0.456 | -0.008 | 0.001 | 1.3E-08 | 7.13E-05 | 32 |
| rs1704190 | 2 | 200760629 | C2orf69 | G | A | 0.364 | -0.008 | 0.001 | 4.5E-09 | 7.59E-05 | 34 |
| rs4482463 | 2 | 205375909 | PARD3B | C | A | 0.077 | 0.019 | 0.003 | 2.3E-13 | 1.19E-04 | 54 |
| rs79675564 | 2 | 211286896 | LANCL1 | C | A | 0.922 | -0.015 | 0.003 | 3.4E-09 | 7.70E-05 | 35 |
| rs13427822 | 2 | 213414265 | ERBB4 | A | G | 0.729 | 0.009 | 0.002 | 1.3E-09 | 8.14E-05 | 37 |
| rs55658481 | 2 | 219284215 | VIL1 | G | A | 0.661 | -0.009 | 0.001 | 5.3E-10 | 8.51E-05 | 39 |
| rs2433733 | 2 | 230816703 | FBXO36 | G | A | 0.322 | 0.011 | 0.001 | 7.3E-13 | 1.14E-04 | 51 |
| rs4663213 | 2 | 236807893 | AGAP1 | G | A | 0.222 | 0.009 | 0.002 | 1.2E-08 | 7.17E-05 | 33 |
| rs112380819 | 3 | 9498519 | SETD5 | G | A | 0.898 | -0.014 | 0.002 | 1.3E-09 | 8.11E-05 | 37 |
| rs34373881 | 3 | 20432033 | SGOL1 | G | A | 0.725 | 0.009 | 0.002 | 9E-09 | 7.29E-05 | 33 |
| rs7619139 | 3 | 25110415 | RARB | T | A | 0.411 | -0.009 | 0.001 | 2.4E-10 | 8.86E-05 | 40 |
| rs80082536 | 3 | 35195311 | ARPP21 | A | G | 0.880 | -0.013 | 0.002 | 1.3E-09 | 8.13E-05 | 37 |
| rs111768603 | 3 | 42329113 | CCK | G | T | 0.890 | 0.016 | 0.002 | 3.8E-13 | 1.16E-04 | 53 |
| rs28350 | 3 | 42418446 | LYZL4 | A | G | 0.179 | 0.013 | 0.002 | 1.7E-12 | 1.10E-04 | 50 |
| rs78517245 | 3 | 42587865 | SEC22C | T | C | 0.985 | -0.034 | 0.006 | 4.1E-09 | 7.63E-05 | 35 |
| rs113706999 | 3 | 44159156 | TOPAZ1 | T | A | 0.977 | -0.029 | 0.005 | 1.1E-09 | 8.18E-05 | 37 |
| rs9852062 | 3 | 45373442 | LARS2 | T | A | 0.443 | 0.008 | 0.001 | 3.5E-09 | 7.70E-05 | 35 |
| rs113569731 | 3 | 47093206 | SETD2 | C | A | 0.906 | -0.015 | 0.002 | 3.1E-10 | 8.73E-05 | 40 |
| rs9843653 | 3 | 49920571 | MST1R | T | C | 0.488 | -0.018 | 0.001 | 1.5E-37 | 3.62E-04 | 164 |
| rs62259692 | 3 | 51847709 | IQCF3 | G | A | 0.939 | -0.018 | 0.003 | 1.4E-09 | 8.10E-05 | 37 |
| rs6798941 | 3 | 52893465 | TMEM110 | C | T | 0.704 | -0.011 | 0.002 | 2.7E-13 | 1.18E-04 | 53 |
| rs6445198 | 3 | 61219865 | FHIT | G | T | 0.584 | 0.011 | 0.001 | 3.5E-15 | 1.37E-04 | 62 |
| rs6445258 | 3 | 62112198 | PTPRG | T | C | 0.208 | 0.010 | 0.002 | 8.6E-09 | 7.31E-05 | 33 |
| rs76824303 | 3 | 62459819 | CADPS | A | C | 0.900 | 0.015 | 0.002 | 9.2E-11 | 9.26E-05 | 42 |
| rs557951 | 3 | 62713263 | CADPS | T | G | 0.687 | -0.008 | 0.001 | 3.9E-08 | 6.67E-05 | 30 |
| rs11708540 | 3 | 70593081 | FOXP1 | G | A | 0.842 | -0.011 | 0.002 | 2E-08 | 6.96E-05 | 32 |
| rs1598121 | 3 | 82694710 | GBE1 | A | G | 0.637 | -0.009 | 0.001 | 3E-10 | 8.76E-05 | 40 |
| rs114593013 | 3 | 84113491 | CADM2 | A | G | 0.942 | 0.020 | 0.003 | 2.1E-11 | 9.89E-05 | 45 |

| | | | | | | | | | | | |
|---|---|---|---|---|---|---|---|---|---|---|---|
| rs11915747 | 3 | 85699040 | CADM2 | C | G | 0.647 | 0.011 | 0.001 | 5E-14 | 1.25E-04 | 57 |
| rs4858940 | 3 | 88254820 | C3orf38 | T | C | 0.114 | -0.013 | 0.002 | 6E-10 | 8.45E-05 | 38 |
| rs1454687 | 3 | 94038085 | NSUN3 | C | G | 0.485 | 0.012 | 0.001 | 3E-19 | 1.77E-04 | 80 |
| rs1436348 | 3 | 104612668 | ALCAM | A | G | 0.417 | -0.010 | 0.001 | 3.5E-12 | 1.07E-04 | 48 |
| rs36131051 | 3 | 107888841 | IFT57 | T | G | 0.795 | 0.011 | 0.002 | 2.9E-10 | 8.78E-05 | 40 |
| rs9814758 | 3 | 123062657 | ADCY5 | T | G | 0.644 | 0.008 | 0.001 | 5.4E-09 | 7.51E-05 | 34 |
| rs1320903 | 3 | 131758077 | CPNE4 | G | A | 0.680 | -0.015 | 0.001 | 2.6E-23 | 2.18E-04 | 99 |
| rs10935143 | 3 | 134665159 | EPHB1 | G | A | 0.551 | 0.008 | 0.001 | 1.1E-09 | 8.18E-05 | 37 |
| rs2343681 | 3 | 136535024 | SLC35G2 | G | A | 0.211 | -0.012 | 0.002 | 1.8E-13 | 1.20E-04 | 54 |
| rs2035936 | 3 | 141298124 | RASA2 | G | T | 0.944 | -0.022 | 0.003 | 2.3E-13 | 1.19E-04 | 54 |
| rs12634936 | 3 | 147716498 | ZIC1 | T | C | 0.943 | -0.018 | 0.003 | 8.8E-09 | 7.30E-05 | 33 |
| rs1568488 | 3 | 153657951 | ARHGEF26 | G | C | 0.405 | -0.011 | 0.001 | 4.4E-15 | 1.36E-04 | 61 |
| rs9834519 | 3 | 156379637 | TIPARP | C | T | 0.919 | 0.016 | 0.003 | 8.4E-10 | 8.31E-05 | 38 |
| rs12630209 | 3 | 156881392 | CCNL1 | T | G | 0.747 | -0.009 | 0.002 | 3E-08 | 6.77E-05 | 31 |
| rs8192675 | 3 | 170724883 | SLC2A2 | T | C | 0.711 | -0.012 | 0.002 | 8.7E-15 | 1.33E-04 | 60 |
| rs529200 | 3 | 173114305 | NLGN1 | A | G | 0.472 | -0.010 | 0.001 | 5.8E-14 | 1.25E-04 | 56 |
| rs13061117 | 3 | 181186466 | SOX2 | T | C | 0.910 | -0.015 | 0.002 | 1.3E-09 | 8.13E-05 | 37 |
| rs262956 | 3 | 183486117 | YEATS2 | T | G | 0.353 | 0.009 | 0.001 | 2.2E-10 | 8.89E-05 | 40 |
| rs869400 | 3 | 185826740 | ETV5 | T | G | 0.185 | -0.018 | 0.002 | 3.5E-25 | 2.37E-04 | 107 |
| rs80236973 | 3 | 188001014 | LPP | C | T | 0.865 | 0.012 | 0.002 | 1.4E-09 | 8.09E-05 | 37 |
| rs4677813 | 3 | 194863860 | XXYLT1 | T | C | 0.750 | 0.009 | 0.002 | 1.6E-08 | 7.04E-05 | 32 |
| rs6583310 | 3 | 196170985 | UBXN7 | G | C | 0.561 | -0.009 | 0.001 | 2.2E-10 | 8.89E-05 | 40 |
| rs2051559 | 4 | 3298800 | RGS12 | T | C | 0.868 | -0.014 | 0.002 | 8.3E-12 | 1.03E-04 | 47 |
| rs35852935 | 4 | 17991522 | LCORL | A | C | 0.964 | -0.022 | 0.004 | 4.9E-09 | 7.55E-05 | 34 |
| rs1477890 | 4 | 18511738 | LCORL | A | G | 0.507 | -0.010 | 0.001 | 1.1E-12 | 1.12E-04 | 51 |
| rs34811474 | 4 | 25408838 | ANAPC4 | G | A | 0.769 | 0.017 | 0.002 | 2E-24 | 2.30E-04 | 104 |
| rs73213484 | 4 | 28489339 | RP11-180C1.1 | A | T | 0.859 | 0.015 | 0.002 | 1.1E-13 | 1.22E-04 | 55 |
| rs4527444 | 4 | 30842780 | PCDH7 | A | G | 0.459 | -0.009 | 0.001 | 9E-12 | 1.03E-04 | 47 |
| rs36023504 | 4 | 38698924 | KLF3 | C | T | 0.617 | 0.008 | 0.001 | 4.6E-08 | 6.59E-05 | 30 |
| rs12507026 | 4 | 45181334 | GNPDA2 | A | T | 0.565 | -0.019 | 0.001 | 1.8E-41 | 4.01E-04 | 182 |
| rs2237025 | 4 | 55541879 | KIT | T | C | 0.445 | 0.010 | 0.001 | 1.8E-13 | 1.20E-04 | 54 |
| rs28462076 | 4 | 65696174 | TECRL | A | G | 0.762 | 0.009 | 0.002 | 6.7E-09 | 7.42E-05 | 34 |
| rs2164300 | 4 | 67813017 | CENPC | C | T | 0.481 | 0.008 | 0.001 | 4.2E-08 | 6.64E-05 | 30 |
| rs13104584 | 4 | 80811227 | ANTXR2 | G | A | 0.592 | -0.009 | 0.001 | 6.2E-10 | 8.44E-05 | 38 |
| rs4148155 | 4 | 89054667 | ABCG2 | A | G | 0.887 | 0.014 | 0.002 | 2.9E-11 | 9.76E-05 | 44 |
| rs4419475 | 4 | 96150044 | UNC5C | A | T | 0.593 | -0.008 | 0.001 | 6.5E-09 | 7.43E-05 | 34 |
| rs1229984 | 4 | 100239319 | ADH1B | T | C | 0.027 | -0.023 | 0.004 | 3.2E-08 | 6.75E-05 | 31 |
| rs2583410 | 4 | 102182199 | PPP3CA | A | C | 0.853 | -0.013 | 0.002 | 5.3E-12 | 1.05E-04 | 48 |
| rs13107325 | 4 | 103188709 | SLC39A8 | C | T | 0.925 | -0.029 | 0.003 | 3.1E-28 | 2.68E-04 | 121 |
| rs1381010 | 4 | 112677085 | C4orf32 | G | A | 0.695 | 0.008 | 0.001 | 3E-08 | 6.77E-05 | 31 |
| rs2952863 | 4 | 130759647 | C4orf33 | T | G | 0.291 | 0.010 | 0.002 | 1.6E-10 | 9.02E-05 | 41 |
| rs1296328 | 4 | 137083193 | PCDH18 | A | C | 0.441 | 0.012 | 0.001 | 2.6E-17 | 1.58E-04 | 72 |
| rs809955 | 4 | 140874760 | MAML3 | G | A | 0.634 | 0.010 | 0.001 | 1.4E-12 | 1.11E-04 | 50 |
| rs35390852 | 4 | 143067054 | INPP4B | G | A | 0.876 | -0.011 | 0.002 | 4.8E-08 | 6.57E-05 | 30 |
| rs12644329 | 4 | 143634746 | INPP4B | G | A | 0.372 | 0.008 | 0.001 | 2.5E-08 | 6.86E-05 | 31 |

| | | | | | | | | | | | |
|---|---|---|---|---|---|---|---|---|---|---|---|
| rs6823268 | 4 | 145982563 | ANAPC10 | A | G | 0.629 | -0.008 | 0.001 | 8.2E-09 | 7.33E-05 | 33 |
| rs113079574 | 4 | 147354089 | SLC10A7 | C | T | 0.807 | 0.010 | 0.002 | 3.3E-08 | 6.74E-05 | 31 |
| rs6843852 | 4 | 162132758 | FSTL5 | C | T | 0.492 | -0.009 | 0.001 | 2E-10 | 8.92E-05 | 40 |
| rs698147 | 5 | 3513485 | IRX1 | A | G | 0.456 | 0.009 | 0.001 | 1.8E-10 | 8.97E-05 | 41 |
| rs67913249 | 5 | 43204126 | NIM1K | C | G | 0.658 | 0.009 | 0.001 | 1.2E-10 | 9.17E-05 | 42 |
| rs114263339 | 5 | 50932343 | ISL1 | C | T | 0.974 | -0.025 | 0.004 | 5.2E-09 | 7.53E-05 | 34 |
| rs10805383 | 5 | 63034606 | HTR1A | G | A | 0.519 | -0.010 | 0.001 | 2.6E-14 | 1.28E-04 | 58 |
| rs9291822 | 5 | 64076515 | CWC27 | C | T | 0.485 | 0.008 | 0.001 | 1.1E-08 | 7.20E-05 | 33 |
| rs27215 | 5 | 66219318 | MAST4 | C | A | 0.275 | -0.010 | 0.002 | 5.1E-11 | 9.52E-05 | 43 |
| rs2307111 | 5 | 75003678 | POC5 | T | C | 0.605 | 0.017 | 0.001 | 6.9E-33 | 3.15E-04 | 143 |
| rs252749 | 5 | 77389973 | AP3B1 | G | A | 0.754 | 0.010 | 0.002 | 2.1E-09 | 7.91E-05 | 36 |
| rs59893724 | 5 | 80830788 | SSBP2 | A | G | 0.756 | 0.011 | 0.002 | 8.7E-13 | 1.13E-04 | 51 |
| rs79236537 | 5 | 86727690 | CCNH | G | T | 0.980 | -0.030 | 0.005 | 1E-09 | 8.23E-05 | 37 |
| rs6870983 | 5 | 87697533 | TMEM161B | C | T | 0.786 | 0.014 | 0.002 | 1.6E-16 | 1.50E-04 | 68 |
| rs1477290 | 5 | 87988934 | MEF2C | T | C | 0.863 | -0.020 | 0.002 | 3.5E-23 | 2.17E-04 | 98 |
| rs142503704 | 5 | 92622421 | NR2F1 | G | A | 0.977 | -0.026 | 0.005 | 1.9E-08 | 6.97E-05 | 32 |
| rs62379271 | 5 | 105870033 | EFNA5 | T | G | 0.421 | -0.008 | 0.001 | 2E-08 | 6.94E-05 | 31 |
| rs149457 | 5 | 107438057 | FBXL17 | C | T | 0.830 | 0.015 | 0.002 | 2.1E-16 | 1.49E-04 | 67 |
| rs12517187 | 5 | 112444682 | MCC | C | T | 0.565 | -0.008 | 0.001 | 2.8E-09 | 7.79E-05 | 35 |
| rs347551 | 5 | 119389031 | PRR16 | C | G | 0.528 | -0.009 | 0.001 | 1.8E-10 | 8.97E-05 | 41 |
| rs1582931 | 5 | 122657199 | CEP120 | G | A | 0.527 | 0.010 | 0.001 | 3E-12 | 1.07E-04 | 49 |
| rs4836133 | 5 | 124332103 | ZNF608 | C | A | 0.510 | -0.008 | 0.001 | 3.1E-09 | 7.75E-05 | 35 |
| rs329118 | 5 | 133861663 | JADE2 | C | T | 0.581 | 0.009 | 0.001 | 8.8E-12 | 1.03E-04 | 47 |
| rs13174863 | 5 | 139080745 | CXXC5 | A | G | 0.852 | -0.014 | 0.002 | 2.4E-12 | 1.08E-04 | 49 |
| rs7719067 | 5 | 153538241 | MFAP3 | A | G | 0.428 | 0.009 | 0.001 | 1.7E-11 | 9.99E-05 | 45 |
| rs11134512 | 5 | 167847460 | WWC1 | T | G | 0.677 | 0.008 | 0.001 | 1.9E-08 | 6.96E-05 | 32 |
| rs11134679 | 5 | 170623391 | RANBP17 | A | G | 0.315 | -0.012 | 0.001 | 5.2E-17 | 1.55E-04 | 70 |
| rs4467770 | 6 | 12086826 | HIVEP1 | G | A | 0.269 | -0.010 | 0.002 | 2E-10 | 8.92E-05 | 40 |
| rs9395520 | 6 | 13183523 | PHACTR1 | C | T | 0.696 | 0.010 | 0.001 | 5.9E-11 | 9.46E-05 | 43 |
| rs3806114 | 6 | 20482335 | E2F3 | G | A | 0.332 | 0.009 | 0.001 | 2.3E-09 | 7.87E-05 | 36 |
| rs75499503 | 6 | 26145217 | HIST1H2AC | C | T | 0.780 | 0.011 | 0.002 | 8E-12 | 1.03E-04 | 47 |
| rs34388845 | 6 | 28578286 | SCAND3 | A | G | 0.786 | -0.012 | 0.002 | 1.9E-13 | 1.19E-04 | 54 |
| rs2260051 | 6 | 31591918 | PRRC2A | A | T | 0.438 | -0.012 | 0.001 | 1.6E-17 | 1.60E-04 | 73 |
| rs9277992 | 6 | 33312455 | DAXX | G | A | 0.810 | -0.013 | 0.002 | 5.7E-14 | 1.25E-04 | 56 |
| rs9366863 | 6 | 34688946 | C6orf106 | T | C | 0.328 | 0.017 | 0.001 | 8.5E-33 | 3.14E-04 | 142 |
| rs34298980 | 6 | 40409243 | LRFN2 | T | C | 0.491 | 0.013 | 0.001 | 4.4E-20 | 1.86E-04 | 84 |
| rs9462670 | 6 | 41014309 | OARD1 | G | C | 0.769 | -0.009 | 0.002 | 7.1E-09 | 7.39E-05 | 34 |
| rs72892910 | 6 | 50816887 | TFAP2B | G | T | 0.828 | -0.025 | 0.002 | 5.7E-44 | 4.27E-04 | 193 |
| rs1327259 | 6 | 51177811 | PKHD1 | A | G | 0.612 | 0.010 | 0.001 | 1.3E-13 | 1.21E-04 | 55 |
| rs1547026 | 6 | 51825622 | PKHD1 | T | C | 0.709 | -0.010 | 0.002 | 6.8E-12 | 1.04E-04 | 47 |
| rs72910629 | 6 | 69761994 | BAI3 | A | G | 0.864 | -0.013 | 0.002 | 4.3E-11 | 9.59E-05 | 43 |
| rs1040046 | 6 | 83473573 | UBE3D | C | A | 0.851 | -0.011 | 0.002 | 4.6E-09 | 7.58E-05 | 34 |
| rs1324110 | 6 | 93913200 | EPHA7 | G | C | 0.557 | 0.008 | 0.001 | 2E-08 | 6.95E-05 | 32 |
| rs10499014 | 6 | 97947755 | MMS22L | C | G | 0.731 | 0.010 | 0.002 | 1.1E-10 | 9.20E-05 | 42 |
| rs6938973 | 6 | 98421721 | MMS22L | T | C | 0.399 | -0.012 | 0.001 | 3.5E-17 | 1.57E-04 | 71 |

| | | | | | | | | | | | |
|---|---|---|---|---|---|---|---|---|---|---|---|
| rs9496567 | 6 | 100602753 | MCHR2 | G | A | 0.757 | -0.009 | 0.002 | 4.4E-09 | 7.60E-05 | 34 |
| rs156126 | 6 | 104810083 | HACE1 | T | C | 0.196 | -0.010 | 0.002 | 2.7E-09 | 7.82E-05 | 35 |
| rs2253310 | 6 | 108888593 | FOXO3 | C | G | 0.374 | -0.011 | 0.001 | 9E-14 | 1.23E-04 | 56 |
| rs13218383 | 6 | 120173501 | MAN1A1 | C | G | 0.665 | 0.009 | 0.001 | 1.4E-10 | 9.09E-05 | 41 |
| rs2875762 | 6 | 124925032 | NKAIN2 | G | C | 0.757 | -0.011 | 0.002 | 6E-12 | 1.04E-04 | 47 |
| rs10457469 | 6 | 126083658 | HEY2 | G | A | 0.476 | -0.008 | 0.001 | 2.6E-08 | 6.83E-05 | 31 |
| rs12213441 | 6 | 143208838 | HIVEP2 | C | T | 0.771 | -0.011 | 0.002 | 3.3E-10 | 8.71E-05 | 39 |
| rs7749708 | 6 | 153375907 | RGS17 | C | T | 0.706 | -0.010 | 0.002 | 3.7E-11 | 9.66E-05 | 44 |
| rs9478496 | 6 | 154333183 | OPRM1 | T | C | 0.836 | -0.012 | 0.002 | 3.8E-10 | 8.65E-05 | 39 |
| rs9480184 | 6 | 155987788 | NOX3 | C | T | 0.789 | -0.010 | 0.002 | 4.9E-09 | 7.55E-05 | 34 |
| rs36007635 | 6 | 163009335 | PARK2 | G | A | 0.862 | 0.014 | 0.002 | 5.6E-12 | 1.05E-04 | 47 |
| rs6950388 | 7 | 1270699 | UNCX | G | A | 0.205 | -0.009 | 0.002 | 2.6E-08 | 6.83E-05 | 31 |
| rs2056477 | 7 | 2079744 | MAD1L1 | G | C | 0.772 | 0.012 | 0.002 | 1.5E-12 | 1.10E-04 | 50 |
| rs4307239 | 7 | 24354300 | NPY | A | G | 0.541 | -0.008 | 0.001 | 2.4E-08 | 6.86E-05 | 31 |
| rs215634 | 7 | 32369148 | PDE1C | A | G | 0.388 | 0.010 | 0.001 | 1.4E-13 | 1.21E-04 | 55 |
| rs2237402 | 7 | 39449768 | POU6F2 | G | A | 0.661 | 0.009 | 0.001 | 1.8E-10 | 8.98E-05 | 41 |
| rs2289379 | 7 | 44804225 | ZMIZ2 | C | T | 0.604 | 0.010 | 0.001 | 4.1E-12 | 1.06E-04 | 48 |
| rs3823674 | 7 | 50571996 | DDC | C | T | 0.572 | 0.008 | 0.001 | 3.5E-08 | 6.71E-05 | 30 |
| rs11765062 | 7 | 54417515 | VSTM2A | T | C | 0.459 | 0.008 | 0.001 | 4.6E-08 | 6.59E-05 | 30 |
| rs2866720 | 7 | 70106310 | AUTS2 | C | T | 0.615 | -0.009 | 0.001 | 1.2E-10 | 9.14E-05 | 41 |
| rs1852006 | 7 | 77829768 | MAGI2 | G | A | 0.643 | 0.009 | 0.001 | 6.7E-10 | 8.41E-05 | 38 |
| rs6963840 | 7 | 78144371 | MAGI2 | C | T | 0.844 | -0.013 | 0.002 | 8.5E-12 | 1.03E-04 | 47 |
| rs12538826 | 7 | 99030228 | PTCD1 | T | C | 0.886 | 0.017 | 0.002 | 7.2E-15 | 1.34E-04 | 61 |
| rs2074686 | 7 | 100800635 | AP1S1 | G | A | 0.410 | 0.008 | 0.001 | 2.9E-09 | 7.78E-05 | 35 |
| rs11496125 | 7 | 103417557 | RELN | C | T | 0.575 | -0.011 | 0.001 | 4.8E-15 | 1.35E-04 | 61 |
| rs2396625 | 7 | 113028634 | TSRM | T | A | 0.579 | 0.010 | 0.001 | 1.1E-12 | 1.12E-04 | 51 |
| rs12705894 | 7 | 113351252 | PPP1R3A | G | A | 0.448 | 0.008 | 0.001 | 1.6E-08 | 7.03E-05 | 32 |
| rs1840660 | 7 | 114352615 | FOXP2 | G | A | 0.612 | -0.010 | 0.001 | 2E-12 | 1.09E-04 | 50 |
| rs1899689 | 7 | 121964349 | CADPS2 | C | T | 0.610 | -0.008 | 0.001 | 3.7E-08 | 6.69E-05 | 30 |
| rs35775580 | 7 | 130420740 | KLF14 | A | G | 0.953 | 0.018 | 0.003 | 2.7E-08 | 6.82E-05 | 31 |
| rs11976084 | 7 | 137437156 | DGKI | C | T | 0.712 | -0.009 | 0.002 | 1.7E-08 | 7.02E-05 | 32 |
| rs11525873 | 7 | 138817193 | TTC26 | T | C | 0.902 | 0.015 | 0.002 | 1.4E-10 | 9.07E-05 | 41 |
| rs1805123 | 7 | 150645534 | KCNH2 | T | G | 0.755 | 0.011 | 0.002 | 4.1E-12 | 1.06E-04 | 48 |
| rs7827182 | 8 | 8380471 | SGK223 | G | C | 0.511 | -0.011 | 0.001 | 2.9E-16 | 1.48E-04 | 67 |
| rs6601451 | 8 | 10243681 | MSRA | C | G | 0.485 | 0.012 | 0.001 | 3.7E-17 | 1.56E-04 | 71 |
| rs55896564 | 8 | 11447093 | BLK | G | A | 0.465 | 0.012 | 0.001 | 2.6E-17 | 1.58E-04 | 72 |
| rs6530737 | 8 | 14095763 | SGCZ | A | G | 0.349 | 0.009 | 0.001 | 3.5E-10 | 8.69E-05 | 39 |
| rs2616143 | 8 | 20632022 | LZTS1 | G | A | 0.680 | 0.009 | 0.001 | 1.9E-09 | 7.95E-05 | 36 |
| rs117176448 | 8 | 27261138 | PTK2B | C | G | 0.904 | -0.015 | 0.002 | 1.2E-10 | 9.16E-05 | 42 |
| rs4266606 | 8 | 28043673 | ELP3 | C | T | 0.143 | 0.011 | 0.002 | 4.1E-08 | 6.64E-05 | 30 |
| rs2725371 | 8 | 30854033 | PURG | A | G | 0.304 | 0.010 | 0.001 | 2E-11 | 9.92E-05 | 45 |
| rs4739558 | 8 | 38337264 | FGFR1 | A | G | 0.399 | 0.008 | 0.001 | 7.4E-09 | 7.38E-05 | 33 |
| rs35894137 | 8 | 43071838 | HGSNAT | C | T | 0.919 | 0.014 | 0.003 | 1.6E-08 | 7.03E-05 | 32 |
| rs143662847 | 8 | 48804722 | PRKDC | C | T | 0.960 | 0.019 | 0.004 | 4.6E-08 | 6.59E-05 | 30 |
| rs473837 | 8 | 60906881 | CA8 | G | T | 0.648 | 0.009 | 0.001 | 1.8E-09 | 7.97E-05 | 36 |

| | | | | | | | | | | | |
|---|---|---|---|---|---|---|---|---|---|---|---|
| rs12681792 | 8 | 62054463 | CLVS1 | C | A | 0.807 | -0.010 | 0.002 | 4.3E-08 | 6.62E-05 | 30 |
| rs4737188 | 8 | 64756657 | YTHDF3 | A | T | 0.526 | 0.008 | 0.001 | 1.5E-09 | 8.06E-05 | 37 |
| rs35957544 | 8 | 73440371 | KCNB2 | G | T | 0.426 | 0.013 | 0.001 | 7.4E-20 | 1.84E-04 | 83 |
| rs2941432 | 8 | 76532219 | HNF4G | T | A | 0.528 | -0.008 | 0.001 | 1.3E-08 | 7.13E-05 | 32 |
| rs78565420 | 8 | 85703065 | RALYL | C | T | 0.948 | -0.019 | 0.003 | 2.7E-09 | 7.81E-05 | 35 |
| rs17619860 | 8 | 87779603 | CNGB3 | T | C | 0.837 | -0.010 | 0.002 | 2.2E-08 | 6.90E-05 | 31 |
| rs1905616 | 8 | 93235675 | RUNX1T1 | G | A | 0.669 | 0.008 | 0.001 | 1.8E-08 | 6.99E-05 | 32 |
| rs2114210 | 8 | 95595162 | KIAA1429 | G | A | 0.664 | -0.010 | 0.001 | 1.2E-11 | 1.02E-04 | 46 |
| rs17716502 | 8 | 116659731 | TRPS1 | C | T | 0.796 | 0.016 | 0.002 | 2.7E-21 | 1.98E-04 | 90 |
| rs72673947 | 8 | 118884379 | EXT1 | A | G | 0.893 | -0.014 | 0.002 | 3.3E-10 | 8.72E-05 | 40 |
| rs112875651 | 8 | 126506694 | TRIB1 | G | A | 0.609 | -0.008 | 0.001 | 4.3E-09 | 7.60E-05 | 34 |
| rs11782074 | 8 | 142617096 | AC138647.1 | G | T | 0.616 | -0.010 | 0.001 | 1.2E-11 | 1.01E-04 | 46 |
| rs1865341 | 9 | 8845911 | PTPRD | C | T | 0.238 | -0.009 | 0.002 | 2.5E-08 | 6.85E-05 | 31 |
| rs10960276 | 9 | 11819686 | TYRP1 | C | A | 0.645 | 0.008 | 0.001 | 1.6E-08 | 7.05E-05 | 32 |
| rs7020196 | 9 | 12289527 | TYRP1 | C | T | 0.420 | 0.008 | 0.001 | 3.6E-08 | 6.70E-05 | 30 |
| rs13292699 | 9 | 15910044 | CCDC171 | A | C | 0.566 | 0.013 | 0.001 | 1.5E-19 | 1.80E-04 | 82 |
| rs1411432 | 9 | 16728532 | BNC2 | A | C | 0.814 | -0.012 | 0.002 | 2E-12 | 1.09E-04 | 49 |
| rs17770336 | 9 | 28414625 | LINGO2 | C | T | 0.678 | -0.016 | 0.001 | 3.1E-26 | 2.48E-04 | 112 |
| rs10969334 | 9 | 29717279 | LINGO2 | C | A | 0.606 | 0.009 | 0.001 | 6.9E-10 | 8.40E-05 | 38 |
| rs10973159 | 9 | 36992547 | PAX5 | G | T | 0.380 | -0.008 | 0.001 | 1.2E-08 | 7.16E-05 | 32 |
| rs7038966 | 9 | 73777777 | TRPM3 | C | T | 0.589 | -0.009 | 0.001 | 1.4E-11 | 1.01E-04 | 46 |
| rs1547205 | 9 | 98815145 | ERCC6L2 | G | C | 0.900 | 0.013 | 0.002 | 1.3E-08 | 7.14E-05 | 32 |
| rs4989244 | 9 | 102100348 | SEC61B | G | A | 0.568 | 0.008 | 0.001 | 3.6E-08 | 6.70E-05 | 30 |
| rs2135745 | 9 | 109150784 | ZNF462 | C | G | 0.249 | 0.009 | 0.002 | 1.1E-08 | 7.22E-05 | 33 |
| rs2417998 | 9 | 111958746 | EPB41L4B | C | G | 0.292 | 0.008 | 0.002 | 2.8E-08 | 6.80E-05 | 31 |
| rs12376870 | 9 | 117890567 | TNC | G | A | 0.762 | 0.009 | 0.002 | 4.1E-08 | 6.64E-05 | 30 |
| rs7038943 | 9 | 120377178 | TLR4 | T | C | 0.661 | 0.009 | 0.001 | 1.6E-10 | 9.04E-05 | 41 |
| rs6478538 | 9 | 124627012 | TTLL11 | A | G | 0.323 | 0.009 | 0.001 | 1.9E-09 | 7.96E-05 | 36 |
| rs10760277 | 9 | 126093999 | CRB2 | C | T | 0.615 | -0.009 | 0.001 | 3.6E-10 | 8.68E-05 | 39 |
| rs7030609 | 9 | 129415775 | LMX1B | A | G | 0.910 | -0.015 | 0.002 | 3.5E-10 | 8.69E-05 | 39 |
| rs113132247 | 9 | 131026108 | GOLGA2 | G | A | 0.847 | -0.012 | 0.002 | 1.1E-10 | 9.18E-05 | 42 |
| rs7913496 | 10 | 10257277 | CELF2 | C | T | 0.180 | 0.010 | 0.002 | 2E-08 | 6.96E-05 | 32 |
| rs7893571 | 10 | 16750129 | RSU1 | G | T | 0.334 | -0.010 | 0.001 | 2.9E-12 | 1.08E-04 | 49 |
| rs12253527 | 10 | 21819824 | MLLT10 | G | A | 0.678 | -0.014 | 0.001 | 2E-20 | 1.89E-04 | 86 |
| rs71495049 | 10 | 34014435 | PARD3 | G | A | 0.916 | -0.017 | 0.002 | 5.7E-12 | 1.05E-04 | 47 |
| rs3125326 | 10 | 63053788 | TMEM26 | A | C | 0.392 | -0.008 | 0.001 | 2.7E-08 | 6.82E-05 | 31 |
| rs7924036 | 10 | 65191645 | JMJD1C | G | T | 0.497 | 0.010 | 0.001 | 2.2E-12 | 1.09E-04 | 49 |
| rs11000993 | 10 | 76084111 | ADK | T | C | 0.876 | -0.014 | 0.002 | 3.1E-11 | 9.74E-05 | 44 |
| rs1250597 | 10 | 81010250 | ZMIZ1 | A | G | 0.405 | -0.009 | 0.001 | 1E-09 | 8.22E-05 | 37 |
| rs17399739 | 10 | 87490850 | GRID1 | A | G | 0.931 | -0.018 | 0.003 | 5.9E-11 | 9.45E-05 | 43 |
| rs2450444 | 10 | 93010383 | PCGF5 | G | A | 0.651 | 0.008 | 0.001 | 2E-08 | 6.96E-05 | 32 |
| rs41310284 | 10 | 102447647 | PAX2 | C | A | 0.899 | 0.018 | 0.002 | 9.8E-15 | 1.32E-04 | 60 |
| rs10736156 | 10 | 104019447 | GBF1 | C | A | 0.156 | -0.012 | 0.002 | 3.8E-10 | 8.65E-05 | 39 |
| rs7086898 | 10 | 104386152 | SUFU | A | G | 0.920 | -0.014 | 0.003 | 4.3E-08 | 6.62E-05 | 30 |
| rs4575195 | 10 | 114765747 | TCF7L2 | C | A | 0.687 | 0.010 | 0.001 | 9E-11 | 9.28E-05 | 42 |

| | | | | | | | | | | | |
|---|---|---|---|---|---|---|---|---|---|---|---|
| rs9421249 | 10 | 118623322 | ENO4 | C | T | 0.739 | -0.010 | 0.002 | 2.5E-11 | 9.83E-05 | 45 |
| rs845084 | 10 | 125220036 | GPR26 | G | A | 0.742 | -0.010 | 0.002 | 3.7E-10 | 8.66E-05 | 39 |
| rs4962725 | 10 | 126733321 | CTBP2 | T | C | 0.572 | -0.010 | 0.001 | 6.6E-13 | 1.14E-04 | 52 |
| rs2542615 | 10 | 131128952 | MGMT | C | T | 0.342 | 0.008 | 0.001 | 7.3E-09 | 7.38E-05 | 33 |
| rs2035806 | 10 | 133984916 | JAKMIP3 | G | A | 0.435 | 0.010 | 0.001 | 5.7E-12 | 1.05E-04 | 47 |
| rs67257872 | 11 | 8530218 | STK33 | A | G | 0.553 | 0.011 | 0.001 | 1.5E-14 | 1.30E-04 | 59 |
| rs28711392 | 11 | 13349559 | ARNTL | T | C | 0.633 | 0.012 | 0.001 | 2.1E-16 | 1.49E-04 | 67 |
| rs6265 | 11 | 27679916 | BDNF | C | T | 0.812 | 0.025 | 0.002 | 1.5E-44 | 4.32E-04 | 196 |
| rs10835498 | 11 | 29258947 | KCNA4 | G | A | 0.569 | -0.008 | 0.001 | 8.9E-09 | 7.29E-05 | 33 |
| rs1222216 | 11 | 30346052 | ARL14EP | C | T | 0.774 | 0.012 | 0.002 | 1.1E-13 | 1.22E-04 | 55 |
| rs59227842 | 11 | 43692423 | HSD17B12 | A | G | 0.688 | -0.015 | 0.001 | 1.8E-24 | 2.30E-04 | 104 |
| rs868784 | 11 | 43944388 | C11orf96 | G | A | 0.619 | 0.008 | 0.001 | 2.7E-08 | 6.81E-05 | 31 |
| rs6416134 | 11 | 45352781 | SYT13 | G | C | 0.181 | -0.010 | 0.002 | 4.6E-08 | 6.59E-05 | 30 |
| rs12798028 | 11 | 47604639 | NDUFS3 | C | T | 0.591 | -0.015 | 0.001 | 3.4E-27 | 2.57E-04 | 117 |
| rs12363672 | 11 | 55684028 | OR5W2 | A | C | 0.974 | -0.024 | 0.004 | 2.8E-08 | 6.81E-05 | 31 |
| rs34292685 | 11 | 64049021 | GPR137 | C | T | 0.838 | 0.013 | 0.002 | 7.9E-12 | 1.03E-04 | 47 |
| rs2234458 | 11 | 65639374 | EFEMP2 | C | T | 0.360 | 0.013 | 0.001 | 2.5E-19 | 1.78E-04 | 81 |
| rs667515 | 11 | 69449076 | CCND1 | G | C | 0.614 | 0.009 | 0.001 | 8.4E-10 | 8.31E-05 | 38 |
| rs10160769 | 11 | 76474827 | TSKU | G | C | 0.783 | 0.009 | 0.002 | 4.6E-08 | 6.59E-05 | 30 |
| rs7102934 | 11 | 84648068 | DLG2 | T | C | 0.688 | -0.009 | 0.001 | 1.2E-09 | 8.15E-05 | 37 |
| rs61903695 | 11 | 89922417 | NAALAD2 | A | G | 0.745 | -0.011 | 0.002 | 6.6E-13 | 1.14E-04 | 52 |
| rs2658797 | 11 | 93212254 | SMCO4 | C | T | 0.518 | 0.008 | 0.001 | 1.3E-08 | 7.12E-05 | 32 |
| rs680071 | 11 | 103088414 | DYNC2H1 | T | C | 0.119 | -0.012 | 0.002 | 2.8E-08 | 6.80E-05 | 31 |
| rs719802 | 11 | 113234679 | TTC12 | T | C | 0.386 | 0.008 | 0.001 | 5.7E-09 | 7.49E-05 | 34 |
| rs11607476 | 11 | 115037061 | CADM1 | A | C | 0.513 | -0.010 | 0.001 | 9.2E-13 | 1.13E-04 | 51 |
| rs7928320 | 11 | 116942753 | SIK3 | C | G | 0.943 | -0.017 | 0.003 | 6.1E-09 | 7.46E-05 | 34 |
| rs12281009 | 11 | 117032959 | PAFAH1B2 | A | G | 0.942 | -0.018 | 0.003 | 1.3E-09 | 8.11E-05 | 37 |
| rs7925100 | 11 | 118941596 | VPS11 | G | A | 0.604 | -0.009 | 0.001 | 6.3E-11 | 9.43E-05 | 43 |
| rs11218510 | 11 | 121922587 | BLID | G | A | 0.600 | 0.008 | 0.001 | 7.8E-09 | 7.36E-05 | 33 |
| rs10791113 | 11 | 130873165 | SNX19 | A | G | 0.494 | -0.009 | 0.001 | 4.6E-10 | 8.57E-05 | 39 |
| rs12788343 | 11 | 131452912 | NTM | T | C | 0.591 | -0.010 | 0.001 | 4.3E-12 | 1.06E-04 | 48 |
| rs11223204 | 11 | 132652554 | OPCML | A | G | 0.566 | -0.009 | 0.001 | 3E-10 | 8.76E-05 | 40 |
| rs329651 | 11 | 133767622 | IGSF9B | G | T | 0.196 | -0.011 | 0.002 | 1.7E-10 | 9.01E-05 | 41 |
| rs61909165 | 11 | 134589355 | AP003062.1 | T | A | 0.826 | -0.012 | 0.002 | 1.6E-11 | 1.00E-04 | 45 |
| rs55726687 | 12 | 991306 | WNK1 | G | A | 0.790 | -0.014 | 0.002 | 3E-16 | 1.47E-04 | 67 |
| rs10774018 | 12 | 2157925 | CACNA1C | G | C | 0.780 | -0.009 | 0.002 | 1.1E-08 | 7.19E-05 | 33 |
| rs1799507 | 12 | 16427314 | SLC15A5 | G | A | 0.856 | -0.011 | 0.002 | 3.1E-08 | 6.77E-05 | 31 |
| rs10505836 | 12 | 19288508 | PLEKHA5 | A | C | 0.140 | -0.012 | 0.002 | 1.1E-09 | 8.19E-05 | 37 |
| rs10842231 | 12 | 23986029 | SOX5 | A | T | 0.917 | -0.014 | 0.003 | 8.4E-09 | 7.32E-05 | 33 |
| rs1458156 | 12 | 41887940 | PDZRN4 | C | T | 0.512 | -0.009 | 0.001 | 1.3E-11 | 1.01E-04 | 46 |
| rs1126930 | 12 | 49399132 | PRKAG1 | G | C | 0.965 | -0.021 | 0.004 | 8.6E-09 | 7.31E-05 | 33 |
| rs7132908 | 12 | 50263148 | FAIM2 | G | A | 0.616 | -0.019 | 0.001 | 8.4E-40 | 3.85E-04 | 174 |
| rs4077093 | 12 | 51593616 | POU6F1 | T | G | 0.216 | 0.010 | 0.002 | 7.9E-09 | 7.35E-05 | 33 |
| rs4759073 | 12 | 54653258 | CBX5 | G | A | 0.590 | 0.009 | 0.001 | 1.9E-11 | 9.95E-05 | 45 |
| rs4759228 | 12 | 56508409 | PA2G4 | G | C | 0.705 | 0.012 | 0.002 | 1.3E-14 | 1.31E-04 | 59 |

| | | | | | | | | | | | |
|---|---|---|---|---|---|---|---|---|---|---|---|
| rs12821416 | 12 | 58580570 | XRCC6BP1 | C | T | 0.864 | -0.011 | 0.002 | 1.2E-08 | 7.16E-05 | 32 |
| rs61754230 | 12 | 72179446 | RAB21 | C | T | 0.980 | -0.027 | 0.005 | 2.8E-08 | 6.81E-05 | 31 |
| rs12427047 | 12 | 90213070 | ATP2B1 | C | T | 0.757 | 0.011 | 0.002 | 8.9E-12 | 1.03E-04 | 47 |
| rs2712667 | 12 | 99588917 | ANKS1B | G | C | 0.356 | 0.010 | 0.001 | 2.1E-11 | 9.89E-05 | 45 |
| rs4764949 | 12 | 103658096 | C12orf42 | A | G | 0.674 | 0.012 | 0.001 | 3.8E-16 | 1.46E-04 | 66 |
| rs6606686 | 12 | 110903380 | GPN3 | G | C | 0.319 | 0.010 | 0.001 | 3.4E-11 | 9.69E-05 | 44 |
| rs11513729 | 12 | 112273499 | MAPKAPK5 | C | T | 0.587 | 0.008 | 0.001 | 1.5E-08 | 7.08E-05 | 32 |
| rs181617194 | 12 | 122011598 | KDM2B | T | C | 0.959 | 0.023 | 0.004 | 1.7E-09 | 8.00E-05 | 36 |
| rs147730268 | 12 | 123024476 | KNTC1 | G | T | 0.913 | 0.023 | 0.002 | 4E-20 | 1.86E-04 | 84 |
| rs9579775 | 13 | 20616557 | ZMYM2 | A | C | 0.864 | -0.015 | 0.002 | 2.1E-12 | 1.09E-04 | 49 |
| rs9507791 | 13 | 27352678 | GPR12 | G | A | 0.213 | 0.010 | 0.002 | 4.3E-09 | 7.61E-05 | 34 |
| rs1967772 | 13 | 28036062 | MTIF3 | G | A | 0.715 | 0.011 | 0.002 | 8.2E-13 | 1.13E-04 | 51 |
| rs35193668 | 13 | 33092929 | N4BP2L2 | C | T | 0.639 | 0.010 | 0.001 | 2.5E-13 | 1.18E-04 | 54 |
| rs61954177 | 13 | 40787036 | AL133318.1 | G | C | 0.673 | -0.009 | 0.001 | 2.7E-09 | 7.81E-05 | 35 |
| rs12429545 | 13 | 54102206 | OLFM4 | G | A | 0.871 | -0.020 | 0.002 | 2.6E-22 | 2.08E-04 | 94 |
| rs7321285 | 13 | 54319327 | OLFM4 | A | C | 0.201 | 0.011 | 0.002 | 4.1E-11 | 9.61E-05 | 44 |
| rs2576135 | 13 | 54691442 | OLFM4 | T | A | 0.093 | 0.013 | 0.002 | 3.5E-08 | 6.71E-05 | 30 |
| rs9317002 | 13 | 59175727 | PCDH17 | C | A | 0.485 | -0.011 | 0.001 | 1.9E-16 | 1.49E-04 | 68 |
| rs9529148 | 13 | 67419495 | PCDH9 | G | A | 0.377 | -0.008 | 0.001 | 1.9E-08 | 6.97E-05 | 32 |
| rs1576655 | 13 | 79587841 | RBM26 | A | C | 0.404 | -0.011 | 0.001 | 1.9E-15 | 1.39E-04 | 63 |
| rs61971082 | 13 | 86494667 | SLITRK6 | T | G | 0.716 | -0.010 | 0.002 | 3E-11 | 9.75E-05 | 44 |
| rs7331420 | 13 | 99236471 | STK24 | G | A | 0.715 | 0.009 | 0.002 | 2.1E-09 | 7.92E-05 | 36 |
| rs9888533 | 13 | 107854612 | FAM155A | C | T | 0.462 | -0.008 | 0.001 | 4.5E-08 | 6.60E-05 | 30 |
| rs9522180 | 13 | 111970212 | TEX29 | C | T | 0.447 | 0.009 | 0.001 | 2.4E-11 | 9.85E-05 | 45 |
| rs9515446 | 13 | 112217108 | RP11-65D24.2 | A | G | 0.552 | -0.010 | 0.001 | 2.2E-12 | 1.09E-04 | 49 |
| rs8015400 | 14 | 25930988 | STXBP6 | C | A | 0.323 | -0.013 | 0.001 | 6.9E-18 | 1.64E-04 | 74 |
| rs9788550 | 14 | 29681138 | PRKD1 | G | C | 0.753 | 0.014 | 0.002 | 5.6E-18 | 1.65E-04 | 75 |
| rs12883788 | 14 | 33303540 | AKAP6 | C | T | 0.540 | -0.012 | 0.001 | 2.3E-19 | 1.79E-04 | 81 |
| rs7141912 | 14 | 35649431 | KIAA0391 | A | T | 0.874 | 0.012 | 0.002 | 1.1E-08 | 7.21E-05 | 33 |
| rs8011566 | 14 | 42939471 | LRFN5 | T | A | 0.574 | -0.009 | 0.001 | 3.5E-10 | 8.69E-05 | 39 |
| rs724623 | 14 | 47303577 | MDGA2 | A | C | 0.492 | 0.010 | 0.001 | 9E-14 | 1.23E-04 | 56 |
| rs217672 | 14 | 62361021 | SYT16 | A | C | 0.728 | -0.012 | 0.002 | 2.3E-14 | 1.29E-04 | 58 |
| rs3902951 | 14 | 69789755 | GALNT16 | T | G | 0.763 | -0.011 | 0.002 | 1.1E-10 | 9.18E-05 | 42 |
| rs61986330 | 14 | 73314450 | DPF3 | C | A | 0.726 | 0.009 | 0.002 | 1.4E-08 | 7.10E-05 | 32 |
| rs10146997 | 14 | 79945162 | NRXN3 | A | G | 0.778 | -0.016 | 0.002 | 3.2E-22 | 2.07E-04 | 94 |
| rs8008772 | 14 | 88321884 | GALC | A | T | 0.749 | -0.009 | 0.002 | 5.7E-09 | 7.49E-05 | 34 |
| rs1286138 | 14 | 91485445 | RPS6KA5 | T | G | 0.325 | -0.009 | 0.001 | 8.4E-10 | 8.31E-05 | 38 |
| rs6575340 | 14 | 94023972 | UNC79 | G | A | 0.364 | -0.013 | 0.001 | 1.2E-20 | 1.92E-04 | 87 |
| rs12885251 | 14 | 99670791 | BCL11B | G | A | 0.542 | 0.008 | 0.001 | 4.1E-08 | 6.64E-05 | 30 |
| rs12147845 | 14 | 101144596 | DLK1 | C | T | 0.884 | -0.014 | 0.002 | 2.6E-10 | 8.81E-05 | 40 |
| rs61992671 | 14 | 101531854 | AL117190.3 | A | G | 0.508 | 0.010 | 0.001 | 5.7E-12 | 1.05E-04 | 47 |
| rs7145882 | 14 | 103255461 | TRAF3 | T | C | 0.342 | 0.011 | 0.001 | 2.2E-15 | 1.39E-04 | 63 |
| rs3759584 | 14 | 103990799 | CKB | T | C | 0.637 | 0.010 | 0.001 | 1.1E-11 | 1.02E-04 | 46 |
| rs76520838 | 15 | 47916618 | SEMA6D | C | T | 0.967 | -0.023 | 0.004 | 1.9E-09 | 7.95E-05 | 36 |
| rs7182917 | 15 | 52080803 | TMOD2 | T | C | 0.550 | 0.009 | 0.001 | 2.8E-10 | 8.78E-05 | 40 |

| | | | | | | | | | | | |
|---|---|---|---|---|---|---|---|---|---|---|---|
| rs2247401 | 15 | 53156672 | ONECUT1 | G | A | 0.733 | -0.009 | 0.002 | 4.7E-08 | 6.58E-05 | 30 |
| rs28465175 | 15 | 53427155 | ONECUT1 | A | G | 0.930 | 0.015 | 0.003 | 2.6E-08 | 6.84E-05 | 31 |
| rs7175642 | 15 | 59450079 | MYO1E | T | G | 0.342 | 0.009 | 0.001 | 5.2E-09 | 7.52E-05 | 34 |
| rs28408562 | 15 | 60917079 | RORA | C | G | 0.553 | -0.008 | 0.001 | 1.3E-08 | 7.13E-05 | 32 |
| rs1369159 | 15 | 66360842 | MEGF11 | C | T | 0.420 | 0.008 | 0.001 | 1.7E-08 | 7.02E-05 | 32 |
| rs111584879 | 15 | 66678173 | TIPIN | T | C | 0.761 | 0.009 | 0.002 | 4.8E-08 | 6.57E-05 | 30 |
| rs2241420 | 15 | 68082816 | MAP2K5 | G | A | 0.774 | 0.018 | 0.002 | 1.9E-29 | 2.80E-04 | 127 |
| rs62004865 | 15 | 74207695 | LOXL1 | T | A | 0.894 | -0.014 | 0.002 | 1.4E-09 | 8.08E-05 | 37 |
| rs11856579 | 15 | 78012688 | LINGO1 | G | A | 0.732 | 0.009 | 0.002 | 3.4E-09 | 7.71E-05 | 35 |
| rs57488047 | 15 | 79403002 | RASGRF1 | T | C | 0.532 | 0.010 | 0.001 | 1.1E-12 | 1.12E-04 | 51 |
| rs34994596 | 15 | 80991447 | ABHD17C | T | C | 0.703 | 0.011 | 0.002 | 1.1E-13 | 1.22E-04 | 55 |
| rs7498044 | 15 | 92573639 | SLCO3A1 | G | A | 0.783 | 0.010 | 0.002 | 3.2E-09 | 7.73E-05 | 35 |
| rs8038574 | 15 | 95275890 | MCTP2 | T | C | 0.346 | 0.009 | 0.001 | 4E-10 | 8.63E-05 | 39 |
| rs56803094 | 15 | 99222509 | IGF1R | A | G | 0.773 | 0.010 | 0.002 | 5.8E-09 | 7.48E-05 | 34 |
| rs412243 | 16 | 339672 | AXIN1 | T | C | 0.621 | 0.010 | 0.001 | 1.4E-13 | 1.21E-04 | 55 |
| rs2516726 | 16 | 2095065 | NTHL1 | T | C | 0.772 | 0.010 | 0.002 | 1.7E-09 | 8.01E-05 | 36 |
| rs879620 | 16 | 4015729 | ADCY9 | C | T | 0.387 | -0.014 | 0.001 | 5.9E-24 | 2.25E-04 | 102 |
| rs2660241 | 16 | 4940023 | PPL | T | C | 0.635 | -0.008 | 0.001 | 1.2E-08 | 7.16E-05 | 32 |
| rs11642387 | 16 | 6753239 | RBFOX1 | A | G | 0.899 | 0.013 | 0.002 | 2.1E-08 | 6.94E-05 | 31 |
| rs39674 | 16 | 9413210 | C16orf72 | C | G | 0.304 | -0.009 | 0.001 | 6E-09 | 7.47E-05 | 34 |
| rs12927792 | 16 | 9713194 | RP11-297M9.1 | C | T | 0.574 | -0.009 | 0.001 | 5.2E-10 | 8.52E-05 | 39 |
| rs8054082 | 16 | 19975418 | GPR139 | C | T | 0.859 | 0.017 | 0.002 | 2.9E-18 | 1.68E-04 | 76 |
| rs4432271 | 16 | 20245283 | GP2 | C | T | 0.131 | -0.012 | 0.002 | 2.1E-08 | 6.92E-05 | 31 |
| rs11864909 | 16 | 20400839 | PDILT | C | T | 0.714 | -0.009 | 0.002 | 5.1E-09 | 7.54E-05 | 34 |
| rs9922288 | 16 | 24550930 | RBBP6 | A | G | 0.236 | 0.009 | 0.002 | 9.8E-09 | 7.25E-05 | 33 |
| rs7498665 | 16 | 28883241 | SH2B1 | A | G | 0.600 | -0.017 | 0.001 | 5.9E-35 | 3.36E-04 | 152 |
| rs3814883 | 16 | 29994922 | TAOK2 | C | T | 0.518 | -0.015 | 0.001 | 8.1E-27 | 2.54E-04 | 115 |
| rs34898535 | 16 | 31025641 | STX1B | C | T | 0.622 | 0.014 | 0.001 | 4.8E-24 | 2.26E-04 | 102 |
| rs56094641 | 16 | 53806453 | FTO | A | G | 0.595 | -0.047 | 0.001 | 4E-243 | 2.44E-03 | 1109 |
| rs862320 | 16 | 69651866 | NFAT5 | C | T | 0.590 | 0.014 | 0.001 | 6.2E-25 | 2.35E-04 | 106 |
| rs12149660 | 16 | 70309237 | AARS | G | A | 0.885 | 0.016 | 0.002 | 6.2E-14 | 1.24E-04 | 56 |
| rs811054 | 16 | 72251132 | PMFBP1 | C | T | 0.463 | -0.009 | 0.001 | 4.4E-10 | 8.59E-05 | 39 |
| rs4500770 | 16 | 74658430 | RFWD3 | A | T | 0.637 | 0.008 | 0.001 | 4.3E-08 | 6.62E-05 | 30 |
| rs9673839 | 16 | 76895693 | RP11-58C22.1 | A | G | 0.509 | -0.008 | 0.001 | 1.7E-09 | 8.00E-05 | 36 |
| rs12926506 | 16 | 81722413 | CMIP | C | T | 0.830 | 0.011 | 0.002 | 1.6E-09 | 8.03E-05 | 36 |
| rs11150462 | 16 | 82451679 | CDH13 | T | A | 0.368 | 0.008 | 0.001 | 3.1E-09 | 7.75E-05 | 35 |
| rs7206608 | 16 | 82872628 | CDH13 | C | G | 0.678 | -0.010 | 0.001 | 2.6E-11 | 9.80E-05 | 44 |
| rs4790292 | 17 | 1824305 | RTN4RL1 | C | A | 0.846 | 0.016 | 0.002 | 1.3E-17 | 1.61E-04 | 73 |
| rs58351927 | 17 | 5297038 | NUP88 | A | G | 0.698 | -0.010 | 0.001 | 1.2E-11 | 1.01E-04 | 46 |
| rs4792716 | 17 | 15943144 | NCOR1 | A | G | 0.438 | -0.008 | 0.001 | 4.8E-09 | 7.57E-05 | 34 |
| rs1320251 | 17 | 21264396 | KCNJ12 | C | T | 0.545 | 0.012 | 0.001 | 3.2E-19 | 1.77E-04 | 80 |
| rs1017529 | 17 | 27912415 | GIT1 | C | A | 0.825 | -0.011 | 0.002 | 7.7E-09 | 7.36E-05 | 33 |
| rs73982435 | 17 | 31473455 | ASIC2 | C | T | 0.784 | 0.009 | 0.002 | 1.6E-08 | 7.05E-05 | 32 |
| rs113962925 | 17 | 46044446 | CDK5RAP3 | C | T | 0.926 | -0.017 | 0.003 | 8.6E-11 | 9.29E-05 | 42 |
| rs11079849 | 17 | 47090785 | IGF2BP1 | C | T | 0.671 | 0.012 | 0.001 | 6.4E-17 | 1.54E-04 | 70 |

| | | | | | | | | | | | |
|---|---|---|---|---|---|---|---|---|---|---|---|
| rs78369934 | 17 | 61739101 | MAP3K3 | T | C | 0.945 | 0.020 | 0.003 | 6E-11 | 9.45E-05 | 43 |
| rs11150745 | 17 | 78757626 | RPTOR | A | G | 0.682 | 0.014 | 0.001 | 9.2E-21 | 1.93E-04 | 87 |
| rs2083323 | 18 | 1856272 | METTL4 | G | A | 0.822 | -0.010 | 0.002 | 7.3E-09 | 7.38E-05 | 33 |
| rs512121 | 18 | 7548501 | PTPRM | T | C | 0.808 | 0.011 | 0.002 | 1.5E-09 | 8.06E-05 | 37 |
| rs1788808 | 18 | 21090023 | NPC1 | A | G | 0.505 | 0.013 | 0.001 | 1.9E-20 | 1.89E-04 | 86 |
| rs16940823 | 18 | 22137319 | HRH4 | C | A | 0.815 | 0.011 | 0.002 | 1.6E-09 | 8.03E-05 | 36 |
| rs6507054 | 18 | 31248323 | ASXL3 | T | C | 0.416 | -0.009 | 0.001 | 2.7E-11 | 9.80E-05 | 44 |
| rs559231 | 18 | 39644247 | PIK3C3 | G | T | 0.607 | -0.010 | 0.001 | 1.5E-11 | 1.00E-04 | 46 |
| rs1834144 | 18 | 40744790 | RIT2 | C | A | 0.627 | 0.008 | 0.001 | 2.5E-08 | 6.86E-05 | 31 |
| rs7230240 | 18 | 42597978 | SETBP1 | C | T | 0.705 | 0.009 | 0.002 | 4.2E-10 | 8.61E-05 | 39 |
| rs58243949 | 18 | 52509833 | RAB27B | C | T | 0.767 | -0.012 | 0.002 | 1.3E-13 | 1.21E-04 | 55 |
| rs11659764 | 18 | 53335512 | TCF4 | T | A | 0.947 | 0.018 | 0.003 | 1.9E-09 | 7.96E-05 | 36 |
| rs1517037 | 18 | 56878274 | GRP | C | T | 0.812 | 0.010 | 0.002 | 1.6E-08 | 7.03E-05 | 32 |
| rs58084604 | 18 | 57849429 | MC4R | C | T | 0.767 | -0.035 | 0.002 | 2.6E-104 | 1.04E-03 | 470 |
| rs17773370 | 18 | 57951433 | MC4R | G | A | 0.934 | -0.016 | 0.003 | 8.7E-09 | 7.31E-05 | 33 |
| rs57636386 | 18 | 58048295 | MC4R | T | C | 0.916 | 0.025 | 0.002 | 2.7E-23 | 2.18E-04 | 99 |
| rs12454712 | 18 | 60845884 | BCL2 | T | C | 0.623 | -0.008 | 0.001 | 1E-08 | 7.24E-05 | 33 |
| rs1373349 | 18 | 63282992 | CDH7 | C | T | 0.316 | 0.010 | 0.001 | 3.5E-11 | 9.68E-05 | 44 |
| rs8089514 | 18 | 69224478 | RP11-723G8.2 | T | A | 0.631 | -0.008 | 0.001 | 1.4E-08 | 7.11E-05 | 32 |
| rs45521740 | 19 | 2245622 | SF3A2 | G | A | 0.943 | -0.018 | 0.003 | 1E-09 | 8.23E-05 | 37 |
| rs72976986 | 19 | 4050424 | ZBTB7A | G | A | 0.810 | 0.014 | 0.002 | 4.4E-15 | 1.36E-04 | 62 |
| rs75957461 | 19 | 11166163 | SMARCA4 | C | T | 0.946 | -0.018 | 0.003 | 6.6E-09 | 7.43E-05 | 34 |
| rs6511826 | 19 | 12706991 | ZNF490 | G | A | 0.087 | 0.014 | 0.002 | 1.3E-08 | 7.15E-05 | 32 |
| rs273505 | 19 | 18217147 | MAST3 | T | C | 0.578 | -0.010 | 0.001 | 3.1E-13 | 1.17E-04 | 53 |
| rs113230003 | 19 | 18460956 | PGPEP1 | G | A | 0.739 | 0.013 | 0.002 | 1.2E-15 | 1.41E-04 | 64 |
| rs10404726 | 19 | 18834514 | CRTC1 | C | T | 0.535 | 0.012 | 0.001 | 4.7E-19 | 1.76E-04 | 80 |
| rs112253053 | 19 | 19425145 | SUGP1 | T | A | 0.839 | 0.014 | 0.002 | 1.8E-13 | 1.20E-04 | 54 |
| rs12462975 | 19 | 30272202 | CCNE1 | G | A | 0.670 | -0.011 | 0.001 | 5.9E-15 | 1.34E-04 | 61 |
| rs73026723 | 19 | 31017177 | ZNF536 | C | T | 0.846 | 0.014 | 0.002 | 1.3E-13 | 1.21E-04 | 55 |
| rs7255223 | 19 | 32824310 | ZNF507 | C | A | 0.735 | 0.009 | 0.002 | 8.5E-09 | 7.32E-05 | 33 |
| rs429358 | 19 | 45411941 | APOE | T | C | 0.846 | 0.016 | 0.002 | 2.9E-17 | 1.58E-04 | 71 |
| rs12971645 | 19 | 45807945 | MARK4 | G | A | 0.725 | 0.009 | 0.002 | 2.6E-08 | 6.84E-05 | 31 |
| rs10423928 | 19 | 46182304 | GIPR | T | A | 0.806 | 0.021 | 0.002 | 4.2E-33 | 3.17E-04 | 144 |
| rs3810291 | 19 | 47569003 | ZC3H4 | G | A | 0.325 | -0.016 | 0.001 | 4.1E-27 | 2.57E-04 | 116 |
| rs4545921 | 19 | 49646006 | PPFIA3 | A | G | 0.400 | 0.008 | 0.001 | 4.3E-08 | 6.62E-05 | 30 |
| rs61746970 | 19 | 51132746 | SYT3 | G | A | 0.961 | -0.021 | 0.004 | 3.8E-09 | 7.66E-05 | 35 |
| rs8111074 | 19 | 51776117 | SIGLECL1 | G | T | 0.714 | 0.008 | 0.002 | 3.2E-08 | 6.74E-05 | 31 |
| rs6075658 | 20 | 2094078 | STK35 | T | C | 0.531 | 0.008 | 0.001 | 6.8E-10 | 8.40E-05 | 38 |
| rs2206925 | 20 | 6634895 | BMP2 | T | C | 0.364 | -0.012 | 0.001 | 2E-16 | 1.49E-04 | 68 |
| rs4813224 | 20 | 16546846 | KIF16B | T | C | 0.262 | -0.009 | 0.002 | 5.4E-09 | 7.51E-05 | 34 |
| rs8124896 | 20 | 21385659 | NKX2-4 | T | C | 0.899 | -0.013 | 0.002 | 8.1E-09 | 7.34E-05 | 33 |
| rs6050446 | 20 | 25195509 | ENTPD6 | A | G | 0.033 | -0.026 | 0.004 | 1.2E-11 | 1.02E-04 | 46 |
| rs201475383 | 20 | 26273991 | FAM182B | G | A | 0.966 | 0.024 | 0.004 | 8.8E-10 | 8.29E-05 | 38 |
| rs1987960 | 20 | 30649834 | HCK | T | C | 0.048 | -0.019 | 0.003 | 1.1E-08 | 7.21E-05 | 33 |
| rs4911382 | 20 | 32553095 | RALY | C | T | 0.415 | -0.008 | 0.001 | 2.6E-09 | 7.83E-05 | 35 |

| | | | | | | | | | | | |
|---|---|---|---|---|---|---|---|---|---|---|---|
| rs6029180 | 20 | 39178923 | MAFB | A | G | 0.674 | -0.008 | 0.001 | 3.1E-08 | 6.76E-05 | 31 |
| rs6030803 | 20 | 41986507 | SRSF6 | T | C | 0.873 | 0.013 | 0.002 | 4.5E-10 | 8.58E-05 | 39 |
| rs2425856 | 20 | 44911954 | CDH22 | A | G | 0.444 | 0.008 | 0.001 | 1.1E-09 | 8.20E-05 | 37 |
| rs112852122 | 20 | 47498117 | ARFGEF2 | G | A | 0.842 | 0.013 | 0.002 | 2.7E-12 | 1.08E-04 | 49 |
| rs66460909 | 20 | 51195387 | ZFP64 | G | A | 0.808 | 0.016 | 0.002 | 2.9E-19 | 1.78E-04 | 81 |
| rs559390 | 20 | 54157546 | CBLN4 | A | T | 0.658 | -0.008 | 0.001 | 7.9E-09 | 7.35E-05 | 33 |
| rs8134638 | 21 | 40644170 | BRWD1 | T | C | 0.624 | -0.008 | 0.001 | 4E-08 | 6.65E-05 | 30 |
| rs2837398 | 21 | 41427168 | DSCAM | A | C | 0.596 | -0.008 | 0.001 | 4.8E-09 | 7.56E-05 | 34 |
| rs1964926 | 21 | 42653121 | BACE2 | A | G | 0.354 | -0.008 | 0.001 | 1.1E-08 | 7.21E-05 | 33 |
| rs403694 | 21 | 46567625 | ADARB1 | C | T | 0.463 | -0.012 | 0.001 | 1.3E-18 | 1.71E-04 | 78 |
| rs1296685 | 22 | 18230964 | BID | A | G | 0.789 | -0.009 | 0.002 | 2.2E-08 | 6.90E-05 | 31 |
| rs12484438 | 22 | 40558064 | TNRC6B | T | C | 0.661 | 0.013 | 0.001 | 6.7E-19 | 1.74E-04 | 79 |
| rs738140 | 22 | 41884954 | ACO2 | A | G | 0.690 | 0.009 | 0.001 | 2.6E-09 | 7.82E-05 | 35 |
| rs9615723 | 22 | 48386670 | FAM19A5 | C | T | 0.441 | 0.008 | 0.001 | 3.8E-08 | 6.67E-05 | 30 |
| rs34778589 | 22 | 50709957 | MAPK11 | A | C | 0.917 | -0.014 | 0.003 | 9.8E-09 | 7.26E-05 | 33 |
| Genome-wide significant SNPs for adult body size in UK Biobank (men only) | | | | | | | | | | | |
| SNP | Chr | Position | Closest gene | EA | OA | EAF | Beta | SE | pvalue | $R^2$ | F statistic |
| rs3762444 | 1 | 2427712 | PLCH2 | C | T | 0.546 | 0.012 | 0.002 | 2.1E-10 | 1.95E-04 | 40 |
| rs1284373 | 1 | 33355923 | HPCA | C | T | 0.801 | 0.014 | 0.002 | 6.9E-09 | 1.62E-04 | 34 |
| rs34052145 | 1 | 38824262 | POU3F1 | A | G | 0.647 | -0.011 | 0.002 | 1.4E-08 | 1.56E-04 | 32 |
| rs1167311 | 1 | 49996959 | AGBL4 | G | A | 0.319 | 0.012 | 0.002 | 7E-09 | 1.62E-04 | 34 |
| rs12140153 | 1 | 62579891 | INADL | G | T | 0.905 | 0.024 | 0.003 | 1.1E-12 | 2.45E-04 | 51 |
| rs11208779 | 1 | 66417145 | PDE4B | G | C | 0.471 | -0.012 | 0.002 | 1.9E-09 | 1.75E-04 | 36 |
| rs61765650 | 1 | 72753112 | NEGR1 | A | G | 0.810 | 0.022 | 0.002 | 2.8E-19 | 3.90E-04 | 81 |
| rs34517439 | 1 | 78450517 | DNAJB4 | C | A | 0.879 | -0.024 | 0.003 | 3.5E-16 | 3.22E-04 | 67 |
| rs2181375 | 1 | 96940119 | PTBP2 | A | G | 0.403 | -0.012 | 0.002 | 2.6E-10 | 1.93E-04 | 40 |
| rs17024258 | 1 | 110147321 | GNAT2 | C | T | 0.974 | -0.047 | 0.006 | 1.4E-14 | 2.86E-04 | 59 |
| rs984225 | 1 | 119504284 | TBX15 | G | A | 0.389 | 0.011 | 0.002 | 3.4E-08 | 1.47E-04 | 30 |
| rs61813324 | 1 | 156049877 | MEX3A | C | T | 0.864 | -0.016 | 0.003 | 2.4E-08 | 1.51E-04 | 31 |
| rs543874 | 1 | 177889480 | SEC16B | A | G | 0.794 | -0.022 | 0.002 | 1.8E-20 | 4.16E-04 | 86 |
| rs2125232 | 1 | 243759210 | AKT3 | C | T | 0.318 | 0.012 | 0.002 | 2.7E-09 | 1.71E-04 | 35 |
| rs1529897 | 2 | 25086827 | ADCY3 | T | G | 0.569 | -0.016 | 0.002 | 1E-16 | 3.34E-04 | 69 |
| rs935166 | 2 | 26949366 | KCNK3 | G | A | 0.493 | 0.013 | 0.002 | 2.5E-11 | 2.16E-04 | 45 |
| rs1861410 | 2 | 58933591 | FANCL | C | T | 0.446 | 0.013 | 0.002 | 1.5E-11 | 2.20E-04 | 46 |
| rs3552 | 2 | 69698158 | AAK1 | G | A | 0.458 | -0.012 | 0.002 | 2E-09 | 1.74E-04 | 36 |
| rs396354 | 2 | 86850022 | CHMP3 | T | C | 0.285 | 0.012 | 0.002 | 3.1E-08 | 1.48E-04 | 31 |
| rs6753397 | 2 | 105466651 | POU3F3 | C | T | 0.725 | -0.012 | 0.002 | 3E-08 | 1.49E-04 | 31 |
| rs13405033 | 2 | 113994595 | PAX8 | C | T | 0.846 | -0.018 | 0.003 | 5.6E-12 | 2.30E-04 | 47 |
| rs1451077 | 2 | 147901207 | ACVR2A | G | A | 0.414 | 0.013 | 0.002 | 2.7E-11 | 2.15E-04 | 44 |
| rs7581907 | 2 | 205385454 | PARD3B | A | G | 0.127 | 0.016 | 0.003 | 2E-08 | 1.52E-04 | 31 |
| rs6436661 | 2 | 228056670 | COL4A3 | C | T | 0.113 | -0.018 | 0.003 | 2.6E-08 | 1.50E-04 | 31 |
| rs7619139 | 3 | 25110415 | RARB | T | A | 0.411 | -0.012 | 0.002 | 9.2E-10 | 1.81E-04 | 37 |
| rs2526389 | 3 | 50192826 | SEMA3F | C | T | 0.573 | -0.019 | 0.002 | 1E-22 | 4.66E-04 | 96 |
| rs2336558 | 3 | 52886240 | TMEM110 | C | T | 0.384 | 0.012 | 0.002 | 3.8E-10 | 1.90E-04 | 39 |

| | | | | | | | | | | | |
|---|---|---|---|---|---|---|---|---|---|---|---|
| rs11708540 | 3 | 70593081 | FOXP1 | G | A | 0.843 | -0.015 | 0.003 | 1.6E-08 | 1.55E-04 | 32 |
| rs55782528 | 3 | 85893825 | CADM2 | C | A | 0.646 | 0.013 | 0.002 | 1.2E-10 | 2.00E-04 | 41 |
| rs9861443 | 3 | 88196211 | CGGBP1 | A | C | 0.285 | -0.013 | 0.002 | 2.8E-09 | 1.71E-04 | 35 |
| rs13066686 | 3 | 94075026 | NSUN3 | C | A | 0.594 | 0.013 | 0.002 | 1.2E-11 | 2.22E-04 | 46 |
| rs2918217 | 3 | 115546380 | LSAMP | C | T | 0.859 | -0.018 | 0.003 | 5.7E-11 | 2.08E-04 | 43 |
| rs73875019 | 3 | 131521276 | CPNE4 | T | A | 0.883 | 0.019 | 0.003 | 1.1E-10 | 2.02E-04 | 42 |
| rs1320903 | 3 | 131758077 | CPNE4 | G | A | 0.681 | -0.016 | 0.002 | 1.2E-14 | 2.88E-04 | 60 |
| rs10935143 | 3 | 134665159 | EPHB1 | G | A | 0.551 | 0.011 | 0.002 | 2.2E-08 | 1.52E-04 | 31 |
| rs61789562 | 3 | 135926784 | MSL2 | T | C | 0.882 | 0.017 | 0.003 | 1.5E-08 | 1.55E-04 | 32 |
| rs1568488 | 3 | 153657951 | ARHGEF26 | G | C | 0.405 | -0.012 | 0.002 | 2.7E-09 | 1.71E-04 | 35 |
| rs8192675 | 3 | 170724883 | SLC2A2 | T | C | 0.712 | -0.014 | 0.002 | 1.7E-10 | 1.98E-04 | 41 |
| rs894000 | 3 | 183536063 | MAP6D1 | T | C | 0.372 | 0.013 | 0.002 | 3.7E-11 | 2.12E-04 | 44 |
| rs55742087 | 3 | 185830488 | DGKG | C | T | 0.815 | 0.019 | 0.002 | 1.5E-14 | 2.86E-04 | 59 |
| rs10938398 | 4 | 45186139 | GNPDA2 | G | A | 0.565 | -0.021 | 0.002 | 2.3E-28 | 5.90E-04 | 122 |
| rs2537860 | 4 | 55484718 | KIT | A | C | 0.437 | 0.011 | 0.002 | 6E-09 | 1.64E-04 | 34 |
| rs13107325 | 4 | 103188709 | SLC39A8 | C | T | 0.924 | -0.029 | 0.004 | 4.7E-16 | 3.19E-04 | 66 |
| rs1296328 | 4 | 137083193 | PCDH18 | A | C | 0.440 | 0.013 | 0.002 | 7E-12 | 2.28E-04 | 47 |
| rs2307111 | 5 | 75003678 | POC5 | T | C | 0.605 | 0.013 | 0.002 | 3.7E-11 | 2.12E-04 | 44 |
| rs7703782 | 5 | 87938557 | MEF2C | T | A | 0.868 | -0.024 | 0.003 | 7.4E-17 | 3.37E-04 | 70 |
| rs1459843 | 5 | 95867223 | CAST | C | A | 0.392 | 0.012 | 0.002 | 4.2E-09 | 1.67E-04 | 35 |
| rs2591496 | 5 | 170516965 | RANBP17 | G | A | 0.270 | -0.013 | 0.002 | 2.4E-09 | 1.72E-04 | 36 |
| rs9379829 | 6 | 26172219 | HIST1H2BD | C | T | 0.782 | 0.013 | 0.002 | 3.1E-08 | 1.48E-04 | 31 |
| rs2260051 | 6 | 31591918 | PRRC2A | A | T | 0.439 | -0.012 | 0.002 | 1.4E-09 | 1.77E-04 | 37 |
| rs9277992 | 6 | 33312455 | DAXX | G | A | 0.810 | -0.016 | 0.002 | 6.8E-11 | 2.06E-04 | 43 |
| rs9469899 | 6 | 34793124 | UHRF1BP1 | G | A | 0.640 | -0.013 | 0.002 | 8.2E-11 | 2.04E-04 | 42 |
| rs9471333 | 6 | 40362023 | LRFN2 | C | T | 0.447 | 0.012 | 0.002 | 3.8E-10 | 1.90E-04 | 39 |
| rs3798519 | 6 | 50788778 | TFAP2B | A | C | 0.820 | -0.024 | 0.003 | 3.8E-21 | 4.31E-04 | 89 |
| rs74621225 | 6 | 97804399 | MMS22L | A | G | 0.866 | 0.016 | 0.003 | 8.9E-09 | 1.60E-04 | 33 |
| rs9320823 | 6 | 98429337 | MMS22L | T | C | 0.397 | -0.013 | 0.002 | 1.3E-11 | 2.21E-04 | 46 |
| rs9478671 | 6 | 155987825 | NOX3 | A | G | 0.789 | -0.014 | 0.002 | 3.1E-09 | 1.70E-04 | 35 |
| rs34714518 | 6 | 163080778 | PARK2 | G | A | 0.877 | 0.016 | 0.003 | 2.6E-08 | 1.50E-04 | 31 |
| rs12666574 | 7 | 26526960 | KIAA0087 | G | A | 0.327 | -0.012 | 0.002 | 1.7E-08 | 1.54E-04 | 32 |
| rs62457529 | 7 | 32349219 | PDE1C | A | G | 0.891 | -0.019 | 0.003 | 9.4E-10 | 1.81E-04 | 37 |
| rs6962280 | 7 | 44788657 | ZMIZ2 | A | G | 0.442 | -0.012 | 0.002 | 3.8E-10 | 1.90E-04 | 39 |
| rs17145600 | 7 | 73994733 | GTF2IRD1 | C | T | 0.949 | -0.025 | 0.004 | 2E-08 | 1.52E-04 | 31 |
| rs12538826 | 7 | 99030228 | PTCD1 | T | C | 0.885 | 0.017 | 0.003 | 2E-08 | 1.52E-04 | 31 |
| rs10236214 | 7 | 150668070 | KCNH2 | C | T | 0.358 | -0.011 | 0.002 | 2.1E-08 | 1.52E-04 | 31 |
| rs4840941 | 8 | 8216986 | SGK223 | A | G | 0.470 | -0.014 | 0.002 | 2.1E-12 | 2.39E-04 | 49 |
| rs9329197 | 8 | 9259568 | TNKS | A | T | 0.380 | 0.013 | 0.002 | 1.8E-10 | 1.97E-04 | 41 |
| rs3750310 | 8 | 10283426 | MSRA | G | A | 0.488 | 0.012 | 0.002 | 3.6E-10 | 1.90E-04 | 39 |
| rs11780420 | 8 | 11446421 | BLK | G | A | 0.415 | 0.014 | 0.002 | 3.6E-12 | 2.34E-04 | 48 |
| rs7827210 | 8 | 30871656 | PURG | G | A | 0.604 | -0.011 | 0.002 | 1.7E-08 | 1.54E-04 | 32 |
| rs35732620 | 8 | 73445881 | KCNB2 | G | T | 0.425 | 0.012 | 0.002 | 2.6E-09 | 1.71E-04 | 35 |
| rs1812736 | 8 | 76299138 | HNF4G | G | A | 0.174 | -0.014 | 0.003 | 2.3E-08 | 1.51E-04 | 31 |
| rs72674843 | 8 | 95533000 | KIAA1429 | T | C | 0.761 | 0.015 | 0.002 | 2.8E-11 | 2.14E-04 | 44 |

| | | | | | | | | | | | |
|---|---|---|---|---|---|---|---|---|---|---|---|
| rs800526 | 8 | 116845729 | TRPS1 | A | C | 0.223 | -0.014 | 0.002 | 7.1E-10 | 1.84E-04 | 38 |
| rs1412239 | 9 | 28425515 | LINGO2 | C | G | 0.678 | -0.015 | 0.002 | 4.2E-14 | 2.76E-04 | 57 |
| rs10828247 | 10 | 21822856 | MLLT10 | A | G | 0.657 | -0.011 | 0.002 | 2.7E-08 | 1.49E-04 | 31 |
| rs10824218 | 10 | 76421216 | ADK | A | T | 0.563 | 0.011 | 0.002 | 4.7E-08 | 1.44E-04 | 30 |
| rs10883026 | 10 | 99793865 | CRTAC1 | C | T | 0.479 | 0.013 | 0.002 | 1E-10 | 2.02E-04 | 42 |
| rs117597828 | 10 | 102416055 | PAX2 | C | T | 0.782 | -0.014 | 0.002 | 1E-09 | 1.80E-04 | 37 |
| rs4962671 | 10 | 126305434 | LHPP | T | C | 0.532 | 0.011 | 0.002 | 2.7E-08 | 1.49E-04 | 31 |
| rs72867447 | 11 | 13301875 | ARNTL | C | G | 0.428 | -0.011 | 0.002 | 9.5E-09 | 1.59E-04 | 33 |
| rs6265 | 11 | 27679916 | BDNF | C | T | 0.811 | 0.028 | 0.002 | 3E-31 | 6.54E-04 | 135 |
| rs1222216 | 11 | 30346052 | ARL14EP | C | T | 0.773 | 0.014 | 0.002 | 2.6E-09 | 1.71E-04 | 35 |
| rs4755726 | 11 | 43642130 | HSD17B12 | T | G | 0.310 | 0.014 | 0.002 | 2.5E-11 | 2.15E-04 | 45 |
| rs12798028 | 11 | 47604639 | NDUFS3 | C | T | 0.592 | -0.015 | 0.002 | 9.6E-15 | 2.90E-04 | 60 |
| rs7940691 | 11 | 65640906 | EFEMP2 | C | T | 0.361 | 0.013 | 0.002 | 1.8E-11 | 2.19E-04 | 45 |
| rs10898317 | 11 | 84613479 | DLG2 | C | T | 0.487 | 0.011 | 0.002 | 3E-08 | 1.49E-04 | 31 |
| rs55726687 | 12 | 991306 | WNK1 | G | A | 0.792 | -0.013 | 0.002 | 4.7E-08 | 1.44E-04 | 30 |
| rs76895963 | 12 | 4384844 | CCND2 | T | G | 0.979 | -0.042 | 0.007 | 1.3E-08 | 1.57E-04 | 32 |
| rs7132908 | 12 | 50263148 | FAIM2 | G | A | 0.615 | -0.019 | 0.002 | 5.3E-22 | 4.50E-04 | 93 |
| rs4759228 | 12 | 56508409 | PA2G4 | G | C | 0.703 | 0.012 | 0.002 | 2.5E-08 | 1.50E-04 | 31 |
| rs7308188 | 12 | 103701537 | C12orf42 | T | C | 0.745 | 0.014 | 0.002 | 5.3E-10 | 1.87E-04 | 39 |
| rs6490030 | 12 | 116106755 | MED13L | C | A | 0.626 | -0.011 | 0.002 | 3.8E-08 | 1.46E-04 | 30 |
| rs147730268 | 12 | 123024476 | KNTC1 | G | T | 0.913 | 0.022 | 0.003 | 6.1E-10 | 1.85E-04 | 38 |
| rs11619722 | 13 | 27998600 | GTF3A | T | C | 0.699 | 0.012 | 0.002 | 7.1E-09 | 1.62E-04 | 34 |
| rs61954177 | 13 | 40787036 | AL133318.1 | G | C | 0.673 | -0.012 | 0.002 | 2E-08 | 1.53E-04 | 32 |
| rs4477562 | 13 | 54104968 | OLFM4 | C | T | 0.871 | -0.021 | 0.003 | 8E-13 | 2.48E-04 | 51 |
| rs9317002 | 13 | 59175727 | PCDH17 | C | A | 0.486 | -0.015 | 0.002 | 3E-14 | 2.79E-04 | 58 |
| rs7983454 | 13 | 111850539 | ARHGEF7 | T | C | 0.477 | 0.011 | 0.002 | 1.2E-08 | 1.57E-04 | 33 |
| rs9522279 | 13 | 112221296 | RP11-65D24.2 | C | T | 0.577 | -0.013 | 0.002 | 7.3E-11 | 2.05E-04 | 42 |
| rs10132280 | 14 | 25928179 | STXBP6 | C | A | 0.698 | 0.016 | 0.002 | 9.9E-15 | 2.90E-04 | 60 |
| rs9788550 | 14 | 29681138 | PRKD1 | G | C | 0.753 | 0.015 | 0.002 | 2E-11 | 2.18E-04 | 45 |
| rs2143975 | 14 | 33297398 | AKAP6 | C | G | 0.466 | 0.013 | 0.002 | 4.6E-12 | 2.31E-04 | 48 |
| rs10131761 | 14 | 40752651 | FBXO33 | T | A | 0.812 | 0.014 | 0.002 | 4E-08 | 1.46E-04 | 30 |
| rs4898556 | 14 | 47299082 | MDGA2 | A | C | 0.492 | 0.013 | 0.002 | 1.9E-11 | 2.18E-04 | 45 |
| rs217669 | 14 | 62360075 | SYT16 | T | C | 0.728 | -0.014 | 0.002 | 3.4E-10 | 1.91E-04 | 39 |
| rs8008910 | 14 | 79944099 | NRXN3 | G | A | 0.778 | -0.017 | 0.002 | 2.3E-13 | 2.60E-04 | 54 |
| rs8008772 | 14 | 88321884 | GALC | A | T | 0.750 | -0.013 | 0.002 | 1.6E-08 | 1.54E-04 | 32 |
| rs1887197 | 14 | 94187832 | PRIMA1 | C | T | 0.347 | -0.014 | 0.002 | 2.3E-11 | 2.16E-04 | 45 |
| rs61992671 | 14 | 101531854 | AL117190.3 | A | G | 0.508 | 0.011 | 0.002 | 4.9E-08 | 1.44E-04 | 30 |
| rs11631651 | 15 | 73646487 | HCN4 | A | C | 0.927 | 0.022 | 0.004 | 1.3E-09 | 1.78E-04 | 37 |
| rs57488047 | 15 | 79403002 | RASGRF1 | T | C | 0.532 | 0.012 | 0.002 | 3E-09 | 1.70E-04 | 35 |
| rs2238435 | 16 | 4014282 | ADCY9 | C | G | 0.386 | -0.011 | 0.002 | 2.5E-08 | 1.50E-04 | 31 |
| rs1990573 | 16 | 9713688 | RP11-297M9.1 | A | G | 0.306 | -0.013 | 0.002 | 1.7E-09 | 1.76E-04 | 36 |
| rs8054079 | 16 | 19975407 | GPR139 | C | T | 0.860 | 0.016 | 0.003 | 8.8E-09 | 1.60E-04 | 33 |
| rs27741 | 16 | 28504181 | CLN3 | G | A | 0.584 | -0.017 | 0.002 | 8E-19 | 3.80E-04 | 78 |
| rs62048402 | 16 | 53803223 | FTO | G | A | 0.594 | -0.047 | 0.002 | 6.9E-130 | 2.84E-03 | 588 |
| rs12923231 | 16 | 69572892 | NFAT5 | C | T | 0.590 | 0.014 | 0.002 | 3.9E-13 | 2.55E-04 | 53 |

| | | | | | | | | | | | |
|---|---|---|---|---|---|---|---|---|---|---|---|
| rs12149660 | 16 | 70309237 | AARS | G | A | 0.885 | 0.020 | 0.003 | 3E-11 | 2.14E-04 | 44 |
| rs3923783 | 17 | 1843189 | RTN4RL1 | C | A | 0.815 | 0.016 | 0.002 | 1E-10 | 2.02E-04 | 42 |
| rs9901404 | 17 | 21213782 | MAP2K3 | A | G | 0.473 | 0.016 | 0.002 | 1.1E-13 | 2.67E-04 | 55 |
| rs12941009 | 17 | 34961051 | MRM1 | C | T | 0.598 | 0.011 | 0.002 | 1.2E-08 | 1.57E-04 | 32 |
| rs11079849 | 17 | 47090785 | IGF2BP1 | C | T | 0.672 | 0.012 | 0.002 | 6.4E-09 | 1.63E-04 | 34 |
| rs11150745 | 17 | 78757626 | RPTOR | A | G | 0.683 | 0.013 | 0.002 | 6.8E-11 | 2.06E-04 | 43 |
| rs1652376 | 18 | 21109466 | NPC1 | G | T | 0.538 | 0.015 | 0.002 | 2.6E-14 | 2.80E-04 | 58 |
| rs7232171 | 18 | 31251221 | ASXL3 | G | T | 0.418 | -0.011 | 0.002 | 2.2E-08 | 1.51E-04 | 31 |
| rs58243949 | 18 | 52509833 | RAB27B | C | T | 0.767 | -0.014 | 0.002 | 2.3E-09 | 1.73E-04 | 36 |
| rs7240682 | 18 | 57824038 | MC4R | C | G | 0.772 | -0.031 | 0.002 | 8.7E-42 | 8.87E-04 | 183 |
| rs8112818 | 19 | 18812785 | CRTC1 | A | G | 0.601 | 0.012 | 0.002 | 4.6E-09 | 1.66E-04 | 34 |
| rs10423928 | 19 | 46182304 | GIPR | T | A | 0.806 | 0.019 | 0.002 | 1.2E-14 | 2.88E-04 | 60 |
| rs3810291 | 19 | 47569003 | ZC3H4 | G | A | 0.326 | -0.016 | 0.002 | 6.3E-15 | 2.94E-04 | 61 |
| rs6054427 | 20 | 6635266 | BMP2 | G | A | 0.378 | -0.014 | 0.002 | 8.9E-12 | 2.25E-04 | 47 |
| rs6096886 | 20 | 50951298 | ZFP64 | A | G | 0.810 | 0.016 | 0.002 | 2.4E-10 | 1.94E-04 | 40 |
| rs9977825 | 21 | 46494995 | ADARB1 | T | C | 0.361 | 0.012 | 0.002 | 5.8E-09 | 1.64E-04 | 34 |
| rs17421586 | 22 | 40591312 | TNRC6B | T | A | 0.660 | 0.012 | 0.002 | 3.4E-09 | 1.69E-04 | 35 |
| Genome-wide significant SNPs for adult body size in UK Biobank (women only) | | | | | | | | | | | |
| SNP | Chr | Position | Closest gene | EA | OA | EAF | Beta | SE | pvalue | $R^2$ | F statistic |
| rs74892851 | 1 | 1563789 | MIB2 | C | A | 0.623 | 0.012 | 0.002 | 1.9E-08 | 1.28E-04 | 32 |
| rs78886584 | 1 | 16859325 | FAM231B | A | G | 0.509 | -0.013 | 0.002 | 2.6E-11 | 1.80E-04 | 44 |
| rs72660086 | 1 | 39571992 | MACF1 | T | G | 0.789 | -0.013 | 0.002 | 3.2E-08 | 1.24E-04 | 31 |
| rs12144626 | 1 | 47670525 | TAL1 | T | C | 0.417 | 0.012 | 0.002 | 4.4E-09 | 1.40E-04 | 34 |
| rs1494461 | 1 | 49787196 | AGBL4 | C | T | 0.321 | 0.013 | 0.002 | 1.4E-09 | 1.49E-04 | 37 |
| rs12140153 | 1 | 62579891 | INADL | G | T | 0.906 | 0.019 | 0.003 | 2.3E-08 | 1.27E-04 | 31 |
| rs2815757 | 1 | 72764289 | NEGR1 | C | T | 0.191 | -0.017 | 0.002 | 2E-11 | 1.82E-04 | 45 |
| rs1514173 | 1 | 74995110 | TNNI3K | C | T | 0.598 | -0.013 | 0.002 | 1.9E-11 | 1.83E-04 | 45 |
| rs34517439 | 1 | 78450517 | DNAJB4 | C | A | 0.878 | -0.025 | 0.003 | 8.4E-17 | 2.81E-04 | 69 |
| rs10922911 | 1 | 91205831 | BARHL2 | C | T | 0.629 | -0.014 | 0.002 | 1.1E-11 | 1.87E-04 | 46 |
| rs653958 | 1 | 96884006 | PTBP2 | A | G | 0.629 | -0.012 | 0.002 | 1.1E-08 | 1.33E-04 | 33 |
| rs75641275 | 1 | 98327133 | DPYD | A | C | 0.857 | -0.018 | 0.003 | 4.7E-11 | 1.76E-04 | 43 |
| rs41279738 | 1 | 110082551 | GPR61 | T | G | 0.974 | -0.041 | 0.006 | 3.4E-11 | 1.78E-04 | 44 |
| rs12033257 | 1 | 112318484 | KCND3 | A | G | 0.618 | 0.012 | 0.002 | 1.9E-09 | 1.46E-04 | 36 |
| rs3753639 | 1 | 154986091 | ZBTB7B | T | C | 0.756 | -0.013 | 0.002 | 3.7E-08 | 1.23E-04 | 30 |
| rs61813324 | 1 | 156049877 | MEX3A | C | T | 0.865 | -0.021 | 0.003 | 7.6E-13 | 2.08E-04 | 51 |
| rs539515 | 1 | 177889025 | SEC16B | A | C | 0.795 | -0.038 | 0.002 | 9.6E-54 | 9.65E-04 | 238 |
| rs815163 | 1 | 190294726 | BRINP3 | T | C | 0.438 | 0.013 | 0.002 | 2.4E-11 | 1.81E-04 | 45 |
| rs2678204 | 1 | 201800511 | IPO9 | T | G | 0.659 | -0.013 | 0.002 | 1.2E-10 | 1.68E-04 | 42 |
| rs2994320 | 1 | 243641247 | SDCCAG8 | A | G | 0.804 | 0.017 | 0.002 | 2.4E-11 | 1.81E-04 | 45 |
| rs62106258 | 2 | 417167 | FAM150B | T | C | 0.951 | 0.067 | 0.005 | 2E-49 | 8.85E-04 | 218 |
| rs6548237 | 2 | 621461 | TMEM18 | A | C | 0.173 | -0.036 | 0.003 | 3.4E-44 | 7.88E-04 | 194 |
| rs6749422 | 2 | 25150011 | ADCY3 | C | G | 0.514 | -0.024 | 0.002 | 1.2E-34 | 6.11E-04 | 151 |
| rs34606703 | 2 | 47014522 | SOCS5 | G | A | 0.642 | 0.012 | 0.002 | 1.1E-08 | 1.32E-04 | 33 |
| rs13420048 | 2 | 50751414 | NRXN1 | C | A | 0.635 | 0.012 | 0.002 | 4.4E-09 | 1.40E-04 | 34 |

| | | | | | | | | | | | |
|---|---|---|---|---|---|---|---|---|---|---|---|
| rs6545468 | 2 | 55277641 | RTN4 | C | G | 0.587 | 0.012 | 0.002 | 2.2E-09 | 1.45E-04 | 36 |
| rs4671328 | 2 | 58935282 | FANCL | T | G | 0.448 | 0.014 | 0.002 | 6.5E-12 | 1.91E-04 | 47 |
| rs13416992 | 2 | 59298298 | FANCL | A | C | 0.396 | 0.015 | 0.002 | 1.9E-13 | 2.19E-04 | 54 |
| rs10192894 | 2 | 62838936 | EHBP1 | A | G | 0.562 | -0.011 | 0.002 | 2.5E-08 | 1.26E-04 | 31 |
| rs12477088 | 2 | 67841326 | ETAA1 | T | C | 0.590 | 0.012 | 0.002 | 2.7E-09 | 1.43E-04 | 35 |
| rs11691869 | 2 | 100805996 | AFF3 | C | A | 0.637 | 0.016 | 0.002 | 3.3E-14 | 2.33E-04 | 58 |
| rs7602120 | 2 | 144033069 | ARHGAP15 | C | T | 0.534 | -0.013 | 0.002 | 1.2E-11 | 1.87E-04 | 46 |
| rs1083472 | 2 | 147873492 | ACVR2A | C | G | 0.386 | 0.011 | 0.002 | 2.1E-08 | 1.27E-04 | 31 |
| rs4482463 | 2 | 205375909 | PARD3B | C | A | 0.077 | 0.022 | 0.004 | 3.7E-09 | 1.41E-04 | 35 |
| rs4673553 | 2 | 211608379 | CPS1 | T | G | 0.552 | -0.012 | 0.002 | 3.2E-09 | 1.42E-04 | 35 |
| rs2433733 | 2 | 230816703 | FBXO36 | G | A | 0.322 | 0.012 | 0.002 | 2.3E-08 | 1.27E-04 | 31 |
| rs113706999 | 3 | 44159156 | TOPAZ1 | T | A | 0.976 | -0.039 | 0.007 | 1E-08 | 1.33E-04 | 33 |
| rs72906474 | 3 | 47817007 | SMARCC1 | G | T | 0.416 | 0.012 | 0.002 | 1.1E-08 | 1.33E-04 | 33 |
| rs9843653 | 3 | 49920571 | MST1R | T | C | 0.490 | -0.018 | 0.002 | 4.2E-19 | 3.24E-04 | 80 |
| rs6774533 | 3 | 62471086 | CADPS | C | T | 0.296 | -0.012 | 0.002 | 2.8E-08 | 1.25E-04 | 31 |
| rs13066308 | 3 | 82713150 | GBE1 | C | G | 0.639 | -0.012 | 0.002 | 3.7E-09 | 1.41E-04 | 35 |
| rs1454687 | 3 | 94038085 | NSUN3 | C | G | 0.484 | 0.012 | 0.002 | 4.1E-10 | 1.59E-04 | 39 |
| rs13081671 | 3 | 135876549 | MSL2 | C | T | 0.728 | -0.013 | 0.002 | 1.2E-09 | 1.50E-04 | 37 |
| rs2035936 | 3 | 141298124 | RASA2 | G | T | 0.944 | -0.027 | 0.004 | 4.4E-10 | 1.58E-04 | 39 |
| rs529200 | 3 | 173114305 | NLGN1 | A | G | 0.473 | -0.012 | 0.002 | 4E-09 | 1.40E-04 | 35 |
| rs73052033 | 3 | 185828465 | DGKG | T | C | 0.815 | 0.017 | 0.003 | 1.1E-11 | 1.87E-04 | 46 |
| rs61218008 | 3 | 194881130 | XXYLT1 | A | G | 0.721 | 0.012 | 0.002 | 4.7E-08 | 1.21E-04 | 30 |
| rs2643450 | 4 | 18547417 | LCORL | A | G | 0.459 | -0.011 | 0.002 | 4.5E-08 | 1.21E-04 | 30 |
| rs9684942 | 4 | 20233035 | SLIT2 | G | A | 0.853 | -0.015 | 0.003 | 3.8E-08 | 1.23E-04 | 30 |
| rs34811474 | 4 | 25408838 | ANAPC4 | G | A | 0.770 | 0.020 | 0.002 | 3.2E-17 | 2.89E-04 | 71 |
| rs73213484 | 4 | 28489339 | RP11-180C1.1 | A | T | 0.858 | 0.019 | 0.003 | 2.3E-11 | 1.81E-04 | 45 |
| rs4527444 | 4 | 30842780 | PCDH7 | A | G | 0.458 | -0.012 | 0.002 | 3.3E-10 | 1.60E-04 | 39 |
| rs12641981 | 4 | 45179883 | GNPDA2 | C | T | 0.566 | -0.017 | 0.002 | 2.8E-17 | 2.90E-04 | 71 |
| rs148712344 | 4 | 55476318 | KIT | G | T | 0.960 | 0.030 | 0.005 | 5.1E-09 | 1.38E-04 | 34 |
| rs925422 | 4 | 60254101 | LPHN3 | T | G | 0.257 | 0.013 | 0.002 | 2.7E-08 | 1.25E-04 | 31 |
| rs1603179 | 4 | 67805347 | CENPC | A | C | 0.356 | 0.011 | 0.002 | 2.9E-08 | 1.25E-04 | 31 |
| rs11098965 | 4 | 80888040 | ANTXR2 | C | T | 0.293 | 0.014 | 0.002 | 1.6E-09 | 1.48E-04 | 36 |
| rs4148155 | 4 | 89054667 | ABCG2 | A | G | 0.887 | 0.018 | 0.003 | 3.8E-09 | 1.41E-04 | 35 |
| rs13107325 | 4 | 103188709 | SLC39A8 | C | T | 0.926 | -0.029 | 0.004 | 1.9E-14 | 2.38E-04 | 59 |
| rs769668 | 4 | 140858717 | MAML3 | T | C | 0.653 | 0.014 | 0.002 | 1.8E-11 | 1.83E-04 | 45 |
| rs35390852 | 4 | 143067054 | INPP4B | G | A | 0.876 | -0.017 | 0.003 | 2.4E-08 | 1.26E-04 | 31 |
| rs828550 | 5 | 3539923 | IRX1 | C | T | 0.355 | -0.012 | 0.002 | 1.5E-08 | 1.30E-04 | 32 |
| rs10514963 | 5 | 63027870 | HTR1A | G | A | 0.519 | -0.014 | 0.002 | 3E-12 | 1.97E-04 | 49 |
| rs34341 | 5 | 74934009 | ANKDD1B | A | T | 0.425 | -0.020 | 0.002 | 1.1E-23 | 4.08E-04 | 101 |
| rs59893724 | 5 | 80830788 | SSBP2 | A | G | 0.756 | 0.013 | 0.002 | 2.4E-08 | 1.26E-04 | 31 |
| rs7442885 | 5 | 87682877 | TMEM161B | C | G | 0.786 | 0.017 | 0.002 | 1.5E-12 | 2.03E-04 | 50 |
| rs1477290 | 5 | 87988934 | MEF2C | T | C | 0.863 | -0.017 | 0.003 | 1.4E-09 | 1.49E-04 | 37 |
| rs10038055 | 5 | 88783421 | MEF2C | G | T | 0.632 | -0.012 | 0.002 | 8.3E-09 | 1.35E-04 | 33 |
| rs288187 | 5 | 107344426 | FBXL17 | C | T | 0.827 | 0.020 | 0.003 | 4E-14 | 2.32E-04 | 57 |
| rs1366334 | 5 | 122683163 | CEP120 | C | G | 0.293 | 0.012 | 0.002 | 4.7E-08 | 1.21E-04 | 30 |

| | | | | | | | | | | | |
|---|---|---|---|---|---|---|---|---|---|---|---|
| rs13174863 | 5 | 139080745 | CXXC5 | A | G | 0.852 | -0.019 | 0.003 | 1.8E-11 | 1.83E-04 | 45 |
| rs251353 | 5 | 140228164 | PCDHA1 | C | A | 0.471 | 0.012 | 0.002 | 1.1E-08 | 1.33E-04 | 33 |
| rs11134679 | 5 | 170623391 | RANBP17 | A | G | 0.315 | -0.013 | 0.002 | 8E-10 | 1.53E-04 | 38 |
| rs9395520 | 6 | 13183523 | PHACTR1 | C | T | 0.695 | 0.013 | 0.002 | 4.6E-09 | 1.39E-04 | 34 |
| rs35778344 | 6 | 28599105 | SCAND3 | G | A | 0.803 | -0.014 | 0.002 | 1.8E-08 | 1.29E-04 | 32 |
| rs3130048 | 6 | 31613739 | BAG6 | T | C | 0.720 | -0.016 | 0.002 | 4.6E-13 | 2.12E-04 | 52 |
| rs34298980 | 6 | 40409243 | LRFN2 | T | C | 0.492 | 0.014 | 0.002 | 8.8E-12 | 1.89E-04 | 47 |
| rs72892910 | 6 | 50816887 | TFAP2B | G | T | 0.828 | -0.026 | 0.003 | 9.7E-24 | 4.09E-04 | 101 |
| rs1547026 | 6 | 51825622 | PKHD1 | T | C | 0.709 | -0.012 | 0.002 | 1.6E-08 | 1.29E-04 | 32 |
| rs2253310 | 6 | 108888593 | FOXO3 | C | G | 0.374 | -0.012 | 0.002 | 6.1E-09 | 1.37E-04 | 34 |
| rs9387640 | 6 | 119508871 | MAN1A1 | C | T | 0.637 | 0.012 | 0.002 | 9.2E-09 | 1.34E-04 | 33 |
| rs73046311 | 7 | 1854159 | MAD1L1 | C | G | 0.839 | 0.016 | 0.003 | 5.7E-09 | 1.38E-04 | 34 |
| rs2866720 | 7 | 70106310 | AUTS2 | C | T | 0.615 | -0.012 | 0.002 | 7E-09 | 1.36E-04 | 34 |
| rs11976018 | 7 | 99122437 | ZKSCAN5 | G | A | 0.846 | 0.015 | 0.003 | 2.8E-08 | 1.25E-04 | 31 |
| rs12375196 | 7 | 103416541 | RELN | C | A | 0.576 | -0.012 | 0.002 | 5.5E-09 | 1.38E-04 | 34 |
| rs2396625 | 7 | 113028634 | TSRM | T | A | 0.578 | 0.012 | 0.002 | 3.4E-09 | 1.42E-04 | 35 |
| rs1840661 | 7 | 114352682 | FOXP2 | T | A | 0.575 | -0.012 | 0.002 | 1.2E-09 | 1.50E-04 | 37 |
| rs7853 | 8 | 8890814 | ERI1 | A | G | 0.553 | -0.012 | 0.002 | 1.3E-09 | 1.50E-04 | 37 |
| rs11250094 | 8 | 10802001 | XKR6 | G | C | 0.453 | 0.012 | 0.002 | 1.1E-09 | 1.51E-04 | 37 |
| rs6557829 | 8 | 21973970 | HR | C | A | 0.595 | -0.012 | 0.002 | 6.9E-09 | 1.36E-04 | 34 |
| rs117176448 | 8 | 27261138 | PTK2B | C | G | 0.904 | -0.019 | 0.003 | 2E-08 | 1.28E-04 | 32 |
| rs10957605 | 8 | 73433886 | KCNB2 | C | T | 0.318 | 0.016 | 0.002 | 9.8E-14 | 2.25E-04 | 55 |
| rs17716502 | 8 | 116659731 | TRPS1 | C | T | 0.795 | 0.018 | 0.002 | 3.5E-13 | 2.15E-04 | 53 |
| rs4740442 | 9 | 10153245 | PTPRD | C | T | 0.715 | -0.012 | 0.002 | 2.6E-08 | 1.26E-04 | 31 |
| rs13292699 | 9 | 15910044 | CCDC171 | A | C | 0.567 | 0.015 | 0.002 | 4.3E-14 | 2.31E-04 | 57 |
| rs17770336 | 9 | 28414625 | LINGO2 | C | T | 0.676 | -0.016 | 0.002 | 2.5E-14 | 2.36E-04 | 58 |
| rs2398851 | 9 | 96398508 | PHF2 | A | G | 0.320 | 0.012 | 0.002 | 8E-09 | 1.35E-04 | 33 |
| rs7047694 | 9 | 103141037 | TEX10 | G | A | 0.681 | -0.012 | 0.002 | 3.7E-09 | 1.41E-04 | 35 |
| rs6478538 | 9 | 124627012 | TTLL11 | A | G | 0.323 | 0.012 | 0.002 | 4.6E-08 | 1.21E-04 | 30 |
| rs777676 | 9 | 129691913 | RALGPS1 | T | A | 0.511 | 0.011 | 0.002 | 3.7E-08 | 1.23E-04 | 30 |
| rs3003578 | 9 | 130994179 | DNM1 | C | T | 0.441 | 0.011 | 0.002 | 1.8E-08 | 1.29E-04 | 32 |
| rs1270799 | 10 | 21907423 | MLLT10 | T | G | 0.696 | -0.017 | 0.002 | 1.2E-14 | 2.42E-04 | 60 |
| rs113585475 | 10 | 33985434 | NRP1 | C | T | 0.900 | 0.022 | 0.003 | 3.6E-11 | 1.78E-04 | 44 |
| rs3125326 | 10 | 63053788 | TMEM26 | A | C | 0.391 | -0.011 | 0.002 | 4.7E-08 | 1.21E-04 | 30 |
| rs7090758 | 10 | 65335315 | REEP3 | T | C | 0.526 | -0.013 | 0.002 | 5.7E-11 | 1.74E-04 | 43 |
| rs11000942 | 10 | 76025256 | ADK | G | A | 0.876 | -0.016 | 0.003 | 4E-08 | 1.22E-04 | 30 |
| rs1250535 | 10 | 81016112 | ZMIZ1 | C | G | 0.325 | -0.012 | 0.002 | 1.8E-08 | 1.28E-04 | 32 |
| rs17399739 | 10 | 87490850 | GRID1 | A | G | 0.931 | -0.023 | 0.004 | 6.1E-09 | 1.37E-04 | 34 |
| rs10510025 | 10 | 118650996 | KIAA1598 | C | T | 0.753 | -0.013 | 0.002 | 6.9E-09 | 1.36E-04 | 34 |
| rs4962725 | 10 | 126733321 | CTBP2 | T | C | 0.572 | -0.011 | 0.002 | 2E-08 | 1.28E-04 | 31 |
| rs11146233 | 10 | 134000962 | DPYSL4 | G | A | 0.439 | 0.011 | 0.002 | 9.4E-09 | 1.34E-04 | 33 |
| rs7950166 | 11 | 8642218 | TRIM66 | C | T | 0.354 | -0.014 | 0.002 | 3E-12 | 1.97E-04 | 49 |
| rs11022766 | 11 | 13348249 | ARNTL | T | G | 0.641 | 0.014 | 0.002 | 2.5E-11 | 1.81E-04 | 45 |
| rs1013402 | 11 | 27712381 | BDNF | A | G | 0.681 | -0.021 | 0.002 | 6E-24 | 4.13E-04 | 102 |
| rs34292685 | 11 | 64049021 | GPR137 | C | T | 0.839 | 0.018 | 0.003 | 1.6E-11 | 1.84E-04 | 45 |

| | | | | | | | | | | | |
|---|---|---|---|---|---|---|---|---|---|---|---|
| rs10896012 | 11 | 65278461 | SCYL1 | T | C | 0.784 | -0.016 | 0.002 | 3.7E-11 | 1.77E-04 | 44 |
| rs3802858 | 11 | 115078492 | CADM1 | T | C | 0.573 | 0.012 | 0.002 | 1.1E-09 | 1.51E-04 | 37 |
| rs11218510 | 11 | 121922587 | BLID | G | A | 0.599 | 0.011 | 0.002 | 2.9E-08 | 1.25E-04 | 31 |
| rs2512884 | 11 | 131467856 | NTM | C | A | 0.484 | -0.012 | 0.002 | 3.3E-10 | 1.60E-04 | 40 |
| rs11223204 | 11 | 132652554 | OPCML | A | G | 0.565 | -0.011 | 0.002 | 1.1E-08 | 1.32E-04 | 33 |
| rs12364470 | 11 | 134601012 | AP003062.1 | T | G | 0.836 | -0.015 | 0.003 | 2.9E-08 | 1.25E-04 | 31 |
| rs55726687 | 12 | 991306 | WNK1 | G | A | 0.789 | -0.015 | 0.002 | 1.3E-09 | 1.49E-04 | 37 |
| rs7976757 | 12 | 19207948 | PLEKHA5 | T | C | 0.173 | -0.015 | 0.003 | 6.1E-09 | 1.37E-04 | 34 |
| rs7132908 | 12 | 50263148 | FAIM2 | G | A | 0.616 | -0.019 | 0.002 | 8.2E-21 | 3.55E-04 | 88 |
| rs2292238 | 12 | 56493822 | ERBB3 | A | C | 0.593 | 0.012 | 0.002 | 1.3E-09 | 1.50E-04 | 37 |
| rs770082 | 12 | 89776485 | DUSP6 | G | A | 0.563 | -0.013 | 0.002 | 2.1E-11 | 1.82E-04 | 45 |
| rs10849900 | 12 | 110974890 | PPTC7 | T | C | 0.690 | 0.012 | 0.002 | 1.6E-08 | 1.29E-04 | 32 |
| rs111828690 | 12 | 117576767 | FBXO21 | C | T | 0.779 | -0.014 | 0.002 | 1E-08 | 1.33E-04 | 33 |
| rs181617194 | 12 | 122011598 | KDM2B | T | C | 0.959 | 0.032 | 0.005 | 4.2E-09 | 1.40E-04 | 35 |
| rs3803005 | 12 | 123110654 | KNTC1 | T | C | 0.270 | -0.017 | 0.002 | 3.6E-14 | 2.33E-04 | 57 |
| rs9579775 | 13 | 20616557 | ZMYM2 | A | C | 0.864 | -0.019 | 0.003 | 1.4E-10 | 1.67E-04 | 41 |
| rs1933440 | 13 | 28676971 | FLT3 | A | C | 0.837 | -0.015 | 0.003 | 1.2E-08 | 1.32E-04 | 33 |
| rs2761366 | 13 | 33011872 | N4BP2L2 | C | T | 0.358 | -0.011 | 0.002 | 3.2E-08 | 1.24E-04 | 31 |
| rs9568867 | 13 | 54107352 | OLFM4 | G | A | 0.871 | -0.019 | 0.003 | 6.8E-11 | 1.73E-04 | 43 |
| rs12866691 | 13 | 58623783 | PCDH17 | A | T | 0.773 | 0.016 | 0.002 | 3.9E-11 | 1.77E-04 | 44 |
| rs1576655 | 13 | 79587841 | RBM26 | A | C | 0.404 | -0.012 | 0.002 | 2.4E-09 | 1.45E-04 | 36 |
| rs7331420 | 13 | 99236471 | STK24 | G | A | 0.715 | 0.012 | 0.002 | 4.8E-08 | 1.21E-04 | 30 |
| rs9522180 | 13 | 111970212 | TEX29 | C | T | 0.447 | 0.011 | 0.002 | 4.4E-08 | 1.21E-04 | 30 |
| rs10142359 | 14 | 73884540 | NUMB | A | G | 0.518 | -0.011 | 0.002 | 1.7E-08 | 1.29E-04 | 32 |
| rs8022132 | 14 | 79955864 | NRXN3 | A | T | 0.697 | -0.015 | 0.002 | 1E-11 | 1.88E-04 | 46 |
| rs6575340 | 14 | 94023972 | UNC79 | G | A | 0.365 | -0.015 | 0.002 | 1.1E-12 | 2.05E-04 | 51 |
| rs7145882 | 14 | 103255461 | TRAF3 | T | C | 0.342 | 0.012 | 0.002 | 6E-09 | 1.37E-04 | 34 |
| rs12891477 | 14 | 104332759 | PPP1R13B | C | T | 0.631 | -0.013 | 0.002 | 1.8E-10 | 1.65E-04 | 41 |
| rs1466276 | 15 | 52025950 | LYSMD2 | C | G | 0.524 | 0.011 | 0.002 | 1.9E-08 | 1.28E-04 | 32 |
| rs4776985 | 15 | 68123021 | SKOR1 | T | G | 0.774 | 0.020 | 0.002 | 8.9E-17 | 2.81E-04 | 69 |
| rs67962220 | 15 | 74188926 | TBC1D21 | T | G | 0.831 | -0.015 | 0.003 | 9.8E-09 | 1.33E-04 | 33 |
| rs715724 | 15 | 80984293 | ABHD17C | A | G | 0.649 | 0.012 | 0.002 | 1.4E-08 | 1.30E-04 | 32 |
| rs939624 | 15 | 99480551 | IGF1R | C | T | 0.538 | -0.012 | 0.002 | 4.7E-10 | 1.57E-04 | 39 |
| rs7200589 | 16 | 349331 | AXIN1 | G | A | 0.727 | 0.016 | 0.002 | 2E-12 | 2.01E-04 | 49 |
| rs879620 | 16 | 4015729 | ADCY9 | C | T | 0.387 | -0.017 | 0.002 | 9.7E-18 | 2.98E-04 | 74 |
| rs57790054 | 16 | 20006986 | GPR139 | A | G | 0.739 | -0.012 | 0.002 | 3.7E-08 | 1.23E-04 | 30 |
| rs11074452 | 16 | 20370168 | PDILT | C | G | 0.492 | 0.015 | 0.002 | 8.3E-14 | 2.26E-04 | 56 |
| rs62031562 | 16 | 28609329 | SULT1A2 | A | T | 0.626 | -0.018 | 0.002 | 1.3E-19 | 3.33E-04 | 82 |
| rs3814883 | 16 | 29994922 | TAOK2 | C | T | 0.517 | -0.015 | 0.002 | 3.5E-14 | 2.33E-04 | 57 |
| rs34898535 | 16 | 31025641 | STX1B | C | T | 0.623 | 0.019 | 0.002 | 2.8E-21 | 3.64E-04 | 90 |
| rs56094641 | 16 | 53806453 | FTO | A | G | 0.597 | -0.046 | 0.002 | 4.3E-114 | 2.09E-03 | 515 |
| rs3751859 | 16 | 81735012 | CMIP | G | A | 0.843 | 0.017 | 0.003 | 2.4E-10 | 1.63E-04 | 40 |
| rs4790292 | 17 | 1824305 | RTN4RL1 | C | A | 0.846 | 0.017 | 0.003 | 1.8E-10 | 1.65E-04 | 41 |
| rs1914889 | 17 | 21267590 | KCNJ12 | G | A | 0.547 | 0.012 | 0.002 | 7.8E-10 | 1.53E-04 | 38 |
| rs2306593 | 17 | 34866546 | MYO19 | C | T | 0.510 | 0.011 | 0.002 | 7.2E-09 | 1.36E-04 | 33 |

| | | | | | | | | | | | |
|---|---|---|---|---|---|---|---|---|---|---|---|
| rs11079849 | 17 | 47090785 | IGF2BP1 | C | T | 0.671 | 0.013 | 0.002 | 1.6E-09 | 1.48E-04 | 36 |
| rs77706698 | 17 | 65953348 | BPTF | G | A | 0.868 | -0.017 | 0.003 | 4.2E-09 | 1.40E-04 | 35 |
| rs2619976 | 17 | 71754545 | SDK2 | C | T | 0.587 | -0.012 | 0.002 | 5.4E-09 | 1.38E-04 | 34 |
| rs11150745 | 17 | 78757626 | RPTOR | A | G | 0.682 | 0.014 | 0.002 | 1.5E-11 | 1.85E-04 | 46 |
| rs891386 | 18 | 21103971 | NPC1 | T | G | 0.542 | 0.012 | 0.002 | 3.4E-09 | 1.42E-04 | 35 |
| rs11660335 | 18 | 22154235 | HRH4 | T | C | 0.810 | 0.016 | 0.003 | 2.5E-10 | 1.62E-04 | 40 |
| rs784257 | 18 | 53397199 | TCF4 | T | C | 0.188 | -0.014 | 0.003 | 2.9E-08 | 1.25E-04 | 31 |
| rs66922415 | 18 | 57848651 | MC4R | A | G | 0.767 | -0.039 | 0.002 | 7.4E-62 | 1.12E-03 | 275 |
| rs17066856 | 18 | 58049656 | MC4R | T | C | 0.909 | 0.028 | 0.003 | 2.9E-16 | 2.71E-04 | 67 |
| rs9962947 | 18 | 72903636 | ZADH2 | C | T | 0.362 | -0.011 | 0.002 | 4.6E-08 | 1.21E-04 | 30 |
| rs12974664 | 19 | 1856186 | KLF16 | G | A | 0.464 | -0.012 | 0.002 | 6E-10 | 1.55E-04 | 38 |
| rs350832 | 19 | 4069426 | ZBTB7A | G | A | 0.229 | -0.014 | 0.002 | 6.5E-09 | 1.37E-04 | 34 |
| rs12986231 | 19 | 18469017 | PGPEP1 | T | C | 0.732 | 0.015 | 0.002 | 6.2E-11 | 1.73E-04 | 43 |
| rs10404726 | 19 | 18834514 | CRTC1 | C | T | 0.534 | 0.014 | 0.002 | 2.5E-12 | 1.99E-04 | 49 |
| rs56212061 | 19 | 19394640 | SUGP1 | C | T | 0.849 | 0.015 | 0.003 | 3.9E-08 | 1.23E-04 | 30 |
| rs111640872 | 19 | 30290357 | CCNE1 | G | C | 0.669 | -0.013 | 0.002 | 1.8E-09 | 1.47E-04 | 36 |
| rs11880064 | 19 | 33964181 | PEPD | T | C | 0.636 | -0.011 | 0.002 | 4.8E-08 | 1.21E-04 | 30 |
| rs429358 | 19 | 45411941 | APOE | T | C | 0.846 | 0.018 | 0.003 | 1.4E-10 | 1.67E-04 | 41 |
| rs12971645 | 19 | 45807945 | MARK4 | G | A | 0.725 | 0.012 | 0.002 | 4.3E-08 | 1.22E-04 | 30 |
| rs1800437 | 19 | 46181392 | GIPR | G | C | 0.805 | 0.023 | 0.002 | 1.8E-20 | 3.49E-04 | 86 |
| rs3810291 | 19 | 47569003 | ZC3H4 | G | A | 0.324 | -0.015 | 0.002 | 4.7E-13 | 2.12E-04 | 52 |
| rs8124896 | 20 | 21385659 | NKX2-4 | T | C | 0.899 | -0.020 | 0.003 | 6.9E-10 | 1.54E-04 | 38 |
| rs116948922 | 20 | 25534854 | NINL | C | T | 0.967 | 0.034 | 0.006 | 1.5E-09 | 1.48E-04 | 37 |
| rs34966255 | 20 | 51193862 | ZFP64 | T | C | 0.808 | 0.016 | 0.003 | 6.2E-11 | 1.73E-04 | 43 |
| rs915814 | 21 | 46493003 | AP001579.1 | G | A | 0.761 | 0.013 | 0.002 | 1.5E-08 | 1.30E-04 | 32 |
| rs400997 | 21 | 46564154 | ADARB1 | T | A | 0.441 | -0.014 | 0.002 | 8E-13 | 2.08E-04 | 51 |
| rs738140 | 22 | 41884954 | ACO2 | A | G | 0.689 | 0.012 | 0.002 | 1.80E-08 | 1.28E-04 | 32 |
| Abbreviations: Chr Chromosome; EA effect allele; EAF effect allele frequency; OA other allele; SE standard error | | | | | | | | | | | |

| Supplementary table 2: Summary information on colorectal cancer risk for the SNPs used in the analysis | | | | | | | | | | | | |
|---|---|---|---|---|---|---|---|---|---|---|---|---|
| Early life body size SNPs (overall) | | | | | | | | | | | | |
| SNP | EA | OA | beta_ca | se_ca | beta_co | se_co | beta_prox | se_prox | beta_dist | se_dist | beta_re | se_re |
| rs2229330 | T | G | -0.018 | 0.021 | -0.024 | 0.025 | -0.029 | 0.031 | -0.019 | 0.032 | -0.019 | 0.033 |
| rs2175171 | G | C | 0.005 | 0.010 | 0.000 | 0.012 | -0.001 | 0.015 | -0.006 | 0.015 | -0.001 | 0.015 |
| rs6577497 | A | T | -0.007 | 0.010 | -0.004 | 0.012 | 0.009 | 0.015 | -0.012 | 0.016 | -0.014 | 0.016 |
| rs12045879 | C | T | -0.018 | 0.010 | -0.014 | 0.013 | -0.014 | 0.016 | -0.012 | 0.016 | -0.014 | 0.016 |
| rs212517 | T | A | 0.009 | 0.010 | 0.003 | 0.012 | 0.013 | 0.015 | -0.019 | 0.015 | 0.011 | 0.015 |
| rs2356864 | G | A | 0.009 | 0.010 | 0.022 | 0.012 | 0.035 | 0.015 | 0.014 | 0.015 | -0.003 | 0.015 |
| rs630602 | G | C | -0.004 | 0.010 | -0.014 | 0.012 | -0.007 | 0.015 | -0.017 | 0.016 | 0.015 | 0.016 |
| rs12140153 | G | T | 0.015 | 0.019 | 0.026 | 0.022 | 0.041 | 0.028 | -0.003 | 0.029 | -0.024 | 0.029 |
| rs2767486 | A | G | -0.013 | 0.012 | -0.016 | 0.014 | -0.004 | 0.018 | -0.029 | 0.018 | -0.022 | 0.019 |
| rs7522014 | A | G | -0.008 | 0.011 | 0.007 | 0.013 | -0.011 | 0.016 | 0.025 | 0.016 | -0.015 | 0.016 |
| rs2755253 | C | T | 0.033 | 0.011 | 0.029 | 0.013 | 0.026 | 0.016 | 0.038 | 0.017 | 0.042 | 0.017 |
| rs11209943 | A | G | 0.017 | 0.010 | 0.011 | 0.012 | 0.009 | 0.015 | 0.005 | 0.016 | 0.035 | 0.016 |
| rs12042908 | A | G | 0.001 | 0.010 | 0.011 | 0.012 | 0.010 | 0.015 | 0.019 | 0.015 | 0.006 | 0.015 |
| rs34517439 | C | A | -0.007 | 0.017 | -0.016 | 0.020 | -0.011 | 0.025 | -0.023 | 0.026 | -0.015 | 0.026 |
| rs11165687 | C | T | -0.014 | 0.010 | -0.016 | 0.012 | -0.006 | 0.015 | -0.024 | 0.015 | -0.015 | 0.015 |
| rs7550711 | C | T | 0.003 | 0.029 | -0.012 | 0.034 | -0.014 | 0.044 | -0.004 | 0.045 | -0.004 | 0.044 |
| rs3013431 | C | T | -0.008 | 0.010 | -0.003 | 0.012 | -0.020 | 0.015 | 0.013 | 0.015 | -0.031 | 0.016 |
| rs12132598 | A | G | -0.017 | 0.010 | -0.015 | 0.012 | -0.005 | 0.015 | -0.021 | 0.016 | -0.028 | 0.016 |
| rs7536458 | T | G | 0.031 | 0.011 | 0.030 | 0.013 | 0.042 | 0.017 | 0.012 | 0.017 | 0.042 | 0.017 |
| rs11205303 | T | C | 0.011 | 0.010 | 0.031 | 0.012 | 0.017 | 0.016 | 0.037 | 0.016 | 0.004 | 0.016 |
| rs35588936 | C | T | -0.005 | 0.019 | -0.002 | 0.022 | 0.021 | 0.028 | -0.026 | 0.028 | 0.016 | 0.029 |
| rs12748436 | C | G | -0.027 | 0.018 | -0.006 | 0.021 | -0.030 | 0.027 | 0.007 | 0.028 | -0.030 | 0.028 |
| rs543874 | A | G | -0.003 | 0.012 | 0.002 | 0.015 | -0.005 | 0.019 | 0.006 | 0.019 | 0.002 | 0.019 |
| rs78444298 | G | A | 0.014 | 0.042 | 0.038 | 0.050 | 0.072 | 0.063 | 0.008 | 0.065 | -0.009 | 0.065 |
| rs4074404 | T | A | -0.018 | 0.013 | -0.015 | 0.016 | -0.014 | 0.020 | -0.019 | 0.020 | -0.025 | 0.020 |
| rs16839832 | G | T | 0.014 | 0.018 | 0.009 | 0.022 | -0.005 | 0.027 | 0.029 | 0.029 | 0.001 | 0.029 |
| rs9438393 | A | G | 0.033 | 0.010 | 0.033 | 0.012 | 0.021 | 0.015 | 0.043 | 0.015 | 0.040 | 0.015 |
| rs7354849 | A | G | -0.009 | 0.010 | -0.005 | 0.012 | -0.003 | 0.015 | -0.008 | 0.015 | -0.017 | 0.015 |
| rs62106258 | T | C | 0.046 | 0.026 | 0.050 | 0.030 | 0.047 | 0.038 | 0.068 | 0.040 | 0.043 | 0.040 |
| rs12992672 | G | A | -0.018 | 0.013 | -0.024 | 0.015 | -0.022 | 0.019 | -0.024 | 0.020 | -0.023 | 0.020 |
| rs2867116 | C | A | -0.012 | 0.015 | -0.015 | 0.017 | -0.014 | 0.022 | -0.027 | 0.022 | -0.029 | 0.022 |
| rs2141004 | A | C | 0.010 | 0.011 | 0.009 | 0.013 | 0.001 | 0.016 | 0.016 | 0.017 | 0.015 | 0.017 |
| rs10182458 | A | G | -0.005 | 0.010 | -0.011 | 0.012 | -0.018 | 0.015 | -0.009 | 0.015 | -0.013 | 0.015 |
| rs146910503 | G | A | 0.075 | 0.037 | 0.082 | 0.044 | 0.110 | 0.057 | 0.039 | 0.058 | 0.011 | 0.056 |
| rs6719507 | G | A | -0.007 | 0.010 | 0.005 | 0.012 | 0.016 | 0.015 | -0.002 | 0.015 | -0.018 | 0.015 |
| rs62134189 | A | G | -0.006 | 0.017 | -0.007 | 0.020 | -0.012 | 0.025 | -0.005 | 0.026 | -0.044 | 0.026 |
| rs2902142 | C | T | -0.016 | 0.010 | -0.007 | 0.012 | -0.002 | 0.015 | -0.010 | 0.015 | -0.006 | 0.016 |
| rs2539692 | T | A | 0.003 | 0.010 | -0.003 | 0.012 | -0.001 | 0.015 | -0.002 | 0.015 | 0.002 | 0.016 |
| rs1177279 | A | G | 0.005 | 0.011 | 0.027 | 0.013 | 0.011 | 0.017 | 0.042 | 0.017 | -0.017 | 0.017 |
| rs7565437 | T | C | 0.007 | 0.010 | 0.006 | 0.012 | 0.006 | 0.015 | 0.006 | 0.015 | 0.019 | 0.015 |
| rs12713889 | T | C | -0.001 | 0.010 | 0.002 | 0.012 | -0.002 | 0.015 | 0.005 | 0.016 | 0.001 | 0.016 |
| rs772175 | G | A | -0.004 | 0.010 | -0.007 | 0.012 | -0.023 | 0.016 | 0.012 | 0.016 | -0.005 | 0.016 |

| | | | | | | | | | | | |
|---|---|---|---|---|---|---|---|---|---|---|---|
| rs1384660 | G | A | -0.015 | 0.012 | -0.015 | 0.015 | 0.003 | 0.019 | -0.040 | 0.019 | -0.032 | 0.020 |
| rs62175963 | T | C | 0.014 | 0.010 | 0.021 | 0.012 | 0.004 | 0.015 | 0.034 | 0.016 | 0.004 | 0.016 |
| rs115319174 | G | C | -0.025 | 0.022 | -0.011 | 0.026 | 0.021 | 0.033 | -0.038 | 0.033 | -0.033 | 0.034 |
| rs11891707 | T | C | 0.008 | 0.014 | 0.006 | 0.017 | -0.006 | 0.021 | 0.011 | 0.022 | 0.005 | 0.022 |
| rs3791478 | T | C | -0.006 | 0.016 | -0.014 | 0.019 | -0.016 | 0.024 | -0.007 | 0.025 | -0.014 | 0.025 |
| rs1476698 | A | G | 0.010 | 0.010 | 0.012 | 0.012 | 0.000 | 0.015 | 0.027 | 0.016 | 0.011 | 0.016 |
| rs2594994 | T | A | 0.007 | 0.013 | 0.003 | 0.015 | -0.004 | 0.019 | 0.013 | 0.019 | 0.036 | 0.020 |
| rs7619139 | T | A | 0.001 | 0.010 | 0.002 | 0.012 | -0.005 | 0.015 | 0.007 | 0.015 | -0.012 | 0.015 |
| rs1402989 | C | T | -0.001 | 0.010 | 0.003 | 0.012 | 0.009 | 0.015 | -0.003 | 0.015 | -0.001 | 0.015 |
| rs2268762 | A | G | 0.014 | 0.010 | 0.017 | 0.012 | 0.023 | 0.015 | 0.007 | 0.015 | -0.012 | 0.016 |
| rs754635 | C | G | -0.016 | 0.015 | -0.021 | 0.018 | -0.012 | 0.023 | -0.026 | 0.024 | -0.029 | 0.024 |
| rs2034963 | G | C | -0.002 | 0.010 | 0.001 | 0.012 | -0.006 | 0.016 | 0.018 | 0.016 | -0.012 | 0.016 |
| rs35926495 | C | T | -0.012 | 0.010 | -0.014 | 0.012 | -0.014 | 0.015 | -0.011 | 0.016 | -0.014 | 0.016 |
| rs3774604 | C | T | -0.020 | 0.010 | -0.015 | 0.012 | -0.018 | 0.015 | -0.013 | 0.016 | -0.026 | 0.016 |
| rs2629881 | C | T | 0.007 | 0.012 | 0.010 | 0.015 | 0.009 | 0.019 | 0.011 | 0.019 | 0.013 | 0.019 |
| rs538579 | G | C | -0.014 | 0.010 | -0.020 | 0.012 | -0.009 | 0.016 | -0.029 | 0.016 | -0.017 | 0.016 |
| rs115903965 | G | A | 0.031 | 0.044 | 0.008 | 0.052 | -0.021 | 0.064 | 0.046 | 0.069 | 0.022 | 0.069 |
| rs4677156 | A | T | 0.033 | 0.012 | 0.037 | 0.014 | 0.030 | 0.018 | 0.052 | 0.018 | 0.042 | 0.018 |
| rs1666132 | C | T | -0.010 | 0.010 | -0.019 | 0.012 | -0.009 | 0.015 | -0.031 | 0.015 | -0.001 | 0.015 |
| rs1357798 | C | T | 0.000 | 0.012 | -0.004 | 0.014 | -0.002 | 0.018 | -0.005 | 0.018 | 0.018 | 0.018 |
| rs6783281 | A | G | 0.005 | 0.011 | 0.015 | 0.013 | 0.006 | 0.016 | 0.021 | 0.017 | 0.019 | 0.017 |
| rs7355953 | T | C | 0.020 | 0.012 | 0.029 | 0.014 | 0.038 | 0.018 | 0.024 | 0.018 | 0.024 | 0.018 |
| rs2735556 | T | C | 0.005 | 0.016 | -0.004 | 0.019 | -0.016 | 0.024 | 0.005 | 0.025 | 0.035 | 0.026 |
| rs11925138 | G | A | -0.001 | 0.016 | 0.002 | 0.019 | -0.008 | 0.024 | 0.011 | 0.025 | -0.031 | 0.025 |
| rs7625768 | G | A | -0.002 | 0.010 | 0.009 | 0.013 | -0.006 | 0.016 | 0.021 | 0.016 | -0.034 | 0.016 |
| rs1199333 | G | T | 0.018 | 0.012 | 0.035 | 0.015 | 0.038 | 0.018 | 0.027 | 0.019 | -0.030 | 0.019 |
| rs59714050 | T | A | 0.006 | 0.019 | 0.000 | 0.024 | -0.033 | 0.030 | 0.051 | 0.031 | 0.002 | 0.031 |
| rs355748 | G | T | 0.000 | 0.010 | 0.004 | 0.012 | 0.004 | 0.015 | 0.013 | 0.015 | -0.009 | 0.015 |
| rs7633995 | A | G | -0.001 | 0.017 | -0.026 | 0.020 | -0.027 | 0.026 | -0.023 | 0.026 | 0.037 | 0.027 |
| rs10937241 | A | G | 0.014 | 0.013 | 0.020 | 0.016 | 0.031 | 0.020 | 0.012 | 0.021 | -0.011 | 0.021 |
| rs34811474 | G | A | 0.022 | 0.013 | 0.038 | 0.016 | 0.033 | 0.020 | 0.043 | 0.021 | 0.016 | 0.021 |
| rs7656673 | A | G | -0.001 | 0.010 | -0.006 | 0.012 | 0.003 | 0.015 | -0.014 | 0.015 | -0.003 | 0.015 |
| rs34722008 | G | A | 0.004 | 0.011 | 0.009 | 0.013 | 0.018 | 0.016 | 0.002 | 0.016 | 0.015 | 0.016 |
| rs7439324 | C | T | 0.008 | 0.013 | 0.034 | 0.016 | 0.033 | 0.020 | 0.042 | 0.020 | -0.011 | 0.020 |
| rs12641981 | C | T | 0.000 | 0.010 | -0.005 | 0.012 | -0.008 | 0.015 | 0.002 | 0.015 | -0.001 | 0.015 |
| rs788858 | A | G | -0.008 | 0.011 | -0.012 | 0.013 | -0.020 | 0.016 | -0.004 | 0.017 | 0.001 | 0.017 |
| rs7377083 | C | A | 0.000 | 0.010 | -0.004 | 0.012 | -0.004 | 0.015 | -0.004 | 0.016 | 0.006 | 0.016 |
| rs72675820 | A | C | 0.000 | 0.010 | -0.002 | 0.012 | 0.002 | 0.015 | -0.003 | 0.016 | 0.006 | 0.016 |
| rs35189091 | A | G | -0.009 | 0.010 | -0.005 | 0.012 | -0.019 | 0.015 | 0.011 | 0.015 | -0.018 | 0.016 |
| rs11727676 | T | C | -0.078 | 0.017 | -0.091 | 0.020 | -0.090 | 0.025 | -0.094 | 0.026 | -0.077 | 0.026 |
| rs3936511 | A | G | -0.009 | 0.012 | -0.019 | 0.015 | -0.026 | 0.019 | -0.010 | 0.019 | 0.013 | 0.019 |
| rs6449532 | C | T | 0.006 | 0.010 | 0.008 | 0.012 | -0.002 | 0.015 | 0.028 | 0.016 | 0.007 | 0.016 |
| rs9291816 | C | T | 0.006 | 0.010 | 0.007 | 0.012 | 0.003 | 0.016 | 0.010 | 0.016 | 0.010 | 0.016 |
| rs39862 | T | C | 0.010 | 0.011 | 0.003 | 0.013 | 0.000 | 0.016 | -0.001 | 0.017 | 0.009 | 0.017 |
| rs2307111 | T | C | 0.016 | 0.010 | 0.008 | 0.012 | -0.003 | 0.015 | 0.018 | 0.015 | 0.009 | 0.015 |

| | | | | | | | | | | | |
|---|---|---|---|---|---|---|---|---|---|---|---|
| rs1422067 | C | T | -0.012 | 0.012 | -0.001 | 0.014 | 0.014 | 0.018 | -0.028 | 0.018 | -0.030 | 0.018 |
| rs2115885 | G | A | -0.031 | 0.012 | -0.036 | 0.014 | -0.034 | 0.017 | -0.040 | 0.018 | -0.011 | 0.018 |
| rs77960 | G | A | 0.017 | 0.010 | 0.019 | 0.012 | 0.026 | 0.016 | 0.016 | 0.016 | 0.021 | 0.016 |
| rs4958568 | G | A | -0.002 | 0.011 | -0.002 | 0.013 | -0.003 | 0.016 | -0.009 | 0.017 | -0.013 | 0.017 |
| rs7719067 | A | G | 0.012 | 0.010 | 0.018 | 0.012 | 0.014 | 0.015 | 0.009 | 0.015 | -0.011 | 0.015 |
| rs7711823 | A | G | 0.017 | 0.010 | 0.016 | 0.012 | 0.012 | 0.015 | 0.018 | 0.016 | 0.036 | 0.016 |
| rs918472 | G | A | 0.001 | 0.011 | 0.000 | 0.013 | 0.000 | 0.016 | -0.002 | 0.016 | -0.017 | 0.017 |
| rs12214497 | G | T | 0.002 | 0.010 | 0.006 | 0.012 | 0.031 | 0.015 | -0.019 | 0.016 | -0.027 | 0.016 |
| rs10498713 | G | T | -0.006 | 0.014 | -0.004 | 0.016 | 0.026 | 0.021 | -0.033 | 0.021 | -0.012 | 0.021 |
| rs35162296 | C | T | -0.064 | 0.019 | -0.073 | 0.022 | -0.098 | 0.027 | -0.062 | 0.028 | -0.089 | 0.028 |
| rs34196306 | G | C | -0.064 | 0.019 | -0.073 | 0.022 | -0.107 | 0.028 | -0.043 | 0.029 | -0.087 | 0.029 |
| rs3131336 | C | T | -0.059 | 0.018 | -0.069 | 0.021 | -0.107 | 0.026 | -0.037 | 0.027 | -0.075 | 0.027 |
| rs3129942 | G | T | -0.022 | 0.011 | -0.020 | 0.014 | -0.024 | 0.017 | -0.012 | 0.018 | -0.031 | 0.018 |
| rs9366803 | T | C | -0.023 | 0.016 | -0.018 | 0.020 | -0.010 | 0.025 | -0.028 | 0.025 | -0.051 | 0.024 |
| rs686431 | C | T | -0.113 | 0.037 | -0.130 | 0.043 | -0.156 | 0.053 | -0.139 | 0.055 | -0.147 | 0.057 |
| rs73422097 | A | G | 0.012 | 0.010 | 0.007 | 0.013 | 0.010 | 0.016 | 0.007 | 0.016 | 0.026 | 0.016 |
| rs76187039 | G | T | 0.012 | 0.015 | 0.003 | 0.017 | 0.005 | 0.022 | -0.006 | 0.023 | 0.016 | 0.023 |
| rs3798519 | A | C | 0.009 | 0.012 | 0.006 | 0.015 | 0.020 | 0.019 | -0.005 | 0.019 | 0.025 | 0.020 |
| rs1775255 | G | T | -0.011 | 0.010 | -0.009 | 0.012 | -0.010 | 0.015 | -0.004 | 0.015 | -0.010 | 0.015 |
| rs1342831 | T | C | 0.009 | 0.018 | 0.024 | 0.023 | 0.028 | 0.029 | 0.019 | 0.030 | 0.005 | 0.030 |
| rs12110721 | G | A | -0.030 | 0.013 | -0.041 | 0.016 | -0.061 | 0.020 | -0.019 | 0.021 | -0.026 | 0.021 |
| rs9370527 | G | A | 0.007 | 0.011 | 0.006 | 0.014 | 0.011 | 0.017 | 0.008 | 0.018 | 0.020 | 0.018 |
| rs435775 | A | G | 0.000 | 0.011 | -0.010 | 0.013 | -0.018 | 0.017 | -0.009 | 0.017 | -0.020 | 0.017 |
| rs6931604 | C | T | 0.030 | 0.010 | 0.039 | 0.012 | 0.014 | 0.015 | 0.059 | 0.015 | 0.032 | 0.015 |
| rs34260097 | T | G | -0.014 | 0.012 | -0.011 | 0.014 | -0.017 | 0.018 | -0.014 | 0.018 | -0.007 | 0.018 |
| rs7759938 | C | T | 0.001 | 0.010 | 0.000 | 0.012 | 0.000 | 0.016 | 0.000 | 0.016 | -0.003 | 0.016 |
| rs7753558 | C | A | -0.001 | 0.010 | -0.008 | 0.012 | 0.003 | 0.015 | -0.014 | 0.016 | 0.017 | 0.016 |
| rs1452991 | G | A | 0.012 | 0.010 | 0.009 | 0.012 | 0.017 | 0.015 | 0.008 | 0.016 | 0.020 | 0.016 |
| rs796915 | C | G | 0.016 | 0.010 | 0.019 | 0.012 | 0.011 | 0.016 | 0.026 | 0.016 | 0.024 | 0.016 |
| rs62425398 | C | A | -0.007 | 0.017 | 0.008 | 0.020 | 0.005 | 0.025 | 0.010 | 0.026 | -0.029 | 0.026 |
| rs2349179 | T | A | -0.026 | 0.010 | -0.034 | 0.012 | -0.018 | 0.015 | -0.045 | 0.016 | -0.019 | 0.016 |
| rs2722406 | C | T | -0.002 | 0.011 | -0.009 | 0.013 | -0.010 | 0.016 | -0.014 | 0.016 | 0.010 | 0.017 |
| rs4723263 | G | C | -0.001 | 0.010 | -0.007 | 0.012 | -0.014 | 0.015 | -0.008 | 0.015 | 0.007 | 0.015 |
| rs10234366 | G | A | -0.016 | 0.016 | -0.021 | 0.018 | 0.002 | 0.023 | -0.045 | 0.024 | -0.032 | 0.024 |
| rs1852006 | G | A | -0.011 | 0.010 | -0.007 | 0.012 | -0.014 | 0.015 | 0.001 | 0.016 | -0.009 | 0.016 |
| rs6974282 | C | T | -0.035 | 0.012 | -0.024 | 0.015 | -0.031 | 0.018 | -0.012 | 0.019 | -0.052 | 0.019 |
| rs7808296 | C | T | -0.002 | 0.010 | 0.011 | 0.013 | 0.010 | 0.016 | 0.006 | 0.016 | -0.037 | 0.016 |
| rs262338 | G | T | -0.003 | 0.010 | -0.002 | 0.012 | -0.001 | 0.015 | -0.004 | 0.015 | -0.030 | 0.015 |
| rs10953577 | T | C | 0.008 | 0.010 | 0.011 | 0.012 | 0.014 | 0.015 | 0.020 | 0.015 | -0.011 | 0.015 |
| rs67679818 | C | T | -0.017 | 0.010 | -0.024 | 0.012 | -0.039 | 0.015 | -0.009 | 0.015 | -0.004 | 0.015 |
| rs6979832 | A | G | 0.001 | 0.010 | -0.006 | 0.012 | -0.003 | 0.015 | -0.013 | 0.015 | 0.020 | 0.015 |
| rs11525873 | T | C | 0.017 | 0.016 | 0.015 | 0.020 | 0.008 | 0.026 | 0.028 | 0.027 | 0.043 | 0.027 |
| rs10503246 | A | G | -0.003 | 0.011 | 0.001 | 0.013 | 0.006 | 0.016 | -0.003 | 0.017 | -0.001 | 0.017 |
| rs77976727 | C | T | -0.005 | 0.016 | -0.017 | 0.019 | -0.013 | 0.024 | -0.031 | 0.025 | 0.012 | 0.025 |
| rs7814267 | A | G | -0.018 | 0.012 | -0.021 | 0.015 | -0.030 | 0.019 | -0.015 | 0.019 | -0.019 | 0.019 |

| | | | | | | | | | | | |
|---|---|---|---|---|---|---|---|---|---|---|---|
| rs11777719 | A | G | -0.012 | 0.011 | 0.002 | 0.013 | 0.023 | 0.017 | -0.024 | 0.017 | -0.041 | 0.018 |
| rs13256357 | C | T | -0.006 | 0.012 | -0.010 | 0.015 | 0.001 | 0.019 | -0.017 | 0.019 | -0.001 | 0.020 |
| rs2409743 | C | G | -0.001 | 0.011 | -0.017 | 0.013 | -0.032 | 0.016 | 0.004 | 0.017 | 0.017 | 0.017 |
| rs10503555 | A | G | -0.004 | 0.010 | 0.002 | 0.012 | -0.003 | 0.015 | 0.008 | 0.015 | -0.025 | 0.015 |
| rs884152 | G | T | 0.008 | 0.010 | 0.013 | 0.012 | 0.006 | 0.015 | 0.029 | 0.016 | 0.011 | 0.016 |
| rs7012648 | G | A | -0.001 | 0.010 | 0.001 | 0.012 | -0.015 | 0.015 | 0.018 | 0.015 | 0.002 | 0.015 |
| rs4739558 | A | G | 0.007 | 0.010 | 0.011 | 0.012 | 0.004 | 0.015 | 0.014 | 0.015 | 0.008 | 0.016 |
| rs10095724 | G | A | 0.015 | 0.010 | 0.020 | 0.012 | 0.020 | 0.015 | 0.018 | 0.016 | 0.023 | 0.016 |
| rs10111937 | C | T | 0.004 | 0.010 | 0.002 | 0.012 | 0.000 | 0.016 | 0.008 | 0.016 | 0.009 | 0.016 |
| rs7840305 | A | G | 0.002 | 0.010 | 0.007 | 0.012 | 0.004 | 0.015 | 0.016 | 0.016 | -0.007 | 0.016 |
| rs13254613 | A | C | 0.001 | 0.010 | 0.013 | 0.012 | 0.030 | 0.015 | -0.006 | 0.016 | 0.001 | 0.016 |
| rs7817581 | G | A | 0.016 | 0.011 | 0.024 | 0.014 | 0.018 | 0.017 | 0.030 | 0.018 | 0.027 | 0.018 |
| rs10962279 | T | C | 0.004 | 0.012 | 0.008 | 0.015 | 0.005 | 0.019 | -0.005 | 0.019 | -0.006 | 0.019 |
| rs3118252 | G | C | 0.001 | 0.010 | -0.001 | 0.012 | -0.019 | 0.015 | 0.018 | 0.015 | -0.017 | 0.016 |
| rs1935354 | T | C | -0.010 | 0.010 | -0.020 | 0.012 | -0.022 | 0.015 | -0.018 | 0.015 | -0.011 | 0.015 |
| rs1619120 | A | G | -0.001 | 0.010 | 0.007 | 0.012 | 0.008 | 0.015 | 0.002 | 0.015 | -0.029 | 0.015 |
| rs4744246 | A | G | 0.015 | 0.010 | 0.016 | 0.012 | 0.053 | 0.015 | -0.023 | 0.016 | -0.014 | 0.016 |
| rs7020564 | A | T | 0.010 | 0.011 | 0.016 | 0.013 | 0.016 | 0.016 | 0.015 | 0.016 | 0.007 | 0.017 |
| rs957512 | T | C | 0.018 | 0.010 | 0.028 | 0.012 | 0.023 | 0.015 | 0.034 | 0.016 | 0.026 | 0.016 |
| rs10116891 | G | A | 0.043 | 0.017 | 0.032 | 0.019 | 0.054 | 0.025 | 0.006 | 0.025 | 0.056 | 0.026 |
| rs2275241 | G | A | -0.012 | 0.010 | -0.025 | 0.012 | -0.026 | 0.015 | -0.021 | 0.016 | 0.007 | 0.016 |
| rs117911387 | G | A | 0.008 | 0.026 | 0.021 | 0.031 | -0.010 | 0.038 | 0.062 | 0.040 | -0.025 | 0.040 |
| rs7084503 | T | C | 0.020 | 0.010 | 0.018 | 0.012 | 0.010 | 0.015 | 0.024 | 0.015 | 0.037 | 0.015 |
| rs11256627 | G | A | 0.024 | 0.011 | 0.020 | 0.013 | 0.028 | 0.016 | 0.015 | 0.017 | 0.024 | 0.017 |
| rs4572029 | A | G | -0.014 | 0.011 | -0.019 | 0.014 | -0.021 | 0.018 | -0.026 | 0.018 | -0.013 | 0.018 |
| rs10823504 | G | A | -0.012 | 0.019 | -0.016 | 0.023 | -0.034 | 0.029 | 0.010 | 0.030 | 0.019 | 0.030 |
| rs2242258 | T | C | -0.001 | 0.011 | -0.012 | 0.014 | -0.015 | 0.017 | -0.002 | 0.018 | 0.008 | 0.018 |
| rs17399739 | A | G | 0.022 | 0.019 | 0.034 | 0.023 | 0.048 | 0.029 | 0.033 | 0.030 | 0.031 | 0.030 |
| rs10887571 | C | T | -0.002 | 0.010 | -0.007 | 0.012 | -0.019 | 0.016 | 0.007 | 0.016 | 0.005 | 0.016 |
| rs41310284 | C | A | -0.011 | 0.017 | -0.006 | 0.020 | -0.013 | 0.025 | 0.006 | 0.026 | -0.010 | 0.026 |
| rs75387636 | G | A | -0.007 | 0.023 | -0.014 | 0.029 | 0.051 | 0.037 | -0.068 | 0.037 | -0.044 | 0.037 |
| rs2939931 | T | C | 0.015 | 0.010 | 0.008 | 0.012 | -0.003 | 0.015 | 0.018 | 0.015 | 0.008 | 0.015 |
| rs1061072 | G | A | -0.006 | 0.016 | -0.003 | 0.019 | -0.003 | 0.024 | -0.003 | 0.024 | -0.023 | 0.024 |
| rs56133711 | G | A | 0.024 | 0.012 | 0.021 | 0.014 | 0.012 | 0.017 | 0.026 | 0.018 | 0.033 | 0.018 |
| rs4267058 | T | C | 0.025 | 0.010 | 0.014 | 0.012 | -0.003 | 0.015 | 0.033 | 0.016 | 0.047 | 0.016 |
| rs661878 | A | G | 0.003 | 0.014 | -0.002 | 0.016 | -0.001 | 0.021 | -0.004 | 0.021 | 0.012 | 0.022 |
| rs3181269 | C | T | -0.014 | 0.012 | -0.004 | 0.014 | -0.027 | 0.017 | 0.020 | 0.018 | -0.023 | 0.018 |
| rs7951870 | T | C | 0.017 | 0.013 | 0.017 | 0.016 | 0.012 | 0.020 | 0.021 | 0.020 | 0.031 | 0.021 |
| rs12798028 | C | T | -0.008 | 0.010 | -0.004 | 0.012 | -0.013 | 0.015 | 0.000 | 0.015 | -0.006 | 0.015 |
| rs3862342 | C | T | 0.012 | 0.011 | 0.013 | 0.013 | 0.013 | 0.016 | 0.006 | 0.017 | 0.005 | 0.017 |
| rs2958542 | C | T | -0.005 | 0.010 | -0.012 | 0.012 | -0.019 | 0.015 | -0.012 | 0.016 | -0.001 | 0.016 |
| rs10791902 | C | T | 0.005 | 0.011 | 0.001 | 0.013 | -0.001 | 0.016 | 0.004 | 0.017 | 0.012 | 0.017 |
| rs10896348 | T | C | -0.012 | 0.011 | -0.011 | 0.013 | -0.015 | 0.016 | -0.003 | 0.017 | -0.025 | 0.017 |
| rs10796828 | T | G | -0.013 | 0.010 | -0.017 | 0.012 | -0.006 | 0.016 | -0.026 | 0.016 | 0.005 | 0.016 |
| rs11215403 | G | A | -0.018 | 0.012 | -0.019 | 0.014 | -0.016 | 0.017 | -0.013 | 0.018 | -0.016 | 0.018 |

| | | | | | | | | | | | |
|---|---|---|---|---|---|---|---|---|---|---|---|
| rs7123283 | C | T | -0.008 | 0.010 | -0.006 | 0.012 | -0.026 | 0.015 | 0.013 | 0.015 | -0.027 | 0.015 |
| rs10790809 | A | G | -0.005 | 0.010 | 0.008 | 0.012 | 0.019 | 0.016 | 0.002 | 0.016 | -0.022 | 0.016 |
| rs55726687 | G | A | -0.015 | 0.012 | -0.026 | 0.014 | -0.033 | 0.018 | -0.016 | 0.019 | 0.007 | 0.019 |
| rs2187642 | A | C | 0.000 | 0.010 | 0.005 | 0.012 | 0.008 | 0.015 | 0.000 | 0.016 | -0.001 | 0.016 |
| rs10841379 | A | G | 0.018 | 0.010 | 0.022 | 0.012 | 0.016 | 0.016 | 0.032 | 0.016 | 0.010 | 0.016 |
| rs10842356 | A | T | -0.018 | 0.010 | -0.017 | 0.012 | -0.019 | 0.015 | -0.020 | 0.015 | -0.039 | 0.015 |
| rs61937656 | G | A | 0.001 | 0.012 | 0.002 | 0.014 | 0.005 | 0.017 | -0.002 | 0.018 | -0.002 | 0.018 |
| rs7958241 | A | G | 0.034 | 0.010 | 0.036 | 0.012 | 0.035 | 0.016 | 0.042 | 0.016 | 0.040 | 0.016 |
| rs7132908 | G | A | -0.012 | 0.010 | -0.006 | 0.012 | 0.004 | 0.015 | -0.014 | 0.015 | -0.006 | 0.016 |
| rs836179 | A | G | 0.041 | 0.010 | 0.038 | 0.012 | 0.030 | 0.015 | 0.050 | 0.015 | 0.053 | 0.016 |
| rs78607331 | C | T | 0.003 | 0.028 | 0.029 | 0.034 | 0.029 | 0.043 | 0.026 | 0.044 | -0.016 | 0.043 |
| rs7306710 | T | C | -0.010 | 0.010 | -0.007 | 0.012 | -0.026 | 0.015 | 0.015 | 0.015 | -0.014 | 0.015 |
| rs10860295 | T | C | 0.006 | 0.010 | 0.016 | 0.012 | 0.011 | 0.015 | 0.012 | 0.015 | -0.001 | 0.015 |
| rs1552759 | T | C | -0.005 | 0.010 | -0.006 | 0.012 | -0.011 | 0.015 | -0.005 | 0.016 | -0.021 | 0.016 |
| rs12817542 | C | T | -0.017 | 0.023 | -0.016 | 0.027 | 0.000 | 0.034 | -0.029 | 0.036 | -0.016 | 0.036 |
| rs61936936 | A | T | -0.027 | 0.017 | -0.017 | 0.020 | -0.033 | 0.025 | 0.005 | 0.026 | -0.071 | 0.026 |
| rs7305424 | A | T | 0.002 | 0.010 | 0.015 | 0.012 | 0.012 | 0.015 | 0.019 | 0.016 | -0.004 | 0.016 |
| rs12308065 | A | G | -0.011 | 0.011 | -0.013 | 0.013 | -0.023 | 0.016 | 0.004 | 0.016 | -0.025 | 0.016 |
| rs28629903 | T | C | -0.006 | 0.010 | -0.008 | 0.012 | -0.008 | 0.015 | -0.005 | 0.015 | 0.023 | 0.015 |
| rs7989098 | T | C | 0.007 | 0.011 | 0.003 | 0.014 | 0.002 | 0.017 | -0.001 | 0.018 | 0.016 | 0.018 |
| rs9652090 | G | T | 0.023 | 0.010 | 0.023 | 0.012 | 0.031 | 0.015 | 0.024 | 0.015 | 0.023 | 0.015 |
| rs1933437 | G | A | -0.011 | 0.010 | -0.013 | 0.012 | 0.000 | 0.015 | -0.026 | 0.015 | 0.003 | 0.015 |
| rs9603697 | C | T | 0.011 | 0.010 | 0.018 | 0.012 | 0.010 | 0.016 | 0.022 | 0.016 | 0.015 | 0.016 |
| rs9594686 | C | T | -0.001 | 0.013 | 0.003 | 0.016 | 0.018 | 0.020 | -0.013 | 0.020 | -0.003 | 0.020 |
| rs12429545 | G | A | 0.005 | 0.014 | 0.015 | 0.017 | 0.016 | 0.022 | 0.006 | 0.022 | 0.020 | 0.023 |
| rs9538141 | G | A | -0.009 | 0.010 | -0.007 | 0.012 | -0.002 | 0.015 | -0.018 | 0.015 | -0.012 | 0.015 |
| rs1333010 | G | A | 0.017 | 0.010 | 0.010 | 0.012 | 0.015 | 0.015 | 0.003 | 0.016 | 0.034 | 0.016 |
| rs1576655 | A | C | -0.017 | 0.010 | -0.021 | 0.012 | -0.020 | 0.015 | -0.024 | 0.016 | -0.009 | 0.016 |
| rs61978655 | G | A | -0.029 | 0.030 | -0.014 | 0.030 | 0.001 | 0.038 | -0.012 | 0.039 | -0.052 | 0.038 |
| rs7161424 | G | A | -0.008 | 0.012 | -0.007 | 0.012 | -0.007 | 0.015 | -0.005 | 0.015 | -0.009 | 0.015 |
| rs1865719 | A | G | 0.011 | 0.012 | 0.014 | 0.012 | 0.015 | 0.015 | 0.011 | 0.016 | 0.040 | 0.016 |
| rs10133279 | C | T | -0.003 | 0.012 | 0.010 | 0.012 | 0.001 | 0.015 | 0.023 | 0.016 | -0.018 | 0.016 |
| rs7145052 | C | T | 0.004 | 0.011 | 0.007 | 0.012 | -0.006 | 0.015 | 0.015 | 0.015 | 0.004 | 0.015 |
| rs7159126 | T | C | 0.013 | 0.012 | -0.008 | 0.013 | -0.002 | 0.016 | -0.018 | 0.017 | 0.004 | 0.017 |
| rs78420139 | G | A | 0.047 | 0.026 | 0.030 | 0.027 | 0.055 | 0.034 | 0.014 | 0.035 | 0.069 | 0.035 |
| rs12436513 | C | A | -0.009 | 0.012 | 0.001 | 0.012 | 0.002 | 0.015 | 0.002 | 0.016 | -0.011 | 0.016 |
| rs824207 | A | G | 0.008 | 0.010 | 0.006 | 0.012 | -0.004 | 0.015 | 0.012 | 0.016 | 0.008 | 0.016 |
| rs62048187 | G | C | -0.031 | 0.011 | -0.035 | 0.013 | -0.036 | 0.016 | -0.037 | 0.017 | -0.029 | 0.017 |
| rs10519136 | C | T | -0.011 | 0.010 | -0.020 | 0.012 | -0.018 | 0.015 | -0.025 | 0.015 | -0.004 | 0.015 |
| rs8030456 | C | T | 0.044 | 0.011 | 0.044 | 0.014 | 0.058 | 0.017 | 0.028 | 0.018 | 0.011 | 0.018 |
| rs7162542 | C | G | -0.013 | 0.010 | -0.017 | 0.012 | -0.017 | 0.015 | -0.014 | 0.015 | -0.012 | 0.015 |
| rs3817428 | C | G | -0.007 | 0.011 | 0.001 | 0.014 | 0.011 | 0.017 | -0.011 | 0.018 | -0.019 | 0.018 |
| rs1000471 | C | T | 0.008 | 0.012 | 0.004 | 0.015 | 0.005 | 0.018 | 0.003 | 0.019 | -0.003 | 0.019 |
| rs2970356 | C | G | -0.005 | 0.011 | -0.016 | 0.013 | -0.001 | 0.016 | -0.022 | 0.017 | 0.009 | 0.017 |
| rs72755233 | G | A | 0.024 | 0.018 | 0.030 | 0.022 | 0.043 | 0.028 | 0.025 | 0.029 | 0.032 | 0.029 |

| | | | | | | | | | | | |
|---|---|---|---|---|---|---|---|---|---|---|---|
| rs2238435 | C | G | -0.009 | 0.010 | -0.004 | 0.012 | -0.004 | 0.015 | -0.012 | 0.015 | -0.011 | 0.016 |
| rs55880046 | T | G | -0.011 | 0.014 | -0.018 | 0.017 | 0.002 | 0.021 | -0.040 | 0.022 | 0.013 | 0.022 |
| rs4432271 | C | T | -0.008 | 0.014 | -0.004 | 0.017 | -0.006 | 0.022 | 0.014 | 0.022 | -0.001 | 0.022 |
| rs9922288 | A | G | -0.011 | 0.012 | -0.020 | 0.015 | -0.016 | 0.018 | -0.018 | 0.019 | 0.018 | 0.019 |
| rs62037365 | C | G | -0.011 | 0.010 | -0.017 | 0.012 | -0.022 | 0.015 | -0.016 | 0.016 | -0.007 | 0.016 |
| rs4889630 | T | C | 0.025 | 0.012 | 0.019 | 0.014 | 0.021 | 0.018 | 0.009 | 0.019 | 0.024 | 0.019 |
| rs4783789 | T | C | -0.012 | 0.011 | -0.008 | 0.013 | -0.017 | 0.017 | 0.004 | 0.017 | -0.017 | 0.018 |
| rs1421085 | T | C | 0.004 | 0.010 | -0.003 | 0.012 | 0.000 | 0.015 | -0.008 | 0.015 | 0.008 | 0.015 |
| rs594585 | T | G | 0.020 | 0.010 | 0.029 | 0.012 | 0.028 | 0.015 | 0.031 | 0.015 | 0.011 | 0.015 |
| rs117903946 | G | A | -0.001 | 0.029 | 0.009 | 0.034 | -0.014 | 0.043 | 0.030 | 0.044 | -0.059 | 0.043 |
| rs7672 | C | G | -0.019 | 0.011 | -0.013 | 0.013 | -0.004 | 0.016 | -0.010 | 0.017 | -0.011 | 0.017 |
| rs4985555 | A | G | 0.004 | 0.010 | 0.008 | 0.012 | 0.014 | 0.015 | -0.003 | 0.016 | -0.001 | 0.016 |
| rs11642090 | T | C | 0.008 | 0.010 | 0.004 | 0.012 | 0.010 | 0.015 | -0.006 | 0.016 | -0.002 | 0.016 |
| rs72819571 | G | T | 0.005 | 0.010 | -0.001 | 0.012 | 0.011 | 0.015 | -0.004 | 0.016 | 0.006 | 0.016 |
| rs67603370 | G | A | 0.016 | 0.019 | 0.023 | 0.023 | 0.025 | 0.029 | 0.030 | 0.030 | 0.042 | 0.030 |
| rs3815156 | A | G | -0.021 | 0.013 | -0.032 | 0.015 | -0.032 | 0.019 | -0.039 | 0.020 | -0.018 | 0.020 |
| rs12601380 | A | C | 0.002 | 0.010 | 0.003 | 0.012 | 0.017 | 0.015 | -0.011 | 0.015 | 0.016 | 0.015 |
| rs9299 | C | T | -0.003 | 0.010 | -0.006 | 0.012 | -0.005 | 0.015 | -0.004 | 0.016 | 0.009 | 0.016 |
| rs17637472 | G | A | 0.028 | 0.010 | 0.024 | 0.012 | 0.013 | 0.015 | 0.044 | 0.015 | 0.028 | 0.015 |
| rs7217460 | G | A | 0.017 | 0.011 | 0.015 | 0.014 | -0.001 | 0.017 | 0.027 | 0.018 | 0.034 | 0.018 |
| rs12941038 | C | T | 0.012 | 0.011 | 0.007 | 0.014 | 0.013 | 0.017 | 0.004 | 0.018 | 0.035 | 0.018 |
| rs2246623 | C | T | -0.026 | 0.010 | -0.022 | 0.012 | -0.009 | 0.015 | -0.036 | 0.016 | -0.044 | 0.016 |
| rs11150745 | A | G | -0.007 | 0.010 | -0.008 | 0.012 | 0.000 | 0.016 | -0.008 | 0.016 | 0.007 | 0.016 |
| rs7503580 | C | T | -0.005 | 0.013 | -0.019 | 0.016 | -0.014 | 0.020 | -0.030 | 0.021 | 0.025 | 0.021 |
| rs1013737 | G | C | 0.001 | 0.010 | -0.007 | 0.012 | 0.003 | 0.015 | -0.027 | 0.015 | -0.021 | 0.015 |
| rs1808579 | C | T | 0.020 | 0.010 | 0.012 | 0.012 | 0.006 | 0.015 | 0.020 | 0.015 | 0.030 | 0.015 |
| rs7237444 | G | A | -0.017 | 0.011 | -0.021 | 0.013 | -0.035 | 0.017 | -0.006 | 0.017 | -0.023 | 0.017 |
| rs7239114 | G | A | -0.001 | 0.010 | -0.008 | 0.012 | -0.009 | 0.015 | -0.004 | 0.015 | 0.012 | 0.015 |
| rs68015088 | G | A | 0.007 | 0.010 | 0.016 | 0.012 | 0.014 | 0.015 | 0.026 | 0.016 | -0.012 | 0.016 |
| rs12606230 | T | C | 0.002 | 0.011 | 0.001 | 0.014 | -0.003 | 0.018 | 0.016 | 0.018 | 0.017 | 0.018 |
| rs663129 | G | A | -0.019 | 0.011 | -0.010 | 0.014 | -0.007 | 0.017 | -0.010 | 0.018 | -0.025 | 0.018 |
| rs113728099 | G | A | 0.024 | 0.032 | 0.022 | 0.038 | -0.011 | 0.048 | 0.038 | 0.050 | 0.028 | 0.050 |
| rs8096658 | C | G | -0.008 | 0.011 | -0.021 | 0.013 | -0.002 | 0.016 | -0.032 | 0.017 | -0.012 | 0.017 |
| rs62621197 | C | T | -0.035 | 0.029 | -0.025 | 0.034 | 0.022 | 0.044 | -0.079 | 0.044 | -0.020 | 0.046 |
| rs4545941 | T | C | 0.008 | 0.015 | 0.005 | 0.018 | 0.023 | 0.022 | -0.021 | 0.023 | -0.008 | 0.023 |
| rs116399833 | C | A | -0.001 | 0.012 | -0.005 | 0.014 | 0.012 | 0.018 | -0.014 | 0.019 | 0.011 | 0.019 |
| rs4808961 | C | G | -0.021 | 0.010 | -0.016 | 0.012 | -0.014 | 0.015 | -0.019 | 0.016 | -0.022 | 0.016 |
| rs3810304 | A | G | -0.004 | 0.012 | 0.003 | 0.014 | 0.013 | 0.018 | -0.011 | 0.018 | -0.004 | 0.018 |
| rs1800437 | G | C | -0.005 | 0.012 | -0.002 | 0.014 | -0.002 | 0.018 | 0.001 | 0.018 | -0.007 | 0.019 |
| rs3810291 | G | A | 0.000 | 0.010 | 0.003 | 0.013 | -0.008 | 0.016 | 0.017 | 0.016 | -0.005 | 0.016 |
| rs601338 | G | A | -0.018 | 0.010 | -0.028 | 0.012 | -0.044 | 0.015 | -0.010 | 0.015 | -0.008 | 0.015 |
| rs16996644 | C | G | 0.009 | 0.015 | 0.015 | 0.018 | 0.026 | 0.023 | 0.013 | 0.023 | 0.044 | 0.023 |
| rs947088 | G | T | -0.002 | 0.011 | 0.001 | 0.013 | -0.007 | 0.016 | 0.012 | 0.017 | -0.010 | 0.017 |
| rs8117463 | G | A | 0.005 | 0.011 | -0.002 | 0.012 | -0.016 | 0.016 | 0.011 | 0.016 | 0.025 | 0.016 |
| rs73085586 | G | A | 0.021 | 0.012 | 0.026 | 0.014 | 0.021 | 0.018 | 0.034 | 0.019 | -0.006 | 0.019 |

| | | | | | | | | | | | | |
|---|---|---|---|---|---|---|---|---|---|---|---|---|
| rs2281148 | T | C | -0.006 | 0.011 | -0.014 | 0.014 | -0.024 | 0.017 | -0.004 | 0.018 | 0.014 | 0.018 |
| rs2207894 | C | T | -0.005 | 0.012 | 0.002 | 0.014 | 0.000 | 0.018 | 0.002 | 0.019 | -0.025 | 0.019 |
| rs117455294 | C | A | 0.034 | 0.026 | 0.056 | 0.031 | 0.039 | 0.039 | 0.080 | 0.041 | -0.017 | 0.040 |
| rs8130408 | A | C | -0.005 | 0.012 | 0.007 | 0.014 | -0.006 | 0.018 | 0.027 | 0.018 | -0.026 | 0.019 |
| rs13047416 | C | G | -0.017 | 0.010 | -0.003 | 0.012 | 0.015 | 0.015 | -0.019 | 0.016 | -0.038 | 0.016 |
| rs78907487 | A | C | -0.009 | 0.014 | -0.016 | 0.016 | -0.005 | 0.020 | -0.033 | 0.021 | 0.038 | 0.021 |
| rs9610387 | G | A | 0.011 | 0.018 | 0.015 | 0.021 | 0.040 | 0.027 | -0.007 | 0.028 | -0.006 | 0.028 |
| rs6001872 | A | G | 0.017 | 0.010 | 0.008 | 0.012 | 0.004 | 0.015 | 0.012 | 0.016 | 0.021 | 0.016 |
| rs9611560 | T | C | -0.018 | 0.011 | -0.023 | 0.014 | -0.022 | 0.017 | -0.015 | 0.018 | -0.032 | 0.018 |
| Early life body size SNPs (men only) | | | | | | | | | | | | |
| SNP | EA | OA | beta_ca | se_ca | beta_co | se_co | beta_prox | se_prox | beta_dist | se_dist | beta_re | se_re |
| rs12140153 | G | T | 0.000 | 0.026 | 0.012 | 0.032 | -0.008 | 0.041 | 0.027 | 0.040 | -0.034 | 0.038 |
| rs2012697 | T | C | 0.025 | 0.014 | 0.023 | 0.017 | 0.008 | 0.022 | 0.023 | 0.022 | 0.033 | 0.021 |
| rs4650277 | A | G | 0.010 | 0.014 | 0.023 | 0.017 | 0.028 | 0.022 | 0.022 | 0.021 | 0.016 | 0.021 |
| rs7550711 | C | T | 0.002 | 0.040 | -0.025 | 0.049 | -0.008 | 0.063 | 0.008 | 0.062 | -0.030 | 0.059 |
| rs539515 | A | C | 0.013 | 0.017 | 0.000 | 0.022 | -0.024 | 0.028 | 0.009 | 0.027 | 0.031 | 0.026 |
| rs78444298 | G | A | -0.004 | 0.060 | 0.023 | 0.073 | 0.055 | 0.095 | 0.037 | 0.093 | 0.009 | 0.090 |
| rs1772143 | T | A | 0.036 | 0.014 | 0.041 | 0.017 | 0.028 | 0.022 | 0.049 | 0.021 | 0.032 | 0.021 |
| rs77165542 | C | T | 0.033 | 0.043 | 0.007 | 0.051 | 0.046 | 0.067 | -0.015 | 0.065 | 0.051 | 0.065 |
| rs6749422 | C | G | 0.002 | 0.014 | -0.016 | 0.017 | -0.030 | 0.021 | -0.008 | 0.021 | 0.001 | 0.020 |
| rs2862874 | G | T | 0.033 | 0.014 | 0.028 | 0.017 | 0.001 | 0.022 | 0.052 | 0.021 | 0.019 | 0.021 |
| rs10496885 | G | A | -0.007 | 0.018 | 0.021 | 0.022 | 0.058 | 0.028 | -0.011 | 0.027 | -0.013 | 0.026 |
| rs115319174 | G | C | -0.023 | 0.031 | -0.018 | 0.037 | 0.007 | 0.048 | -0.022 | 0.046 | -0.046 | 0.045 |
| rs9880272 | T | G | 0.007 | 0.016 | 0.016 | 0.019 | 0.013 | 0.025 | 0.021 | 0.024 | 0.011 | 0.024 |
| rs34722008 | G | A | -0.017 | 0.015 | -0.006 | 0.018 | 0.007 | 0.023 | -0.007 | 0.023 | -0.001 | 0.022 |
| rs10938398 | G | A | 0.017 | 0.014 | 0.014 | 0.017 | 0.015 | 0.022 | 0.014 | 0.021 | 0.013 | 0.020 |
| rs13107325 | C | T | -0.009 | 0.026 | -0.003 | 0.032 | -0.012 | 0.040 | -0.009 | 0.039 | 0.000 | 0.039 |
| rs3212519 | A | G | -0.010 | 0.026 | -0.028 | 0.032 | -0.018 | 0.041 | -0.051 | 0.039 | 0.025 | 0.039 |
| rs75577466 | G | C | 0.003 | 0.019 | 0.017 | 0.023 | 0.014 | 0.029 | 0.029 | 0.028 | -0.034 | 0.027 |
| rs25842 | C | T | 0.001 | 0.015 | 0.003 | 0.019 | -0.004 | 0.024 | 0.000 | 0.024 | -0.011 | 0.023 |
| rs55654862 | A | G | -0.015 | 0.017 | -0.007 | 0.020 | 0.034 | 0.026 | -0.052 | 0.025 | -0.021 | 0.024 |
| rs4958361 | C | G | 0.001 | 0.014 | 0.010 | 0.017 | 0.015 | 0.021 | 0.002 | 0.021 | -0.004 | 0.020 |
| rs2240071 | C | G | -0.012 | 0.016 | -0.013 | 0.020 | -0.012 | 0.025 | -0.019 | 0.025 | 0.001 | 0.024 |
| rs62405422 | T | C | 0.005 | 0.017 | 0.006 | 0.021 | 0.018 | 0.028 | -0.006 | 0.027 | 0.005 | 0.026 |
| rs1775255 | G | T | -0.030 | 0.014 | -0.029 | 0.017 | -0.027 | 0.022 | -0.030 | 0.021 | -0.025 | 0.020 |
| rs115597956 | G | A | 0.043 | 0.026 | 0.049 | 0.034 | 0.064 | 0.043 | 0.045 | 0.042 | 0.075 | 0.042 |
| rs12110721 | G | A | -0.049 | 0.019 | -0.050 | 0.023 | -0.068 | 0.030 | -0.034 | 0.029 | -0.062 | 0.028 |
| rs2693560 | A | G | 0.015 | 0.014 | 0.007 | 0.018 | 0.018 | 0.023 | 0.005 | 0.022 | 0.016 | 0.021 |
| rs1452991 | G | A | 0.003 | 0.014 | -0.009 | 0.017 | -0.011 | 0.022 | -0.005 | 0.022 | 0.006 | 0.021 |
| rs16120 | A | G | 0.002 | 0.014 | 0.017 | 0.017 | 0.032 | 0.021 | 0.012 | 0.021 | -0.010 | 0.020 |
| rs7796922 | A | G | -0.006 | 0.024 | -0.011 | 0.029 | -0.011 | 0.038 | -0.002 | 0.037 | 0.011 | 0.036 |
| rs2979139 | A | G | -0.016 | 0.015 | -0.019 | 0.018 | -0.031 | 0.024 | -0.014 | 0.023 | -0.016 | 0.022 |
| rs12674871 | C | T | 0.003 | 0.017 | -0.012 | 0.022 | -0.011 | 0.028 | -0.006 | 0.027 | 0.026 | 0.027 |
| rs10504620 | T | C | 0.008 | 0.015 | -0.009 | 0.018 | -0.010 | 0.024 | -0.011 | 0.023 | 0.036 | 0.022 |
| rs10968101 | G | A | -0.014 | 0.014 | -0.025 | 0.017 | -0.029 | 0.022 | -0.016 | 0.021 | -0.007 | 0.021 |

| | | | | | | | | | | | | |
|---|---|---|---|---|---|---|---|---|---|---|---|---|
| rs11790060 | T | C | 0.015 | 0.014 | 0.008 | 0.018 | 0.030 | 0.023 | -0.009 | 0.022 | -0.006 | 0.022 |
| rs41310284 | C | A | -0.002 | 0.024 | -0.002 | 0.029 | 0.001 | 0.038 | -0.010 | 0.037 | 0.005 | 0.036 |
| rs11030102 | C | G | 0.022 | 0.016 | 0.023 | 0.020 | 0.007 | 0.025 | 0.026 | 0.025 | 0.018 | 0.024 |
| rs3817334 | C | T | -0.016 | 0.014 | -0.007 | 0.017 | -0.010 | 0.022 | -0.013 | 0.021 | -0.013 | 0.021 |
| rs10896348 | T | C | -0.008 | 0.015 | -0.011 | 0.019 | -0.033 | 0.024 | 0.016 | 0.024 | -0.019 | 0.023 |
| rs11218734 | A | G | 0.003 | 0.016 | 0.006 | 0.020 | 0.031 | 0.025 | -0.013 | 0.025 | 0.003 | 0.024 |
| rs7978659 | G | T | 0.021 | 0.015 | 0.024 | 0.018 | 0.048 | 0.023 | 0.003 | 0.022 | 0.030 | 0.022 |
| rs7132908 | G | A | 0.001 | 0.014 | 0.005 | 0.017 | 0.006 | 0.022 | 0.004 | 0.022 | 0.018 | 0.021 |
| rs7306710 | T | C | -0.010 | 0.014 | -0.005 | 0.017 | -0.032 | 0.022 | 0.023 | 0.021 | -0.016 | 0.021 |
| rs7316962 | A | G | 0.006 | 0.014 | 0.002 | 0.017 | 0.005 | 0.022 | -0.005 | 0.021 | 0.022 | 0.020 |
| rs9568868 | G | T | 0.009 | 0.020 | 0.039 | 0.025 | 0.063 | 0.032 | 0.014 | 0.031 | -0.007 | 0.030 |
| rs1576655 | A | C | -0.018 | 0.014 | -0.016 | 0.018 | -0.014 | 0.023 | -0.021 | 0.022 | -0.018 | 0.021 |
| rs61978655 | G | A | -0.022 | 0.036 | -0.037 | 0.051 | -0.054 | 0.066 | -0.037 | 0.066 | -0.015 | 0.066 |
| rs2143975 | C | G | 0.021 | 0.014 | 0.033 | 0.020 | 0.042 | 0.025 | 0.025 | 0.025 | 0.037 | 0.025 |
| rs7159126 | T | C | -0.002 | 0.015 | -0.003 | 0.018 | 0.015 | 0.024 | -0.023 | 0.023 | -0.017 | 0.022 |
| rs3784710 | T | C | 0.063 | 0.016 | 0.061 | 0.020 | 0.074 | 0.025 | 0.061 | 0.025 | 0.022 | 0.024 |
| rs7190603 | T | C | -0.015 | 0.020 | -0.006 | 0.024 | 0.016 | 0.031 | -0.024 | 0.030 | -0.017 | 0.029 |
| rs56094641 | A | G | 0.001 | 0.014 | 0.002 | 0.017 | -0.005 | 0.022 | 0.010 | 0.021 | 0.003 | 0.020 |
| rs17637472 | G | A | 0.006 | 0.014 | 0.008 | 0.017 | 0.002 | 0.022 | 0.022 | 0.021 | 0.006 | 0.021 |
| rs2250081 | G | A | -0.032 | 0.014 | -0.037 | 0.017 | -0.032 | 0.022 | -0.043 | 0.022 | -0.032 | 0.021 |
| rs7239114 | G | A | -0.014 | 0.014 | -0.017 | 0.017 | -0.016 | 0.022 | -0.014 | 0.021 | -0.001 | 0.021 |
| rs3764516 | A | C | 0.002 | 0.016 | -0.002 | 0.020 | -0.004 | 0.026 | -0.009 | 0.025 | -0.019 | 0.025 |
| rs663129 | G | A | -0.019 | 0.016 | -0.020 | 0.020 | -0.021 | 0.025 | -0.012 | 0.025 | -0.018 | 0.024 |
| rs1532127 | G | A | 0.020 | 0.015 | 0.027 | 0.018 | 0.017 | 0.023 | 0.044 | 0.023 | 0.012 | 0.022 |
| rs1321434 | A | G | 0.050 | 0.014 | 0.055 | 0.017 | 0.032 | 0.022 | 0.075 | 0.021 | 0.058 | 0.021 |
| rs73898513 | C | T | 0.052 | 0.022 | 0.058 | 0.027 | 0.060 | 0.034 | 0.074 | 0.034 | 0.063 | 0.032 |
| Early life body size SNPs (women only) | | | | | | | | | | | | |
| SNP | EA | OA | beta_ca | se_ca | beta_co | se_co | beta_prox | se_prox | beta_dist | se_dist | beta_re | se_re |
| rs212540 | C | T | 0.004 | 0.014 | 0.005 | 0.016 | 0.016 | 0.020 | -0.024 | 0.022 | -0.005 | 0.023 |
| rs582220 | A | G | 0.002 | 0.014 | -0.007 | 0.016 | 0.001 | 0.020 | -0.018 | 0.022 | 0.034 | 0.023 |
| rs12140153 | G | T | 0.028 | 0.027 | 0.038 | 0.031 | 0.080 | 0.039 | -0.038 | 0.042 | -0.026 | 0.044 |
| rs2767486 | A | G | 0.005 | 0.017 | 0.011 | 0.020 | 0.013 | 0.024 | 0.021 | 0.027 | 0.008 | 0.028 |
| rs7522014 | A | G | -0.009 | 0.015 | 0.008 | 0.018 | 0.002 | 0.022 | 0.017 | 0.024 | -0.035 | 0.025 |
| rs11209943 | A | G | 0.006 | 0.014 | -0.003 | 0.017 | 0.009 | 0.020 | -0.019 | 0.023 | 0.023 | 0.024 |
| rs12042908 | A | G | -0.010 | 0.014 | -0.001 | 0.016 | -0.003 | 0.020 | 0.011 | 0.022 | -0.006 | 0.023 |
| rs41279738 | T | G | 0.002 | 0.042 | 0.002 | 0.049 | -0.015 | 0.060 | -0.020 | 0.066 | 0.029 | 0.069 |
| rs543874 | A | G | -0.017 | 0.017 | 0.003 | 0.021 | 0.006 | 0.025 | 0.007 | 0.028 | -0.038 | 0.029 |
| rs10798139 | C | T | 0.010 | 0.016 | 0.010 | 0.020 | -0.008 | 0.024 | 0.024 | 0.027 | -0.014 | 0.028 |
| rs815339 | T | A | 0.006 | 0.014 | 0.011 | 0.016 | 0.005 | 0.020 | 0.023 | 0.022 | -0.002 | 0.023 |
| rs4971239 | G | A | -0.001 | 0.022 | -0.009 | 0.027 | -0.017 | 0.033 | 0.019 | 0.037 | 0.021 | 0.038 |
| rs62106258 | T | C | 0.041 | 0.036 | 0.060 | 0.042 | 0.027 | 0.051 | 0.121 | 0.058 | 0.000 | 0.059 |
| rs12992672 | G | A | -0.008 | 0.018 | -0.006 | 0.021 | -0.030 | 0.026 | 0.023 | 0.028 | 0.001 | 0.030 |
| rs6738433 | G | C | -0.020 | 0.014 | -0.017 | 0.016 | -0.019 | 0.020 | -0.023 | 0.022 | -0.031 | 0.023 |
| rs146910503 | G | A | 0.119 | 0.053 | 0.114 | 0.062 | 0.103 | 0.076 | 0.114 | 0.086 | -0.010 | 0.085 |
| rs1446725 | T | G | -0.007 | 0.014 | -0.019 | 0.016 | -0.039 | 0.020 | -0.005 | 0.022 | 0.020 | 0.023 |

| | | | | | | | | | | | |
|---|---|---|---|---|---|---|---|---|---|---|---|
| rs1483153 | C | T | 0.018 | 0.016 | 0.027 | 0.020 | 0.010 | 0.024 | 0.057 | 0.026 | 0.037 | 0.028 |
| rs55959207 | A | C | 0.031 | 0.014 | 0.052 | 0.017 | 0.032 | 0.020 | 0.068 | 0.023 | 0.016 | 0.024 |
| rs17464221 | C | T | 0.009 | 0.015 | 0.001 | 0.017 | 0.017 | 0.021 | -0.011 | 0.024 | 0.018 | 0.025 |
| rs115319174 | G | C | -0.017 | 0.031 | -0.001 | 0.036 | 0.031 | 0.044 | -0.053 | 0.048 | -0.024 | 0.051 |
| rs2594989 | C | T | 0.003 | 0.018 | -0.002 | 0.021 | -0.006 | 0.026 | -0.005 | 0.028 | 0.030 | 0.029 |
| rs754635 | C | G | 0.004 | 0.021 | -0.007 | 0.026 | -0.020 | 0.031 | 0.001 | 0.035 | 0.018 | 0.036 |
| rs2034963 | G | C | 0.018 | 0.015 | 0.022 | 0.017 | 0.009 | 0.021 | 0.039 | 0.023 | 0.031 | 0.025 |
| rs2629881 | C | T | 0.005 | 0.017 | -0.003 | 0.020 | -0.012 | 0.025 | 0.002 | 0.028 | -0.014 | 0.029 |
| rs79569013 | T | G | -0.009 | 0.020 | -0.016 | 0.023 | -0.012 | 0.028 | -0.024 | 0.031 | 0.011 | 0.033 |
| rs818219 | T | C | -0.017 | 0.014 | -0.018 | 0.016 | 0.000 | 0.020 | -0.035 | 0.022 | 0.028 | 0.023 |
| rs2735556 | T | C | -0.031 | 0.023 | -0.025 | 0.027 | 0.001 | 0.033 | -0.060 | 0.036 | -0.019 | 0.039 |
| rs1199328 | G | A | 0.029 | 0.017 | 0.046 | 0.020 | 0.040 | 0.024 | 0.050 | 0.027 | -0.031 | 0.029 |
| rs76152047 | A | G | -0.025 | 0.026 | -0.024 | 0.033 | -0.077 | 0.039 | 0.051 | 0.045 | -0.081 | 0.046 |
| rs7656673 | A | G | 0.000 | 0.014 | -0.001 | 0.017 | 0.006 | 0.020 | -0.010 | 0.022 | -0.017 | 0.023 |
| rs12641981 | C | T | -0.019 | 0.014 | -0.025 | 0.016 | -0.027 | 0.020 | -0.013 | 0.022 | -0.011 | 0.023 |
| rs1349641 | T | G | -0.003 | 0.014 | -0.020 | 0.017 | -0.030 | 0.020 | -0.015 | 0.022 | 0.024 | 0.023 |
| rs7377083 | C | A | 0.014 | 0.014 | 0.005 | 0.017 | -0.002 | 0.021 | 0.016 | 0.023 | 0.027 | 0.024 |
| rs3936511 | A | G | -0.035 | 0.018 | -0.049 | 0.021 | -0.023 | 0.025 | -0.072 | 0.027 | 0.006 | 0.029 |
| rs10050620 | C | T | 0.002 | 0.014 | 0.003 | 0.017 | 0.000 | 0.021 | 0.004 | 0.023 | 0.014 | 0.024 |
| rs13190020 | G | A | -0.012 | 0.015 | -0.028 | 0.017 | -0.016 | 0.021 | -0.047 | 0.023 | -0.033 | 0.024 |
| rs9293494 | T | G | -0.037 | 0.016 | -0.039 | 0.019 | -0.045 | 0.023 | -0.036 | 0.025 | -0.025 | 0.026 |
| rs4235642 | A | G | 0.021 | 0.014 | 0.020 | 0.017 | 0.031 | 0.020 | 0.015 | 0.022 | 0.016 | 0.024 |
| rs6860760 | A | G | -0.003 | 0.014 | -0.007 | 0.016 | -0.020 | 0.020 | 0.012 | 0.022 | -0.017 | 0.023 |
| rs815610 | C | G | 0.015 | 0.014 | 0.021 | 0.016 | 0.011 | 0.020 | 0.015 | 0.022 | -0.028 | 0.023 |
| rs12214497 | G | T | 0.016 | 0.014 | 0.025 | 0.017 | 0.038 | 0.021 | -0.008 | 0.023 | -0.015 | 0.024 |
| rs75782365 | T | G | -0.092 | 0.026 | -0.094 | 0.031 | -0.142 | 0.037 | -0.034 | 0.042 | -0.119 | 0.043 |
| rs34196306 | G | C | -0.083 | 0.027 | -0.084 | 0.031 | -0.140 | 0.037 | -0.014 | 0.042 | -0.103 | 0.043 |
| rs3749971 | G | A | -0.093 | 0.024 | -0.091 | 0.028 | -0.133 | 0.034 | -0.043 | 0.038 | -0.112 | 0.039 |
| rs3131934 | T | C | -0.031 | 0.020 | -0.048 | 0.023 | -0.062 | 0.028 | -0.027 | 0.032 | -0.088 | 0.033 |
| rs9268235 | C | T | -0.043 | 0.023 | -0.041 | 0.027 | -0.055 | 0.033 | -0.021 | 0.037 | -0.107 | 0.038 |
| rs141127771 | G | A | -0.035 | 0.025 | -0.017 | 0.030 | 0.001 | 0.037 | -0.048 | 0.040 | -0.027 | 0.041 |
| rs3798544 | G | A | -0.003 | 0.019 | -0.014 | 0.023 | 0.004 | 0.028 | -0.035 | 0.031 | 0.012 | 0.033 |
| rs2206277 | C | T | 0.019 | 0.017 | 0.012 | 0.021 | 0.026 | 0.026 | -0.002 | 0.028 | 0.060 | 0.030 |
| rs1775255 | G | T | 0.005 | 0.014 | 0.005 | 0.016 | -0.001 | 0.020 | 0.020 | 0.022 | 0.007 | 0.023 |
| rs12110721 | G | A | -0.008 | 0.019 | -0.033 | 0.022 | -0.053 | 0.027 | 0.001 | 0.030 | 0.014 | 0.032 |
| rs34260097 | T | G | -0.031 | 0.016 | -0.035 | 0.019 | -0.035 | 0.024 | -0.045 | 0.026 | -0.017 | 0.027 |
| rs7759938 | C | T | 0.014 | 0.015 | 0.010 | 0.017 | 0.010 | 0.021 | 0.017 | 0.023 | -0.002 | 0.025 |
| rs796915 | C | G | 0.021 | 0.015 | 0.023 | 0.017 | 0.016 | 0.021 | 0.031 | 0.023 | 0.028 | 0.025 |
| rs62425122 | G | A | -0.022 | 0.015 | -0.039 | 0.018 | -0.037 | 0.022 | -0.048 | 0.024 | -0.051 | 0.025 |
| rs983949 | T | G | 0.009 | 0.015 | 0.006 | 0.018 | -0.002 | 0.022 | 0.013 | 0.024 | 0.035 | 0.025 |
| rs7808296 | C | T | -0.013 | 0.015 | -0.017 | 0.017 | -0.008 | 0.021 | -0.042 | 0.024 | -0.055 | 0.025 |
| rs6979832 | A | G | -0.006 | 0.014 | -0.016 | 0.016 | -0.029 | 0.020 | -0.006 | 0.022 | 0.023 | 0.023 |
| rs13233916 | C | G | 0.024 | 0.025 | 0.019 | 0.031 | 0.021 | 0.038 | 0.032 | 0.043 | 0.096 | 0.045 |
| rs7005216 | G | C | 0.019 | 0.015 | -0.001 | 0.017 | -0.030 | 0.021 | 0.045 | 0.024 | 0.041 | 0.025 |
| rs351776 | A | C | -0.034 | 0.014 | -0.043 | 0.016 | -0.050 | 0.020 | -0.046 | 0.022 | -0.065 | 0.023 |

| | | | | | | | | | | | |
|---|---|---|---|---|---|---|---|---|---|---|---|
| rs62515439 | C | T | -0.006 | 0.014 | 0.007 | 0.017 | -0.008 | 0.021 | 0.033 | 0.023 | -0.014 | 0.024 |
| rs13254613 | A | C | 0.011 | 0.014 | 0.043 | 0.017 | 0.065 | 0.021 | 0.021 | 0.023 | 0.000 | 0.024 |
| rs2126474 | G | T | 0.006 | 0.014 | 0.009 | 0.016 | 0.021 | 0.020 | -0.011 | 0.022 | 0.026 | 0.023 |
| rs10821163 | G | C | 0.009 | 0.014 | 0.021 | 0.017 | 0.062 | 0.021 | -0.025 | 0.023 | -0.023 | 0.024 |
| rs957512 | T | C | 0.003 | 0.014 | 0.013 | 0.017 | 0.004 | 0.021 | 0.019 | 0.023 | -0.011 | 0.024 |
| rs2275241 | G | A | -0.013 | 0.015 | -0.017 | 0.017 | -0.012 | 0.021 | -0.023 | 0.023 | 0.003 | 0.024 |
| rs7084503 | T | C | 0.025 | 0.014 | 0.018 | 0.016 | 0.001 | 0.020 | 0.043 | 0.022 | 0.048 | 0.023 |
| rs76971642 | T | C | 0.034 | 0.033 | 0.053 | 0.039 | 0.078 | 0.048 | 0.022 | 0.053 | 0.039 | 0.055 |
| rs962369 | T | C | 0.005 | 0.015 | -0.010 | 0.018 | -0.013 | 0.022 | -0.015 | 0.024 | 0.028 | 0.026 |
| rs661878 | A | G | -0.012 | 0.019 | -0.014 | 0.023 | -0.018 | 0.028 | -0.008 | 0.031 | -0.013 | 0.033 |
| rs11039307 | C | T | 0.002 | 0.014 | 0.001 | 0.016 | -0.011 | 0.020 | 0.011 | 0.022 | 0.001 | 0.023 |
| rs678653 | C | G | -0.011 | 0.015 | -0.009 | 0.017 | 0.003 | 0.021 | -0.021 | 0.023 | -0.013 | 0.024 |
| rs11215403 | G | A | -0.045 | 0.016 | -0.032 | 0.019 | -0.026 | 0.024 | -0.023 | 0.026 | -0.061 | 0.027 |
| rs11611246 | G | T | -0.017 | 0.017 | -0.028 | 0.020 | -0.043 | 0.025 | -0.004 | 0.027 | 0.040 | 0.029 |
| rs2187642 | A | C | 0.012 | 0.014 | 0.030 | 0.017 | 0.027 | 0.021 | 0.036 | 0.023 | 0.010 | 0.024 |
| rs10876457 | G | A | 0.004 | 0.017 | 0.016 | 0.019 | 0.014 | 0.024 | 0.016 | 0.026 | -0.005 | 0.027 |
| rs10783302 | G | T | 0.052 | 0.014 | 0.046 | 0.017 | 0.037 | 0.021 | 0.062 | 0.023 | 0.038 | 0.024 |
| rs7132908 | G | A | -0.021 | 0.014 | -0.015 | 0.017 | 0.001 | 0.021 | -0.032 | 0.023 | -0.033 | 0.024 |
| rs76919525 | T | A | -0.047 | 0.015 | -0.039 | 0.017 | -0.035 | 0.021 | -0.045 | 0.023 | -0.071 | 0.025 |
| rs78607331 | C | T | -0.016 | 0.041 | -0.008 | 0.048 | -0.041 | 0.058 | 0.028 | 0.065 | -0.028 | 0.066 |
| rs10784514 | C | T | 0.020 | 0.016 | 0.022 | 0.018 | 0.022 | 0.023 | 0.038 | 0.025 | 0.033 | 0.026 |
| rs2364232 | A | C | 0.005 | 0.016 | -0.015 | 0.018 | 0.010 | 0.023 | -0.039 | 0.025 | 0.020 | 0.026 |
| rs12309017 | G | T | -0.016 | 0.016 | -0.023 | 0.019 | -0.054 | 0.023 | 0.011 | 0.026 | 0.015 | 0.027 |
| rs11111647 | G | A | -0.007 | 0.017 | -0.018 | 0.020 | -0.016 | 0.025 | -0.020 | 0.027 | 0.000 | 0.029 |
| rs7305424 | A | T | 0.018 | 0.014 | 0.029 | 0.017 | 0.014 | 0.021 | 0.043 | 0.023 | -0.012 | 0.024 |
| rs35202265 | C | T | 0.010 | 0.014 | 0.003 | 0.017 | 0.020 | 0.021 | -0.008 | 0.023 | 0.045 | 0.024 |
| rs9551428 | C | T | -0.026 | 0.014 | -0.020 | 0.016 | -0.009 | 0.020 | -0.032 | 0.022 | -0.017 | 0.023 |
| rs1336486 | T | G | 0.015 | 0.014 | 0.015 | 0.017 | 0.002 | 0.021 | 0.028 | 0.023 | 0.024 | 0.025 |
| rs4477562 | C | T | 0.003 | 0.020 | -0.007 | 0.024 | -0.017 | 0.029 | -0.002 | 0.032 | 0.049 | 0.034 |
| rs9317002 | C | A | -0.009 | 0.014 | -0.008 | 0.016 | 0.002 | 0.020 | -0.023 | 0.022 | -0.006 | 0.023 |
| rs58681688 | C | G | -0.017 | 0.017 | -0.025 | 0.021 | -0.015 | 0.026 | -0.037 | 0.028 | -0.039 | 0.030 |
| rs9540493 | A | G | 0.019 | 0.014 | 0.017 | 0.016 | 0.020 | 0.020 | 0.012 | 0.022 | 0.050 | 0.023 |
| rs1576655 | A | C | -0.017 | 0.014 | -0.026 | 0.017 | -0.025 | 0.021 | -0.025 | 0.023 | 0.006 | 0.024 |
| rs61980008 | G | A | -0.032 | 0.036 | 0.011 | 0.051 | 0.048 | 0.062 | -0.030 | 0.067 | 0.008 | 0.073 |
| rs1865719 | A | G | 0.006 | 0.014 | 0.006 | 0.018 | 0.025 | 0.024 | 0.017 | 0.027 | 0.059 | 0.029 |
| rs8030456 | C | T | 0.030 | 0.016 | 0.039 | 0.019 | 0.052 | 0.023 | 0.006 | 0.026 | 0.001 | 0.027 |
| rs4932430 | A | C | 0.016 | 0.014 | 0.015 | 0.017 | 0.026 | 0.020 | 0.005 | 0.023 | 0.004 | 0.024 |
| rs2970356 | C | G | 0.002 | 0.015 | -0.006 | 0.018 | 0.008 | 0.022 | -0.010 | 0.025 | 0.019 | 0.026 |
| rs72755233 | G | A | 0.020 | 0.026 | 0.017 | 0.030 | 0.002 | 0.037 | 0.041 | 0.042 | 0.014 | 0.043 |
| rs2531991 | G | A | -0.026 | 0.016 | -0.024 | 0.019 | -0.015 | 0.023 | -0.043 | 0.025 | -0.040 | 0.026 |
| rs148965598 | A | G | -0.015 | 0.021 | -0.036 | 0.024 | -0.014 | 0.030 | -0.066 | 0.033 | 0.033 | 0.035 |
| rs4432271 | C | T | -0.018 | 0.020 | 0.000 | 0.024 | -0.004 | 0.029 | 0.023 | 0.032 | 0.007 | 0.034 |
| rs7189927 | T | C | 0.012 | 0.015 | 0.019 | 0.017 | 0.027 | 0.021 | 0.009 | 0.023 | 0.010 | 0.024 |
| rs4889630 | T | C | 0.049 | 0.017 | 0.045 | 0.020 | 0.049 | 0.024 | 0.034 | 0.027 | 0.031 | 0.028 |
| rs1421085 | T | C | 0.005 | 0.014 | -0.011 | 0.016 | 0.003 | 0.020 | -0.032 | 0.022 | 0.017 | 0.023 |

| | | | | | | | | | | | |
|---|---|---|---|---|---|---|---|---|---|---|---|
| rs11863799 | C | T | -0.001 | 0.015 | -0.016 | 0.017 | -0.003 | 0.021 | -0.036 | 0.023 | 0.003 | 0.024 |
| rs34229857 | C | T | -0.046 | 0.045 | -0.002 | 0.052 | -0.024 | 0.063 | 0.023 | 0.072 | -0.136 | 0.070 |
| rs11642090 | T | C | -0.001 | 0.014 | 0.005 | 0.017 | 0.012 | 0.021 | 0.003 | 0.023 | -0.008 | 0.024 |
| rs242922 | A | C | 0.009 | 0.014 | 0.011 | 0.017 | -0.013 | 0.021 | 0.031 | 0.023 | -0.004 | 0.024 |
| rs999493 | G | A | 0.008 | 0.014 | 0.017 | 0.016 | 0.026 | 0.020 | 0.016 | 0.022 | 0.009 | 0.023 |
| rs12185242 | A | C | 0.029 | 0.014 | 0.029 | 0.016 | 0.022 | 0.020 | 0.043 | 0.022 | 0.025 | 0.023 |
| rs11150745 | A | G | -0.022 | 0.015 | -0.020 | 0.017 | -0.008 | 0.021 | -0.032 | 0.023 | -0.018 | 0.025 |
| rs1013737 | G | C | 0.020 | 0.014 | 0.014 | 0.016 | 0.044 | 0.020 | -0.030 | 0.022 | 0.000 | 0.023 |
| rs303753 | G | A | 0.001 | 0.015 | -0.005 | 0.017 | -0.006 | 0.021 | -0.008 | 0.023 | 0.007 | 0.024 |
| rs7239114 | G | A | 0.010 | 0.014 | 0.001 | 0.016 | -0.002 | 0.020 | 0.006 | 0.022 | 0.029 | 0.023 |
| rs12606230 | T | C | 0.009 | 0.016 | 0.003 | 0.019 | -0.006 | 0.024 | 0.024 | 0.026 | 0.014 | 0.028 |
| rs2168711 | T | C | -0.022 | 0.016 | -0.005 | 0.019 | -0.002 | 0.023 | -0.008 | 0.025 | -0.036 | 0.027 |
| rs17066856 | T | C | -0.002 | 0.023 | 0.003 | 0.028 | -0.016 | 0.034 | 0.043 | 0.038 | -0.009 | 0.039 |
| rs16982345 | G | A | 0.008 | 0.016 | 0.007 | 0.019 | 0.038 | 0.023 | -0.019 | 0.025 | 0.006 | 0.027 |
| rs3810304 | A | G | -0.006 | 0.016 | -0.009 | 0.020 | 0.001 | 0.024 | -0.026 | 0.027 | -0.008 | 0.028 |
| rs4805881 | A | C | 0.020 | 0.014 | 0.032 | 0.017 | 0.039 | 0.021 | 0.012 | 0.023 | -0.010 | 0.024 |
| rs3810291 | G | A | -0.021 | 0.015 | -0.019 | 0.018 | -0.023 | 0.021 | -0.012 | 0.024 | -0.016 | 0.025 |
| rs633372 | G | A | -0.027 | 0.014 | -0.041 | 0.016 | -0.058 | 0.020 | -0.026 | 0.022 | -0.031 | 0.023 |
| rs994308 | C | T | 0.091 | 0.014 | 0.082 | 0.016 | 0.070 | 0.020 | 0.097 | 0.022 | 0.098 | 0.023 |
| rs7268466 | C | T | -0.009 | 0.020 | 0.006 | 0.024 | 0.034 | 0.029 | -0.023 | 0.032 | 0.026 | 0.033 |
| rs947088 | G | T | -0.007 | 0.015 | -0.005 | 0.018 | -0.023 | 0.022 | 0.021 | 0.024 | -0.011 | 0.025 |
| rs66469746 | C | A | -0.011 | 0.015 | -0.011 | 0.018 | -0.006 | 0.022 | -0.020 | 0.024 | -0.040 | 0.025 |
| rs4817973 | G | A | -0.010 | 0.014 | -0.003 | 0.017 | 0.022 | 0.021 | -0.033 | 0.023 | -0.019 | 0.024 |
| rs6001872 | A | G | 0.024 | 0.014 | 0.021 | 0.017 | 0.017 | 0.021 | 0.034 | 0.023 | 0.043 | 0.024 |
| Adult body size SNPs (overall) | | | | | | | | | | | | |
| SNP | EA | OA | beta_ca | se_ca | beta_co | se_co | beta_prox | se_prox | beta_dist | se_dist | beta_re | se_re |
| rs4648450 | C | A | 0.024 | 0.010 | 0.016 | 0.012 | 0.018 | 0.015 | 0.021 | 0.015 | 0.031 | 0.015 |
| rs2076363 | C | G | -0.024 | 0.010 | -0.022 | 0.012 | -0.026 | 0.015 | -0.024 | 0.016 | -0.027 | 0.016 |
| rs4908677 | C | T | -0.004 | 0.010 | -0.001 | 0.012 | -0.003 | 0.015 | 0.000 | 0.015 | -0.014 | 0.015 |
| rs78886584 | A | G | -0.010 | 0.011 | -0.009 | 0.013 | -0.008 | 0.017 | -0.008 | 0.017 | -0.006 | 0.018 |
| rs10799778 | T | G | 0.027 | 0.013 | 0.035 | 0.015 | 0.039 | 0.019 | 0.031 | 0.020 | 0.043 | 0.020 |
| rs7511698 | C | T | -0.019 | 0.011 | -0.031 | 0.013 | -0.034 | 0.016 | -0.029 | 0.017 | -0.013 | 0.017 |
| rs945211 | G | C | 0.004 | 0.010 | 0.011 | 0.012 | 0.026 | 0.015 | -0.011 | 0.016 | 0.009 | 0.016 |
| rs3737992 | G | A | 0.005 | 0.012 | 0.002 | 0.015 | -0.002 | 0.019 | 0.002 | 0.019 | -0.013 | 0.020 |
| rs12031634 | G | A | 0.016 | 0.011 | 0.009 | 0.013 | 0.030 | 0.016 | -0.010 | 0.017 | 0.009 | 0.017 |
| rs116195355 | C | A | 0.011 | 0.032 | 0.002 | 0.037 | -0.039 | 0.046 | 0.027 | 0.049 | 0.023 | 0.049 |
| rs2744801 | C | T | 0.011 | 0.010 | 0.012 | 0.012 | -0.009 | 0.016 | 0.029 | 0.016 | 0.013 | 0.016 |
| rs4660586 | C | T | -0.010 | 0.011 | -0.017 | 0.013 | -0.024 | 0.017 | -0.008 | 0.017 | -0.007 | 0.017 |
| rs6669341 | A | G | -0.012 | 0.010 | -0.018 | 0.012 | -0.017 | 0.015 | -0.014 | 0.015 | -0.033 | 0.015 |
| rs1167311 | G | A | 0.011 | 0.011 | 0.010 | 0.013 | 0.009 | 0.016 | 0.006 | 0.017 | 0.016 | 0.017 |
| rs630602 | G | C | -0.004 | 0.010 | -0.014 | 0.012 | -0.007 | 0.015 | -0.017 | 0.016 | 0.015 | 0.016 |
| rs12140153 | G | T | 0.015 | 0.019 | 0.026 | 0.022 | 0.041 | 0.028 | -0.003 | 0.029 | -0.024 | 0.029 |
| rs11208659 | T | C | 0.004 | 0.016 | -0.005 | 0.019 | -0.001 | 0.025 | 0.002 | 0.025 | 0.036 | 0.026 |
| rs7519259 | G | A | 0.010 | 0.010 | 0.005 | 0.012 | 0.009 | 0.015 | 0.004 | 0.015 | 0.001 | 0.015 |
| rs2613499 | A | G | 0.013 | 0.013 | 0.023 | 0.015 | 0.015 | 0.019 | 0.039 | 0.020 | 0.001 | 0.020 |

| | | | | | | | | | | | |
|---|---|---|---|---|---|---|---|---|---|---|---|
| rs7553158 | G | A | 0.001 | 0.010 | 0.011 | 0.012 | 0.009 | 0.015 | 0.019 | 0.015 | 0.006 | 0.015 |
| rs115778101 | T | C | 0.029 | 0.024 | 0.038 | 0.029 | 0.079 | 0.037 | -0.006 | 0.037 | 0.046 | 0.038 |
| rs34517439 | C | A | -0.007 | 0.017 | -0.016 | 0.020 | -0.011 | 0.025 | -0.023 | 0.026 | -0.015 | 0.026 |
| rs651533 | T | A | 0.011 | 0.011 | 0.023 | 0.014 | 0.020 | 0.017 | 0.017 | 0.018 | 0.011 | 0.018 |
| rs28726372 | T | C | 0.016 | 0.011 | 0.023 | 0.013 | 0.034 | 0.017 | 0.014 | 0.017 | 0.003 | 0.017 |
| rs7548936 | G | C | 0.001 | 0.010 | 0.003 | 0.012 | 0.008 | 0.015 | -0.004 | 0.015 | 0.025 | 0.016 |
| rs6679458 | G | T | 0.001 | 0.010 | -0.002 | 0.012 | 0.005 | 0.015 | -0.008 | 0.015 | 0.013 | 0.015 |
| rs12072739 | A | G | 0.008 | 0.012 | 0.008 | 0.014 | 0.015 | 0.018 | 0.005 | 0.018 | 0.014 | 0.018 |
| rs41279738 | T | G | 0.003 | 0.029 | -0.012 | 0.034 | -0.015 | 0.044 | -0.002 | 0.045 | -0.004 | 0.045 |
| rs12033257 | A | G | 0.015 | 0.010 | 0.011 | 0.012 | 0.016 | 0.016 | 0.001 | 0.016 | 0.016 | 0.016 |
| rs7549358 | G | C | 0.015 | 0.010 | 0.010 | 0.012 | 0.008 | 0.015 | 0.017 | 0.016 | 0.020 | 0.016 |
| rs1409158 | C | T | -0.015 | 0.011 | -0.018 | 0.013 | -0.015 | 0.017 | -0.022 | 0.017 | -0.011 | 0.018 |
| rs142315514 | C | A | 0.001 | 0.030 | -0.001 | 0.036 | 0.021 | 0.045 | -0.017 | 0.046 | 0.011 | 0.047 |
| rs10749659 | C | T | -0.022 | 0.011 | -0.024 | 0.014 | -0.035 | 0.017 | -0.020 | 0.018 | -0.020 | 0.018 |
| rs3753639 | T | C | -0.001 | 0.012 | -0.003 | 0.014 | 0.005 | 0.017 | -0.017 | 0.018 | -0.024 | 0.018 |
| rs61813324 | C | T | -0.047 | 0.017 | -0.045 | 0.020 | -0.041 | 0.025 | -0.060 | 0.026 | -0.047 | 0.027 |
| rs1778830 | G | A | -0.021 | 0.010 | -0.020 | 0.012 | -0.028 | 0.015 | -0.011 | 0.016 | -0.024 | 0.016 |
| rs4916229 | C | G | -0.022 | 0.017 | -0.023 | 0.020 | -0.049 | 0.025 | 0.002 | 0.026 | -0.025 | 0.026 |
| rs148137538 | A | G | -0.049 | 0.036 | -0.058 | 0.042 | -0.004 | 0.054 | -0.100 | 0.055 | -0.033 | 0.056 |
| rs77560793 | G | A | -0.027 | 0.028 | -0.005 | 0.034 | 0.009 | 0.042 | -0.041 | 0.043 | -0.105 | 0.043 |
| rs539515 | A | C | -0.003 | 0.012 | 0.002 | 0.015 | -0.004 | 0.019 | 0.005 | 0.019 | 0.002 | 0.019 |
| rs9425634 | T | C | 0.009 | 0.010 | 0.009 | 0.012 | 0.015 | 0.015 | 0.000 | 0.015 | 0.003 | 0.015 |
| rs815163 | T | C | -0.015 | 0.010 | -0.015 | 0.012 | -0.012 | 0.015 | -0.021 | 0.015 | -0.018 | 0.015 |
| rs76702514 | C | G | 0.016 | 0.012 | 0.023 | 0.015 | 0.035 | 0.018 | 0.011 | 0.019 | 0.007 | 0.019 |
| rs2678204 | T | G | -0.027 | 0.010 | -0.035 | 0.012 | -0.045 | 0.016 | -0.029 | 0.016 | -0.005 | 0.016 |
| rs4971239 | G | A | -0.002 | 0.016 | -0.008 | 0.019 | -0.047 | 0.024 | 0.044 | 0.025 | 0.033 | 0.025 |
| rs7539903 | T | A | 0.007 | 0.010 | -0.001 | 0.012 | 0.002 | 0.015 | -0.011 | 0.015 | 0.004 | 0.016 |
| rs78508049 | T | C | -0.016 | 0.012 | -0.018 | 0.015 | -0.010 | 0.019 | -0.032 | 0.019 | -0.027 | 0.019 |
| rs12037905 | C | T | 0.005 | 0.010 | 0.001 | 0.012 | -0.009 | 0.015 | 0.013 | 0.015 | 0.008 | 0.015 |
| rs7518221 | T | C | 0.009 | 0.010 | 0.012 | 0.012 | 0.017 | 0.015 | 0.005 | 0.016 | -0.020 | 0.016 |
| rs10779835 | T | C | 0.002 | 0.010 | -0.008 | 0.012 | 0.002 | 0.015 | -0.018 | 0.015 | 0.009 | 0.015 |
| rs10927006 | T | C | 0.000 | 0.014 | -0.003 | 0.017 | 0.008 | 0.021 | -0.015 | 0.021 | -0.002 | 0.022 |
| rs4658403 | C | T | -0.001 | 0.013 | 0.001 | 0.015 | 0.011 | 0.019 | -0.005 | 0.020 | 0.011 | 0.020 |
| rs6752378 | C | A | -0.004 | 0.010 | -0.010 | 0.012 | -0.016 | 0.015 | -0.007 | 0.015 | -0.013 | 0.015 |
| rs1631026 | C | T | -0.024 | 0.010 | -0.027 | 0.012 | -0.033 | 0.015 | -0.022 | 0.015 | -0.027 | 0.015 |
| rs10204994 | G | A | 0.007 | 0.012 | 0.012 | 0.014 | -0.002 | 0.017 | 0.022 | 0.018 | 0.010 | 0.018 |
| rs10185199 | G | A | -0.009 | 0.011 | -0.008 | 0.013 | -0.008 | 0.017 | 0.005 | 0.017 | -0.006 | 0.017 |
| rs10169594 | T | C | -0.006 | 0.010 | -0.006 | 0.012 | 0.007 | 0.015 | -0.022 | 0.016 | -0.008 | 0.016 |
| rs35809007 | G | A | -0.001 | 0.010 | -0.007 | 0.012 | -0.021 | 0.015 | 0.002 | 0.016 | -0.004 | 0.016 |
| rs72618637 | T | A | -0.008 | 0.012 | -0.023 | 0.015 | -0.023 | 0.019 | -0.021 | 0.019 | -0.025 | 0.020 |
| rs6761463 | G | C | 0.000 | 0.013 | 0.008 | 0.016 | -0.007 | 0.020 | 0.035 | 0.021 | -0.010 | 0.021 |
| rs59428052 | A | G | 0.015 | 0.014 | 0.018 | 0.017 | 0.011 | 0.021 | 0.021 | 0.022 | 0.010 | 0.022 |
| rs7601895 | C | G | 0.010 | 0.011 | 0.013 | 0.013 | 0.022 | 0.016 | 0.003 | 0.017 | 0.019 | 0.017 |
| rs4671328 | T | G | -0.007 | 0.010 | -0.007 | 0.012 | -0.006 | 0.015 | -0.008 | 0.015 | 0.013 | 0.015 |
| rs4672338 | C | T | 0.006 | 0.010 | 0.009 | 0.012 | 0.018 | 0.016 | 0.000 | 0.016 | -0.007 | 0.016 |

| | | | | | | | | | | | |
|---|---|---|---|---|---|---|---|---|---|---|---|
| rs12477088 | T | C | -0.010 | 0.010 | -0.016 | 0.012 | -0.028 | 0.015 | 0.002 | 0.015 | -0.031 | 0.015 |
| rs6752979 | G | A | 0.009 | 0.010 | 0.009 | 0.012 | -0.012 | 0.016 | 0.024 | 0.016 | -0.008 | 0.016 |
| rs396354 | T | C | 0.008 | 0.011 | -0.001 | 0.013 | 0.004 | 0.016 | -0.009 | 0.017 | 0.009 | 0.017 |
| rs11691869 | C | A | 0.026 | 0.010 | 0.025 | 0.012 | 0.012 | 0.016 | 0.038 | 0.016 | 0.033 | 0.016 |
| rs1451533 | G | A | 0.009 | 0.011 | 0.002 | 0.013 | -0.008 | 0.016 | 0.023 | 0.017 | 0.013 | 0.017 |
| rs72851476 | A | C | -0.006 | 0.015 | -0.014 | 0.017 | 0.000 | 0.022 | -0.032 | 0.023 | 0.009 | 0.023 |
| rs62171698 | C | A | -0.007 | 0.014 | -0.005 | 0.016 | 0.002 | 0.021 | -0.015 | 0.021 | -0.007 | 0.022 |
| rs75706763 | A | G | -0.016 | 0.022 | 0.004 | 0.026 | -0.014 | 0.032 | 0.002 | 0.033 | -0.070 | 0.033 |
| rs409696 | G | A | 0.014 | 0.010 | 0.018 | 0.012 | 0.013 | 0.015 | 0.022 | 0.015 | 0.006 | 0.015 |
| rs12692596 | C | T | -0.013 | 0.010 | -0.017 | 0.012 | 0.003 | 0.015 | -0.037 | 0.016 | -0.010 | 0.016 |
| rs12477385 | G | T | 0.003 | 0.012 | -0.004 | 0.014 | -0.008 | 0.018 | 0.003 | 0.018 | 0.017 | 0.018 |
| rs788163 | A | C | 0.017 | 0.011 | 0.012 | 0.013 | 0.004 | 0.016 | 0.017 | 0.017 | 0.006 | 0.017 |
| rs72917533 | T | C | 0.004 | 0.013 | -0.006 | 0.015 | 0.003 | 0.019 | -0.012 | 0.020 | 0.013 | 0.020 |
| rs7570446 | C | A | -0.002 | 0.010 | -0.003 | 0.012 | 0.012 | 0.015 | -0.018 | 0.015 | -0.004 | 0.015 |
| rs1704190 | G | A | 0.022 | 0.010 | 0.032 | 0.012 | 0.031 | 0.015 | 0.032 | 0.016 | 0.017 | 0.016 |
| rs4482463 | C | A | 0.001 | 0.017 | 0.006 | 0.021 | -0.006 | 0.027 | 0.009 | 0.027 | -0.018 | 0.028 |
| rs79675564 | C | A | -0.004 | 0.020 | -0.011 | 0.024 | -0.010 | 0.030 | -0.009 | 0.031 | -0.019 | 0.031 |
| rs13427822 | A | G | 0.024 | 0.011 | 0.030 | 0.013 | 0.030 | 0.016 | 0.035 | 0.017 | 0.014 | 0.017 |
| rs55658481 | G | A | 0.013 | 0.010 | 0.010 | 0.012 | 0.003 | 0.016 | 0.013 | 0.016 | 0.035 | 0.016 |
| rs2433733 | G | A | 0.000 | 0.010 | -0.001 | 0.012 | -0.001 | 0.015 | -0.007 | 0.016 | 0.003 | 0.016 |
| rs4663213 | G | A | 0.015 | 0.012 | 0.021 | 0.014 | 0.035 | 0.017 | 0.004 | 0.018 | 0.028 | 0.018 |
| rs112380819 | G | A | 0.007 | 0.016 | 0.011 | 0.019 | 0.017 | 0.024 | 0.009 | 0.024 | 0.004 | 0.024 |
| rs34373881 | G | A | 0.001 | 0.011 | -0.013 | 0.013 | 0.009 | 0.017 | -0.034 | 0.017 | -0.006 | 0.017 |
| rs7619139 | T | A | 0.001 | 0.010 | 0.002 | 0.012 | -0.005 | 0.015 | 0.007 | 0.015 | -0.012 | 0.015 |
| rs80082536 | A | G | 0.006 | 0.016 | 0.007 | 0.019 | -0.004 | 0.024 | 0.014 | 0.025 | 0.034 | 0.025 |
| rs111768603 | G | T | 0.012 | 0.015 | 0.019 | 0.019 | 0.008 | 0.024 | 0.026 | 0.024 | 0.026 | 0.024 |
| rs28350 | A | G | -0.004 | 0.013 | 0.007 | 0.015 | -0.012 | 0.019 | 0.037 | 0.019 | -0.025 | 0.020 |
| rs78517245 | T | C | 0.001 | 0.047 | 0.004 | 0.056 | 0.040 | 0.072 | -0.076 | 0.074 | -0.085 | 0.073 |
| rs113706999 | T | A | 0.002 | 0.039 | 0.024 | 0.047 | 0.000 | 0.058 | 0.068 | 0.062 | -0.002 | 0.062 |
| rs9852062 | T | A | -0.003 | 0.010 | 0.001 | 0.012 | 0.003 | 0.015 | -0.007 | 0.015 | 0.012 | 0.015 |
| rs113569731 | C | A | 0.024 | 0.020 | 0.036 | 0.023 | 0.036 | 0.030 | 0.056 | 0.031 | 0.014 | 0.031 |
| rs9843653 | T | C | -0.008 | 0.010 | -0.008 | 0.012 | 0.000 | 0.015 | -0.014 | 0.015 | -0.033 | 0.015 |
| rs62259692 | G | A | 0.027 | 0.025 | 0.021 | 0.029 | -0.004 | 0.037 | 0.060 | 0.038 | 0.033 | 0.038 |
| rs6798941 | C | T | -0.015 | 0.010 | -0.018 | 0.013 | -0.019 | 0.016 | -0.017 | 0.016 | -0.008 | 0.017 |
| rs6445198 | G | T | 0.015 | 0.010 | 0.005 | 0.012 | -0.005 | 0.015 | 0.013 | 0.015 | 0.024 | 0.015 |
| rs6445258 | T | C | 0.002 | 0.013 | 0.010 | 0.015 | 0.004 | 0.019 | 0.019 | 0.020 | -0.017 | 0.020 |
| rs76824303 | A | C | 0.008 | 0.016 | 0.016 | 0.019 | 0.021 | 0.025 | 0.005 | 0.025 | 0.025 | 0.026 |
| rs557951 | T | G | -0.015 | 0.010 | -0.020 | 0.012 | -0.009 | 0.016 | -0.030 | 0.016 | -0.017 | 0.016 |
| rs11708540 | G | A | -0.027 | 0.014 | -0.023 | 0.016 | -0.033 | 0.021 | -0.010 | 0.021 | -0.032 | 0.021 |
| rs1598121 | A | G | -0.005 | 0.010 | 0.001 | 0.012 | 0.005 | 0.015 | 0.000 | 0.016 | -0.003 | 0.016 |
| rs114593013 | A | G | -0.003 | 0.021 | -0.026 | 0.025 | -0.030 | 0.032 | -0.009 | 0.033 | -0.001 | 0.033 |
| rs11915747 | C | G | -0.015 | 0.010 | -0.026 | 0.012 | -0.021 | 0.015 | -0.035 | 0.016 | -0.021 | 0.016 |
| rs4858940 | T | C | -0.006 | 0.017 | 0.000 | 0.020 | 0.014 | 0.025 | -0.010 | 0.026 | -0.036 | 0.026 |
| rs1454687 | C | G | -0.012 | 0.010 | -0.012 | 0.012 | -0.011 | 0.015 | -0.014 | 0.015 | -0.005 | 0.015 |
| rs1436348 | A | G | -0.011 | 0.010 | -0.016 | 0.012 | -0.011 | 0.015 | -0.023 | 0.015 | 0.004 | 0.015 |

| | | | | | | | | | | | |
|---|---|---|---|---|---|---|---|---|---|---|---|
| rs36131051 | T | G | 0.013 | 0.013 | 0.013 | 0.015 | 0.013 | 0.019 | 0.022 | 0.020 | 0.023 | 0.020 |
| rs9814758 | T | G | 0.001 | 0.010 | -0.002 | 0.012 | -0.005 | 0.015 | 0.005 | 0.016 | 0.011 | 0.016 |
| rs1320903 | G | A | -0.002 | 0.010 | 0.009 | 0.012 | -0.006 | 0.016 | 0.021 | 0.016 | -0.033 | 0.016 |
| rs10935143 | G | A | 0.003 | 0.010 | -0.009 | 0.012 | -0.007 | 0.015 | -0.014 | 0.015 | -0.002 | 0.015 |
| rs2343681 | G | A | -0.018 | 0.012 | -0.021 | 0.014 | -0.014 | 0.018 | -0.018 | 0.018 | -0.016 | 0.018 |
| rs2035936 | G | T | -0.001 | 0.020 | -0.016 | 0.026 | -0.055 | 0.033 | 0.042 | 0.035 | -0.004 | 0.034 |
| rs12634936 | T | C | 0.012 | 0.026 | 0.037 | 0.031 | 0.001 | 0.039 | 0.097 | 0.042 | -0.018 | 0.041 |
| rs1568488 | G | C | 0.012 | 0.010 | 0.017 | 0.012 | 0.035 | 0.015 | 0.003 | 0.016 | 0.012 | 0.016 |
| rs9834519 | C | T | 0.000 | 0.019 | -0.002 | 0.023 | 0.012 | 0.029 | -0.011 | 0.029 | 0.001 | 0.029 |
| rs12630209 | T | G | -0.002 | 0.011 | -0.014 | 0.013 | -0.010 | 0.017 | -0.021 | 0.017 | 0.008 | 0.018 |
| rs8192675 | T | C | 0.012 | 0.010 | 0.012 | 0.013 | -0.005 | 0.016 | 0.025 | 0.016 | 0.013 | 0.016 |
| rs529200 | A | G | -0.003 | 0.010 | -0.004 | 0.012 | -0.007 | 0.015 | -0.001 | 0.015 | -0.017 | 0.015 |
| rs13061117 | T | C | 0.012 | 0.016 | 0.025 | 0.019 | 0.020 | 0.025 | 0.032 | 0.025 | 0.003 | 0.025 |
| rs262956 | T | G | -0.006 | 0.010 | -0.008 | 0.012 | -0.009 | 0.015 | -0.001 | 0.016 | -0.028 | 0.016 |
| rs869400 | T | G | 0.021 | 0.013 | 0.020 | 0.015 | 0.025 | 0.019 | 0.013 | 0.020 | 0.002 | 0.020 |
| rs80236973 | C | T | 0.013 | 0.015 | 0.035 | 0.018 | 0.028 | 0.022 | 0.050 | 0.023 | 0.017 | 0.023 |
| rs4677813 | T | C | 0.027 | 0.011 | 0.020 | 0.013 | 0.021 | 0.017 | 0.012 | 0.018 | 0.025 | 0.018 |
| rs6583310 | G | C | -0.001 | 0.010 | -0.008 | 0.012 | -0.013 | 0.015 | -0.001 | 0.016 | 0.008 | 0.016 |
| rs2051559 | T | C | -0.017 | 0.014 | -0.010 | 0.017 | 0.003 | 0.022 | -0.034 | 0.022 | -0.059 | 0.022 |
| rs35852935 | A | C | 0.027 | 0.027 | 0.066 | 0.032 | 0.045 | 0.041 | 0.082 | 0.042 | 0.032 | 0.041 |
| rs1477890 | A | G | -0.017 | 0.010 | -0.013 | 0.012 | -0.010 | 0.015 | -0.017 | 0.015 | -0.039 | 0.015 |
| rs34811474 | G | A | 0.022 | 0.013 | 0.038 | 0.016 | 0.033 | 0.020 | 0.043 | 0.021 | 0.016 | 0.021 |
| rs73213484 | A | T | 0.008 | 0.013 | 0.014 | 0.016 | 0.010 | 0.020 | 0.020 | 0.021 | 0.036 | 0.021 |
| rs4527444 | A | G | -0.002 | 0.010 | -0.005 | 0.012 | 0.013 | 0.015 | -0.024 | 0.015 | 0.001 | 0.015 |
| rs36023504 | C | T | 0.005 | 0.010 | 0.009 | 0.012 | 0.021 | 0.016 | -0.003 | 0.016 | 0.010 | 0.016 |
| rs12507026 | A | T | 0.000 | 0.010 | -0.005 | 0.012 | -0.007 | 0.015 | 0.002 | 0.015 | 0.000 | 0.015 |
| rs2237025 | T | C | -0.043 | 0.010 | -0.024 | 0.012 | -0.024 | 0.015 | -0.029 | 0.015 | -0.067 | 0.015 |
| rs28462076 | A | G | 0.008 | 0.011 | -0.001 | 0.014 | -0.005 | 0.017 | -0.005 | 0.018 | -0.005 | 0.018 |
| rs2164300 | C | T | -0.005 | 0.010 | -0.005 | 0.012 | 0.000 | 0.015 | -0.012 | 0.015 | -0.005 | 0.015 |
| rs13104584 | G | A | -0.012 | 0.010 | -0.011 | 0.012 | -0.011 | 0.015 | -0.011 | 0.015 | -0.005 | 0.015 |
| rs4148155 | A | G | 0.004 | 0.015 | 0.014 | 0.019 | 0.029 | 0.024 | 0.003 | 0.025 | 0.018 | 0.025 |
| rs4419475 | A | T | -0.001 | 0.010 | -0.007 | 0.012 | -0.025 | 0.015 | 0.019 | 0.016 | -0.012 | 0.016 |
| rs1229984 | T | C | -0.039 | 0.021 | -0.039 | 0.026 | -0.061 | 0.034 | -0.015 | 0.034 | -0.063 | 0.035 |
| rs2583410 | A | C | -0.008 | 0.013 | -0.022 | 0.016 | -0.050 | 0.020 | 0.007 | 0.021 | -0.004 | 0.021 |
| rs13107325 | C | T | 0.008 | 0.019 | 0.012 | 0.022 | -0.027 | 0.028 | 0.044 | 0.029 | -0.006 | 0.029 |
| rs1381010 | G | A | -0.006 | 0.010 | -0.004 | 0.013 | -0.012 | 0.016 | 0.001 | 0.016 | 0.002 | 0.016 |
| rs2952863 | T | G | -0.005 | 0.010 | -0.003 | 0.013 | -0.005 | 0.016 | 0.002 | 0.016 | -0.003 | 0.016 |
| rs1296328 | A | C | -0.012 | 0.010 | -0.001 | 0.012 | -0.009 | 0.015 | 0.008 | 0.015 | -0.025 | 0.015 |
| rs809955 | G | A | 0.010 | 0.010 | 0.006 | 0.012 | -0.015 | 0.015 | 0.027 | 0.016 | 0.013 | 0.016 |
| rs35390852 | G | A | -0.009 | 0.015 | -0.007 | 0.018 | -0.005 | 0.023 | -0.006 | 0.024 | -0.040 | 0.024 |
| rs12644329 | G | A | -0.014 | 0.010 | -0.016 | 0.012 | -0.009 | 0.015 | -0.031 | 0.015 | -0.002 | 0.016 |
| rs6823268 | A | G | 0.020 | 0.010 | 0.014 | 0.012 | 0.016 | 0.015 | 0.013 | 0.015 | 0.034 | 0.016 |
| rs113079574 | C | T | 0.005 | 0.012 | 0.016 | 0.014 | 0.027 | 0.018 | 0.000 | 0.019 | -0.004 | 0.019 |
| rs6843852 | C | T | -0.020 | 0.010 | -0.028 | 0.012 | -0.018 | 0.015 | -0.032 | 0.015 | -0.001 | 0.015 |
| rs698147 | A | G | -0.003 | 0.010 | -0.011 | 0.012 | -0.009 | 0.015 | -0.010 | 0.015 | 0.001 | 0.015 |

| | | | | | | | | | | | |
|---|---|---|---|---|---|---|---|---|---|---|---|
| rs67913249 | C | G | 0.020 | 0.010 | 0.022 | 0.012 | 0.022 | 0.015 | 0.016 | 0.016 | 0.038 | 0.016 |
| rs114263339 | C | T | 0.028 | 0.032 | 0.022 | 0.038 | 0.006 | 0.048 | 0.063 | 0.050 | -0.035 | 0.049 |
| rs10805383 | G | A | 0.025 | 0.010 | 0.022 | 0.012 | 0.020 | 0.015 | 0.027 | 0.015 | 0.018 | 0.015 |
| rs9291822 | C | T | 0.006 | 0.010 | 0.013 | 0.012 | 0.008 | 0.015 | 0.018 | 0.015 | -0.009 | 0.015 |
| rs27215 | C | A | -0.006 | 0.011 | -0.002 | 0.014 | -0.005 | 0.017 | 0.008 | 0.018 | -0.008 | 0.018 |
| rs2307111 | T | C | 0.016 | 0.010 | 0.008 | 0.012 | -0.003 | 0.015 | 0.018 | 0.015 | 0.009 | 0.015 |
| rs252749 | G | A | -0.008 | 0.011 | 0.003 | 0.014 | 0.015 | 0.017 | -0.020 | 0.018 | -0.022 | 0.018 |
| rs59893724 | A | G | 0.004 | 0.012 | 0.005 | 0.014 | 0.015 | 0.017 | -0.013 | 0.018 | 0.003 | 0.018 |
| rs79236537 | G | T | 0.012 | 0.032 | 0.019 | 0.038 | -0.005 | 0.048 | 0.051 | 0.051 | 0.104 | 0.051 |
| rs6870983 | C | T | -0.031 | 0.012 | -0.036 | 0.014 | -0.032 | 0.017 | -0.043 | 0.018 | -0.011 | 0.018 |
| rs1477290 | T | C | 0.004 | 0.013 | -0.009 | 0.016 | -0.002 | 0.020 | -0.020 | 0.021 | 0.020 | 0.021 |
| rs142503704 | G | A | 0.041 | 0.042 | 0.044 | 0.049 | 0.049 | 0.063 | 0.029 | 0.065 | 0.011 | 0.066 |
| rs62379271 | T | G | -0.008 | 0.010 | -0.008 | 0.012 | -0.009 | 0.015 | -0.003 | 0.015 | -0.009 | 0.015 |
| rs149457 | C | T | 0.008 | 0.013 | 0.008 | 0.015 | 0.020 | 0.019 | -0.003 | 0.020 | 0.014 | 0.020 |
| rs12517187 | C | T | -0.004 | 0.010 | -0.010 | 0.012 | -0.019 | 0.015 | -0.008 | 0.015 | 0.008 | 0.015 |
| rs347551 | C | G | 0.000 | 0.010 | -0.002 | 0.012 | 0.001 | 0.015 | -0.006 | 0.016 | 0.003 | 0.016 |
| rs1582931 | G | A | -0.008 | 0.010 | -0.016 | 0.012 | -0.006 | 0.015 | -0.024 | 0.015 | -0.017 | 0.015 |
| rs4836133 | C | A | -0.004 | 0.010 | -0.014 | 0.012 | -0.022 | 0.015 | -0.006 | 0.015 | 0.002 | 0.015 |
| rs329118 | C | T | 0.005 | 0.010 | 0.008 | 0.012 | 0.018 | 0.015 | 0.000 | 0.015 | 0.025 | 0.015 |
| rs13174863 | A | G | -0.029 | 0.013 | -0.032 | 0.016 | -0.027 | 0.020 | -0.034 | 0.021 | -0.035 | 0.021 |
| rs7719067 | A | G | 0.012 | 0.010 | 0.018 | 0.012 | 0.014 | 0.015 | 0.009 | 0.015 | -0.011 | 0.015 |
| rs11134512 | T | G | 0.009 | 0.010 | 0.006 | 0.012 | 0.015 | 0.016 | -0.009 | 0.016 | 0.009 | 0.016 |
| rs11134679 | A | G | 0.005 | 0.010 | 0.009 | 0.012 | 0.009 | 0.016 | 0.009 | 0.016 | -0.009 | 0.016 |
| rs4467770 | G | A | 0.018 | 0.011 | 0.014 | 0.013 | 0.009 | 0.016 | 0.025 | 0.017 | 0.032 | 0.017 |
| rs9395520 | C | T | 0.006 | 0.010 | 0.005 | 0.013 | 0.006 | 0.016 | -0.001 | 0.016 | 0.016 | 0.016 |
| rs3806114 | G | A | -0.008 | 0.010 | -0.004 | 0.013 | 0.030 | 0.016 | -0.038 | 0.016 | -0.010 | 0.016 |
| rs75499503 | C | T | 0.017 | 0.012 | 0.026 | 0.014 | 0.035 | 0.017 | 0.023 | 0.018 | 0.035 | 0.018 |
| rs34388845 | A | G | -0.029 | 0.012 | -0.022 | 0.015 | -0.035 | 0.018 | -0.015 | 0.019 | -0.039 | 0.019 |
| rs2260051 | A | T | -0.030 | 0.010 | -0.034 | 0.012 | -0.036 | 0.015 | -0.034 | 0.016 | -0.043 | 0.016 |
| rs9277992 | G | A | 0.003 | 0.012 | -0.011 | 0.015 | -0.014 | 0.019 | -0.005 | 0.019 | 0.009 | 0.019 |
| rs9366863 | T | C | -0.007 | 0.010 | -0.002 | 0.012 | -0.034 | 0.015 | 0.028 | 0.016 | -0.039 | 0.016 |
| rs34298980 | T | C | -0.005 | 0.010 | -0.014 | 0.012 | -0.020 | 0.016 | -0.003 | 0.016 | 0.003 | 0.016 |
| rs9462670 | G | C | -0.005 | 0.011 | 0.001 | 0.014 | 0.007 | 0.017 | -0.010 | 0.018 | -0.015 | 0.018 |
| rs72892910 | G | T | 0.011 | 0.013 | 0.008 | 0.015 | 0.019 | 0.019 | 0.001 | 0.020 | 0.026 | 0.020 |
| rs1327259 | A | G | 0.012 | 0.010 | 0.013 | 0.012 | 0.015 | 0.015 | 0.008 | 0.015 | 0.004 | 0.016 |
| rs1547026 | T | C | 0.003 | 0.011 | 0.016 | 0.013 | 0.000 | 0.016 | 0.032 | 0.017 | -0.004 | 0.017 |
| rs72910629 | A | G | -0.014 | 0.014 | -0.032 | 0.017 | -0.025 | 0.022 | -0.032 | 0.022 | 0.014 | 0.023 |
| rs1040046 | C | A | 0.004 | 0.014 | 0.001 | 0.017 | 0.001 | 0.021 | 0.004 | 0.022 | 0.011 | 0.022 |
| rs1324110 | G | C | 0.005 | 0.010 | 0.009 | 0.012 | 0.018 | 0.015 | 0.001 | 0.015 | -0.011 | 0.015 |
| rs10499014 | C | G | -0.013 | 0.011 | -0.011 | 0.013 | -0.017 | 0.017 | -0.006 | 0.017 | -0.007 | 0.017 |
| rs6938973 | T | C | -0.029 | 0.010 | -0.023 | 0.012 | -0.002 | 0.015 | -0.046 | 0.015 | -0.035 | 0.015 |
| rs9496567 | G | A | 0.003 | 0.012 | -0.007 | 0.014 | -0.012 | 0.017 | 0.008 | 0.018 | 0.010 | 0.018 |
| rs156126 | T | C | -0.001 | 0.012 | -0.007 | 0.015 | -0.012 | 0.018 | -0.007 | 0.019 | 0.000 | 0.019 |
| rs2253310 | C | G | -0.014 | 0.010 | -0.017 | 0.012 | -0.034 | 0.015 | -0.008 | 0.015 | -0.007 | 0.015 |
| rs13218383 | C | G | -0.001 | 0.010 | -0.006 | 0.012 | -0.001 | 0.015 | -0.012 | 0.016 | -0.005 | 0.016 |

| | | | | | | | | | | | |
|---|---|---|---|---|---|---|---|---|---|---|---|
| rs2875762 | G | C | 0.003 | 0.011 | -0.002 | 0.014 | -0.013 | 0.017 | 0.006 | 0.018 | 0.001 | 0.018 |
| rs10457469 | G | A | -0.004 | 0.010 | 0.000 | 0.012 | 0.001 | 0.015 | 0.001 | 0.015 | -0.013 | 0.015 |
| rs12213441 | C | T | -0.003 | 0.012 | -0.010 | 0.015 | 0.006 | 0.018 | -0.018 | 0.019 | 0.009 | 0.019 |
| rs7749708 | C | T | -0.011 | 0.010 | 0.001 | 0.013 | 0.019 | 0.016 | -0.019 | 0.016 | -0.017 | 0.016 |
| rs9478496 | T | C | -0.018 | 0.013 | -0.012 | 0.016 | -0.017 | 0.020 | -0.003 | 0.020 | -0.007 | 0.020 |
| rs9480184 | C | T | 0.014 | 0.012 | 0.018 | 0.014 | 0.024 | 0.018 | 0.011 | 0.018 | 0.005 | 0.018 |
| rs36007635 | G | A | 0.029 | 0.014 | 0.042 | 0.017 | 0.061 | 0.022 | 0.025 | 0.022 | 0.016 | 0.022 |
| rs6950388 | G | A | -0.010 | 0.012 | -0.012 | 0.015 | -0.015 | 0.018 | -0.012 | 0.019 | -0.026 | 0.019 |
| rs2056477 | G | C | 0.010 | 0.011 | 0.009 | 0.013 | 0.009 | 0.017 | 0.010 | 0.017 | 0.021 | 0.017 |
| rs4307239 | A | G | -0.008 | 0.010 | -0.017 | 0.012 | -0.019 | 0.015 | -0.022 | 0.015 | 0.016 | 0.015 |
| rs215634 | A | G | -0.003 | 0.010 | -0.010 | 0.012 | -0.007 | 0.015 | -0.012 | 0.015 | -0.010 | 0.015 |
| rs2237402 | G | A | 0.013 | 0.010 | 0.022 | 0.012 | 0.036 | 0.015 | 0.006 | 0.016 | 0.004 | 0.016 |
| rs2289379 | C | T | 0.015 | 0.010 | 0.009 | 0.012 | 0.002 | 0.015 | 0.020 | 0.015 | 0.000 | 0.015 |
| rs3823674 | C | T | 0.000 | 0.010 | -0.017 | 0.012 | -0.014 | 0.015 | -0.020 | 0.015 | 0.035 | 0.015 |
| rs11765062 | T | C | -0.022 | 0.010 | -0.023 | 0.012 | -0.025 | 0.015 | -0.018 | 0.015 | 0.004 | 0.015 |
| rs2866720 | C | T | 0.008 | 0.010 | 0.005 | 0.012 | 0.019 | 0.015 | -0.015 | 0.016 | -0.009 | 0.016 |
| rs1852006 | G | A | -0.011 | 0.010 | -0.007 | 0.012 | -0.014 | 0.015 | 0.001 | 0.016 | -0.009 | 0.016 |
| rs6963840 | C | T | -0.004 | 0.014 | -0.007 | 0.016 | -0.004 | 0.020 | -0.018 | 0.021 | -0.017 | 0.021 |
| rs12538826 | T | C | 0.013 | 0.015 | 0.004 | 0.018 | -0.005 | 0.023 | 0.011 | 0.023 | 0.026 | 0.024 |
| rs2074686 | G | A | -0.016 | 0.010 | -0.021 | 0.012 | -0.017 | 0.015 | -0.026 | 0.015 | 0.016 | 0.015 |
| rs11496125 | C | T | 0.004 | 0.010 | 0.004 | 0.012 | 0.003 | 0.015 | 0.003 | 0.015 | -0.025 | 0.015 |
| rs2396625 | T | A | 0.010 | 0.010 | 0.011 | 0.012 | 0.017 | 0.015 | 0.003 | 0.015 | -0.018 | 0.015 |
| rs12705894 | G | A | -0.007 | 0.010 | -0.003 | 0.012 | -0.003 | 0.015 | 0.002 | 0.016 | -0.013 | 0.016 |
| rs1840660 | G | A | 0.003 | 0.010 | 0.011 | 0.012 | 0.006 | 0.015 | 0.017 | 0.016 | -0.013 | 0.016 |
| rs1899689 | C | T | -0.004 | 0.010 | 0.005 | 0.012 | -0.011 | 0.015 | 0.016 | 0.015 | -0.006 | 0.015 |
| rs35775580 | A | G | 0.029 | 0.026 | 0.033 | 0.031 | 0.049 | 0.039 | 0.024 | 0.040 | 0.040 | 0.040 |
| rs11976084 | C | T | -0.012 | 0.011 | -0.013 | 0.013 | -0.006 | 0.017 | -0.016 | 0.017 | -0.026 | 0.018 |
| rs11525873 | T | C | 0.017 | 0.016 | 0.015 | 0.020 | 0.008 | 0.026 | 0.028 | 0.027 | 0.043 | 0.027 |
| rs1805123 | T | G | 0.000 | 0.012 | 0.006 | 0.014 | 0.034 | 0.017 | -0.032 | 0.018 | -0.012 | 0.018 |
| rs7827182 | G | C | -0.009 | 0.011 | 0.005 | 0.013 | 0.025 | 0.016 | -0.018 | 0.017 | -0.026 | 0.017 |
| rs6601451 | C | G | 0.009 | 0.011 | -0.006 | 0.013 | -0.028 | 0.016 | 0.018 | 0.017 | 0.033 | 0.017 |
| rs55896564 | G | A | 0.010 | 0.011 | -0.010 | 0.013 | -0.018 | 0.016 | 0.003 | 0.017 | 0.029 | 0.017 |
| rs6530737 | A | G | -0.009 | 0.010 | -0.018 | 0.012 | -0.030 | 0.015 | -0.018 | 0.016 | 0.004 | 0.016 |
| rs2616143 | G | A | 0.016 | 0.011 | 0.017 | 0.013 | 0.013 | 0.016 | 0.010 | 0.016 | 0.016 | 0.016 |
| rs117176448 | C | G | 0.007 | 0.017 | 0.019 | 0.020 | 0.011 | 0.025 | 0.022 | 0.026 | 0.001 | 0.026 |
| rs4266606 | C | T | -0.001 | 0.014 | -0.005 | 0.017 | 0.011 | 0.021 | -0.024 | 0.022 | -0.005 | 0.022 |
| rs2725371 | A | G | 0.011 | 0.011 | 0.018 | 0.013 | 0.012 | 0.016 | 0.030 | 0.016 | -0.005 | 0.017 |
| rs4739558 | A | G | 0.007 | 0.010 | 0.011 | 0.012 | 0.004 | 0.015 | 0.014 | 0.015 | 0.008 | 0.016 |
| rs35894137 | C | T | 0.009 | 0.019 | 0.016 | 0.022 | -0.002 | 0.028 | 0.020 | 0.029 | 0.011 | 0.029 |
| rs143662847 | C | T | -0.025 | 0.032 | -0.032 | 0.038 | -0.081 | 0.047 | 0.025 | 0.051 | 0.002 | 0.053 |
| rs473837 | G | T | -0.012 | 0.010 | -0.016 | 0.012 | -0.013 | 0.015 | -0.015 | 0.016 | -0.005 | 0.016 |
| rs12681792 | C | A | -0.019 | 0.012 | -0.010 | 0.015 | -0.021 | 0.019 | 0.002 | 0.020 | -0.015 | 0.020 |
| rs4737188 | A | T | 0.010 | 0.010 | 0.020 | 0.012 | 0.031 | 0.015 | 0.008 | 0.015 | 0.020 | 0.015 |
| rs35957544 | G | T | -0.009 | 0.010 | -0.014 | 0.012 | -0.008 | 0.015 | -0.025 | 0.015 | -0.008 | 0.015 |
| rs2941432 | T | A | -0.018 | 0.010 | -0.010 | 0.012 | -0.016 | 0.015 | -0.011 | 0.015 | -0.035 | 0.016 |

| | | | | | | | | | | | |
|---|---|---|---|---|---|---|---|---|---|---|---|
| rs78565420 | C | T | -0.016 | 0.030 | -0.063 | 0.035 | -0.099 | 0.044 | -0.022 | 0.047 | -0.015 | 0.049 |
| rs17619860 | T | C | 0.009 | 0.013 | 0.005 | 0.016 | -0.002 | 0.020 | 0.011 | 0.021 | 0.029 | 0.021 |
| rs1905616 | G | A | 0.001 | 0.010 | 0.001 | 0.012 | -0.007 | 0.016 | 0.010 | 0.016 | 0.008 | 0.016 |
| rs2114210 | G | A | -0.010 | 0.010 | -0.009 | 0.012 | 0.022 | 0.016 | -0.036 | 0.016 | -0.005 | 0.016 |
| rs17716502 | C | T | 0.009 | 0.013 | 0.015 | 0.015 | 0.018 | 0.019 | 0.014 | 0.019 | 0.009 | 0.020 |
| rs72673947 | A | G | -0.006 | 0.015 | 0.002 | 0.018 | -0.007 | 0.023 | 0.003 | 0.023 | -0.006 | 0.024 |
| rs112875651 | G | A | 0.022 | 0.010 | 0.033 | 0.012 | 0.020 | 0.015 | 0.054 | 0.016 | -0.007 | 0.016 |
| rs11782074 | G | T | -0.007 | 0.010 | 0.001 | 0.012 | 0.007 | 0.015 | -0.009 | 0.016 | 0.000 | 0.016 |
| rs1865341 | C | T | -0.012 | 0.011 | -0.010 | 0.014 | -0.019 | 0.017 | -0.007 | 0.018 | -0.014 | 0.018 |
| rs10960276 | C | A | -0.018 | 0.010 | -0.019 | 0.012 | -0.019 | 0.015 | -0.019 | 0.016 | -0.005 | 0.016 |
| rs7020196 | C | T | 0.000 | 0.010 | 0.010 | 0.012 | 0.010 | 0.015 | 0.018 | 0.016 | -0.032 | 0.016 |
| rs13292699 | A | C | 0.021 | 0.010 | 0.027 | 0.012 | 0.044 | 0.015 | 0.001 | 0.015 | 0.008 | 0.015 |
| rs1411432 | A | C | -0.020 | 0.012 | -0.013 | 0.015 | -0.039 | 0.018 | 0.023 | 0.019 | 0.008 | 0.019 |
| rs17770336 | C | T | -0.027 | 0.010 | -0.022 | 0.013 | -0.036 | 0.016 | -0.023 | 0.016 | -0.018 | 0.016 |
| rs10969334 | C | A | 0.013 | 0.010 | 0.003 | 0.012 | -0.015 | 0.015 | 0.020 | 0.015 | 0.021 | 0.015 |
| rs10973159 | G | T | -0.004 | 0.010 | 0.004 | 0.012 | 0.014 | 0.015 | -0.006 | 0.016 | -0.017 | 0.016 |
| rs7038966 | C | T | -0.001 | 0.010 | 0.005 | 0.012 | -0.008 | 0.015 | 0.009 | 0.015 | -0.024 | 0.015 |
| rs1547205 | G | C | 0.025 | 0.017 | 0.014 | 0.020 | 0.036 | 0.026 | 0.001 | 0.026 | 0.045 | 0.027 |
| rs4989244 | G | A | -0.002 | 0.010 | 0.000 | 0.012 | -0.003 | 0.015 | -0.002 | 0.015 | 0.005 | 0.015 |
| rs2135745 | C | G | 0.012 | 0.011 | 0.006 | 0.013 | 0.007 | 0.017 | 0.013 | 0.017 | 0.025 | 0.017 |
| rs2417998 | C | G | -0.015 | 0.011 | -0.022 | 0.013 | -0.028 | 0.016 | -0.021 | 0.017 | -0.020 | 0.017 |
| rs12376870 | G | A | 0.018 | 0.011 | 0.021 | 0.014 | 0.013 | 0.017 | 0.030 | 0.018 | 0.013 | 0.018 |
| rs7038943 | T | C | 0.020 | 0.010 | 0.032 | 0.012 | 0.030 | 0.015 | 0.037 | 0.015 | 0.025 | 0.016 |
| rs6478538 | A | G | -0.027 | 0.010 | -0.024 | 0.012 | -0.015 | 0.016 | -0.035 | 0.016 | -0.042 | 0.016 |
| rs10760277 | C | T | 0.001 | 0.010 | -0.008 | 0.012 | -0.025 | 0.015 | 0.003 | 0.016 | 0.011 | 0.016 |
| rs7030609 | A | G | -0.024 | 0.017 | -0.018 | 0.021 | 0.016 | 0.027 | -0.051 | 0.027 | -0.028 | 0.027 |
| rs113132247 | G | A | -0.002 | 0.013 | 0.021 | 0.016 | 0.024 | 0.020 | 0.015 | 0.021 | -0.012 | 0.021 |
| rs7913496 | C | T | 0.001 | 0.013 | -0.004 | 0.015 | -0.008 | 0.019 | -0.007 | 0.020 | 0.006 | 0.020 |
| rs7893571 | G | T | 0.001 | 0.010 | 0.005 | 0.012 | -0.009 | 0.016 | 0.020 | 0.016 | -0.005 | 0.016 |
| rs12253527 | G | A | -0.028 | 0.010 | -0.024 | 0.012 | -0.024 | 0.016 | -0.021 | 0.016 | -0.051 | 0.016 |
| rs71495049 | G | A | -0.012 | 0.017 | -0.023 | 0.021 | -0.021 | 0.026 | -0.029 | 0.027 | -0.038 | 0.027 |
| rs3125326 | A | C | -0.005 | 0.010 | -0.012 | 0.012 | -0.028 | 0.015 | 0.001 | 0.016 | -0.012 | 0.016 |
| rs7924036 | G | T | -0.008 | 0.010 | -0.012 | 0.012 | -0.016 | 0.015 | -0.002 | 0.015 | 0.002 | 0.015 |
| rs11000993 | T | C | 0.011 | 0.015 | 0.025 | 0.018 | 0.004 | 0.023 | 0.039 | 0.023 | -0.033 | 0.023 |
| rs1250597 | A | G | -0.025 | 0.010 | -0.029 | 0.012 | -0.029 | 0.015 | -0.030 | 0.015 | -0.022 | 0.015 |
| rs17399739 | A | G | 0.022 | 0.019 | 0.034 | 0.023 | 0.048 | 0.029 | 0.033 | 0.030 | 0.031 | 0.030 |
| rs2450444 | G | A | -0.019 | 0.010 | -0.019 | 0.012 | -0.024 | 0.016 | -0.013 | 0.016 | -0.007 | 0.016 |
| rs41310284 | C | A | -0.011 | 0.017 | -0.006 | 0.020 | -0.013 | 0.025 | 0.006 | 0.026 | -0.010 | 0.026 |
| rs10736156 | C | A | -0.006 | 0.013 | -0.007 | 0.016 | -0.012 | 0.020 | 0.002 | 0.020 | -0.005 | 0.020 |
| rs7086898 | A | G | -0.022 | 0.016 | -0.032 | 0.020 | -0.042 | 0.025 | -0.016 | 0.026 | 0.002 | 0.026 |
| rs4575195 | C | A | -0.002 | 0.011 | 0.000 | 0.012 | 0.011 | 0.016 | -0.014 | 0.016 | 0.000 | 0.016 |
| rs9421249 | C | T | 0.025 | 0.011 | 0.010 | 0.013 | 0.000 | 0.017 | 0.022 | 0.017 | 0.020 | 0.017 |
| rs845084 | G | A | 0.011 | 0.011 | 0.009 | 0.013 | 0.012 | 0.017 | 0.007 | 0.017 | 0.011 | 0.017 |
| rs4962725 | T | C | -0.005 | 0.010 | -0.008 | 0.012 | -0.011 | 0.015 | -0.007 | 0.015 | -0.006 | 0.015 |
| rs2542615 | C | T | -0.008 | 0.010 | -0.001 | 0.012 | 0.010 | 0.015 | -0.009 | 0.016 | -0.018 | 0.016 |

| | | | | | | | | | | | |
|---|---|---|---|---|---|---|---|---|---|---|---|
| rs2035806 | G | A | 0.009 | 0.010 | 0.004 | 0.012 | 0.006 | 0.015 | 0.005 | 0.015 | 0.013 | 0.015 |
| rs67257872 | A | G | 0.002 | 0.010 | 0.010 | 0.012 | 0.009 | 0.015 | 0.013 | 0.015 | -0.001 | 0.015 |
| rs28711392 | T | C | -0.012 | 0.010 | -0.005 | 0.012 | -0.003 | 0.015 | -0.002 | 0.015 | -0.029 | 0.015 |
| rs6265 | C | T | 0.008 | 0.012 | 0.015 | 0.015 | 0.020 | 0.018 | 0.010 | 0.019 | 0.010 | 0.019 |
| rs10835498 | G | A | -0.001 | 0.010 | 0.001 | 0.012 | 0.014 | 0.015 | -0.019 | 0.015 | -0.006 | 0.015 |
| rs1222216 | C | T | 0.002 | 0.012 | -0.004 | 0.014 | -0.019 | 0.018 | 0.004 | 0.019 | -0.002 | 0.019 |
| rs59227842 | A | G | -0.013 | 0.011 | -0.017 | 0.013 | -0.007 | 0.016 | -0.024 | 0.016 | 0.011 | 0.017 |
| rs868784 | G | A | -0.012 | 0.010 | -0.012 | 0.012 | -0.023 | 0.015 | -0.003 | 0.016 | -0.022 | 0.016 |
| rs6416134 | G | C | 0.002 | 0.012 | 0.014 | 0.015 | 0.025 | 0.018 | 0.007 | 0.019 | 0.000 | 0.019 |
| rs12798028 | C | T | -0.008 | 0.010 | -0.004 | 0.012 | -0.013 | 0.015 | 0.000 | 0.015 | -0.006 | 0.015 |
| rs12363672 | A | C | -0.072 | 0.055 | -0.058 | 0.064 | -0.081 | 0.081 | -0.094 | 0.088 | -0.183 | 0.092 |
| rs34292685 | C | T | 0.023 | 0.013 | 0.023 | 0.016 | 0.005 | 0.020 | 0.033 | 0.021 | 0.007 | 0.021 |
| rs2234458 | C | T | 0.026 | 0.010 | 0.033 | 0.012 | 0.025 | 0.015 | 0.046 | 0.016 | 0.021 | 0.016 |
| rs667515 | G | C | 0.033 | 0.010 | 0.043 | 0.012 | 0.043 | 0.015 | 0.037 | 0.016 | 0.019 | 0.016 |
| rs10160769 | G | C | 0.001 | 0.011 | 0.004 | 0.014 | 0.012 | 0.017 | -0.011 | 0.018 | 0.013 | 0.018 |
| rs7102934 | T | C | -0.002 | 0.010 | 0.004 | 0.012 | -0.010 | 0.016 | 0.025 | 0.016 | 0.002 | 0.016 |
| rs61903695 | A | G | 0.002 | 0.011 | 0.008 | 0.013 | -0.007 | 0.017 | 0.027 | 0.017 | 0.014 | 0.018 |
| rs2658797 | C | T | 0.015 | 0.010 | 0.018 | 0.012 | 0.029 | 0.015 | 0.003 | 0.015 | -0.014 | 0.015 |
| rs680071 | T | C | -0.017 | 0.015 | -0.009 | 0.018 | -0.011 | 0.023 | -0.012 | 0.023 | -0.006 | 0.023 |
| rs719802 | T | C | 0.002 | 0.010 | 0.006 | 0.012 | -0.015 | 0.015 | 0.033 | 0.016 | 0.003 | 0.016 |
| rs11607476 | A | C | 0.010 | 0.010 | 0.015 | 0.012 | 0.018 | 0.015 | 0.006 | 0.015 | 0.000 | 0.015 |
| rs7928320 | C | G | 0.002 | 0.020 | 0.002 | 0.024 | 0.020 | 0.031 | -0.023 | 0.031 | 0.030 | 0.032 |
| rs12281009 | A | G | -0.002 | 0.020 | -0.004 | 0.024 | 0.014 | 0.030 | -0.022 | 0.031 | 0.021 | 0.032 |
| rs7925100 | G | A | -0.018 | 0.010 | -0.013 | 0.012 | -0.001 | 0.015 | -0.023 | 0.015 | -0.028 | 0.016 |
| rs11218510 | G | A | -0.002 | 0.010 | -0.007 | 0.012 | -0.009 | 0.015 | -0.012 | 0.015 | 0.018 | 0.015 |
| rs10791113 | A | G | -0.015 | 0.010 | -0.010 | 0.012 | -0.006 | 0.015 | -0.013 | 0.015 | -0.021 | 0.015 |
| rs12788343 | T | C | 0.005 | 0.010 | 0.004 | 0.012 | 0.009 | 0.015 | -0.003 | 0.015 | 0.013 | 0.016 |
| rs11223204 | A | G | 0.009 | 0.010 | 0.014 | 0.012 | 0.016 | 0.015 | 0.012 | 0.015 | 0.012 | 0.015 |
| rs329651 | G | T | -0.006 | 0.013 | -0.001 | 0.015 | -0.013 | 0.019 | 0.011 | 0.019 | -0.005 | 0.020 |
| rs61909165 | T | A | -0.012 | 0.013 | 0.001 | 0.016 | -0.012 | 0.020 | 0.006 | 0.021 | -0.065 | 0.021 |
| rs55726687 | G | A | -0.015 | 0.012 | -0.026 | 0.014 | -0.033 | 0.018 | -0.016 | 0.019 | 0.007 | 0.019 |
| rs10774018 | G | C | -0.014 | 0.011 | -0.027 | 0.014 | -0.031 | 0.017 | -0.030 | 0.018 | -0.010 | 0.018 |
| rs1799507 | G | A | -0.013 | 0.014 | -0.005 | 0.017 | -0.007 | 0.021 | -0.002 | 0.022 | 0.010 | 0.022 |
| rs10505836 | A | C | -0.023 | 0.013 | -0.002 | 0.016 | 0.001 | 0.020 | -0.001 | 0.021 | -0.042 | 0.021 |
| rs10842231 | A | T | -0.007 | 0.016 | -0.014 | 0.020 | -0.003 | 0.025 | -0.021 | 0.026 | 0.019 | 0.026 |
| rs1458156 | C | T | 0.008 | 0.010 | 0.020 | 0.012 | 0.026 | 0.015 | 0.020 | 0.015 | 0.008 | 0.015 |
| rs1126930 | G | C | -0.004 | 0.029 | -0.020 | 0.035 | -0.007 | 0.044 | -0.050 | 0.046 | -0.034 | 0.046 |
| rs7132908 | G | A | -0.012 | 0.010 | -0.006 | 0.012 | 0.004 | 0.015 | -0.014 | 0.015 | -0.006 | 0.016 |
| rs4077093 | T | G | 0.008 | 0.012 | -0.002 | 0.014 | 0.014 | 0.018 | -0.021 | 0.019 | 0.020 | 0.018 |
| rs4759073 | G | A | -0.012 | 0.010 | -0.011 | 0.012 | -0.008 | 0.015 | -0.006 | 0.016 | -0.019 | 0.016 |
| rs4759228 | G | C | 0.003 | 0.011 | 0.017 | 0.014 | 0.011 | 0.017 | 0.038 | 0.018 | 0.009 | 0.018 |
| rs12821416 | C | T | -0.008 | 0.015 | -0.012 | 0.017 | -0.011 | 0.022 | -0.018 | 0.023 | -0.016 | 0.023 |
| rs61754230 | C | T | 0.014 | 0.049 | -0.006 | 0.058 | -0.021 | 0.073 | 0.016 | 0.077 | 0.056 | 0.077 |
| rs12427047 | C | T | -0.011 | 0.011 | -0.023 | 0.014 | -0.040 | 0.017 | -0.014 | 0.018 | 0.019 | 0.018 |
| rs2712667 | G | C | -0.009 | 0.010 | -0.005 | 0.012 | -0.001 | 0.015 | -0.008 | 0.015 | -0.028 | 0.016 |

| | | | | | | | | | | | |
|---|---|---|---|---|---|---|---|---|---|---|---|
| rs4764949 | A | G | 0.000 | 0.010 | 0.008 | 0.012 | -0.001 | 0.015 | 0.016 | 0.016 | -0.017 | 0.016 |
| rs6606686 | G | C | 0.009 | 0.011 | 0.020 | 0.013 | 0.010 | 0.016 | 0.028 | 0.016 | -0.004 | 0.017 |
| rs11513729 | C | T | 0.065 | 0.010 | 0.074 | 0.012 | 0.080 | 0.015 | 0.069 | 0.016 | 0.065 | 0.016 |
| rs181617194 | T | C | 0.020 | 0.032 | 0.025 | 0.038 | 0.072 | 0.048 | -0.001 | 0.049 | -0.004 | 0.050 |
| rs147730268 | G | T | 0.020 | 0.020 | 0.028 | 0.024 | 0.061 | 0.030 | -0.005 | 0.031 | 0.025 | 0.031 |
| rs9579775 | A | C | -0.034 | 0.016 | -0.043 | 0.020 | -0.039 | 0.025 | -0.044 | 0.025 | -0.021 | 0.026 |
| rs9507791 | G | A | -0.010 | 0.012 | -0.007 | 0.015 | -0.006 | 0.018 | 0.004 | 0.019 | -0.023 | 0.019 |
| rs1967772 | G | A | 0.006 | 0.011 | 0.011 | 0.013 | 0.011 | 0.016 | 0.010 | 0.017 | -0.009 | 0.017 |
| rs35193668 | C | T | -0.003 | 0.010 | 0.006 | 0.012 | -0.007 | 0.015 | 0.019 | 0.016 | -0.021 | 0.016 |
| rs61954177 | G | C | 0.009 | 0.010 | 0.016 | 0.012 | 0.010 | 0.016 | 0.019 | 0.016 | 0.013 | 0.016 |
| rs12429545 | G | A | 0.005 | 0.014 | 0.015 | 0.017 | 0.016 | 0.022 | 0.006 | 0.022 | 0.020 | 0.023 |
| rs7321285 | A | C | 0.016 | 0.012 | 0.016 | 0.014 | 0.008 | 0.018 | 0.024 | 0.019 | 0.014 | 0.019 |
| rs2576135 | T | A | 0.022 | 0.017 | 0.010 | 0.020 | 0.012 | 0.025 | 0.002 | 0.026 | 0.027 | 0.026 |
| rs9317002 | C | A | -0.009 | 0.010 | -0.009 | 0.012 | -0.003 | 0.015 | -0.022 | 0.015 | -0.011 | 0.015 |
| rs9529148 | G | A | 0.001 | 0.010 | 0.007 | 0.012 | -0.011 | 0.015 | 0.032 | 0.015 | 0.013 | 0.016 |
| rs1576655 | A | C | -0.017 | 0.010 | -0.021 | 0.012 | -0.020 | 0.015 | -0.024 | 0.016 | -0.009 | 0.016 |
| rs61971082 | T | G | -0.014 | 0.011 | -0.014 | 0.013 | -0.012 | 0.017 | -0.017 | 0.017 | -0.027 | 0.017 |
| rs7331420 | G | A | -0.011 | 0.011 | -0.021 | 0.013 | -0.003 | 0.016 | -0.038 | 0.017 | -0.002 | 0.017 |
| rs9888533 | C | T | 0.018 | 0.010 | 0.010 | 0.012 | 0.000 | 0.015 | 0.013 | 0.016 | 0.029 | 0.016 |
| rs9522180 | C | T | 0.015 | 0.010 | 0.013 | 0.012 | 0.002 | 0.015 | 0.013 | 0.015 | -0.010 | 0.015 |
| rs9515446 | A | G | 0.000 | 0.010 | 0.002 | 0.012 | 0.009 | 0.015 | -0.007 | 0.015 | -0.013 | 0.015 |
| rs8015400 | C | A | 0.009 | 0.012 | 0.010 | 0.012 | 0.014 | 0.015 | 0.003 | 0.016 | 0.014 | 0.016 |
| rs9788550 | G | C | -0.006 | 0.013 | 0.018 | 0.014 | -0.001 | 0.017 | 0.033 | 0.018 | -0.007 | 0.018 |
| rs12883788 | C | T | -0.007 | 0.011 | -0.007 | 0.012 | -0.008 | 0.015 | -0.004 | 0.015 | -0.010 | 0.015 |
| rs7141912 | A | T | -0.014 | 0.017 | 0.010 | 0.017 | 0.010 | 0.022 | 0.008 | 0.023 | -0.004 | 0.023 |
| rs8011566 | T | A | 0.034 | 0.012 | 0.007 | 0.012 | 0.009 | 0.015 | 0.005 | 0.015 | 0.029 | 0.015 |
| rs724623 | A | C | -0.001 | 0.011 | -0.003 | 0.012 | -0.014 | 0.015 | 0.010 | 0.015 | -0.025 | 0.015 |
| rs217672 | A | C | -0.001 | 0.013 | -0.009 | 0.013 | 0.006 | 0.017 | -0.020 | 0.017 | 0.002 | 0.017 |
| rs3902951 | T | G | 0.010 | 0.013 | 0.019 | 0.014 | 0.020 | 0.017 | 0.021 | 0.018 | 0.013 | 0.018 |
| rs61986330 | C | A | 0.009 | 0.013 | 0.028 | 0.013 | 0.021 | 0.017 | 0.036 | 0.017 | -0.013 | 0.017 |
| rs10146997 | A | G | -0.012 | 0.015 | -0.003 | 0.014 | 0.003 | 0.018 | -0.003 | 0.019 | 0.003 | 0.019 |
| rs8008772 | A | T | 0.013 | 0.014 | 0.005 | 0.014 | -0.013 | 0.017 | 0.025 | 0.018 | -0.005 | 0.018 |
| rs1286138 | T | G | -0.008 | 0.012 | -0.007 | 0.012 | -0.020 | 0.016 | 0.006 | 0.016 | 0.001 | 0.016 |
| rs6575340 | G | A | -0.004 | 0.012 | 0.012 | 0.012 | 0.004 | 0.015 | 0.024 | 0.016 | -0.004 | 0.016 |
| rs12885251 | G | A | 0.016 | 0.012 | 0.027 | 0.012 | 0.036 | 0.015 | 0.016 | 0.015 | 0.022 | 0.015 |
| rs12147845 | C | T | -0.035 | 0.019 | 0.001 | 0.019 | 0.014 | 0.023 | -0.012 | 0.024 | -0.020 | 0.024 |
| rs61992671 | A | G | 0.012 | 0.012 | 0.021 | 0.013 | 0.025 | 0.016 | 0.014 | 0.016 | 0.011 | 0.016 |
| rs7145882 | T | C | -0.009 | 0.012 | -0.004 | 0.012 | -0.006 | 0.015 | -0.002 | 0.016 | -0.010 | 0.016 |
| rs3759584 | T | C | 0.011 | 0.012 | 0.005 | 0.012 | -0.005 | 0.015 | 0.011 | 0.016 | 0.020 | 0.016 |
| rs76520838 | C | T | -0.040 | 0.027 | -0.073 | 0.032 | -0.099 | 0.040 | -0.054 | 0.041 | -0.011 | 0.042 |
| rs7182917 | T | C | 0.001 | 0.010 | -0.010 | 0.012 | 0.000 | 0.015 | -0.028 | 0.015 | 0.025 | 0.015 |
| rs2247401 | G | A | -0.015 | 0.011 | -0.022 | 0.013 | -0.030 | 0.017 | -0.010 | 0.017 | 0.010 | 0.017 |
| rs28465175 | A | G | 0.017 | 0.019 | 0.027 | 0.022 | 0.034 | 0.028 | 0.026 | 0.029 | 0.028 | 0.029 |
| rs7175642 | T | G | -0.013 | 0.011 | -0.019 | 0.013 | -0.022 | 0.016 | -0.014 | 0.017 | -0.004 | 0.017 |
| rs28408562 | C | G | -0.024 | 0.010 | -0.024 | 0.012 | -0.022 | 0.015 | -0.028 | 0.015 | -0.017 | 0.015 |

| | | | | | | | | | | | |
|---|---|---|---|---|---|---|---|---|---|---|---|
| rs1369159 | C | T | 0.002 | 0.010 | 0.007 | 0.012 | 0.017 | 0.015 | -0.001 | 0.016 | 0.020 | 0.016 |
| rs111584879 | T | C | -0.011 | 0.011 | -0.008 | 0.014 | 0.000 | 0.017 | -0.023 | 0.018 | -0.046 | 0.018 |
| rs2241420 | G | A | 0.044 | 0.011 | 0.043 | 0.014 | 0.057 | 0.017 | 0.028 | 0.018 | 0.012 | 0.018 |
| rs62004865 | T | A | -0.021 | 0.017 | -0.023 | 0.020 | -0.034 | 0.025 | -0.010 | 0.026 | -0.033 | 0.026 |
| rs11856579 | G | A | -0.010 | 0.011 | -0.019 | 0.013 | -0.021 | 0.017 | -0.024 | 0.017 | -0.011 | 0.017 |
| rs57488047 | T | C | 0.014 | 0.010 | 0.015 | 0.012 | 0.003 | 0.015 | 0.027 | 0.015 | 0.011 | 0.015 |
| rs34994596 | T | C | -0.030 | 0.011 | -0.017 | 0.013 | -0.018 | 0.016 | -0.010 | 0.017 | -0.046 | 0.017 |
| rs7498044 | G | A | 0.007 | 0.012 | 0.006 | 0.014 | 0.005 | 0.018 | 0.005 | 0.019 | 0.008 | 0.019 |
| rs8038574 | T | C | 0.002 | 0.010 | 0.002 | 0.012 | 0.001 | 0.015 | -0.001 | 0.016 | -0.005 | 0.016 |
| rs56803094 | A | G | -0.002 | 0.012 | -0.004 | 0.014 | -0.007 | 0.018 | -0.004 | 0.018 | -0.009 | 0.018 |
| rs412243 | T | C | 0.007 | 0.010 | 0.002 | 0.012 | 0.011 | 0.015 | -0.005 | 0.016 | 0.017 | 0.016 |
| rs2516726 | T | C | 0.013 | 0.012 | 0.014 | 0.014 | -0.003 | 0.018 | 0.028 | 0.019 | 0.001 | 0.018 |
| rs879620 | C | T | -0.010 | 0.010 | -0.004 | 0.012 | -0.004 | 0.015 | -0.012 | 0.015 | -0.012 | 0.015 |
| rs2660241 | T | C | 0.021 | 0.010 | 0.012 | 0.012 | -0.001 | 0.015 | 0.021 | 0.016 | 0.026 | 0.016 |
| rs11642387 | A | G | -0.029 | 0.017 | -0.030 | 0.020 | -0.054 | 0.025 | -0.004 | 0.027 | -0.036 | 0.026 |
| rs39674 | C | G | -0.002 | 0.010 | -0.013 | 0.012 | 0.004 | 0.016 | -0.035 | 0.016 | 0.000 | 0.016 |
| rs12927792 | C | T | -0.001 | 0.010 | -0.011 | 0.012 | -0.015 | 0.015 | -0.006 | 0.015 | 0.016 | 0.015 |
| rs8054082 | C | T | -0.010 | 0.014 | -0.018 | 0.017 | 0.004 | 0.022 | -0.041 | 0.022 | 0.017 | 0.022 |
| rs4432271 | C | T | -0.008 | 0.014 | -0.004 | 0.017 | -0.006 | 0.022 | 0.014 | 0.022 | -0.001 | 0.022 |
| rs11864909 | C | T | 0.002 | 0.011 | -0.009 | 0.013 | -0.003 | 0.016 | -0.012 | 0.017 | 0.012 | 0.017 |
| rs9922288 | A | G | -0.011 | 0.012 | -0.020 | 0.015 | -0.016 | 0.018 | -0.018 | 0.019 | 0.018 | 0.019 |
| rs7498665 | A | G | -0.011 | 0.010 | -0.016 | 0.012 | -0.020 | 0.015 | -0.017 | 0.016 | -0.007 | 0.016 |
| rs3814883 | C | T | -0.002 | 0.010 | -0.006 | 0.012 | 0.012 | 0.015 | -0.021 | 0.015 | 0.015 | 0.015 |
| rs34898535 | C | T | 0.021 | 0.010 | 0.019 | 0.012 | 0.033 | 0.015 | 0.009 | 0.016 | 0.017 | 0.016 |
| rs56094641 | A | G | 0.004 | 0.010 | -0.002 | 0.012 | 0.000 | 0.015 | -0.007 | 0.015 | 0.008 | 0.015 |
| rs862320 | C | T | 0.019 | 0.010 | 0.026 | 0.012 | 0.033 | 0.015 | 0.020 | 0.015 | -0.003 | 0.015 |
| rs12149660 | G | A | -0.030 | 0.016 | -0.030 | 0.019 | -0.010 | 0.025 | -0.045 | 0.025 | -0.021 | 0.026 |
| rs811054 | C | T | -0.010 | 0.010 | -0.014 | 0.012 | -0.030 | 0.015 | 0.001 | 0.015 | 0.003 | 0.015 |
| rs4500770 | A | T | 0.024 | 0.010 | 0.025 | 0.012 | 0.020 | 0.016 | 0.026 | 0.016 | 0.030 | 0.016 |
| rs9673839 | A | G | -0.004 | 0.010 | -0.006 | 0.012 | -0.010 | 0.015 | 0.002 | 0.015 | 0.007 | 0.015 |
| rs12926506 | C | T | -0.011 | 0.013 | -0.007 | 0.016 | -0.004 | 0.021 | -0.012 | 0.021 | -0.005 | 0.022 |
| rs11150462 | T | A | 0.008 | 0.010 | 0.014 | 0.012 | 0.016 | 0.015 | 0.019 | 0.016 | -0.007 | 0.016 |
| rs7206608 | C | G | -0.028 | 0.011 | -0.029 | 0.013 | -0.028 | 0.016 | -0.034 | 0.016 | -0.032 | 0.016 |
| rs4790292 | C | A | -0.007 | 0.014 | -0.007 | 0.017 | 0.000 | 0.021 | -0.023 | 0.021 | 0.014 | 0.022 |
| rs58351927 | A | G | 0.004 | 0.010 | 0.003 | 0.013 | -0.002 | 0.016 | 0.014 | 0.016 | -0.002 | 0.016 |
| rs4792716 | A | G | 0.007 | 0.010 | 0.006 | 0.012 | -0.001 | 0.015 | 0.013 | 0.015 | 0.005 | 0.015 |
| rs1320251 | C | T | 0.018 | 0.010 | 0.021 | 0.012 | 0.024 | 0.015 | 0.023 | 0.015 | -0.001 | 0.015 |
| rs1017529 | C | A | 0.009 | 0.014 | 0.007 | 0.017 | 0.002 | 0.021 | 0.005 | 0.022 | -0.004 | 0.022 |
| rs73982435 | C | T | -0.014 | 0.011 | -0.022 | 0.014 | -0.008 | 0.017 | -0.041 | 0.018 | 0.003 | 0.018 |
| rs113962925 | C | T | -0.005 | 0.017 | 0.002 | 0.021 | 0.011 | 0.026 | -0.004 | 0.027 | -0.003 | 0.027 |
| rs11079849 | C | T | -0.009 | 0.010 | -0.008 | 0.012 | -0.019 | 0.015 | 0.007 | 0.016 | -0.013 | 0.016 |
| rs78369934 | T | C | -0.010 | 0.023 | -0.008 | 0.027 | -0.023 | 0.034 | -0.006 | 0.035 | -0.061 | 0.035 |
| rs11150745 | A | G | -0.007 | 0.010 | -0.008 | 0.012 | 0.000 | 0.016 | -0.008 | 0.016 | 0.007 | 0.016 |
| rs2083323 | G | A | -0.007 | 0.013 | -0.003 | 0.015 | -0.030 | 0.019 | 0.033 | 0.020 | 0.000 | 0.020 |
| rs512121 | T | C | -0.006 | 0.012 | 0.006 | 0.014 | -0.001 | 0.018 | 0.019 | 0.019 | -0.008 | 0.019 |

| | | | | | | | | | | | |
|---|---|---|---|---|---|---|---|---|---|---|---|
| rs1788808 | A | G | 0.017 | 0.010 | 0.014 | 0.012 | 0.004 | 0.015 | 0.026 | 0.015 | 0.027 | 0.015 |
| rs16940823 | C | A | 0.006 | 0.013 | 0.018 | 0.016 | 0.028 | 0.020 | 0.012 | 0.020 | 0.009 | 0.020 |
| rs6507054 | T | C | 0.009 | 0.010 | 0.015 | 0.012 | 0.010 | 0.015 | 0.027 | 0.015 | -0.006 | 0.015 |
| rs559231 | G | T | 0.005 | 0.010 | 0.008 | 0.012 | 0.020 | 0.015 | -0.007 | 0.016 | -0.002 | 0.016 |
| rs1834144 | C | A | 0.020 | 0.010 | 0.032 | 0.012 | 0.025 | 0.015 | 0.045 | 0.016 | 0.015 | 0.016 |
| rs7230240 | C | T | -0.008 | 0.011 | -0.006 | 0.013 | -0.011 | 0.016 | 0.007 | 0.017 | 0.004 | 0.017 |
| rs58243949 | C | T | 0.003 | 0.012 | 0.003 | 0.014 | 0.004 | 0.018 | 0.016 | 0.019 | 0.016 | 0.019 |
| rs11659764 | T | A | 0.001 | 0.022 | 0.006 | 0.026 | 0.039 | 0.033 | -0.027 | 0.033 | 0.019 | 0.034 |
| rs1517037 | C | T | 0.005 | 0.012 | -0.001 | 0.015 | -0.008 | 0.019 | 0.009 | 0.019 | -0.004 | 0.019 |
| rs58084604 | C | T | -0.019 | 0.011 | -0.011 | 0.014 | -0.009 | 0.017 | -0.011 | 0.018 | -0.026 | 0.018 |
| rs17773370 | G | A | 0.010 | 0.022 | 0.006 | 0.027 | 0.010 | 0.034 | 0.004 | 0.035 | 0.019 | 0.035 |
| rs57636386 | T | C | 0.018 | 0.017 | 0.012 | 0.021 | 0.004 | 0.026 | 0.022 | 0.027 | 0.031 | 0.027 |
| rs12454712 | T | C | -0.006 | 0.010 | 0.000 | 0.013 | 0.003 | 0.016 | 0.002 | 0.016 | -0.017 | 0.016 |
| rs1373349 | C | T | -0.001 | 0.010 | -0.015 | 0.012 | -0.006 | 0.015 | -0.034 | 0.016 | 0.002 | 0.016 |
| rs8089514 | T | A | -0.024 | 0.010 | -0.024 | 0.012 | -0.028 | 0.015 | -0.021 | 0.016 | -0.023 | 0.016 |
| rs45521740 | G | A | -0.016 | 0.025 | -0.001 | 0.030 | -0.021 | 0.037 | 0.021 | 0.039 | 0.044 | 0.040 |
| rs72976986 | G | A | -0.033 | 0.013 | -0.022 | 0.015 | 0.009 | 0.019 | -0.056 | 0.019 | -0.048 | 0.019 |
| rs75957461 | C | T | -0.023 | 0.021 | -0.043 | 0.024 | -0.052 | 0.031 | -0.037 | 0.032 | 0.006 | 0.032 |
| rs6511826 | G | A | 0.006 | 0.019 | -0.010 | 0.023 | 0.012 | 0.029 | -0.032 | 0.030 | 0.021 | 0.030 |
| rs273505 | T | C | 0.002 | 0.010 | 0.001 | 0.012 | 0.001 | 0.015 | 0.000 | 0.015 | -0.007 | 0.015 |
| rs113230003 | G | A | 0.003 | 0.011 | -0.001 | 0.013 | 0.020 | 0.017 | -0.030 | 0.017 | 0.003 | 0.018 |
| rs10404726 | C | T | 0.023 | 0.010 | 0.027 | 0.012 | 0.039 | 0.015 | 0.016 | 0.015 | 0.039 | 0.015 |
| rs112253053 | T | A | -0.028 | 0.013 | -0.031 | 0.016 | -0.038 | 0.020 | -0.026 | 0.020 | -0.026 | 0.020 |
| rs12462975 | G | A | -0.025 | 0.010 | -0.028 | 0.012 | -0.013 | 0.016 | -0.035 | 0.016 | -0.028 | 0.016 |
| rs73026723 | C | T | 0.010 | 0.014 | 0.014 | 0.017 | 0.013 | 0.021 | 0.016 | 0.022 | 0.009 | 0.022 |
| rs7255223 | C | A | 0.009 | 0.011 | 0.004 | 0.013 | -0.005 | 0.016 | 0.016 | 0.017 | 0.001 | 0.017 |
| rs429358 | T | C | 0.005 | 0.014 | 0.017 | 0.017 | 0.029 | 0.022 | -0.001 | 0.022 | 0.007 | 0.022 |
| rs12971645 | G | A | 0.009 | 0.011 | 0.010 | 0.013 | 0.021 | 0.017 | -0.006 | 0.017 | 0.019 | 0.017 |
| rs10423928 | T | A | -0.005 | 0.012 | -0.002 | 0.014 | -0.002 | 0.018 | 0.001 | 0.018 | -0.007 | 0.019 |
| rs3810291 | G | A | 0.000 | 0.010 | 0.003 | 0.013 | -0.008 | 0.016 | 0.017 | 0.016 | -0.005 | 0.016 |
| rs4545921 | A | G | 0.003 | 0.010 | -0.003 | 0.012 | 0.002 | 0.015 | -0.006 | 0.015 | -0.019 | 0.016 |
| rs61746970 | G | A | 0.022 | 0.029 | 0.028 | 0.034 | 0.011 | 0.043 | 0.042 | 0.045 | 0.033 | 0.045 |
| rs8111074 | G | T | 0.003 | 0.011 | 0.005 | 0.013 | 0.009 | 0.017 | 0.007 | 0.017 | -0.004 | 0.017 |
| rs6075658 | T | C | -0.004 | 0.010 | 0.001 | 0.011 | 0.013 | 0.015 | -0.016 | 0.015 | -0.015 | 0.015 |
| rs2206925 | T | C | 0.049 | 0.010 | 0.031 | 0.012 | 0.024 | 0.015 | 0.037 | 0.016 | 0.061 | 0.016 |
| rs4813224 | T | C | 0.007 | 0.011 | 0.002 | 0.013 | 0.016 | 0.017 | -0.009 | 0.017 | 0.019 | 0.017 |
| rs8124896 | T | C | -0.011 | 0.016 | -0.020 | 0.019 | -0.030 | 0.024 | 0.000 | 0.025 | 0.030 | 0.025 |
| rs6050446 | A | G | -0.004 | 0.032 | -0.017 | 0.038 | -0.053 | 0.048 | 0.030 | 0.049 | 0.032 | 0.049 |
| rs201475383 | G | A | 0.008 | 0.037 | 0.024 | 0.044 | 0.054 | 0.056 | -0.014 | 0.056 | -0.028 | 0.057 |
| rs1987960 | T | C | -0.015 | 0.032 | -0.035 | 0.037 | -0.054 | 0.047 | -0.020 | 0.049 | 0.019 | 0.050 |
| rs4911382 | C | T | 0.011 | 0.010 | 0.000 | 0.012 | -0.012 | 0.015 | 0.005 | 0.015 | 0.028 | 0.015 |
| rs6029180 | A | G | -0.003 | 0.010 | 0.011 | 0.013 | 0.004 | 0.016 | 0.023 | 0.017 | 0.005 | 0.017 |
| rs6030803 | T | C | 0.018 | 0.014 | 0.022 | 0.017 | 0.026 | 0.022 | 0.027 | 0.022 | 0.028 | 0.022 |
| rs2425856 | A | G | 0.002 | 0.010 | -0.003 | 0.012 | 0.001 | 0.015 | -0.008 | 0.015 | 0.012 | 0.015 |
| rs112852122 | G | A | -0.010 | 0.014 | -0.014 | 0.017 | -0.016 | 0.021 | -0.013 | 0.022 | -0.003 | 0.022 |

| | | | | | | | | | | | |
|---|---|---|---|---|---|---|---|---|---|---|---|
| rs66460909 | G | A | 0.024 | 0.013 | 0.025 | 0.015 | 0.032 | 0.019 | 0.017 | 0.019 | 0.020 | 0.020 |
| rs559390 | A | T | -0.008 | 0.010 | -0.002 | 0.012 | 0.005 | 0.015 | -0.012 | 0.016 | -0.014 | 0.016 |
| rs8134638 | T | C | 0.000 | 0.010 | -0.002 | 0.012 | 0.000 | 0.015 | 0.009 | 0.016 | 0.018 | 0.016 |
| rs2837398 | A | C | 0.002 | 0.010 | 0.003 | 0.012 | -0.009 | 0.015 | 0.013 | 0.015 | -0.012 | 0.015 |
| rs1964926 | A | G | -0.005 | 0.010 | -0.003 | 0.012 | -0.003 | 0.015 | 0.005 | 0.016 | -0.014 | 0.016 |
| rs403694 | C | T | 0.003 | 0.010 | 0.003 | 0.012 | 0.004 | 0.015 | 0.004 | 0.015 | 0.012 | 0.015 |
| rs1296685 | A | G | 0.017 | 0.012 | 0.000 | 0.015 | -0.003 | 0.019 | 0.000 | 0.019 | 0.003 | 0.019 |
| rs12484438 | T | C | 0.015 | 0.010 | 0.008 | 0.012 | 0.004 | 0.016 | 0.013 | 0.016 | 0.018 | 0.016 |
| rs738140 | A | G | -0.022 | 0.011 | -0.032 | 0.012 | -0.032 | 0.016 | -0.027 | 0.016 | -0.008 | 0.016 |
| rs9615723 | C | T | -0.019 | 0.010 | -0.031 | 0.012 | -0.016 | 0.015 | -0.042 | 0.016 | -0.008 | 0.016 |
| rs34778589 | A | C | 0.012 | 0.019 | 0.001 | 0.022 | -0.018 | 0.028 | 0.023 | 0.029 | 0.055 | 0.029 |
| Adult body size SNPs (men only) | | | | | | | | | | | |
| SNP | EA | OA | beta_ca | se_ca | beta_co | se_co | beta_prox | se_prox | beta_dist | se_dist | beta_re | se_re |
| rs3762444 | C | T | 0.021 | 0.014 | 0.018 | 0.017 | 0.027 | 0.022 | 0.022 | 0.022 | 0.031 | 0.021 |
| rs1284373 | C | T | 0.008 | 0.016 | 0.000 | 0.020 | -0.014 | 0.026 | 0.013 | 0.025 | -0.012 | 0.024 |
| rs34052145 | A | G | -0.015 | 0.015 | -0.036 | 0.018 | -0.039 | 0.023 | -0.034 | 0.022 | -0.012 | 0.022 |
| rs1167311 | G | A | 0.025 | 0.015 | 0.035 | 0.019 | 0.034 | 0.024 | 0.033 | 0.023 | 0.037 | 0.023 |
| rs12140153 | G | T | 0.000 | 0.026 | 0.012 | 0.032 | -0.008 | 0.041 | 0.027 | 0.040 | -0.034 | 0.038 |
| rs11208779 | G | C | 0.012 | 0.014 | 0.006 | 0.017 | 0.015 | 0.022 | 0.013 | 0.021 | -0.013 | 0.020 |
| rs61765650 | A | G | 0.010 | 0.019 | 0.022 | 0.023 | 0.038 | 0.029 | 0.018 | 0.028 | 0.010 | 0.027 |
| rs34517439 | C | A | -0.008 | 0.024 | -0.004 | 0.029 | 0.014 | 0.037 | -0.030 | 0.036 | -0.018 | 0.035 |
| rs2181375 | A | G | 0.022 | 0.014 | 0.020 | 0.017 | 0.030 | 0.022 | 0.010 | 0.021 | 0.030 | 0.020 |
| rs17024258 | C | T | -0.002 | 0.039 | -0.029 | 0.048 | -0.003 | 0.062 | -0.014 | 0.060 | -0.037 | 0.058 |
| rs984225 | G | A | 0.001 | 0.014 | -0.004 | 0.017 | -0.002 | 0.022 | -0.012 | 0.021 | -0.027 | 0.021 |
| rs61813324 | C | T | -0.037 | 0.024 | -0.036 | 0.029 | -0.020 | 0.037 | -0.066 | 0.036 | -0.026 | 0.036 |
| rs543874 | A | G | 0.013 | 0.017 | -0.001 | 0.022 | -0.025 | 0.028 | 0.008 | 0.027 | 0.031 | 0.026 |
| rs2125232 | C | T | -0.008 | 0.014 | -0.008 | 0.018 | 0.013 | 0.023 | -0.028 | 0.022 | 0.001 | 0.022 |
| rs1529897 | T | G | 0.006 | 0.014 | -0.014 | 0.017 | -0.025 | 0.022 | -0.010 | 0.021 | -0.002 | 0.021 |
| rs935166 | G | A | 0.010 | 0.014 | 0.006 | 0.017 | 0.012 | 0.021 | 0.009 | 0.021 | 0.026 | 0.020 |
| rs1861410 | C | T | -0.009 | 0.014 | -0.013 | 0.017 | -0.017 | 0.021 | -0.016 | 0.021 | 0.000 | 0.020 |
| rs3552 | G | A | 0.015 | 0.014 | 0.023 | 0.017 | 0.008 | 0.022 | 0.031 | 0.021 | 0.001 | 0.020 |
| rs396354 | T | C | 0.001 | 0.015 | -0.012 | 0.019 | -0.003 | 0.024 | -0.022 | 0.024 | 0.010 | 0.023 |
| rs6753397 | C | T | 0.009 | 0.015 | 0.001 | 0.019 | 0.001 | 0.024 | 0.010 | 0.024 | 0.029 | 0.023 |
| rs13405033 | C | T | -0.046 | 0.020 | -0.036 | 0.024 | -0.006 | 0.031 | -0.055 | 0.030 | -0.037 | 0.029 |
| rs1451077 | G | A | -0.003 | 0.014 | 0.004 | 0.017 | -0.014 | 0.022 | 0.021 | 0.021 | -0.023 | 0.021 |
| rs7581907 | A | G | 0.020 | 0.020 | 0.027 | 0.025 | 0.029 | 0.031 | 0.029 | 0.031 | 0.028 | 0.030 |
| rs6436661 | C | T | -0.033 | 0.024 | -0.013 | 0.029 | -0.034 | 0.038 | 0.015 | 0.036 | -0.054 | 0.036 |
| rs7619139 | T | A | 0.005 | 0.014 | 0.005 | 0.017 | -0.019 | 0.022 | 0.023 | 0.021 | 0.001 | 0.021 |
| rs2526389 | C | T | 0.018 | 0.014 | 0.015 | 0.017 | 0.033 | 0.022 | 0.004 | 0.022 | 0.024 | 0.021 |
| rs2336558 | C | T | -0.007 | 0.014 | -0.008 | 0.017 | -0.007 | 0.022 | -0.002 | 0.021 | 0.000 | 0.021 |
| rs11708540 | G | A | -0.036 | 0.020 | -0.046 | 0.024 | -0.064 | 0.030 | -0.014 | 0.030 | -0.037 | 0.029 |
| rs55782528 | C | A | -0.029 | 0.014 | -0.027 | 0.017 | -0.013 | 0.022 | -0.034 | 0.022 | -0.039 | 0.021 |
| rs9861443 | A | C | -0.028 | 0.016 | -0.025 | 0.019 | -0.022 | 0.025 | -0.032 | 0.024 | -0.029 | 0.023 |
| rs13066686 | C | A | 0.006 | 0.014 | 0.000 | 0.017 | -0.002 | 0.022 | -0.003 | 0.022 | 0.016 | 0.021 |
| rs2918217 | C | T | 0.023 | 0.020 | 0.052 | 0.025 | 0.031 | 0.032 | 0.059 | 0.031 | -0.023 | 0.030 |

| | | | | | | | | | | | |
|---|---|---|---|---|---|---|---|---|---|---|---|
| rs73875019 | T | A | -0.032 | 0.022 | -0.045 | 0.026 | -0.034 | 0.034 | -0.057 | 0.033 | -0.055 | 0.032 |
| rs1320903 | G | A | -0.003 | 0.015 | 0.001 | 0.018 | -0.005 | 0.023 | 0.000 | 0.022 | -0.022 | 0.022 |
| rs10935143 | G | A | -0.006 | 0.014 | -0.014 | 0.017 | -0.020 | 0.021 | -0.015 | 0.021 | -0.003 | 0.020 |
| rs61789562 | T | C | 0.023 | 0.022 | 0.020 | 0.026 | 0.037 | 0.034 | -0.011 | 0.033 | 0.021 | 0.032 |
| rs1568488 | G | C | 0.018 | 0.014 | 0.024 | 0.017 | 0.044 | 0.022 | 0.010 | 0.022 | 0.016 | 0.021 |
| rs8192675 | T | C | 0.018 | 0.015 | 0.026 | 0.018 | 0.017 | 0.023 | 0.032 | 0.023 | 0.004 | 0.022 |
| rs894000 | T | C | -0.011 | 0.015 | -0.016 | 0.018 | -0.010 | 0.023 | -0.012 | 0.022 | -0.024 | 0.021 |
| rs55742087 | C | T | -0.029 | 0.018 | -0.022 | 0.022 | -0.043 | 0.028 | -0.001 | 0.028 | 0.000 | 0.027 |
| rs10938398 | G | A | 0.017 | 0.014 | 0.014 | 0.017 | 0.015 | 0.022 | 0.014 | 0.021 | 0.013 | 0.020 |
| rs2537860 | A | C | -0.038 | 0.014 | -0.023 | 0.017 | -0.033 | 0.022 | -0.017 | 0.022 | -0.054 | 0.021 |
| rs13107325 | C | T | -0.009 | 0.026 | -0.003 | 0.032 | -0.012 | 0.040 | -0.009 | 0.039 | 0.000 | 0.039 |
| rs1296328 | A | C | -0.028 | 0.014 | -0.016 | 0.017 | -0.013 | 0.022 | -0.027 | 0.021 | -0.039 | 0.021 |
| rs2307111 | T | C | 0.023 | 0.014 | 0.021 | 0.017 | 0.014 | 0.022 | 0.032 | 0.021 | 0.030 | 0.021 |
| rs7703782 | T | A | -0.009 | 0.019 | -0.010 | 0.024 | -0.002 | 0.031 | -0.026 | 0.030 | 0.013 | 0.029 |
| rs1459843 | C | A | 0.009 | 0.014 | 0.000 | 0.017 | -0.004 | 0.022 | -0.002 | 0.021 | 0.024 | 0.020 |
| rs2591496 | G | A | -0.004 | 0.015 | -0.005 | 0.019 | -0.009 | 0.024 | 0.002 | 0.023 | -0.028 | 0.023 |
| rs9379829 | C | T | 0.033 | 0.016 | 0.049 | 0.020 | 0.055 | 0.025 | 0.052 | 0.025 | 0.044 | 0.024 |
| rs2260051 | A | T | -0.034 | 0.014 | -0.036 | 0.017 | -0.036 | 0.022 | -0.037 | 0.022 | -0.050 | 0.021 |
| rs9277992 | G | A | 0.011 | 0.018 | 0.000 | 0.021 | -0.004 | 0.027 | -0.009 | 0.027 | -0.004 | 0.026 |
| rs9469899 | G | A | -0.011 | 0.014 | -0.001 | 0.017 | 0.035 | 0.022 | -0.025 | 0.022 | 0.045 | 0.021 |
| rs9471333 | C | T | 0.020 | 0.014 | 0.019 | 0.017 | 0.019 | 0.022 | 0.023 | 0.021 | 0.031 | 0.021 |
| rs3798519 | A | C | 0.005 | 0.017 | 0.006 | 0.021 | 0.018 | 0.028 | -0.006 | 0.027 | 0.005 | 0.026 |
| rs74621225 | A | G | -0.023 | 0.021 | -0.018 | 0.025 | -0.007 | 0.032 | -0.035 | 0.031 | -0.019 | 0.030 |
| rs9320823 | T | C | -0.037 | 0.014 | -0.032 | 0.017 | 0.005 | 0.022 | -0.065 | 0.022 | -0.032 | 0.021 |
| rs9478671 | A | G | 0.021 | 0.017 | 0.014 | 0.020 | 0.036 | 0.026 | 0.004 | 0.026 | 0.027 | 0.025 |
| rs34714518 | G | A | 0.025 | 0.021 | 0.042 | 0.026 | 0.048 | 0.034 | 0.046 | 0.033 | 0.038 | 0.031 |
| rs12666574 | G | A | -0.036 | 0.014 | -0.041 | 0.018 | -0.030 | 0.022 | -0.055 | 0.022 | -0.058 | 0.021 |
| rs62457529 | A | G | 0.018 | 0.023 | 0.012 | 0.028 | -0.017 | 0.035 | 0.038 | 0.035 | 0.041 | 0.034 |
| rs6962280 | A | G | -0.032 | 0.014 | -0.019 | 0.017 | 0.006 | 0.022 | -0.047 | 0.021 | -0.030 | 0.020 |
| rs17145600 | C | T | 0.001 | 0.038 | 0.006 | 0.047 | -0.018 | 0.060 | 0.005 | 0.058 | 0.041 | 0.057 |
| rs12538826 | T | C | 0.000 | 0.021 | 0.001 | 0.026 | -0.007 | 0.033 | 0.000 | 0.033 | -0.005 | 0.032 |
| rs10236214 | C | T | -0.011 | 0.015 | -0.021 | 0.018 | -0.036 | 0.023 | -0.004 | 0.022 | -0.010 | 0.021 |
| rs4840941 | A | G | -0.008 | 0.015 | -0.015 | 0.018 | -0.025 | 0.023 | -0.011 | 0.022 | -0.002 | 0.022 |
| rs9329197 | A | T | 0.009 | 0.015 | 0.009 | 0.019 | 0.011 | 0.024 | 0.003 | 0.023 | 0.003 | 0.023 |
| rs3750310 | G | A | 0.009 | 0.015 | 0.012 | 0.019 | 0.001 | 0.024 | 0.018 | 0.023 | 0.009 | 0.023 |
| rs11780420 | G | A | 0.006 | 0.015 | 0.001 | 0.019 | 0.011 | 0.024 | -0.013 | 0.023 | -0.001 | 0.023 |
| rs7827210 | G | A | -0.022 | 0.014 | -0.031 | 0.017 | -0.026 | 0.022 | -0.038 | 0.022 | -0.001 | 0.021 |
| rs35732620 | G | T | -0.020 | 0.014 | -0.022 | 0.017 | -0.008 | 0.022 | -0.040 | 0.021 | -0.014 | 0.021 |
| rs1812736 | G | A | -0.014 | 0.018 | -0.003 | 0.022 | 0.009 | 0.028 | -0.026 | 0.028 | -0.028 | 0.027 |
| rs72674843 | T | C | 0.017 | 0.015 | 0.010 | 0.019 | -0.017 | 0.024 | 0.030 | 0.024 | 0.002 | 0.023 |
| rs800526 | A | C | -0.019 | 0.016 | -0.043 | 0.021 | 0.014 | 0.026 | -0.089 | 0.026 | 0.015 | 0.025 |
| rs1412239 | C | G | -0.024 | 0.015 | -0.015 | 0.018 | -0.023 | 0.023 | -0.026 | 0.023 | -0.017 | 0.022 |
| rs10828247 | A | G | -0.030 | 0.015 | -0.029 | 0.018 | -0.028 | 0.023 | -0.036 | 0.022 | -0.041 | 0.021 |
| rs10824218 | A | T | 0.000 | 0.014 | -0.006 | 0.017 | 0.001 | 0.022 | -0.017 | 0.021 | 0.018 | 0.021 |
| rs10883026 | C | T | 0.011 | 0.014 | -0.008 | 0.017 | 0.001 | 0.022 | -0.013 | 0.021 | 0.027 | 0.021 |

| | | | | | | | | | | | |
|---|---|---|---|---|---|---|---|---|---|---|---|
| rs117597828 | C | T | -0.017 | 0.016 | -0.011 | 0.020 | -0.013 | 0.026 | -0.005 | 0.025 | -0.014 | 0.025 |
| rs4962671 | T | C | 0.009 | 0.014 | 0.002 | 0.017 | -0.007 | 0.022 | 0.019 | 0.021 | 0.025 | 0.021 |
| rs72867447 | C | G | 0.018 | 0.014 | 0.005 | 0.017 | -0.002 | 0.022 | -0.002 | 0.021 | 0.050 | 0.020 |
| rs6265 | C | T | 0.017 | 0.017 | 0.011 | 0.021 | 0.015 | 0.027 | 0.011 | 0.026 | 0.025 | 0.025 |
| rs1222216 | C | T | 0.002 | 0.017 | 0.001 | 0.021 | -0.022 | 0.026 | 0.025 | 0.026 | 0.013 | 0.025 |
| rs4755726 | T | G | 0.001 | 0.015 | 0.012 | 0.018 | 0.007 | 0.023 | 0.020 | 0.023 | -0.040 | 0.022 |
| rs12798028 | C | T | -0.017 | 0.014 | -0.009 | 0.017 | -0.013 | 0.022 | -0.013 | 0.021 | -0.013 | 0.021 |
| rs7940691 | C | T | 0.025 | 0.014 | 0.035 | 0.018 | 0.026 | 0.022 | 0.050 | 0.022 | 0.028 | 0.021 |
| rs10898317 | C | T | 0.002 | 0.014 | 0.003 | 0.017 | 0.005 | 0.022 | -0.013 | 0.021 | -0.017 | 0.021 |
| rs55726687 | G | A | -0.012 | 0.017 | -0.023 | 0.021 | -0.017 | 0.027 | -0.028 | 0.026 | -0.021 | 0.026 |
| rs76895963 | T | G | -0.232 | 0.056 | -0.252 | 0.068 | -0.259 | 0.087 | -0.210 | 0.085 | -0.242 | 0.083 |
| rs7132908 | G | A | 0.001 | 0.014 | 0.005 | 0.017 | 0.006 | 0.022 | 0.004 | 0.022 | 0.018 | 0.021 |
| rs4759228 | G | C | 0.015 | 0.016 | 0.024 | 0.020 | 0.024 | 0.025 | 0.036 | 0.025 | 0.006 | 0.024 |
| rs7308188 | T | C | 0.008 | 0.015 | 0.011 | 0.019 | -0.010 | 0.024 | 0.021 | 0.024 | -0.002 | 0.023 |
| rs6490030 | C | A | 0.028 | 0.014 | 0.031 | 0.017 | 0.029 | 0.022 | 0.034 | 0.022 | 0.029 | 0.021 |
| rs147730268 | G | T | 0.024 | 0.028 | 0.016 | 0.034 | 0.026 | 0.044 | -0.001 | 0.043 | 0.021 | 0.042 |
| rs11619722 | T | C | 0.003 | 0.015 | 0.000 | 0.019 | 0.004 | 0.024 | -0.006 | 0.023 | -0.012 | 0.023 |
| rs61954177 | G | C | 0.000 | 0.015 | 0.016 | 0.018 | 0.019 | 0.023 | 0.013 | 0.023 | 0.004 | 0.022 |
| rs4477562 | C | T | 0.011 | 0.020 | 0.042 | 0.025 | 0.066 | 0.033 | 0.017 | 0.031 | -0.007 | 0.030 |
| rs9317002 | C | A | -0.009 | 0.014 | -0.010 | 0.017 | -0.010 | 0.021 | -0.020 | 0.021 | -0.016 | 0.020 |
| rs7983454 | T | C | 0.001 | 0.014 | 0.010 | 0.017 | 0.013 | 0.021 | 0.005 | 0.021 | -0.005 | 0.020 |
| rs9522279 | C | T | -0.019 | 0.014 | -0.029 | 0.017 | -0.029 | 0.022 | -0.039 | 0.021 | -0.008 | 0.021 |
| rs10132280 | C | A | -0.012 | 0.015 | -0.031 | 0.021 | -0.034 | 0.027 | -0.020 | 0.027 | 0.003 | 0.027 |
| rs9788550 | G | C | 0.008 | 0.016 | 0.045 | 0.023 | 0.033 | 0.029 | 0.058 | 0.029 | -0.018 | 0.029 |
| rs2143975 | C | G | 0.021 | 0.014 | 0.033 | 0.020 | 0.042 | 0.025 | 0.025 | 0.025 | 0.037 | 0.025 |
| rs10131761 | T | A | 0.005 | 0.018 | 0.037 | 0.025 | 0.034 | 0.033 | 0.016 | 0.033 | -0.023 | 0.033 |
| rs4898556 | A | C | -0.005 | 0.014 | -0.002 | 0.020 | 0.005 | 0.025 | -0.011 | 0.025 | -0.039 | 0.025 |
| rs217669 | T | C | 0.012 | 0.015 | 0.010 | 0.023 | 0.060 | 0.029 | -0.021 | 0.025 | 0.018 | 0.029 |
| rs8008910 | G | A | 0.001 | 0.017 | 0.007 | 0.023 | 0.039 | 0.032 | -0.022 | 0.031 | -0.006 | 0.029 |
| rs8008772 | A | T | 0.003 | 0.016 | 0.016 | 0.024 | 0.014 | 0.030 | 0.018 | 0.030 | 0.001 | 0.030 |
| rs1887197 | C | T | 0.002 | 0.014 | 0.008 | 0.019 | -0.019 | 0.025 | 0.001 | 0.026 | 0.017 | 0.023 |
| rs61992671 | A | G | 0.016 | 0.015 | 0.026 | 0.022 | 0.022 | 0.027 | 0.021 | 0.027 | 0.029 | 0.027 |
| rs11631651 | A | C | 0.013 | 0.027 | 0.023 | 0.033 | 0.060 | 0.042 | -0.004 | 0.041 | -0.027 | 0.039 |
| rs57488047 | T | C | 0.021 | 0.014 | 0.031 | 0.017 | 0.028 | 0.022 | 0.033 | 0.021 | 0.006 | 0.020 |
| rs2238435 | C | G | -0.010 | 0.014 | -0.004 | 0.017 | -0.014 | 0.022 | -0.002 | 0.021 | -0.015 | 0.021 |
| rs1990573 | A | G | 0.022 | 0.015 | 0.025 | 0.018 | 0.026 | 0.023 | 0.029 | 0.023 | 0.040 | 0.022 |
| rs8054079 | C | T | -0.005 | 0.020 | -0.001 | 0.025 | 0.023 | 0.032 | -0.021 | 0.031 | 0.002 | 0.030 |
| rs27741 | G | A | 0.002 | 0.014 | -0.006 | 0.017 | 0.003 | 0.022 | -0.017 | 0.022 | 0.007 | 0.021 |
| rs62048402 | G | A | 0.000 | 0.014 | 0.002 | 0.017 | -0.005 | 0.022 | 0.009 | 0.021 | 0.003 | 0.020 |
| rs12923231 | C | T | 0.016 | 0.014 | 0.012 | 0.017 | 0.015 | 0.022 | 0.011 | 0.021 | 0.009 | 0.021 |
| rs12149660 | G | A | -0.014 | 0.023 | 0.009 | 0.028 | 0.027 | 0.037 | -0.007 | 0.036 | -0.002 | 0.035 |
| rs3923783 | C | A | 0.029 | 0.018 | 0.029 | 0.022 | 0.032 | 0.028 | 0.018 | 0.028 | 0.052 | 0.027 |
| rs9901404 | A | G | 0.017 | 0.016 | 0.015 | 0.019 | 0.035 | 0.025 | 0.013 | 0.024 | 0.005 | 0.023 |
| rs12941009 | C | T | 0.001 | 0.014 | -0.004 | 0.017 | 0.004 | 0.022 | -0.015 | 0.022 | 0.021 | 0.021 |
| rs11079849 | C | T | -0.003 | 0.015 | 0.002 | 0.018 | -0.014 | 0.023 | 0.021 | 0.022 | -0.007 | 0.022 |

| | | | | | | | | | | | |
|---|---|---|---|---|---|---|---|---|---|---|---|
| rs11150745 | A | G | 0.011 | 0.015 | 0.006 | 0.018 | 0.009 | 0.023 | 0.018 | 0.023 | 0.030 | 0.022 |
| rs1652376 | G | T | 0.017 | 0.014 | 0.016 | 0.017 | -0.001 | 0.022 | 0.024 | 0.021 | 0.029 | 0.021 |
| rs7232171 | G | T | 0.013 | 0.014 | 0.013 | 0.017 | 0.014 | 0.022 | 0.016 | 0.021 | -0.013 | 0.020 |
| rs58243949 | C | T | -0.002 | 0.017 | 0.002 | 0.021 | 0.012 | 0.027 | 0.003 | 0.026 | 0.013 | 0.025 |
| rs7240682 | C | G | -0.019 | 0.016 | -0.020 | 0.020 | -0.022 | 0.025 | -0.012 | 0.025 | -0.016 | 0.024 |
| rs8112818 | A | G | 0.035 | 0.014 | 0.051 | 0.017 | 0.065 | 0.022 | 0.036 | 0.022 | 0.020 | 0.021 |
| rs10423928 | T | A | 0.006 | 0.017 | -0.006 | 0.020 | -0.012 | 0.026 | 0.001 | 0.026 | 0.007 | 0.025 |
| rs3810291 | G | A | 0.020 | 0.015 | 0.028 | 0.018 | 0.015 | 0.023 | 0.048 | 0.023 | 0.008 | 0.022 |
| rs6054427 | G | A | 0.025 | 0.014 | 0.015 | 0.017 | -0.002 | 0.022 | 0.027 | 0.022 | 0.032 | 0.021 |
| rs6096886 | A | G | 0.028 | 0.017 | 0.025 | 0.021 | 0.028 | 0.027 | 0.019 | 0.027 | 0.026 | 0.026 |
| rs9977825 | T | C | -0.015 | 0.014 | -0.032 | 0.018 | -0.034 | 0.023 | -0.025 | 0.022 | -0.017 | 0.021 |
| rs17421586 | T | A | 0.007 | 0.014 | -0.006 | 0.018 | -0.019 | 0.023 | -0.003 | 0.022 | 0.003 | 0.022 |
| Adult body size SNPs (women only) | | | | | | | | | | | |
| SNP | EA | OA | beta_ca | se_ca | beta_co | se_co | beta_prox | se_prox | beta_dist | se_dist | beta_re | se_re |
| rs74892851 | C | A | 0.008 | 0.017 | 0.020 | 0.021 | 0.042 | 0.025 | -0.017 | 0.028 | 0.004 | 0.028 |
| rs78886584 | A | G | -0.024 | 0.016 | -0.006 | 0.019 | -0.007 | 0.023 | -0.009 | 0.025 | -0.060 | 0.027 |
| rs72660086 | T | G | 0.005 | 0.017 | 0.028 | 0.020 | 0.027 | 0.024 | 0.037 | 0.027 | -0.018 | 0.028 |
| rs12144626 | T | C | -0.013 | 0.014 | -0.009 | 0.017 | -0.009 | 0.020 | -0.010 | 0.022 | -0.066 | 0.023 |
| rs1494461 | C | T | -0.002 | 0.015 | -0.012 | 0.017 | -0.011 | 0.021 | -0.015 | 0.024 | -0.005 | 0.025 |
| rs12140153 | G | T | 0.028 | 0.027 | 0.038 | 0.031 | 0.080 | 0.039 | -0.038 | 0.042 | -0.026 | 0.044 |
| rs2815757 | C | T | -0.016 | 0.018 | -0.022 | 0.021 | 0.005 | 0.026 | -0.055 | 0.029 | 0.005 | 0.030 |
| rs1514173 | C | T | 0.009 | 0.014 | 0.010 | 0.016 | 0.007 | 0.020 | 0.002 | 0.022 | -0.010 | 0.023 |
| rs34517439 | C | A | -0.007 | 0.024 | -0.028 | 0.028 | -0.035 | 0.034 | -0.019 | 0.038 | -0.015 | 0.039 |
| rs10922911 | C | T | 0.009 | 0.014 | 0.006 | 0.017 | -0.008 | 0.020 | 0.017 | 0.022 | 0.030 | 0.023 |
| rs653958 | A | G | -0.025 | 0.014 | -0.027 | 0.017 | -0.013 | 0.021 | -0.035 | 0.023 | -0.019 | 0.025 |
| rs75641275 | A | C | 0.025 | 0.020 | 0.015 | 0.024 | 0.031 | 0.029 | 0.007 | 0.032 | 0.029 | 0.033 |
| rs41279738 | T | G | 0.002 | 0.042 | 0.002 | 0.049 | -0.015 | 0.060 | -0.020 | 0.066 | 0.029 | 0.069 |
| rs12033257 | A | G | 0.017 | 0.014 | 0.007 | 0.017 | 0.025 | 0.021 | -0.013 | 0.023 | 0.039 | 0.024 |
| rs3753639 | T | C | -0.004 | 0.016 | -0.008 | 0.019 | 0.000 | 0.023 | -0.025 | 0.026 | -0.041 | 0.027 |
| rs61813324 | C | T | -0.063 | 0.024 | -0.055 | 0.028 | -0.058 | 0.034 | -0.058 | 0.038 | -0.076 | 0.040 |
| rs539515 | A | C | -0.017 | 0.017 | 0.003 | 0.021 | 0.007 | 0.025 | 0.005 | 0.028 | -0.039 | 0.029 |
| rs815163 | T | C | 0.001 | 0.014 | -0.008 | 0.016 | -0.001 | 0.020 | -0.022 | 0.022 | 0.016 | 0.023 |
| rs2678204 | T | G | -0.027 | 0.015 | -0.037 | 0.017 | -0.043 | 0.021 | -0.033 | 0.024 | -0.003 | 0.025 |
| rs2994320 | A | G | -0.019 | 0.017 | -0.042 | 0.020 | -0.013 | 0.025 | -0.083 | 0.027 | 0.002 | 0.029 |
| rs62106258 | T | C | 0.041 | 0.036 | 0.060 | 0.042 | 0.027 | 0.051 | 0.121 | 0.058 | 0.000 | 0.059 |
| rs6548237 | A | C | -0.004 | 0.018 | -0.007 | 0.021 | -0.033 | 0.026 | 0.027 | 0.028 | 0.006 | 0.030 |
| rs6749422 | C | G | -0.009 | 0.014 | -0.003 | 0.016 | -0.006 | 0.020 | -0.006 | 0.022 | -0.027 | 0.023 |
| rs34606703 | G | A | -0.009 | 0.014 | -0.016 | 0.017 | -0.028 | 0.021 | -0.009 | 0.023 | -0.022 | 0.024 |
| rs13420048 | C | A | 0.021 | 0.015 | 0.022 | 0.017 | 0.033 | 0.021 | 0.012 | 0.023 | 0.033 | 0.024 |
| rs6545468 | C | G | 0.001 | 0.014 | 0.004 | 0.016 | -0.014 | 0.020 | 0.017 | 0.022 | 0.008 | 0.023 |
| rs4671328 | T | G | -0.002 | 0.014 | -0.001 | 0.016 | 0.002 | 0.020 | 0.000 | 0.022 | 0.024 | 0.023 |
| rs13416992 | A | C | 0.006 | 0.014 | 0.007 | 0.017 | -0.003 | 0.020 | 0.020 | 0.022 | 0.027 | 0.023 |
| rs10192894 | A | G | -0.014 | 0.014 | -0.026 | 0.016 | -0.036 | 0.020 | -0.017 | 0.022 | -0.025 | 0.023 |
| rs12477088 | T | C | -0.004 | 0.014 | -0.014 | 0.016 | -0.032 | 0.020 | 0.011 | 0.022 | -0.025 | 0.023 |
| rs11691869 | C | A | 0.015 | 0.014 | 0.018 | 0.017 | -0.008 | 0.021 | 0.043 | 0.023 | 0.019 | 0.024 |

| | | | | | | | | | | | |
|---|---|---|---|---|---|---|---|---|---|---|---|
| rs7602120 | C | T | -0.016 | 0.014 | -0.016 | 0.016 | -0.015 | 0.020 | -0.021 | 0.022 | -0.012 | 0.023 |
| rs1083472 | C | G | 0.028 | 0.014 | 0.030 | 0.016 | 0.034 | 0.020 | 0.023 | 0.022 | 0.034 | 0.023 |
| rs4482463 | C | A | -0.024 | 0.024 | -0.021 | 0.029 | -0.030 | 0.036 | -0.018 | 0.040 | -0.082 | 0.042 |
| rs4673553 | T | G | 0.004 | 0.014 | 0.004 | 0.016 | 0.004 | 0.020 | 0.013 | 0.022 | 0.010 | 0.023 |
| rs2433733 | G | A | -0.009 | 0.014 | -0.007 | 0.017 | -0.005 | 0.021 | -0.009 | 0.023 | -0.014 | 0.024 |
| rs113706999 | T | A | -0.047 | 0.056 | -0.028 | 0.065 | -0.114 | 0.078 | 0.065 | 0.091 | -0.219 | 0.089 |
| rs72906474 | G | T | 0.015 | 0.015 | 0.019 | 0.017 | 0.011 | 0.021 | 0.030 | 0.024 | 0.027 | 0.025 |
| rs9843653 | T | C | -0.015 | 0.014 | -0.010 | 0.016 | -0.005 | 0.020 | -0.011 | 0.022 | -0.046 | 0.023 |
| rs6774533 | C | T | -0.001 | 0.015 | -0.006 | 0.018 | -0.028 | 0.022 | 0.029 | 0.025 | -0.007 | 0.026 |
| rs13066308 | C | G | -0.010 | 0.014 | -0.015 | 0.017 | -0.002 | 0.021 | -0.026 | 0.023 | -0.013 | 0.024 |
| rs1454687 | C | G | -0.009 | 0.014 | -0.008 | 0.016 | 0.006 | 0.020 | -0.019 | 0.022 | 0.001 | 0.023 |
| rs13081671 | C | T | -0.001 | 0.016 | -0.012 | 0.018 | -0.029 | 0.022 | 0.004 | 0.025 | -0.007 | 0.026 |
| rs2035936 | G | T | -0.033 | 0.029 | -0.041 | 0.037 | -0.089 | 0.044 | 0.028 | 0.051 | -0.123 | 0.051 |
| rs529200 | A | G | 0.013 | 0.014 | 0.012 | 0.016 | -0.009 | 0.020 | 0.037 | 0.022 | -0.003 | 0.023 |
| rs73052033 | T | C | -0.011 | 0.018 | -0.017 | 0.021 | -0.014 | 0.026 | -0.030 | 0.028 | -0.008 | 0.030 |
| rs61218008 | A | G | 0.022 | 0.015 | 0.016 | 0.018 | 0.016 | 0.022 | 0.007 | 0.024 | 0.001 | 0.025 |
| rs2643450 | A | G | -0.007 | 0.014 | 0.010 | 0.017 | 0.009 | 0.020 | 0.011 | 0.022 | -0.006 | 0.023 |
| rs9684942 | G | A | 0.002 | 0.020 | 0.003 | 0.023 | 0.014 | 0.028 | -0.007 | 0.031 | 0.012 | 0.033 |
| rs34811474 | G | A | 0.035 | 0.019 | 0.056 | 0.022 | 0.058 | 0.027 | 0.063 | 0.030 | 0.023 | 0.032 |
| rs73213484 | A | T | 0.014 | 0.019 | 0.022 | 0.022 | 0.015 | 0.027 | 0.030 | 0.030 | 0.040 | 0.032 |
| rs4527444 | A | G | -0.001 | 0.014 | -0.006 | 0.016 | 0.011 | 0.020 | -0.032 | 0.022 | 0.000 | 0.023 |
| rs12641981 | C | T | -0.019 | 0.014 | -0.025 | 0.016 | -0.027 | 0.020 | -0.013 | 0.022 | -0.011 | 0.023 |
| rs148712344 | G | T | -0.068 | 0.037 | -0.044 | 0.043 | -0.047 | 0.052 | -0.026 | 0.059 | -0.082 | 0.060 |
| rs925422 | T | G | 0.010 | 0.015 | 0.012 | 0.018 | 0.006 | 0.023 | 0.020 | 0.025 | 0.008 | 0.026 |
| rs1603179 | A | C | 0.014 | 0.014 | 0.020 | 0.017 | 0.040 | 0.020 | -0.006 | 0.022 | 0.028 | 0.024 |
| rs11098965 | C | T | -0.012 | 0.016 | -0.013 | 0.019 | -0.004 | 0.023 | -0.027 | 0.025 | -0.009 | 0.026 |
| rs4148155 | A | G | -0.015 | 0.022 | 0.000 | 0.026 | 0.034 | 0.032 | -0.033 | 0.035 | -0.014 | 0.037 |
| rs13107325 | C | T | 0.028 | 0.027 | 0.026 | 0.031 | -0.041 | 0.037 | 0.101 | 0.043 | -0.021 | 0.044 |
| rs769668 | T | C | -0.005 | 0.015 | -0.007 | 0.017 | -0.007 | 0.021 | -0.001 | 0.023 | -0.004 | 0.024 |
| rs35390852 | G | A | -0.021 | 0.021 | -0.019 | 0.025 | -0.009 | 0.031 | -0.024 | 0.034 | -0.075 | 0.035 |
| rs828550 | C | T | 0.012 | 0.015 | 0.019 | 0.017 | 0.026 | 0.021 | 0.006 | 0.023 | 0.001 | 0.024 |
| rs10514963 | G | A | 0.032 | 0.014 | 0.029 | 0.016 | 0.027 | 0.020 | 0.040 | 0.022 | 0.033 | 0.023 |
| rs34341 | A | T | 0.008 | 0.014 | 0.012 | 0.017 | 0.013 | 0.020 | 0.017 | 0.022 | 0.032 | 0.023 |
| rs59893724 | A | G | -0.013 | 0.016 | -0.013 | 0.019 | 0.001 | 0.024 | -0.042 | 0.026 | -0.050 | 0.027 |
| rs7442885 | C | G | -0.050 | 0.016 | -0.045 | 0.019 | -0.035 | 0.023 | -0.064 | 0.026 | -0.041 | 0.027 |
| rs1477290 | T | C | 0.010 | 0.019 | -0.015 | 0.023 | -0.004 | 0.028 | -0.023 | 0.031 | 0.019 | 0.032 |
| rs10038055 | G | T | -0.008 | 0.014 | -0.010 | 0.017 | -0.011 | 0.020 | -0.010 | 0.023 | 0.012 | 0.024 |
| rs288187 | C | T | 0.002 | 0.018 | 0.001 | 0.022 | 0.014 | 0.026 | -0.007 | 0.029 | 0.023 | 0.031 |
| rs1366334 | C | G | -0.003 | 0.015 | -0.028 | 0.018 | -0.041 | 0.022 | -0.003 | 0.024 | 0.007 | 0.025 |
| rs13174863 | A | G | -0.033 | 0.019 | -0.028 | 0.022 | -0.017 | 0.027 | -0.033 | 0.030 | -0.018 | 0.031 |
| rs251353 | C | A | 0.028 | 0.014 | 0.020 | 0.017 | 0.015 | 0.021 | 0.014 | 0.023 | 0.012 | 0.024 |
| rs11134679 | A | G | 0.007 | 0.015 | 0.017 | 0.017 | 0.021 | 0.021 | 0.016 | 0.023 | 0.004 | 0.024 |
| rs9395520 | C | T | -0.001 | 0.015 | -0.009 | 0.018 | 0.000 | 0.021 | -0.018 | 0.024 | 0.021 | 0.025 |
| rs35778344 | G | A | -0.037 | 0.018 | -0.025 | 0.022 | -0.055 | 0.026 | 0.004 | 0.029 | -0.069 | 0.030 |
| rs3130048 | T | C | -0.020 | 0.016 | -0.016 | 0.018 | -0.030 | 0.022 | 0.003 | 0.025 | -0.063 | 0.026 |

| | | | | | | | | | | | |
|---|---|---|---|---|---|---|---|---|---|---|---|
| rs34298980 | T | C | -0.006 | 0.015 | -0.019 | 0.017 | -0.035 | 0.021 | 0.003 | 0.023 | -0.003 | 0.025 |
| rs72892910 | G | T | 0.017 | 0.018 | 0.006 | 0.021 | 0.019 | 0.026 | -0.006 | 0.029 | 0.059 | 0.030 |
| rs1547026 | T | C | 0.011 | 0.015 | 0.028 | 0.018 | 0.016 | 0.022 | 0.046 | 0.025 | 0.004 | 0.026 |
| rs2253310 | C | G | 0.001 | 0.014 | -0.011 | 0.017 | -0.016 | 0.020 | -0.016 | 0.022 | 0.000 | 0.023 |
| rs9387640 | C | T | -0.004 | 0.014 | -0.001 | 0.017 | -0.015 | 0.020 | 0.032 | 0.023 | -0.004 | 0.024 |
| rs73046311 | C | G | -0.021 | 0.018 | -0.020 | 0.022 | -0.006 | 0.026 | -0.024 | 0.029 | -0.023 | 0.031 |
| rs2866720 | C | T | 0.020 | 0.014 | 0.012 | 0.017 | 0.023 | 0.021 | -0.009 | 0.023 | 0.013 | 0.024 |
| rs11976018 | G | A | 0.033 | 0.018 | 0.019 | 0.022 | 0.011 | 0.027 | 0.019 | 0.030 | 0.031 | 0.032 |
| rs12375196 | C | A | 0.008 | 0.014 | 0.009 | 0.016 | 0.005 | 0.020 | 0.018 | 0.022 | -0.021 | 0.023 |
| rs2396625 | T | A | 0.010 | 0.014 | 0.012 | 0.017 | 0.018 | 0.020 | 0.009 | 0.022 | -0.016 | 0.023 |
| rs1840661 | T | A | -0.001 | 0.014 | 0.005 | 0.017 | 0.018 | 0.020 | -0.009 | 0.022 | -0.030 | 0.023 |
| rs7853 | A | G | 0.009 | 0.015 | 0.046 | 0.018 | 0.069 | 0.022 | 0.023 | 0.024 | -0.035 | 0.025 |
| rs11250094 | G | C | 0.006 | 0.015 | -0.036 | 0.018 | -0.061 | 0.022 | 0.006 | 0.024 | 0.034 | 0.025 |
| rs6557829 | C | A | -0.003 | 0.014 | -0.006 | 0.017 | 0.006 | 0.020 | -0.023 | 0.023 | 0.010 | 0.024 |
| rs117176448 | C | G | 0.038 | 0.024 | 0.048 | 0.028 | 0.042 | 0.034 | 0.047 | 0.038 | 0.043 | 0.039 |
| rs10957605 | C | T | -0.023 | 0.015 | -0.029 | 0.018 | -0.030 | 0.022 | -0.030 | 0.024 | -0.026 | 0.025 |
| rs17716502 | C | T | 0.001 | 0.018 | 0.008 | 0.021 | 0.007 | 0.025 | 0.022 | 0.028 | 0.013 | 0.029 |
| rs4740442 | C | T | -0.013 | 0.015 | 0.006 | 0.018 | 0.021 | 0.022 | -0.006 | 0.024 | -0.038 | 0.025 |
| rs13292699 | A | C | 0.015 | 0.014 | 0.018 | 0.016 | 0.040 | 0.020 | -0.013 | 0.022 | 0.012 | 0.023 |
| rs17770336 | C | T | -0.029 | 0.015 | -0.025 | 0.018 | -0.043 | 0.021 | -0.014 | 0.024 | -0.013 | 0.025 |
| rs2398851 | A | G | -0.008 | 0.014 | -0.015 | 0.017 | 0.010 | 0.021 | -0.050 | 0.023 | 0.021 | 0.024 |
| rs7047694 | G | A | -0.021 | 0.015 | -0.018 | 0.017 | -0.017 | 0.021 | -0.018 | 0.023 | -0.049 | 0.024 |
| rs6478538 | A | G | -0.025 | 0.015 | -0.023 | 0.017 | -0.005 | 0.021 | -0.053 | 0.023 | -0.046 | 0.024 |
| rs777676 | T | A | -0.018 | 0.014 | -0.026 | 0.016 | -0.020 | 0.020 | -0.038 | 0.022 | -0.018 | 0.023 |
| rs3003578 | C | T | 0.015 | 0.014 | 0.005 | 0.016 | 0.013 | 0.020 | -0.009 | 0.022 | 0.054 | 0.023 |
| rs1270799 | T | G | -0.024 | 0.015 | -0.020 | 0.018 | -0.024 | 0.022 | -0.002 | 0.024 | -0.050 | 0.025 |
| rs113585475 | C | T | -0.016 | 0.026 | -0.046 | 0.030 | -0.013 | 0.037 | -0.073 | 0.040 | 0.038 | 0.044 |
| rs3125326 | A | C | 0.007 | 0.014 | 0.003 | 0.017 | -0.019 | 0.021 | 0.026 | 0.023 | -0.010 | 0.024 |
| rs7090758 | T | C | 0.016 | 0.014 | 0.017 | 0.016 | 0.020 | 0.020 | 0.014 | 0.022 | 0.010 | 0.023 |
| rs11000942 | G | A | 0.012 | 0.021 | 0.034 | 0.025 | 0.034 | 0.031 | 0.025 | 0.034 | -0.038 | 0.035 |
| rs1250535 | C | G | -0.028 | 0.015 | -0.034 | 0.017 | -0.044 | 0.021 | -0.025 | 0.023 | -0.002 | 0.024 |
| rs17399739 | A | G | 0.016 | 0.027 | 0.045 | 0.032 | 0.083 | 0.040 | 0.009 | 0.043 | 0.034 | 0.045 |
| rs10510025 | C | T | 0.015 | 0.016 | 0.014 | 0.019 | 0.002 | 0.023 | 0.023 | 0.025 | 0.009 | 0.026 |
| rs4962725 | T | C | -0.010 | 0.014 | -0.011 | 0.016 | 0.000 | 0.020 | -0.027 | 0.022 | 0.000 | 0.023 |
| rs11146233 | G | A | 0.000 | 0.014 | -0.004 | 0.016 | 0.004 | 0.020 | -0.014 | 0.022 | 0.000 | 0.023 |
| rs7950166 | C | T | -0.007 | 0.014 | -0.020 | 0.017 | -0.040 | 0.020 | -0.002 | 0.022 | 0.001 | 0.024 |
| rs11022766 | T | G | -0.013 | 0.014 | -0.009 | 0.017 | -0.015 | 0.020 | -0.003 | 0.023 | -0.023 | 0.024 |
| rs1013402 | A | G | 0.006 | 0.015 | -0.010 | 0.018 | -0.012 | 0.022 | -0.012 | 0.024 | 0.025 | 0.025 |
| rs34292685 | C | T | 0.028 | 0.019 | 0.032 | 0.022 | 0.010 | 0.027 | 0.044 | 0.030 | 0.032 | 0.032 |
| rs10896012 | T | C | 0.017 | 0.017 | 0.029 | 0.020 | 0.031 | 0.025 | 0.040 | 0.027 | 0.025 | 0.028 |
| rs3802858 | T | C | -0.033 | 0.014 | -0.031 | 0.016 | -0.032 | 0.020 | -0.012 | 0.022 | -0.025 | 0.023 |
| rs11218510 | G | A | 0.008 | 0.014 | -0.001 | 0.016 | 0.005 | 0.020 | -0.016 | 0.022 | 0.062 | 0.023 |
| rs2512884 | C | A | 0.002 | 0.014 | -0.019 | 0.016 | -0.022 | 0.020 | -0.028 | 0.022 | 0.035 | 0.023 |
| rs11223204 | A | G | 0.017 | 0.014 | 0.011 | 0.016 | 0.011 | 0.020 | 0.009 | 0.022 | 0.033 | 0.023 |
| rs12364470 | T | G | -0.013 | 0.020 | -0.010 | 0.023 | -0.011 | 0.028 | -0.022 | 0.031 | -0.064 | 0.032 |

| | | | | | | | | | | | |
|---|---|---|---|---|---|---|---|---|---|---|---|
| rs55726687 | G | A | -0.017 | 0.017 | -0.030 | 0.020 | -0.047 | 0.024 | -0.003 | 0.027 | 0.041 | 0.029 |
| rs7976757 | T | C | -0.001 | 0.018 | 0.006 | 0.021 | 0.020 | 0.026 | -0.010 | 0.029 | -0.013 | 0.030 |
| rs7132908 | G | A | -0.021 | 0.014 | -0.015 | 0.017 | 0.001 | 0.021 | -0.032 | 0.023 | -0.033 | 0.024 |
| rs2292238 | A | C | 0.010 | 0.014 | 0.024 | 0.017 | 0.019 | 0.021 | 0.044 | 0.023 | 0.015 | 0.024 |
| rs770082 | G | A | 0.008 | 0.014 | 0.015 | 0.016 | 0.010 | 0.020 | 0.015 | 0.022 | -0.025 | 0.023 |
| rs10849900 | T | C | 0.016 | 0.014 | 0.031 | 0.017 | 0.023 | 0.021 | 0.039 | 0.023 | 0.046 | 0.024 |
| rs111828690 | C | T | -0.080 | 0.017 | -0.074 | 0.020 | -0.066 | 0.024 | -0.085 | 0.027 | -0.066 | 0.028 |
| rs181617194 | T | C | 0.054 | 0.045 | 0.037 | 0.053 | 0.084 | 0.065 | -0.013 | 0.071 | 0.082 | 0.077 |
| rs3803005 | T | C | -0.018 | 0.015 | -0.032 | 0.018 | -0.070 | 0.022 | 0.015 | 0.025 | -0.011 | 0.026 |
| rs9579775 | A | C | -0.039 | 0.023 | -0.038 | 0.027 | -0.047 | 0.033 | -0.013 | 0.037 | -0.008 | 0.039 |
| rs1933440 | A | C | -0.024 | 0.019 | -0.020 | 0.023 | -0.002 | 0.028 | -0.036 | 0.030 | -0.048 | 0.032 |
| rs2761366 | C | T | 0.013 | 0.014 | 0.009 | 0.017 | 0.021 | 0.021 | -0.012 | 0.023 | 0.024 | 0.024 |
| rs9568867 | G | A | 0.003 | 0.020 | -0.006 | 0.024 | -0.016 | 0.029 | -0.002 | 0.032 | 0.050 | 0.034 |
| rs12866691 | A | T | 0.014 | 0.016 | 0.023 | 0.019 | 0.018 | 0.024 | 0.022 | 0.026 | -0.013 | 0.027 |
| rs1576655 | A | C | -0.017 | 0.014 | -0.026 | 0.017 | -0.025 | 0.021 | -0.025 | 0.023 | 0.006 | 0.024 |
| rs7331420 | G | A | 0.016 | 0.015 | 0.020 | 0.018 | 0.032 | 0.022 | 0.013 | 0.024 | -0.004 | 0.025 |
| rs9522180 | C | T | 0.015 | 0.014 | 0.007 | 0.016 | 0.002 | 0.020 | 0.001 | 0.022 | 0.002 | 0.023 |
| rs10142359 | A | G | -0.005 | 0.014 | -0.036 | 0.019 | -0.054 | 0.023 | -0.017 | 0.026 | 0.038 | 0.025 |
| rs8022132 | A | T | -0.012 | 0.015 | 0.000 | 0.020 | -0.013 | 0.026 | 0.006 | 0.029 | -0.007 | 0.031 |
| rs6575340 | G | A | 0.008 | 0.014 | 0.020 | 0.017 | 0.015 | 0.024 | 0.030 | 0.027 | -0.016 | 0.029 |
| rs7145882 | T | C | -0.017 | 0.014 | -0.018 | 0.018 | -0.005 | 0.024 | -0.001 | 0.025 | -0.013 | 0.029 |
| rs12891477 | C | T | 0.024 | 0.016 | 0.024 | 0.022 | 0.048 | 0.027 | -0.006 | 0.027 | 0.046 | 0.035 |
| rs1466276 | C | G | -0.002 | 0.014 | -0.005 | 0.016 | 0.002 | 0.020 | -0.029 | 0.022 | 0.011 | 0.023 |
| rs4776985 | T | G | 0.028 | 0.016 | 0.036 | 0.019 | 0.054 | 0.024 | 0.000 | 0.026 | -0.003 | 0.027 |
| rs67962220 | T | G | -0.013 | 0.019 | -0.030 | 0.022 | -0.005 | 0.027 | -0.055 | 0.030 | -0.014 | 0.031 |
| rs715724 | A | G | 0.004 | 0.014 | -0.004 | 0.017 | -0.009 | 0.021 | 0.004 | 0.023 | 0.014 | 0.024 |
| rs939624 | C | T | -0.020 | 0.014 | 0.007 | 0.016 | -0.007 | 0.020 | 0.011 | 0.022 | -0.027 | 0.023 |
| rs7200589 | G | A | 0.021 | 0.016 | 0.025 | 0.019 | 0.051 | 0.023 | 0.004 | 0.025 | 0.048 | 0.026 |
| rs879620 | C | T | -0.011 | 0.014 | -0.004 | 0.016 | 0.004 | 0.020 | -0.021 | 0.022 | -0.007 | 0.023 |
| rs57790054 | A | G | -0.020 | 0.015 | 0.001 | 0.018 | 0.002 | 0.022 | 0.005 | 0.025 | -0.039 | 0.026 |
| rs11074452 | C | G | 0.003 | 0.014 | 0.008 | 0.016 | -0.003 | 0.020 | 0.016 | 0.022 | 0.049 | 0.023 |
| rs62031562 | A | T | -0.011 | 0.014 | -0.023 | 0.017 | -0.022 | 0.021 | -0.031 | 0.023 | 0.017 | 0.024 |
| rs3814883 | C | T | 0.003 | 0.014 | 0.001 | 0.016 | 0.027 | 0.020 | -0.028 | 0.022 | 0.031 | 0.023 |
| rs34898535 | C | T | 0.027 | 0.014 | 0.015 | 0.017 | 0.025 | 0.020 | 0.001 | 0.023 | 0.026 | 0.024 |
| rs56094641 | A | G | 0.006 | 0.014 | -0.010 | 0.016 | 0.004 | 0.020 | -0.031 | 0.022 | 0.018 | 0.023 |
| rs3751859 | G | A | -0.015 | 0.019 | -0.031 | 0.023 | -0.037 | 0.028 | -0.037 | 0.031 | 0.009 | 0.033 |
| rs4790292 | C | A | -0.023 | 0.020 | -0.020 | 0.023 | 0.002 | 0.028 | -0.044 | 0.031 | -0.025 | 0.033 |
| rs1914889 | G | A | 0.033 | 0.014 | 0.038 | 0.016 | 0.034 | 0.020 | 0.043 | 0.022 | 0.006 | 0.023 |
| rs2306593 | C | T | -0.001 | 0.014 | 0.008 | 0.016 | 0.025 | 0.020 | -0.012 | 0.022 | -0.001 | 0.023 |
| rs11079849 | C | T | -0.016 | 0.015 | -0.016 | 0.017 | -0.024 | 0.021 | -0.007 | 0.023 | -0.019 | 0.024 |
| rs77706698 | G | A | -0.008 | 0.020 | -0.032 | 0.024 | -0.024 | 0.030 | -0.035 | 0.033 | 0.025 | 0.034 |
| rs2619976 | C | T | 0.013 | 0.014 | 0.011 | 0.017 | 0.012 | 0.020 | 0.005 | 0.023 | 0.026 | 0.024 |
| rs11150745 | A | G | -0.022 | 0.015 | -0.020 | 0.017 | -0.008 | 0.021 | -0.032 | 0.023 | -0.018 | 0.025 |
| rs891386 | T | G | 0.019 | 0.014 | 0.010 | 0.016 | 0.014 | 0.020 | 0.009 | 0.022 | 0.026 | 0.023 |
| rs11660335 | T | C | 0.001 | 0.018 | 0.023 | 0.021 | 0.021 | 0.026 | 0.029 | 0.029 | -0.026 | 0.030 |

| | | | | | | | | | | | |
|---|---|---|---|---|---|---|---|---|---|---|---|
| rs784257 | T | C | 0.013 | 0.018 | -0.007 | 0.021 | -0.027 | 0.026 | 0.024 | 0.028 | 0.039 | 0.029 |
| rs66922415 | A | G | -0.022 | 0.016 | -0.005 | 0.019 | -0.002 | 0.023 | -0.008 | 0.025 | -0.035 | 0.027 |
| rs17066856 | T | C | -0.002 | 0.023 | 0.003 | 0.028 | -0.016 | 0.034 | 0.043 | 0.038 | -0.009 | 0.039 |
| rs9962947 | C | T | 0.021 | 0.014 | 0.022 | 0.017 | 0.015 | 0.021 | 0.024 | 0.023 | 0.025 | 0.024 |
| rs12974664 | G | A | -0.015 | 0.014 | -0.020 | 0.016 | -0.037 | 0.020 | -0.007 | 0.022 | -0.019 | 0.023 |
| rs350832 | G | A | 0.050 | 0.017 | 0.029 | 0.019 | -0.012 | 0.024 | 0.080 | 0.026 | 0.072 | 0.027 |
| rs12986231 | T | C | 0.025 | 0.016 | 0.025 | 0.019 | 0.062 | 0.023 | -0.020 | 0.025 | 0.026 | 0.026 |
| rs10404726 | C | T | 0.018 | 0.014 | 0.008 | 0.016 | 0.009 | 0.020 | -0.003 | 0.022 | 0.040 | 0.023 |
| rs56212061 | C | T | -0.037 | 0.019 | -0.040 | 0.022 | -0.042 | 0.027 | -0.047 | 0.030 | -0.029 | 0.031 |
| rs111640872 | G | C | -0.023 | 0.015 | -0.039 | 0.017 | -0.026 | 0.021 | -0.053 | 0.023 | -0.018 | 0.024 |
| rs11880064 | T | C | 0.028 | 0.014 | 0.033 | 0.017 | 0.042 | 0.020 | 0.017 | 0.022 | 0.028 | 0.024 |
| rs429358 | T | C | 0.019 | 0.020 | 0.024 | 0.024 | 0.050 | 0.030 | -0.017 | 0.032 | 0.021 | 0.034 |
| rs12971645 | G | A | 0.014 | 0.016 | 0.003 | 0.018 | 0.022 | 0.023 | -0.027 | 0.025 | 0.034 | 0.026 |
| rs1800437 | G | C | -0.010 | 0.017 | 0.003 | 0.020 | 0.008 | 0.024 | 0.000 | 0.027 | -0.032 | 0.028 |
| rs3810291 | G | A | -0.021 | 0.015 | -0.019 | 0.018 | -0.023 | 0.021 | -0.012 | 0.024 | -0.016 | 0.025 |
| rs8124896 | T | C | -0.008 | 0.022 | -0.011 | 0.026 | -0.017 | 0.032 | 0.011 | 0.036 | 0.032 | 0.038 |
| rs116948922 | C | T | 0.008 | 0.046 | 0.064 | 0.054 | 0.081 | 0.067 | 0.041 | 0.073 | -0.049 | 0.075 |
| rs34966255 | T | C | 0.010 | 0.018 | 0.022 | 0.021 | 0.034 | 0.026 | 0.007 | 0.028 | -0.010 | 0.030 |
| rs915814 | G | A | -0.022 | 0.016 | -0.002 | 0.019 | -0.007 | 0.024 | 0.016 | 0.026 | -0.036 | 0.027 |
| rs400997 | T | A | 0.007 | 0.014 | 0.004 | 0.016 | 0.010 | 0.020 | 0.002 | 0.022 | 0.022 | 0.023 |
| rs738140 | A | G | -0.029 | 0.015 | -0.034 | 0.017 | -0.030 | 0.021 | -0.038 | 0.023 | -0.017 | 0.024 |

Abbreviations: ca cancer; co colon; dist distal; EA effect allele; OA other allele; prox proximal; re rectal; se standard error

| Supplementary table 3: Number of cancer cases by sex and subsite | | | |
|---|---|---|---|
| Cancer | Men & women | Men | Women |
| Overall colorectal | 52,775 | 28,207 | 24,568 |
| Colon | 27,817 | 14,120 | 14,616 |
| Proximal colon | 12,360 | 6,510 | 7,906 |
| Distal colon | 14,016 | 6,853 | 6,026 |
| Rectal | 13,713 | 8,385 | 5,765 |

**Supplementary Table 4: Power calculations for each phenotype and group in the Mendelian randomization study of early and adult life body size and risk of colorectal cancer**

| Exposure/Group | Sample size | Proportion of cases | Power (%) given selected scenarios (Type 1 error of 5%) | | | |
|---|---|---|---|---|---|---|
| | | | OR=1.10 | OR=1.15 | OR=1.20 | OR=1.25 |
| **Early life body size** | | | | | | |
| **Men and women ($r^2$=4.1%)** | | | | | | |
| Overall | 85,638 | 0.50 | 0.81 | 0.99 | 1.00 | 1.00 |
| Colon | 71,488 | 0.40 | 0.72 | 0.96 | 1.00 | 1.00 |
| Proximal colon | 57,168 | 0.25 | 0.53 | 0.86 | 1.00 | 1.00 |
| Distal colon | 55,631 | 0.23 | 0.50 | 0.83 | 0.97 | 1.00 |
| Rectal | 56,902 | 0.25 | 0.53 | 0.86 | 0.98 | 1.00 |
| **Men ($r^2$=2%)** | | | | | | |
| Overall | 43,457 | 0.52 | 0.29 | 0.54 | 0.76 | 0.91 |
| Colon | 35,072 | 0.40 | 0.24 | 0.45 | 0.67 | 0.84 |
| Proximal colon | 27,462 | 0.24 | 0.16 | 0.31 | 0.48 | 0.66 |
| Distal colon | 27,805 | 0.25 | 0.17 | 0.32 | 0.50 | 0.68 |
| Rectal | 29,337 | 0.29 | 0.19 | 0.35 | 0.55 | 0.73 |
| **Women ($r^2$=3.5%)** | | | | | | |
| Overall | 42,181 | 0.48 | 0.45 | 0.77 | 0.94 | 0.99 |
| Colon | 36,416 | 0.40 | 0.39 | 0.70 | 0.90 | 0.98 |
| Proximal colon | 29,706 | 0.27 | 0.29 | 0.54 | 0.78 | 0.92 |
| Distal colon | 27,826 | 0.22 | 0.24 | 0.47 | 0.70 | 0.87 |
| Rectal | 27,565 | 0.21 | 0.24 | 0.45 | 0.68 | 0.86 |
| **Adult life body size** | | | | | | |
| **Men and women ($r^2$=5.5%)** | | | | | | |
| Overall | 85,638 | 0.50 | 0.91 | 1.00 | 1.00 | 1.00 |
| Colon | 71,488 | 0.40 | 0.84 | 0.99 | 1.00 | 1.00 |
| Proximal colon | 57,168 | 0.25 | 0.66 | 0.94 | 1.00 | 1.00 |
| Distal colon | 55,631 | 0.23 | 0.62 | 0.92 | 0.99 | 1.00 |
| Rectal | 56,902 | 0.25 | 0.66 | 0.94 | 1.00 | 1.00 |
| **Men ($r^2$=3.2%)** | | | | | | |
| Overall | 43,457 | 0.52 | 0.43 | 0.74 | 0.92 | 0.99 |
| Colon | 35,072 | 0.40 | 0.35 | 0.64 | 0.86 | 0.96 |

| | | | | | | |
|---|---|---|---|---|---|---|
| Proximal colon | 27,462 | 0.24 | 0.24 | 0.45 | 0.68 | 0.85 |
| Distal colon | 27,805 | 0.25 | 0.24 | 0.46 | 0.69 | 0.86 |
| Rectal | 29,337 | 0.29 | 0.27 | 0.52 | 0.75 | 0.90 |
| **Women ($r^2$=3.6%)** | | | | | | |
| Overall | 42,181 | 0.48 | 0.46 | 0.78 | 0.95 | 0.99 |
| Colon | 36,416 | 0.40 | 0.40 | 0.71 | 0.91 | 0.98 |
| Proximal colon | 29,706 | 0.27 | 0.29 | 0.55 | 0.79 | 0.93 |
| Distal colon | 27,826 | 0.22 | 0.25 | 0.48 | 0.71 | 0.88 |
| Rectal | 27,565 | 0.21 | 0.24 | 0.46 | 0.70 | 0.86 |

Abbreviations: OR, odds ratio

| Supplementary table 5: Beta estimates for early/adult life body size in UK Biobank for the SNPs included in the MVMR analysis (overall, men, women) | | | | | | | | |
|---|---|---|---|---|---|---|---|---|
| Overall | | | | | | | | |
| SNP | EA | OA | beta_early | se_early | pval_early | beta_adult | se_adult | pval_adult |
| rs2229330 | T | G | -0.01976 | 0.002694 | 2.20E-13 | -0.00347 | 0.002635 | 0.19 |
| rs2175171 | G | C | -0.00775 | 0.00141 | 3.90E-08 | -0.00193 | 0.001379 | 0.16 |
| rs78886584 | A | G | -0.00155 | 0.001416 | 0.27 | -0.00849 | 0.001385 | 8.70E-10 |
| rs212517 | T | A | 0.00877 | 0.001431 | 8.90E-10 | 0.001562 | 0.0014 | 0.26 |
| rs10799778 | T | G | 0.007021 | 0.00188 | 0.00019 | 0.012582 | 0.001839 | 7.80E-12 |
| rs3737992 | G | A | 0.003099 | 0.001866 | 0.097 | 0.013732 | 0.001825 | 5.30E-14 |
| rs12031634 | G | A | 0.003598 | 0.001538 | 0.019 | 0.008806 | 0.001505 | 4.80E-09 |
| rs6669341 | A | G | 0.006292 | 0.001418 | 9.10E-06 | 0.010685 | 0.001387 | 1.30E-14 |
| rs1167311 | G | A | 0.004807 | 0.001512 | 0.0015 | 0.012253 | 0.001479 | 1.20E-16 |
| rs12140153 | G | T | 0.021814 | 0.002454 | 6.20E-19 | 0.021447 | 0.002401 | 4.20E-19 |
| rs2767486 | A | G | -0.01538 | 0.001743 | 1.10E-18 | -0.00136 | 0.001705 | 0.42 |
| rs12042908 | A | G | 0.027464 | 0.001411 | 2.30E-84 | 0.010443 | 0.00138 | 3.90E-14 |
| rs34517439 | C | A | -0.01374 | 0.002164 | 2.20E-10 | -0.0247 | 0.002117 | 1.90E-31 |
| rs6679458 | G | T | -0.00669 | 0.00142 | 2.50E-06 | -0.01156 | 0.00139 | 9.10E-17 |
| rs12072739 | A | G | -0.00087 | 0.00168 | 0.6 | -0.01159 | 0.001643 | 1.80E-12 |
| rs7550711 | C | T | -0.04848 | 0.004412 | 4.40E-28 | -0.04264 | 0.004316 | 5.10E-23 |
| rs12033257 | A | G | -0.00072 | 0.001451 | 0.62 | 0.009803 | 0.001419 | 4.90E-12 |
| rs7549358 | G | C | 0.003618 | 0.001461 | 0.013 | 0.00883 | 0.001429 | 6.50E-10 |
| rs142315514 | C | A | -0.01387 | 0.003874 | 0.00034 | -0.02139 | 0.00379 | 1.70E-08 |
| rs61813324 | C | T | -0.00536 | 0.002073 | 0.0097 | -0.0183 | 0.002027 | 1.80E-19 |
| rs1778830 | G | A | -0.00371 | 0.001459 | 0.011 | -0.00991 | 0.001427 | 3.80E-12 |
| rs4916229 | C | G | 0.000356 | 0.002385 | 0.88 | -0.01513 | 0.002333 | 8.90E-11 |
| rs543874 | A | G | -0.04709 | 0.001732 | 8.80E-163 | -0.03012 | 0.001694 | 1.00E-70 |
| rs815163 | T | C | 0.005829 | 0.001409 | 3.50E-05 | 0.010754 | 0.001378 | 6.10E-15 |
| rs76702514 | C | G | 0.003874 | 0.001727 | 0.025 | 0.010137 | 0.001689 | 2.00E-09 |
| rs2678204 | T | G | -0.00575 | 0.001477 | 9.80E-05 | -0.01502 | 0.001445 | 2.70E-25 |
| rs9438393 | A | G | 0.01036 | 0.001422 | 3.10E-13 | 0.005266 | 0.001391 | 0.00015 |
| rs12037905 | C | T | 0.000746 | 0.001419 | 0.6 | 0.007927 | 0.001388 | 1.10E-08 |
| rs7354849 | A | G | -0.00815 | 0.00141 | 7.40E-09 | -0.00426 | 0.00138 | 0.002 |
| rs10927006 | T | C | 0.000898 | 0.001997 | 0.65 | 0.011993 | 0.001954 | 8.30E-10 |
| rs4658403 | C | T | 0.000129 | 0.001881 | 0.95 | 0.013642 | 0.00184 | 1.20E-13 |
| rs62106258 | T | C | 0.080258 | 0.003254 | 2.40E-134 | 0.059139 | 0.003182 | 4.20E-77 |
| rs12992672 | G | A | -0.04332 | 0.001852 | 5.50E-121 | -0.03595 | 0.001811 | 1.20E-87 |
| rs2867116 | C | A | -0.01228 | 0.002028 | 1.40E-09 | -0.00694 | 0.001983 | 0.00047 |
| rs10182458 | A | G | -0.03592 | 0.0014 | 3.10E-145 | -0.01995 | 0.001369 | 4.10E-48 |
| rs146910503 | G | A | 0.033464 | 0.005025 | 2.70E-11 | 0.014782 | 0.004914 | 0.0026 |
| rs10169594 | T | C | -0.00048 | 0.001459 | 0.74 | -0.00788 | 0.001427 | 3.30E-08 |
| rs35809007 | G | A | 0.004337 | 0.001459 | 0.003 | 0.010592 | 0.001427 | 1.20E-13 |
| rs6761463 | G | C | 0.003653 | 0.0019 | 0.054 | 0.012581 | 0.001858 | 1.30E-11 |
| rs4671328 | T | G | 0.005629 | 0.001417 | 7.20E-05 | 0.01317 | 0.001386 | 2.10E-21 |
| rs4672338 | C | T | -0.00458 | 0.001481 | 0.002 | -0.00835 | 0.001448 | 8.00E-09 |

| | | | | | | | | |
|---|---|---|---|---|---|---|---|---|
| rs12713889 | T | C | 0.011692 | 0.001487 | 3.70E-15 | 0.005705 | 0.001454 | 8.70E-05 |
| rs6752979 | G | A | -0.00366 | 0.001503 | 0.015 | -0.00896 | 0.00147 | 1.10E-09 |
| rs396354 | T | C | 0.001351 | 0.001555 | 0.38 | 0.010398 | 0.00152 | 8.00E-12 |
| rs11691869 | C | A | -0.00034 | 0.001459 | 0.82 | 0.011231 | 0.001427 | 3.50E-15 |
| rs1451533 | G | A | -0.00383 | 0.001581 | 0.015 | -0.01097 | 0.001546 | 1.30E-12 |
| rs1384660 | G | A | 0.015794 | 0.001802 | 1.90E-18 | 0.007457 | 0.001763 | 2.30E-05 |
| rs409696 | G | A | 0.001534 | 0.001418 | 0.28 | 0.011422 | 0.001387 | 1.80E-16 |
| rs62175963 | T | C | 0.008974 | 0.001415 | 2.20E-10 | -0.00263 | 0.001383 | 0.057 |
| rs788163 | A | C | -0.00359 | 0.001571 | 0.022 | -0.01006 | 0.001536 | 5.90E-11 |
| rs1704190 | G | A | -0.00121 | 0.001452 | 0.41 | -0.00833 | 0.001421 | 4.50E-09 |
| rs115319174 | G | C | -0.0423 | 0.003036 | 4.00E-44 | -0.01131 | 0.002969 | 0.00014 |
| rs13427822 | A | G | -0.0006 | 0.001591 | 0.71 | 0.009449 | 0.001556 | 1.30E-09 |
| rs55658481 | G | A | -0.00815 | 0.001477 | 3.30E-08 | -0.00897 | 0.001444 | 5.30E-10 |
| rs2433733 | G | A | 0.000678 | 0.001497 | 0.65 | 0.010504 | 0.001464 | 7.30E-13 |
| rs1476698 | A | G | 0.008209 | 0.00145 | 1.50E-08 | 0.001385 | 0.001418 | 0.33 |
| rs2594994 | T | A | 0.015494 | 0.00183 | 2.50E-17 | 0.002107 | 0.00179 | 0.24 |
| rs34373881 | G | A | 0.001937 | 0.001571 | 0.22 | 0.008833 | 0.001537 | 9.00E-09 |
| rs7619139 | T | A | -0.00981 | 0.001427 | 6.20E-12 | -0.00884 | 0.001395 | 2.40E-10 |
| rs80082536 | A | G | -0.00712 | 0.002173 | 0.001 | -0.01289 | 0.002125 | 1.30E-09 |
| rs754635 | C | G | -0.01591 | 0.002206 | 5.50E-13 | -0.01502 | 0.002158 | 3.30E-12 |
| rs28350 | A | G | 0.009691 | 0.001832 | 1.20E-07 | 0.012636 | 0.001791 | 1.70E-12 |
| rs9843653 | T | C | -0.00405 | 0.001402 | 0.0038 | -0.01755 | 0.001371 | 1.50E-37 |
| rs6445198 | G | T | 0.012532 | 0.001424 | 1.40E-18 | 0.010961 | 0.001393 | 3.50E-15 |
| rs6445258 | T | C | 0.001297 | 0.001725 | 0.45 | 0.009711 | 0.001687 | 8.60E-09 |
| rs538579 | G | C | -0.00841 | 0.001509 | 2.50E-08 | -0.00799 | 0.001476 | 6.30E-08 |
| rs11708540 | G | A | 0.002044 | 0.001933 | 0.29 | -0.01062 | 0.001891 | 2.00E-08 |
| rs4677156 | A | T | 0.009241 | 0.00168 | 3.70E-08 | 0.003783 | 0.001642 | 0.021 |
| rs1598121 | A | G | -0.00322 | 0.001455 | 0.027 | -0.00896 | 0.001423 | 3.00E-10 |
| rs6783281 | A | G | -0.00955 | 0.001541 | 5.70E-10 | -0.00331 | 0.001507 | 0.028 |
| rs4858940 | T | C | -0.01696 | 0.002199 | 1.30E-14 | -0.01331 | 0.002151 | 6.00E-10 |
| rs1454687 | C | G | 0.001216 | 0.0014 | 0.39 | 0.012283 | 0.00137 | 3.00E-19 |
| rs1436348 | A | G | -0.00445 | 0.00142 | 0.0017 | -0.00966 | 0.001389 | 3.50E-12 |
| rs9814758 | T | G | 0.004915 | 0.00147 | 0.00082 | 0.008387 | 0.001437 | 5.40E-09 |
| rs1320903 | G | A | -0.01003 | 0.001502 | 2.50E-11 | -0.01462 | 0.001469 | 2.60E-23 |
| rs2343681 | G | A | -0.00449 | 0.001715 | 0.0089 | -0.01236 | 0.001678 | 1.80E-13 |
| rs59714050 | T | A | -0.02282 | 0.002815 | 5.20E-16 | -0.01937 | 0.002753 | 2.00E-12 |
| rs1568488 | G | C | -0.00388 | 0.001438 | 0.007 | -0.01103 | 0.001406 | 4.40E-15 |
| rs8192675 | T | C | -0.00076 | 0.001543 | 0.62 | -0.0117 | 0.001509 | 8.70E-15 |
| rs7633995 | A | G | -0.0145 | 0.002476 | 4.70E-09 | -0.00725 | 0.002421 | 0.0027 |
| rs869400 | T | G | -0.00922 | 0.001806 | 3.30E-07 | -0.01831 | 0.001766 | 3.50E-25 |
| rs6583310 | G | C | -0.00323 | 0.001415 | 0.022 | -0.00878 | 0.001384 | 2.20E-10 |
| rs2051559 | T | C | -0.01031 | 0.002073 | 6.70E-07 | -0.01386 | 0.002029 | 8.30E-12 |
| rs34811474 | G | A | 0.010192 | 0.001663 | 8.80E-10 | 0.016596 | 0.001627 | 2.00E-24 |
| rs73213484 | A | T | 0.001239 | 0.002014 | 0.54 | 0.014638 | 0.001971 | 1.10E-13 |
| rs7439324 | C | T | 0.010792 | 0.001903 | 1.40E-08 | 0.007527 | 0.001862 | 5.30E-05 |

| | | | | | | | | |
|---|---|---|---|---|---|---|---|---|
| rs12641981 | C | T | -0.02219 | 0.001415 | 2.20E-55 | -0.01867 | 0.001385 | 1.90E-41 |
| rs2237025 | T | C | -0.00029 | 0.001422 | 0.84 | 0.010243 | 0.001391 | 1.80E-13 |
| rs28462076 | A | G | 0.006227 | 0.001656 | 0.00017 | 0.009393 | 0.00162 | 6.70E-09 |
| rs788858 | A | G | 0.012315 | 0.001543 | 1.50E-15 | 0.000576 | 0.00151 | 0.7 |
| rs4148155 | A | G | 0.00639 | 0.002205 | 0.0038 | 0.014349 | 0.002158 | 2.90E-11 |
| rs4419475 | A | T | 0.001147 | 0.001426 | 0.42 | -0.0081 | 0.001395 | 6.50E-09 |
| rs1229984 | T | C | 0.000533 | 0.004256 | 0.9 | -0.02304 | 0.004164 | 3.20E-08 |
| rs13107325 | C | T | -0.0215 | 0.002663 | 6.80E-16 | -0.02871 | 0.002606 | 3.10E-28 |
| rs72675820 | A | C | 0.009938 | 0.001461 | 1.00E-11 | 0.008039 | 0.001429 | 1.90E-08 |
| rs1296328 | A | C | 0.008046 | 0.001418 | 1.40E-08 | 0.011738 | 0.001387 | 2.60E-17 |
| rs11727676 | T | C | 0.013114 | 0.002371 | 3.20E-08 | 0.007291 | 0.00232 | 0.0017 |
| rs6843852 | C | T | -0.00477 | 0.001401 | 0.00066 | -0.00872 | 0.001371 | 2.00E-10 |
| rs698147 | A | G | 0.001409 | 0.001407 | 0.32 | 0.00878 | 0.001377 | 1.80E-10 |
| rs67913249 | C | G | 0.003096 | 0.001482 | 0.037 | 0.009346 | 0.00145 | 1.20E-10 |
| rs10805383 | G | A | -0.00154 | 0.001404 | 0.27 | -0.01047 | 0.001374 | 2.60E-14 |
| rs9291816 | C | T | 0.013423 | 0.001498 | 3.20E-19 | 0.007976 | 0.001466 | 5.30E-08 |
| rs2307111 | T | C | 0.008736 | 0.001433 | 1.10E-09 | 0.016755 | 0.001403 | 6.90E-33 |
| rs1422067 | C | T | 0.010831 | 0.001645 | 4.60E-11 | 0.008702 | 0.00161 | 6.50E-08 |
| rs6870983 | C | T | 0.009676 | 0.00171 | 1.50E-08 | 0.013797 | 0.001673 | 1.60E-16 |
| rs1477290 | T | C | -0.00813 | 0.002054 | 7.50E-05 | -0.01994 | 0.00201 | 3.50E-23 |
| rs77960 | G | A | 0.010476 | 0.001493 | 2.20E-12 | -0.00624 | 0.001461 | 1.90E-05 |
| rs149457 | C | T | 0.00199 | 0.00187 | 0.29 | 0.015032 | 0.00183 | 2.10E-16 |
| rs12517187 | C | T | -0.0045 | 0.001418 | 0.0015 | -0.00825 | 0.001388 | 2.80E-09 |
| rs347551 | C | G | -0.00312 | 0.001425 | 0.028 | -0.00889 | 0.001395 | 1.80E-10 |
| rs1582931 | G | A | 0.005279 | 0.001415 | 0.00019 | 0.009657 | 0.001384 | 3.00E-12 |
| rs13174863 | A | G | -0.00402 | 0.001987 | 0.043 | -0.01362 | 0.001944 | 2.40E-12 |
| rs4958568 | G | A | 0.009942 | 0.001567 | 2.20E-10 | 0.004806 | 0.001533 | 0.0017 |
| rs7719067 | A | G | 0.013497 | 0.001414 | 1.40E-21 | 0.00931 | 0.001384 | 1.70E-11 |
| rs11134679 | A | G | -0.00858 | 0.001512 | 1.40E-08 | -0.0124 | 0.00148 | 5.20E-17 |
| rs12214497 | G | T | 0.010509 | 0.001473 | 9.70E-13 | 0.005635 | 0.001442 | 9.30E-05 |
| rs4467770 | G | A | -0.00628 | 0.001586 | 7.60E-05 | -0.00987 | 0.001552 | 2.00E-10 |
| rs3806114 | G | A | 0.001521 | 0.001504 | 0.31 | 0.008789 | 0.001472 | 2.30E-09 |
| rs35162296 | C | T | -0.02003 | 0.002272 | 1.20E-18 | -0.01142 | 0.002223 | 2.80E-07 |
| rs9366863 | T | C | 0.007605 | 0.001489 | 3.30E-07 | 0.017384 | 0.001457 | 8.50E-33 |
| rs34298980 | T | C | 0.005895 | 0.001476 | 6.50E-05 | 0.013257 | 0.001444 | 4.40E-20 |
| rs72892910 | G | T | -0.02465 | 0.001858 | 3.40E-40 | -0.02528 | 0.001818 | 5.70E-44 |
| rs1775255 | G | T | -0.01333 | 0.001405 | 2.30E-21 | -0.00946 | 0.001375 | 6.10E-12 |
| rs1342831 | T | C | -0.0225 | 0.003016 | 8.70E-14 | -0.00659 | 0.002952 | 0.026 |
| rs72910629 | A | G | 0.002266 | 0.002055 | 0.27 | -0.01325 | 0.002011 | 4.30E-11 |
| rs1040046 | C | A | -0.00122 | 0.001963 | 0.53 | -0.01126 | 0.001921 | 4.60E-09 |
| rs10499014 | C | G | -0.00065 | 0.001589 | 0.68 | 0.010046 | 0.001556 | 1.10E-10 |
| rs34260097 | T | G | -0.01782 | 0.001679 | 2.50E-26 | -0.00236 | 0.001643 | 0.15 |
| rs13218383 | C | G | 0.004211 | 0.001483 | 0.0045 | 0.009318 | 0.001452 | 1.40E-10 |
| rs2875762 | G | C | -0.00519 | 0.001639 | 0.0016 | -0.01104 | 0.001604 | 6.00E-12 |
| rs1452991 | G | A | -0.01005 | 0.001458 | 5.40E-12 | -0.00577 | 0.001427 | 5.30E-05 |

| | | | | | | | | |
|---|---|---|---|---|---|---|---|---|
| rs12213441 | C | T | -0.00387 | 0.001708 | 0.024 | -0.01051 | 0.001672 | 3.30E-10 |
| rs7749708 | C | T | -0.00469 | 0.001541 | 0.0024 | -0.00998 | 0.001509 | 3.70E-11 |
| rs796915 | C | G | -0.01289 | 0.001523 | 2.60E-17 | -0.00737 | 0.001491 | 7.70E-07 |
| rs62425398 | C | A | -0.01533 | 0.002284 | 1.90E-11 | -0.00912 | 0.002235 | 4.50E-05 |
| rs6950388 | G | A | 0.001178 | 0.001735 | 0.5 | -0.00945 | 0.001698 | 2.60E-08 |
| rs2722406 | C | T | -0.01218 | 0.001555 | 4.60E-15 | -0.00431 | 0.001522 | 0.0046 |
| rs2289379 | C | T | 0.004568 | 0.001438 | 0.0015 | 0.009759 | 0.001408 | 4.10E-12 |
| rs3823674 | C | T | 0.004829 | 0.001419 | 0.00066 | 0.007657 | 0.001389 | 3.50E-08 |
| rs11765062 | T | C | 0.000958 | 0.001405 | 0.5 | 0.007516 | 0.001375 | 4.60E-08 |
| rs2866720 | C | T | -0.00147 | 0.001449 | 0.31 | -0.00913 | 0.001418 | 1.20E-10 |
| rs1852006 | G | A | 0.008796 | 0.001461 | 1.70E-09 | 0.008831 | 0.00143 | 6.70E-10 |
| rs6963840 | C | T | -0.00683 | 0.001934 | 0.00042 | -0.01293 | 0.001894 | 8.50E-12 |
| rs6974282 | C | T | 0.010058 | 0.001778 | 1.50E-08 | -0.00379 | 0.00174 | 0.03 |
| rs7808296 | C | T | -0.01004 | 0.001508 | 2.80E-11 | -0.00255 | 0.001476 | 0.085 |
| rs11496125 | C | T | -0.00853 | 0.001425 | 2.10E-09 | -0.01092 | 0.001395 | 4.80E-15 |
| rs1840660 | G | A | 0.001444 | 0.001445 | 0.32 | -0.00995 | 0.001415 | 2.00E-12 |
| rs6979832 | A | G | -0.00939 | 0.001409 | 2.70E-11 | -0.0033 | 0.00138 | 0.017 |
| rs11976084 | C | T | -0.00661 | 0.001552 | 2.10E-05 | -0.00857 | 0.001519 | 1.70E-08 |
| rs11525873 | T | C | 0.017431 | 0.002363 | 1.60E-13 | 0.014829 | 0.002313 | 1.40E-10 |
| rs1805123 | T | G | 0.005842 | 0.001628 | 0.00033 | 0.011047 | 0.001593 | 4.10E-12 |
| rs77976727 | C | T | -0.01516 | 0.002409 | 3.10E-10 | -0.00744 | 0.002358 | 0.0016 |
| rs55896564 | G | A | 0.0091 | 0.001412 | 1.20E-10 | 0.011697 | 0.001382 | 2.60E-17 |
| rs6530737 | A | G | 0.001404 | 0.001468 | 0.34 | 0.009019 | 0.001437 | 3.50E-10 |
| rs7012648 | G | A | -0.00974 | 0.001426 | 8.40E-12 | -0.00685 | 0.001396 | 9.30E-07 |
| rs2725371 | A | G | -0.00349 | 0.001529 | 0.022 | 0.010038 | 0.001497 | 2.00E-11 |
| rs4739558 | A | G | 0.0083 | 0.001431 | 6.60E-09 | 0.008097 | 0.001401 | 7.40E-09 |
| rs10095724 | G | A | 0.009381 | 0.001461 | 1.40E-10 | 0.002955 | 0.00143 | 0.039 |
| rs10111937 | C | T | -0.00838 | 0.001528 | 4.20E-08 | -0.0049 | 0.001496 | 0.0011 |
| rs13254613 | A | C | -0.01256 | 0.001475 | 1.60E-17 | 0.005649 | 0.001444 | 9.10E-05 |
| rs35957544 | G | T | 0.00133 | 0.00142 | 0.35 | 0.012679 | 0.00139 | 7.40E-20 |
| rs17619860 | T | C | -0.00572 | 0.0019 | 0.0026 | -0.0104 | 0.00186 | 2.20E-08 |
| rs2114210 | G | A | -0.003 | 0.001485 | 0.043 | -0.00986 | 0.001454 | 1.20E-11 |
| rs17716502 | C | T | 0.004757 | 0.001751 | 0.0066 | 0.016236 | 0.001714 | 2.70E-21 |
| rs112875651 | G | A | -0.0048 | 0.001453 | 0.00095 | -0.00835 | 0.001423 | 4.30E-09 |
| rs11782074 | G | T | 0.000824 | 0.001464 | 0.57 | -0.00972 | 0.001433 | 1.20E-11 |
| rs7020196 | C | T | 0.000549 | 0.001443 | 0.7 | 0.007789 | 0.001413 | 3.60E-08 |
| rs13292699 | A | C | -0.00146 | 0.001416 | 0.3 | 0.012536 | 0.001386 | 1.50E-19 |
| rs1411432 | A | C | 0.001512 | 0.001806 | 0.4 | -0.01244 | 0.001768 | 2.00E-12 |
| rs17770336 | C | T | -0.00383 | 0.001496 | 0.01 | -0.01552 | 0.001464 | 3.10E-26 |
| rs7038966 | C | T | -0.00364 | 0.001431 | 0.011 | -0.00948 | 0.001401 | 1.40E-11 |
| rs4744246 | A | G | -0.01575 | 0.001479 | 1.70E-26 | 0.000944 | 0.001448 | 0.51 |
| rs4989244 | G | A | 0.002625 | 0.001415 | 0.064 | 0.007635 | 0.001386 | 3.60E-08 |
| rs7020564 | A | T | 0.009972 | 0.001553 | 1.40E-10 | 0.004974 | 0.001521 | 0.0011 |
| rs957512 | T | C | 0.010327 | 0.001494 | 4.70E-12 | 0.008797 | 0.001463 | 1.80E-09 |
| rs10760277 | C | T | -0.00171 | 0.001444 | 0.24 | -0.00887 | 0.001414 | 3.60E-10 |

| | | | | | | | | |
|---|---|---|---|---|---|---|---|---|
| rs117911387 | G | A | -0.02458 | 0.003331 | 1.60E-13 | -0.00368 | 0.003261 | 0.26 |
| rs113132247 | G | A | -0.00552 | 0.001952 | 0.0047 | -0.01233 | 0.001911 | 1.10E-10 |
| rs7084503 | T | C | 0.012881 | 0.001408 | 5.70E-20 | 0.004868 | 0.001378 | 0.00041 |
| rs11256627 | G | A | -0.00888 | 0.001546 | 9.30E-09 | -0.00095 | 0.001514 | 0.53 |
| rs7893571 | G | T | -0.00194 | 0.001489 | 0.19 | -0.01018 | 0.001457 | 2.90E-12 |
| rs12253527 | G | A | -4.03E-05 | 0.001501 | 0.98 | -0.01361 | 0.00147 | 2.00E-20 |
| rs71495049 | G | A | -0.00032 | 0.002532 | 0.9 | -0.01707 | 0.002478 | 5.70E-12 |
| rs7924036 | G | T | 0.00413 | 0.001401 | 0.0032 | 0.009634 | 0.001372 | 2.20E-12 |
| rs10823504 | G | A | 0.015808 | 0.002873 | 3.70E-08 | 0.000902 | 0.002812 | 0.75 |
| rs11000993 | T | C | -0.00634 | 0.002124 | 0.0029 | -0.01381 | 0.002079 | 3.10E-11 |
| rs17399739 | A | G | -0.02097 | 0.002771 | 3.80E-14 | -0.01775 | 0.002712 | 5.90E-11 |
| rs10887571 | C | T | -0.00799 | 0.001419 | 1.80E-08 | -0.00682 | 0.001389 | 9.10E-07 |
| rs2450444 | G | A | 0.005248 | 0.001469 | 0.00035 | 0.008074 | 0.001438 | 2.00E-08 |
| rs41310284 | C | A | 0.020588 | 0.002341 | 1.40E-18 | 0.017741 | 0.002291 | 9.80E-15 |
| rs7086898 | A | G | -0.00494 | 0.002587 | 0.056 | -0.01387 | 0.002533 | 4.30E-08 |
| rs4575195 | C | A | 0.004438 | 0.001512 | 0.0033 | 0.009594 | 0.00148 | 9.00E-11 |
| rs75387636 | G | A | -0.02122 | 0.00351 | 1.50E-09 | -0.00746 | 0.003435 | 0.03 |
| rs4962725 | T | C | -0.00145 | 0.001417 | 0.31 | -0.00997 | 0.001387 | 6.60E-13 |
| rs2035806 | G | A | 0.002565 | 0.001416 | 0.07 | 0.009549 | 0.001386 | 5.70E-12 |
| rs67257872 | A | G | 0.007025 | 0.001409 | 6.20E-07 | 0.010598 | 0.001379 | 1.50E-14 |
| rs28711392 | T | C | 0.005801 | 0.001465 | 7.50E-05 | 0.011779 | 0.001434 | 2.10E-16 |
| rs6265 | C | T | 0.011724 | 0.001791 | 5.80E-11 | 0.024542 | 0.001753 | 1.50E-44 |
| rs1222216 | C | T | 0.00506 | 0.001672 | 0.0025 | 0.012152 | 0.001637 | 1.10E-13 |
| rs59227842 | A | G | -0.00056 | 0.001526 | 0.71 | -0.01524 | 0.001494 | 1.80E-24 |
| rs868784 | G | A | 0.002281 | 0.001447 | 0.11 | 0.007872 | 0.001417 | 2.70E-08 |
| rs12798028 | C | T | -0.01458 | 0.001422 | 1.10E-24 | -0.01503 | 0.001392 | 3.40E-27 |
| rs34292685 | C | T | 0.005328 | 0.0019 | 0.005 | 0.012723 | 0.00186 | 7.90E-12 |
| rs2234458 | C | T | 0.012552 | 0.001457 | 6.90E-18 | 0.012819 | 0.001426 | 2.50E-19 |
| rs7102934 | T | C | -0.00507 | 0.001526 | 0.00088 | -0.00908 | 0.001494 | 1.20E-09 |
| rs61903695 | A | G | -0.00189 | 0.001609 | 0.24 | -0.01132 | 0.001575 | 6.60E-13 |
| rs680071 | T | C | -0.00614 | 0.00216 | 0.0044 | -0.01174 | 0.002114 | 2.80E-08 |
| rs11215403 | G | A | 0.013251 | 0.001634 | 5.10E-16 | 0.00591 | 0.0016 | 0.00022 |
| rs7925100 | G | A | -0.00313 | 0.001433 | 0.029 | -0.00917 | 0.001403 | 6.30E-11 |
| rs10791113 | A | G | -0.00394 | 0.001405 | 0.005 | -0.00857 | 0.001375 | 4.60E-10 |
| rs12788343 | T | C | -0.00509 | 0.001423 | 0.00035 | -0.00965 | 0.001393 | 4.30E-12 |
| rs329651 | G | T | -0.00714 | 0.001773 | 5.60E-05 | -0.01109 | 0.001736 | 1.70E-10 |
| rs55726687 | G | A | -0.01454 | 0.001718 | 2.70E-17 | -0.01376 | 0.001683 | 3.00E-16 |
| rs2187642 | A | C | -0.01116 | 0.001445 | 1.10E-14 | -0.00158 | 0.001415 | 0.26 |
| rs10505836 | A | C | -0.00308 | 0.002034 | 0.13 | -0.01214 | 0.001992 | 1.10E-09 |
| rs10842356 | A | T | 0.008236 | 0.0014 | 4.10E-09 | 0.005187 | 0.001371 | 0.00016 |
| rs61937656 | G | A | 0.011731 | 0.001678 | 2.70E-12 | 0.005067 | 0.001644 | 0.002 |
| rs7958241 | A | G | -0.01308 | 0.001476 | 7.90E-19 | -0.00499 | 0.001445 | 0.00055 |
| rs7132908 | G | A | -0.03126 | 0.00144 | 1.70E-104 | -0.01862 | 0.00141 | 8.40E-40 |
| rs836179 | A | G | 0.009835 | 0.001451 | 1.20E-11 | 0.002372 | 0.001421 | 0.095 |
| rs7306710 | T | C | 0.009962 | 0.00141 | 1.60E-12 | 0.000735 | 0.001381 | 0.59 |

| | | | | | | | | |
|---|---|---|---|---|---|---|---|---|
| rs61754230 | C | T | -0.00983 | 0.005032 | 0.051 | -0.02738 | 0.004928 | 2.80E-08 |
| rs12427047 | C | T | 0.001401 | 0.001633 | 0.39 | 0.01091 | 0.001599 | 8.90E-12 |
| rs10860295 | T | C | -0.00848 | 0.00141 | 1.80E-09 | -0.00181 | 0.001381 | 0.19 |
| rs4764949 | A | G | 0.008005 | 0.001496 | 8.70E-08 | 0.011935 | 0.001465 | 3.80E-16 |
| rs7305424 | A | T | -0.0104 | 0.001483 | 2.30E-12 | -0.00388 | 0.001453 | 0.0076 |
| rs147730268 | G | T | -0.00114 | 0.002537 | 0.65 | 0.022832 | 0.002485 | 4.00E-20 |
| rs9507791 | G | A | 0.002853 | 0.001719 | 0.097 | 0.009884 | 0.001683 | 4.30E-09 |
| rs1933437 | G | A | 0.014119 | 0.001446 | 1.60E-22 | 0.007247 | 0.001415 | 3.10E-07 |
| rs35193668 | C | T | 0.005677 | 0.001461 | 1.00E-04 | 0.01047 | 0.001431 | 2.50E-13 |
| rs9603697 | C | T | -0.01294 | 0.001496 | 5.00E-18 | -0.00853 | 0.001465 | 5.70E-09 |
| rs12429545 | G | A | -0.02063 | 0.002106 | 1.20E-22 | -0.02003 | 0.002062 | 2.60E-22 |
| rs7321285 | A | C | 0.001457 | 0.001756 | 0.41 | 0.011348 | 0.001719 | 4.10E-11 |
| rs2576135 | T | A | 0.001255 | 0.002438 | 0.61 | 0.013167 | 0.002387 | 3.50E-08 |
| rs1333010 | G | A | 0.012054 | 0.001438 | 5.20E-17 | 0.006531 | 0.001408 | 3.50E-06 |
| rs1576655 | A | C | -0.01185 | 0.001458 | 4.30E-16 | -0.01135 | 0.001427 | 1.90E-15 |
| rs61971082 | T | G | 0.000158 | 0.001556 | 0.92 | -0.01013 | 0.001523 | 3.00E-11 |
| rs7331420 | G | A | -0.00074 | 0.001558 | 0.63 | 0.009142 | 0.001526 | 2.10E-09 |
| rs9888533 | C | T | -0.00114 | 0.001429 | 0.42 | -0.00765 | 0.001399 | 4.50E-08 |
| rs9522180 | C | T | 0.001565 | 0.001412 | 0.27 | 0.009236 | 0.001382 | 2.40E-11 |
| rs9515446 | A | G | -0.00046 | 0.001409 | 0.75 | -0.00969 | 0.00138 | 2.20E-12 |
| rs9788550 | G | C | 0.007689 | 0.001629 | 2.40E-06 | 0.013787 | 0.001596 | 5.60E-18 |
| rs61978655 | G | A | -0.03396 | 0.003647 | 1.30E-20 | -0.01769 | 0.003572 | 7.30E-07 |
| rs724623 | A | C | 0.00437 | 0.001403 | 0.0018 | 0.010244 | 0.001374 | 9.00E-14 |
| rs217672 | A | C | 0.001024 | 0.001579 | 0.52 | -0.01181 | 0.001547 | 2.30E-14 |
| rs3902951 | T | G | -0.00473 | 0.001666 | 0.0045 | -0.01052 | 0.001632 | 1.10E-10 |
| rs10146997 | A | G | -0.00867 | 0.001686 | 2.70E-07 | -0.016 | 0.001651 | 3.20E-22 |
| rs1286138 | T | G | -0.00543 | 0.001498 | 0.00029 | -0.009 | 0.001467 | 8.40E-10 |
| rs6575340 | G | A | -0.0057 | 0.00146 | 9.40E-05 | -0.01332 | 0.00143 | 1.20E-20 |
| rs61992671 | A | G | 0.007593 | 0.001465 | 2.20E-07 | 0.00988 | 0.001435 | 5.70E-12 |
| rs7145882 | T | C | 0.008194 | 0.001476 | 2.80E-08 | 0.01146 | 0.001445 | 2.20E-15 |
| rs824207 | A | G | -0.00929 | 0.001404 | 3.70E-11 | -0.00221 | 0.001375 | 0.11 |
| rs62048187 | G | C | -0.00841 | 0.001532 | 4.00E-08 | -0.00601 | 0.001501 | 6.10E-05 |
| rs10519136 | C | T | 0.008088 | 0.001432 | 1.60E-08 | 0.006171 | 0.001403 | 1.10E-05 |
| rs7182917 | T | C | 0.002252 | 0.001414 | 0.11 | 0.008737 | 0.001385 | 2.80E-10 |
| rs28408562 | C | G | 4.34E-05 | 0.001408 | 0.98 | -0.00784 | 0.00138 | 1.30E-08 |
| rs8030456 | C | T | 0.021323 | 0.001671 | 2.60E-37 | 0.018343 | 0.001636 | 3.70E-29 |
| rs34994596 | T | C | 0.004793 | 0.001533 | 0.0018 | 0.011151 | 0.001502 | 1.10E-13 |
| rs8038574 | T | C | 0.004853 | 0.001481 | 0.001 | 0.009074 | 0.001451 | 4.00E-10 |
| rs72755233 | G | A | -0.01765 | 0.002225 | 2.10E-15 | -0.00968 | 0.002179 | 8.80E-06 |
| rs2238435 | C | G | -0.01648 | 0.001442 | 2.90E-30 | -0.01415 | 0.001412 | 1.20E-23 |
| rs39674 | C | G | -0.00069 | 0.00153 | 0.65 | -0.00871 | 0.001498 | 6.00E-09 |
| rs55880046 | T | G | 0.029859 | 0.002005 | 3.70E-50 | 0.016863 | 0.001963 | 8.70E-18 |
| rs9922288 | A | G | 0.010499 | 0.001672 | 3.40E-10 | 0.009389 | 0.001637 | 9.80E-09 |
| rs3814883 | C | T | -0.00608 | 0.001406 | 1.50E-05 | -0.01476 | 0.001376 | 8.10E-27 |
| rs4783789 | T | C | 0.009546 | 0.001683 | 1.40E-08 | 0.001654 | 0.001648 | 0.32 |

| | | | | | | | | |
|---|---|---|---|---|---|---|---|---|
| rs56094641 | A | G | -0.04723 | 0.001427 | 3.00E-240 | -0.04652 | 0.001397 | 4.00E-243 |
| rs862320 | C | T | 0.006198 | 0.001426 | 1.40E-05 | 0.014399 | 0.001396 | 6.20E-25 |
| rs12149660 | G | A | 0.003994 | 0.002207 | 0.07 | 0.016218 | 0.002161 | 6.20E-14 |
| rs9673839 | A | G | -0.0032 | 0.001408 | 0.023 | -0.0083 | 0.001379 | 1.70E-09 |
| rs11642090 | T | C | -0.01173 | 0.001459 | 9.20E-16 | -0.00728 | 0.001428 | 3.40E-07 |
| rs11150462 | T | A | 0.003116 | 0.001455 | 0.032 | 0.008444 | 0.001425 | 3.10E-09 |
| rs7206608 | C | G | 4.60E-05 | 0.0015 | 0.98 | -0.00979 | 0.001469 | 2.60E-11 |
| rs4790292 | C | A | 0.007803 | 0.001951 | 6.30E-05 | 0.016329 | 0.001911 | 1.30E-17 |
| rs67603370 | G | A | -0.01586 | 0.002684 | 3.40E-09 | -0.00046 | 0.002629 | 0.86 |
| rs1320251 | C | T | -0.00037 | 0.001412 | 0.79 | 0.012393 | 0.001383 | 3.20E-19 |
| rs1017529 | C | A | -0.00094 | 0.001878 | 0.62 | -0.01062 | 0.00184 | 7.70E-09 |
| rs73982435 | C | T | 0.007541 | 0.001703 | 9.50E-06 | 0.009429 | 0.001668 | 1.60E-08 |
| rs11079849 | C | T | 0.001425 | 0.001495 | 0.34 | 0.012234 | 0.001464 | 6.40E-17 |
| rs78369934 | T | C | -0.00342 | 0.003111 | 0.27 | 0.019936 | 0.003047 | 6.00E-11 |
| rs12941038 | C | T | -0.00956 | 0.001669 | 1.00E-08 | -0.00475 | 0.001634 | 0.0037 |
| rs11150745 | A | G | 0.012163 | 0.001507 | 7.00E-16 | 0.013795 | 0.001476 | 9.20E-21 |
| rs7503580 | C | T | -0.01112 | 0.001933 | 8.70E-09 | -0.00488 | 0.001893 | 0.01 |
| rs1013737 | G | C | -0.01064 | 0.001403 | 3.30E-14 | -0.00585 | 0.001374 | 2.10E-05 |
| rs2083323 | G | A | -0.00502 | 0.001838 | 0.0063 | -0.01041 | 0.0018 | 7.30E-09 |
| rs1788808 | A | G | 0.008793 | 0.001404 | 3.70E-10 | 0.01274 | 0.001375 | 1.90E-20 |
| rs6507054 | T | C | 0.000855 | 0.001422 | 0.55 | -0.00928 | 0.001393 | 2.70E-11 |
| rs7239114 | G | A | -0.01349 | 0.001417 | 1.80E-21 | -0.00644 | 0.001388 | 3.40E-06 |
| rs58084604 | C | T | -0.03316 | 0.001656 | 3.90E-89 | -0.03519 | 0.001622 | 2.60E-104 |
| rs17773370 | G | A | -0.01192 | 0.002819 | 2.30E-05 | -0.01589 | 0.002761 | 8.70E-09 |
| rs57636386 | T | C | 0.023458 | 0.002537 | 2.30E-20 | 0.024702 | 0.002485 | 2.70E-23 |
| rs8089514 | T | A | -0.00733 | 0.00147 | 6.20E-07 | -0.00817 | 0.001439 | 1.40E-08 |
| rs8096658 | C | G | 0.008994 | 0.001419 | 2.30E-10 | 0.004288 | 0.00139 | 0.002 |
| rs72976986 | G | A | 0.000618 | 0.001803 | 0.73 | 0.013851 | 0.001766 | 4.40E-15 |
| rs62621197 | C | T | -0.02349 | 0.003848 | 1.00E-09 | -0.0065 | 0.003769 | 0.085 |
| rs113230003 | G | A | 0.006983 | 0.00161 | 1.40E-05 | 0.012616 | 0.001577 | 1.20E-15 |
| rs10404726 | C | T | 0.00655 | 0.001407 | 3.20E-06 | 0.012293 | 0.001378 | 4.70E-19 |
| rs112253053 | T | A | 0.010125 | 0.001909 | 1.10E-07 | 0.013771 | 0.00187 | 1.80E-13 |
| rs12462975 | G | A | -0.00516 | 0.001501 | 0.00058 | -0.01148 | 0.00147 | 5.90E-15 |
| rs73026723 | C | T | 0.00949 | 0.00194 | 1.00E-06 | 0.014068 | 0.0019 | 1.30E-13 |
| rs429358 | T | C | -0.00346 | 0.001941 | 0.075 | 0.016069 | 0.001902 | 2.90E-17 |
| rs12971645 | G | A | 0.00037 | 0.001576 | 0.81 | 0.008594 | 0.001543 | 2.60E-08 |
| rs10423928 | T | A | 0.011763 | 0.001769 | 2.90E-11 | 0.020764 | 0.001732 | 4.20E-33 |
| rs6075658 | T | C | 0.00244 | 0.001404 | 0.082 | 0.008489 | 0.001376 | 6.80E-10 |
| rs2206925 | T | C | -0.01062 | 0.00146 | 3.40E-13 | -0.01176 | 0.00143 | 2.00E-16 |
| rs16996644 | C | G | -0.01935 | 0.00211 | 4.80E-20 | -0.0106 | 0.002068 | 2.90E-07 |
| rs8124896 | T | C | -0.00576 | 0.002331 | 0.013 | -0.01317 | 0.002284 | 8.10E-09 |
| rs6050446 | A | G | -0.00668 | 0.003975 | 0.093 | -0.02642 | 0.003895 | 1.20E-11 |
| rs4911382 | C | T | -0.00223 | 0.001426 | 0.12 | -0.00832 | 0.001397 | 2.60E-09 |
| rs6029180 | A | G | -0.00454 | 0.001509 | 0.0026 | -0.00819 | 0.001479 | 3.10E-08 |
| rs6030803 | T | C | 0.004327 | 0.002116 | 0.041 | 0.012927 | 0.002074 | 4.50E-10 |

| | | | | | | | | |
|---|---|---|---|---|---|---|---|---|
| rs2425856 | A | G | 0.003461 | 0.00141 | 0.014 | 0.008422 | 0.001381 | 1.10E-09 |
| rs66460909 | G | A | 0.005294 | 0.001785 | 0.003 | 0.015693 | 0.001749 | 2.90E-19 |
| rs2207894 | C | T | 0.014344 | 0.001781 | 8.00E-16 | 0.007601 | 0.001745 | 1.30E-05 |
| rs13047416 | C | G | 0.012376 | 0.001451 | 1.40E-17 | 0.008657 | 0.001422 | 1.10E-09 |
| rs8134638 | T | C | 0.000773 | 0.001449 | 0.59 | -0.0078 | 0.00142 | 4.00E-08 |
| rs403694 | C | T | -0.00185 | 0.001411 | 0.19 | -0.01218 | 0.001383 | 1.30E-18 |
| rs78907487 | A | C | -0.01236 | 0.001977 | 4.10E-10 | -0.00677 | 0.001937 | 0.00047 |
| rs12484438 | T | C | 0.011798 | 0.001481 | 1.60E-15 | 0.012887 | 0.001451 | 6.70E-19 |
| rs738140 | A | G | 0.000927 | 0.001518 | 0.54 | 0.00886 | 0.001488 | 2.60E-09 |
| rs9615723 | C | T | 0.002284 | 0.001431 | 0.11 | 0.007715 | 0.001403 | 3.80E-08 |
| rs34778589 | A | C | -0.0103 | 0.002551 | 5.40E-05 | -0.01434 | 0.0025 | 9.80E-09 |
| Men | | | | | | | | |
| SNP | EA | OA | beta_early | se_early | pval_early | beta_adult | se_adult | pval_adult |
| rs3762444 | C | T | 0.005117 | 0.002078 | 0.014 | 0.012328 | 0.00194 | 2.10E-10 |
| rs1284373 | C | T | 0.00329 | 0.002566 | 0.2 | 0.013881 | 0.002396 | 6.90E-09 |
| rs34052145 | A | G | 0.000152 | 0.00216 | 0.94 | -0.01144 | 0.002017 | 1.40E-08 |
| rs1167311 | G | A | 0.007996 | 0.00222 | 0.00032 | 0.012003 | 0.002073 | 7.00E-09 |
| rs12140153 | G | T | 0.023436 | 0.003586 | 6.40E-11 | 0.023817 | 0.003348 | 1.10E-12 |
| rs11208779 | G | C | -0.00411 | 0.002057 | 0.046 | -0.01154 | 0.00192 | 1.90E-09 |
| rs61765650 | A | G | 0.022662 | 0.002608 | 3.70E-18 | 0.021859 | 0.002435 | 2.80E-19 |
| rs4650277 | A | G | 0.022507 | 0.002068 | 1.40E-27 | 0.007296 | 0.00193 | 0.00016 |
| rs2181375 | A | G | -0.00552 | 0.00209 | 0.0082 | -0.01233 | 0.001951 | 2.60E-10 |
| rs7550711 | C | T | -0.05069 | 0.006492 | 5.80E-15 | -0.04405 | 0.006061 | 3.60E-13 |
| rs61813324 | C | T | -0.00516 | 0.003035 | 0.089 | -0.01581 | 0.002833 | 2.40E-08 |
| rs539515 | A | C | -0.03876 | 0.002539 | 1.30E-52 | -0.02195 | 0.00237 | 2.00E-20 |
| rs1772143 | T | A | 0.014627 | 0.002084 | 2.20E-12 | 0.008697 | 0.001945 | 7.80E-06 |
| rs2125232 | C | T | 0.00216 | 0.002205 | 0.33 | 0.012243 | 0.002058 | 2.70E-09 |
| rs77165542 | C | T | 0.072978 | 0.005625 | 1.70E-38 | 0.049509 | 0.005251 | 4.20E-21 |
| rs6749422 | C | G | -0.0313 | 0.002051 | 1.40E-52 | -0.01524 | 1.91E-03 | 1.70E-15 |
| rs1861410 | C | T | 0.010484 | 0.002073 | 4.20E-07 | 0.013057 | 0.001935 | 1.50E-11 |
| rs3552 | G | A | -0.00616 | 0.002059 | 0.0028 | -0.01152 | 0.001922 | 2.00E-09 |
| rs396354 | T | C | -0.00047 | 0.002279 | 0.83 | 0.011772 | 0.002127 | 3.10E-08 |
| rs6753397 | C | T | -0.00305 | 0.002316 | 0.19 | -0.01198 | 0.002162 | 3.00E-08 |
| rs13405033 | C | T | -0.00429 | 0.002848 | 0.13 | -0.01832 | 0.002659 | 5.60E-12 |
| rs10496885 | G | A | 0.016558 | 0.002636 | 3.30E-10 | 0.00783 | 0.002461 | 0.0015 |
| rs1451077 | G | A | 0.004218 | 0.002085 | 0.043 | 0.012968 | 0.001946 | 2.70E-11 |
| rs115319174 | G | C | -0.04345 | 0.004443 | 1.40E-22 | -0.01618 | 0.004148 | 9.50E-05 |
| rs6436661 | C | T | -0.00542 | 0.003545 | 0.13 | -0.01842 | 0.00331 | 2.60E-08 |
| rs7619139 | T | A | -0.00907 | 0.002093 | 1.50E-05 | -0.01196 | 0.001953 | 9.20E-10 |
| rs2526389 | C | T | -0.01011 | 0.002077 | 1.10E-06 | -0.01902 | 0.001938 | 1.00E-22 |
| rs11708540 | G | A | 0.002594 | 0.002837 | 0.36 | -0.01497 | 0.002648 | 1.60E-08 |
| rs55782528 | C | A | 0.008063 | 0.002148 | 0.00017 | 0.012902 | 0.002005 | 1.20E-10 |
| rs13066686 | C | A | 0.000651 | 0.002098 | 0.76 | 0.013265 | 0.001958 | 1.20E-11 |
| rs2918217 | C | T | -0.00696 | 0.00298 | 0.019 | -0.01822 | 0.002781 | 5.70E-11 |
| rs73875019 | T | A | 0.009583 | 0.003228 | 0.003 | 0.019461 | 0.003013 | 1.10E-10 |

| | | | | | | | | |
|---|---|---|---|---|---|---|---|---|
| rs1320903 | G | A | -0.00867 | 0.002206 | 8.60E-05 | -0.01588 | 0.002059 | 1.20E-14 |
| rs61789562 | T | C | 0.006658 | 0.003178 | 0.036 | 0.016787 | 0.002966 | 1.50E-08 |
| rs1568488 | G | C | -0.00525 | 0.002107 | 0.013 | -0.01169 | 0.001966 | 2.70E-09 |
| rs8192675 | T | C | -0.00343 | 0.002266 | 0.13 | -0.01351 | 2.11E-03 | 1.70E-10 |
| rs55742087 | C | T | 0.013365 | 0.002647 | 4.40E-07 | 0.018996 | 0.002471 | 1.50E-14 |
| rs10938398 | G | A | -0.02074 | 0.002076 | 1.70E-23 | -0.02143 | 0.00194 | 2.30E-28 |
| rs2537860 | A | C | -0.00104 | 0.002093 | 0.62 | 0.011371 | 0.001955 | 6.00E-09 |
| rs13107325 | C | T | -0.02266 | 0.003879 | 5.20E-09 | -0.02942 | 0.003623 | 4.70E-16 |
| rs1296328 | A | C | 0.006762 | 0.002078 | 0.0011 | 0.013313 | 0.001941 | 7.00E-12 |
| rs3212519 | A | G | 0.02244 | 0.004047 | 2.90E-08 | -0.00368 | 0.003781 | 0.33 |
| rs75577466 | G | C | 0.016931 | 0.0027 | 3.60E-10 | 0.003922 | 0.002523 | 0.12 |
| rs2307111 | T | C | 0.009023 | 0.002103 | 1.80E-05 | 0.012999 | 0.001965 | 3.70E-11 |
| rs7703782 | T | A | -0.01218 | 0.003016 | 5.30E-05 | -0.0235 | 0.002818 | 7.40E-17 |
| rs4958361 | C | G | 0.012105 | 0.002054 | 3.80E-09 | 0.007696 | 0.001919 | 6.10E-05 |
| rs2591496 | G | A | -0.0063 | 0.002312 | 0.0064 | -0.01289 | 0.002161 | 2.40E-09 |
| rs9379829 | C | T | -0.00172 | 0.002486 | 0.49 | 0.01286 | 0.002324 | 3.10E-08 |
| rs9277992 | G | A | 0.00026 | 0.002628 | 0.92 | -0.01603 | 0.002457 | 6.80E-11 |
| rs9469899 | G | A | -0.00559 | 0.00214 | 0.009 | -0.013 | 0.002 | 8.20E-11 |
| rs3798519 | A | C | -0.02178 | 0.002675 | 3.90E-16 | -0.0236 | 0.0025 | 3.80E-21 |
| rs1775255 | G | T | -0.0133 | 0.00206 | 1.10E-10 | -0.00925 | 0.001925 | 1.60E-06 |
| rs115597956 | G | A | -0.02993 | 0.004519 | 3.50E-11 | -0.00957 | 0.004224 | 0.023 |
| rs74621225 | A | G | -0.00034 | 0.003024 | 0.91 | 0.016257 | 0.002827 | 8.90E-09 |
| rs9320823 | T | C | 0.003715 | 0.002099 | 0.077 | -0.01327 | 0.001962 | 1.30E-11 |
| rs2693560 | A | G | 0.011595 | 0.002126 | 4.90E-08 | 0.004703 | 0.001987 | 0.018 |
| rs1452991 | G | A | -0.0121 | 0.002139 | 1.50E-08 | -0.00359 | 0.002 | 0.073 |
| rs9478671 | A | G | -0.00827 | 0.002527 | 0.0011 | -0.01399 | 0.002362 | 3.10E-09 |
| rs34714518 | G | A | 0.003403 | 0.003126 | 0.28 | 0.016269 | 0.002922 | 2.60E-08 |
| rs16120 | A | G | 0.01167 | 0.002058 | 1.40E-08 | 0.005929 | 0.001923 | 0.0021 |
| rs62457529 | A | G | -0.00738 | 0.003294 | 0.025 | -0.01884 | 0.003078 | 9.40E-10 |
| rs6962280 | A | G | -0.00546 | 0.002074 | 0.0084 | -0.01214 | 0.001938 | 3.80E-10 |
| rs17145600 | C | T | -0.00693 | 0.004676 | 0.14 | -0.0245 | 0.00437 | 2.00E-08 |
| rs12538826 | T | C | -0.00028 | 0.003216 | 0.93 | 0.016863 | 0.003005 | 2.00E-08 |
| rs10236214 | C | T | -0.00462 | 0.002153 | 0.032 | -0.01128 | 0.002012 | 2.10E-08 |
| rs4840941 | A | G | -0.01132 | 0.002081 | 5.30E-08 | -0.01367 | 0.001945 | 2.10E-12 |
| rs7827210 | G | A | -0.00057 | 0.002104 | 0.79 | -0.0111 | 0.001966 | 1.70E-08 |
| rs10504620 | T | C | -0.01516 | 0.00224 | 1.30E-11 | -0.0048 | 0.002093 | 0.022 |
| rs72674843 | T | C | 0.003086 | 0.002409 | 0.2 | 0.014987 | 0.002252 | 2.80E-11 |
| rs800526 | A | C | -0.00662 | 0.002464 | 0.0072 | -0.0142 | 0.002303 | 7.10E-10 |
| rs1412239 | C | G | -0.00483 | 0.00219 | 0.027 | -0.01546 | 0.002047 | 4.20E-14 |
| rs11790060 | T | C | -0.01283 | 0.002183 | 4.20E-09 | -0.00016 | 0.00204 | 0.94 |
| rs10828247 | A | G | 0.002681 | 0.002163 | 0.22 | -0.01124 | 0.002022 | 2.70E-08 |
| rs10824218 | A | T | 0.005328 | 0.002085 | 0.011 | 0.010646 | 0.001949 | 4.70E-08 |
| rs41310284 | C | A | 0.024196 | 0.003437 | 1.90E-12 | 0.018066 | 0.003213 | 1.90E-08 |
| rs4962671 | T | C | 0.00038 | 0.002067 | 0.85 | 0.010738 | 0.001932 | 2.70E-08 |
| rs72867447 | C | G | -0.00392 | 0.002072 | 0.059 | -0.01111 | 0.001936 | 9.50E-09 |

| | | | | | | | | |
|---|---|---|---|---|---|---|---|---|
| rs6265 | C | T | 0.013661 | 0.002623 | 1.90E-07 | 0.028497 | 0.002451 | 3.00E-31 |
| rs1222216 | C | T | 0.002611 | 0.002449 | 0.29 | 0.013622 | 0.002289 | 2.60E-09 |
| rs4755726 | T | G | -0.00133 | 0.002221 | 0.55 | 0.013849 | 0.002076 | 2.50E-11 |
| rs12798028 | C | T | -0.01362 | 0.002082 | 6.20E-11 | -0.01507 | 0.001946 | 9.60E-15 |
| rs7940691 | C | T | 0.011448 | 0.002134 | 8.10E-08 | 0.013405 | 0.001994 | 1.80E-11 |
| rs10896348 | T | C | 0.013309 | 0.002288 | 6.00E-09 | 0.007009 | 0.002138 | 0.001 |
| rs10898317 | C | T | 0.003485 | 0.002057 | 0.09 | 0.010653 | 0.001922 | 3.00E-08 |
| rs11218734 | A | G | -0.01293 | 0.002347 | 3.60E-08 | -0.00518 | 0.002193 | 0.018 |
| rs55726687 | G | A | -0.01203 | 0.002523 | 1.80E-06 | -0.01289 | 0.002359 | 4.70E-08 |
| rs76895963 | T | G | -0.01649 | 0.007934 | 0.038 | -0.04219 | 0.007417 | 1.30E-08 |
| rs7978659 | G | T | -0.01334 | 0.002163 | 7.00E-10 | -0.00359 | 0.002022 | 0.076 |
| rs7132908 | G | A | -0.03173 | 0.002109 | 3.60E-51 | -0.01901 | 0.001971 | 5.30E-22 |
| rs7306710 | T | C | 0.012402 | 0.002069 | 2.00E-09 | 0.003971 | 0.001934 | 0.04 |
| rs7308188 | T | C | 0.010013 | 0.002354 | 2.10E-05 | 0.013666 | 0.002201 | 5.30E-10 |
| rs6490030 | C | A | -0.00641 | 0.00213 | 0.0026 | -0.01095 | 0.001991 | 3.80E-08 |
| rs147730268 | G | T | -0.00359 | 0.003731 | 0.34 | 0.021582 | 0.003488 | 6.10E-10 |
| rs7316962 | A | G | 0.01291 | 0.002064 | 4.00E-10 | 0.008924 | 0.001929 | 3.70E-06 |
| rs61954177 | G | C | -0.01089 | 0.002195 | 7.00E-07 | -0.01152 | 0.002052 | 2.00E-08 |
| rs9568868 | G | T | -0.02484 | 0.003081 | 7.60E-16 | -0.02052 | 0.00288 | 1.00E-12 |
| rs1576655 | A | C | -0.01177 | 0.002137 | 3.70E-08 | -0.01056 | 0.001998 | 1.20E-07 |
| rs7983454 | T | C | 0.004195 | 0.002056 | 0.041 | 0.010963 | 0.001922 | 1.20E-08 |
| rs9522279 | C | T | -0.00091 | 0.002083 | 0.66 | -0.01269 | 0.001947 | 7.30E-11 |
| rs10132280 | C | A | 0.009174 | 0.002247 | 4.50E-05 | 0.016259 | 0.002101 | 9.90E-15 |
| rs10131761 | T | A | 0.003587 | 0.002632 | 0.17 | 0.013511 | 0.00246 | 4.00E-08 |
| rs4898556 | A | C | 0.002789 | 0.002054 | 0.17 | 0.012895 | 0.00192 | 1.90E-11 |
| rs217669 | T | C | -0.00034 | 0.002311 | 0.88 | -0.01357 | 0.00216 | 3.40E-10 |
| rs8008910 | G | A | -0.01042 | 0.002476 | 2.60E-05 | -0.01697 | 0.002314 | 2.30E-13 |
| rs1887197 | C | T | -0.00451 | 0.002185 | 0.039 | -0.01365 | 0.002043 | 2.30E-11 |
| rs61992671 | A | G | 0.010267 | 0.002148 | 1.80E-06 | 0.010958 | 0.002008 | 4.90E-08 |
| rs3784710 | T | C | 0.019152 | 0.002448 | 5.10E-15 | 0.016403 | 0.002289 | 7.80E-13 |
| rs11631651 | A | C | 0.008112 | 0.003961 | 0.041 | 0.022476 | 0.003705 | 1.30E-09 |
| rs1990573 | A | G | -0.00368 | 0.002228 | 0.098 | -0.01255 | 0.002082 | 1.70E-09 |
| rs7190603 | T | C | 0.029545 | 0.002947 | 1.20E-23 | 0.015601 | 0.002753 | 1.50E-08 |
| rs62048402 | G | A | -0.04616 | 0.002089 | 3.40E-108 | -0.04733 | 0.001952 | 6.90E-130 |
| rs12923231 | C | T | 0.001105 | 0.002092 | 0.6 | 0.014188 | 0.001954 | 3.90E-13 |
| rs12149660 | G | A | 0.004733 | 0.003236 | 0.14 | 0.02009 | 0.003023 | 3.00E-11 |
| rs3923783 | C | A | 0.005273 | 0.002646 | 0.046 | 0.015984 | 0.002474 | 1.00E-10 |
| rs9901404 | A | G | -0.00158 | 0.002232 | 0.48 | 0.015503 | 0.002087 | 1.10E-13 |
| rs12941009 | C | T | 0.009935 | 0.002113 | 2.60E-06 | 0.011248 | 0.001975 | 1.20E-08 |
| rs17637472 | G | A | -0.01237 | 0.002118 | 5.20E-09 | -0.00503 | 0.00198 | 0.011 |
| rs11150745 | A | G | 0.011506 | 0.002206 | 1.80E-07 | 0.01346 | 0.002063 | 6.80E-11 |
| rs1652376 | G | T | 0.007491 | 0.002061 | 0.00028 | 0.014672 | 0.001927 | 2.60E-14 |
| rs7232171 | G | T | -8.73E-05 | 0.002081 | 0.97 | -0.01088 | 0.001946 | 2.20E-08 |
| rs7239114 | G | A | -0.01276 | 0.002077 | 8.10E-10 | -0.00393 | 0.001942 | 0.043 |
| rs7240682 | C | G | -0.03027 | 0.002456 | 6.30E-35 | -0.0311 | 0.002296 | 8.70E-42 |

| | | | | | | | | |
|---|---|---|---|---|---|---|---|---|
| rs8112818 | A | G | 0.00502 | 0.002104 | 0.017 | 0.011532 | 0.001968 | 4.60E-09 |
| rs3810291 | G | A | -0.01526 | 0.002188 | 3.10E-12 | -0.01596 | 0.002047 | 6.30E-15 |
| rs6054427 | G | A | -0.01217 | 0.002127 | 1.10E-08 | -0.01357 | 0.001989 | 8.90E-12 |
| rs73898513 | C | T | -0.01877 | 0.003182 | 3.70E-09 | -0.0112 | 0.002976 | 0.00017 |
| rs6096886 | A | G | 0.006625 | 0.002617 | 0.011 | 0.015501 | 0.002447 | 2.40E-10 |
| rs9977825 | T | C | 0.005075 | 0.002144 | 0.018 | 0.011679 | 0.002006 | 5.80E-09 |
| rs17421586 | T | A | 0.010896 | 0.002168 | 5.00E-07 | 0.011985 | 0.002028 | 3.40E-09 |
| Women | | | | | | | | |
| SNP | EA | OA | beta_early | se_early | pval_early | beta_adult | se_adult | pval_adult |
| rs74892851 | C | A | 0.000122 | 0.002026 | 0.95 | 0.011586 | 0.00206 | 1.90E-08 |
| rs78886584 | A | G | -0.00212 | 0.001958 | 0.28 | -0.01327 | 0.00199 | 2.60E-11 |
| rs212540 | C | T | 0.012688 | 0.002001 | 2.30E-10 | 0.001562 | 0.002034 | 0.44 |
| rs12144626 | T | C | 0.008272 | 0.001964 | 2.50E-05 | 0.011716 | 0.001997 | 4.40E-09 |
| rs1494461 | C | T | 0.001819 | 0.002068 | 0.38 | 0.012736 | 0.002102 | 1.40E-09 |
| rs582220 | A | G | -0.01148 | 0.001958 | 4.50E-09 | -0.00797 | 0.001991 | 6.20E-05 |
| rs2767486 | A | G | -0.02032 | 0.002411 | 3.50E-17 | -0.00124 | 0.00245 | 0.61 |
| rs7522014 | A | G | 0.011509 | 0.0021 | 4.20E-08 | 0.00772 | 0.002134 | 3.00E-04 |
| rs12042908 | A | G | 0.031709 | 0.001951 | 2.10E-59 | 0.013067 | 0.001983 | 4.40E-11 |
| rs34517439 | C | A | -0.01249 | 0.002984 | 2.80E-05 | -0.02525 | 0.003033 | 8.40E-17 |
| rs10922911 | C | T | 0.000466 | 0.002004 | 0.82 | -0.01383 | 0.002037 | 1.10E-11 |
| rs75641275 | A | C | 0.002059 | 0.002765 | 0.46 | -0.01849 | 0.00281 | 4.70E-11 |
| rs41279738 | T | G | -0.04643 | 0.006081 | 2.20E-14 | -0.04097 | 0.006181 | 3.40E-11 |
| rs61813324 | C | T | -0.00474 | 0.002867 | 0.098 | -0.02089 | 0.002914 | 7.60E-13 |
| rs543874 | A | G | -0.05471 | 0.002395 | 1.60E-115 | -0.03753 | 0.002434 | 1.20E-53 |
| rs10798139 | C | T | -0.01371 | 0.002355 | 5.90E-09 | -0.00876 | 0.002394 | 0.00025 |
| rs815163 | T | C | 0.009194 | 0.001947 | 2.30E-06 | 0.01322 | 0.001979 | 2.40E-11 |
| rs2678204 | T | G | -0.00556 | 0.00204 | 0.0065 | -0.01336 | 0.002073 | 1.20E-10 |
| rs2994320 | A | G | -0.00027 | 0.002448 | 0.91 | 0.016622 | 0.002488 | 2.40E-11 |
| rs62106258 | T | C | 0.087151 | 0.004482 | 3.40E-84 | 0.067317 | 0.004555 | 2.00E-49 |
| rs12992672 | G | A | -0.04416 | 0.00256 | 1.10E-66 | -0.0361 | 0.002601 | 8.40E-44 |
| rs6738433 | G | C | -0.04047 | 0.001938 | 7.60E-97 | -0.02355 | 0.001969 | 5.90E-33 |
| rs146910503 | G | A | 0.046977 | 0.006936 | 1.30E-11 | 0.022138 | 0.007048 | 0.0017 |
| rs34606703 | G | A | 0.005027 | 0.002023 | 0.013 | 0.011743 | 0.002056 | 1.10E-08 |
| rs13420048 | C | A | 0.000699 | 0.002012 | 0.73 | 0.011999 | 0.002044 | 4.40E-09 |
| rs4671328 | T | G | 0.001651 | 0.001956 | 0.4 | 0.013654 | 0.001988 | 6.50E-12 |
| rs13416992 | A | C | 0.004401 | 0.001978 | 0.026 | 0.014785 | 0.00201 | 1.90E-13 |
| rs1446725 | T | G | 0.013563 | 0.001962 | 4.80E-12 | 0.008481 | 0.001994 | 2.10E-05 |
| rs11691869 | C | A | 0.001026 | 0.002015 | 0.61 | 0.015537 | 0.002048 | 3.30E-14 |
| rs7602120 | C | T | -0.00271 | 0.001946 | 0.16 | -0.01341 | 0.001977 | 1.20E-11 |
| rs55959207 | A | C | 0.010936 | 0.001949 | 2.00E-08 | -0.00286 | 0.00198 | 0.15 |
| rs17464221 | C | T | 0.012296 | 0.002137 | 8.70E-09 | 0.002653 | 0.002171 | 0.22 |
| rs115319174 | G | C | -0.04113 | 0.004202 | 1.30E-22 | -0.00738 | 0.00427 | 0.084 |
| rs2433733 | G | A | 0.001038 | 0.002068 | 0.62 | 0.011736 | 0.002101 | 2.30E-08 |
| rs754635 | C | G | -0.01662 | 0.003046 | 4.80E-08 | -0.01603 | 0.003096 | 2.30E-07 |
| rs9843653 | T | C | -0.00397 | 0.001936 | 0.04 | -0.01758 | 0.001968 | 4.20E-19 |

| | | | | | | | | |
|---|---|---|---|---|---|---|---|---|
| rs79569013 | T | G | 0.020058 | 0.002689 | 8.60E-14 | 0.014564 | 0.002733 | 9.90E-08 |
| rs6774533 | C | T | -0.00562 | 0.002159 | 0.0092 | -0.01218 | 0.002195 | 2.80E-08 |
| rs818219 | T | C | -0.01337 | 0.001946 | 6.50E-12 | -0.00391 | 0.001978 | 0.048 |
| rs76152047 | A | G | -0.02505 | 0.0039 | 1.30E-10 | -0.02331 | 0.003965 | 4.10E-09 |
| rs529200 | A | G | -0.00302 | 0.00194 | 0.12 | -0.0116 | 0.001971 | 4.00E-09 |
| rs73052033 | T | C | 0.006666 | 0.002495 | 0.0076 | 0.017216 | 0.002537 | 1.10E-11 |
| rs34811474 | G | A | 0.008539 | 0.002296 | 2.00E-04 | 0.019703 | 0.002335 | 3.20E-17 |
| rs73213484 | A | T | 0.002482 | 0.002781 | 0.37 | 0.018902 | 0.002828 | 2.30E-11 |
| rs12641981 | C | T | -0.02326 | 0.001955 | 1.20E-32 | -0.01681 | 0.001988 | 2.80E-17 |
| rs148712344 | G | T | 0.007041 | 0.005019 | 0.16 | 0.029824 | 0.005104 | 5.10E-09 |
| rs1603179 | A | C | -0.0036 | 0.002025 | 0.076 | 0.011425 | 0.002059 | 2.90E-08 |
| rs11098965 | C | T | 0.000651 | 0.00222 | 0.77 | 0.013621 | 0.002258 | 1.60E-09 |
| rs1349641 | T | G | 0.01249 | 0.001982 | 2.90E-10 | 0.00239 | 0.002015 | 0.24 |
| rs7377083 | C | A | -0.01545 | 0.00197 | 4.40E-15 | -0.01084 | 0.002003 | 6.20E-08 |
| rs769668 | T | C | 0.000197 | 0.002043 | 0.92 | 0.013957 | 0.002077 | 1.80E-11 |
| rs35390852 | G | A | -0.00205 | 0.002943 | 0.49 | -0.01669 | 0.002993 | 2.40E-08 |
| rs828550 | C | T | -0.00156 | 0.002039 | 0.44 | -0.01173 | 0.002073 | 1.50E-08 |
| rs10514963 | G | A | -0.0028 | 0.001934 | 0.15 | -0.01372 | 0.001967 | 3.00E-12 |
| rs10050620 | C | T | 0.015942 | 0.002069 | 1.30E-14 | 0.009684 | 0.002103 | 4.10E-06 |
| rs34341 | A | T | -0.00743 | 0.001958 | 0.00015 | -0.01998 | 0.001991 | 1.10E-23 |
| rs7442885 | C | G | 0.011526 | 0.002364 | 1.10E-06 | 0.016992 | 0.002403 | 1.50E-12 |
| rs1477290 | T | C | -0.00588 | 0.002841 | 0.039 | -0.01749 | 0.002889 | 1.40E-09 |
| rs4235642 | A | G | 0.012363 | 0.001996 | 5.80E-10 | -0.0059 | 0.002029 | 0.0037 |
| rs288187 | C | T | 0.003295 | 0.002577 | 0.2 | 0.019809 | 0.00262 | 4.00E-14 |
| rs1366334 | C | G | 0.006107 | 0.00213 | 0.0041 | 0.011828 | 0.002165 | 4.70E-08 |
| rs13174863 | A | G | -0.00348 | 0.002741 | 0.2 | -0.01872 | 0.002787 | 1.80E-11 |
| rs815610 | C | G | 0.015263 | 0.001948 | 4.70E-15 | 0.008674 | 0.001981 | 1.20E-05 |
| rs11134679 | A | G | -0.00842 | 0.002093 | 5.70E-05 | -0.01307 | 0.002128 | 8.00E-10 |
| rs12214497 | G | T | 0.01355 | 0.002033 | 2.70E-11 | 0.006348 | 0.002068 | 0.0021 |
| rs3131934 | T | C | -0.0227 | 0.002581 | 1.40E-18 | -0.00853 | 0.002625 | 0.0012 |
| rs141127771 | G | A | -0.01563 | 0.002601 | 1.80E-09 | -0.00887 | 0.002645 | 8.00E-04 |
| rs34298980 | T | C | 0.007357 | 0.002038 | 0.00031 | 0.014147 | 0.002073 | 8.80E-12 |
| rs2206277 | C | T | -0.02688 | 0.00252 | 1.40E-26 | -0.02533 | 0.002563 | 4.90E-23 |
| rs1775255 | G | T | -0.01366 | 0.00194 | 1.90E-12 | -0.0096 | 0.001973 | 1.20E-06 |
| rs34260097 | T | G | -0.02522 | 0.002317 | 1.30E-27 | -0.00362 | 0.002357 | 0.12 |
| rs9387640 | C | T | 0.003055 | 0.002011 | 0.13 | 0.011752 | 0.002045 | 9.20E-09 |
| rs796915 | C | G | -0.01602 | 0.002104 | 2.60E-14 | -0.0086 | 0.00214 | 5.80E-05 |
| rs62425122 | G | A | 0.012421 | 0.002118 | 4.50E-09 | 0.003302 | 0.002154 | 0.13 |
| rs73046311 | C | G | 0.001704 | 0.002646 | 0.52 | 0.015674 | 0.002691 | 5.70E-09 |
| rs983949 | T | G | -0.01301 | 0.002146 | 1.30E-09 | -0.00559 | 0.002182 | 0.01 |
| rs2866720 | C | T | -0.0016 | 0.002001 | 0.42 | -0.01179 | 0.002036 | 7.00E-09 |
| rs7808296 | C | T | -0.01168 | 0.002082 | 2.00E-08 | -0.00317 | 0.002117 | 0.13 |
| rs12375196 | C | A | -0.01016 | 0.001972 | 2.60E-07 | -0.0117 | 0.002005 | 5.50E-09 |
| rs1840661 | T | A | 0.003183 | 0.001965 | 0.11 | -0.01216 | 0.001999 | 1.20E-09 |
| rs6979832 | A | G | -0.01135 | 0.001948 | 5.70E-09 | -0.00309 | 0.001981 | 0.12 |

| | | | | | | | | |
|---|---|---|---|---|---|---|---|---|
| rs13233916 | C | G | 0.021026 | 0.003394 | 5.80E-10 | 0.014913 | 0.003452 | 1.60E-05 |
| rs11250094 | G | C | 0.009432 | 0.001948 | 1.30E-06 | 0.012082 | 0.001982 | 1.10E-09 |
| rs351776 | A | C | -0.01166 | 0.001943 | 2.00E-09 | -0.0039 | 0.001977 | 0.049 |
| rs62515439 | C | T | 0.012691 | 0.002007 | 2.50E-10 | 0.003678 | 0.002041 | 0.072 |
| rs13254613 | A | C | -0.01664 | 0.002035 | 3.00E-16 | 0.008559 | 0.00207 | 3.60E-05 |
| rs10957605 | C | T | 0.000868 | 0.002085 | 0.68 | 0.015787 | 0.002121 | 9.80E-14 |
| rs2126474 | G | T | 0.019629 | 0.001967 | 1.90E-23 | 0.009625 | 0.002001 | 1.50E-06 |
| rs17716502 | C | T | 0.002814 | 0.002417 | 0.24 | 0.017884 | 0.002459 | 3.50E-13 |
| rs13292699 | A | C | -0.00266 | 0.001956 | 0.17 | 0.015023 | 0.00199 | 4.30E-14 |
| rs17770336 | C | T | -0.00289 | 0.002067 | 0.16 | -0.01603 | 0.002103 | 2.50E-14 |
| rs10821163 | G | C | -0.01843 | 0.002053 | 2.80E-19 | 0.002148 | 0.002089 | 0.3 |
| rs7047694 | G | A | -0.00851 | 0.002076 | 4.10E-05 | -0.01245 | 0.002112 | 3.70E-09 |
| rs957512 | T | C | 0.011541 | 0.002061 | 2.20E-08 | 0.008541 | 0.002097 | 4.60E-05 |
| rs2275241 | G | A | -0.0138 | 0.002002 | 5.50E-12 | -0.00674 | 0.002037 | 0.00094 |
| rs3003578 | C | T | 0.002253 | 0.001962 | 0.25 | 0.011246 | 0.001997 | 1.80E-08 |
| rs7084503 | T | C | 0.015422 | 0.001942 | 2.00E-15 | 0.005656 | 0.001975 | 0.0042 |
| rs1270799 | T | G | -0.00276 | 0.002106 | 0.19 | -0.01653 | 0.002142 | 1.20E-14 |
| rs113585475 | C | T | 0.001865 | 0.003267 | 0.57 | 0.022 | 0.003323 | 3.60E-11 |
| rs3125326 | A | C | -0.00442 | 0.002004 | 0.027 | -0.01113 | 0.002038 | 4.70E-08 |
| rs7090758 | T | C | -0.00572 | 0.00194 | 0.0032 | -0.01293 | 0.001973 | 5.70E-11 |
| rs11000942 | G | A | -0.00641 | 0.002934 | 0.029 | -0.01638 | 0.002983 | 4.00E-08 |
| rs76971642 | T | C | -0.03092 | 0.004586 | 1.60E-11 | -0.02338 | 0.004664 | 5.30E-07 |
| rs10510025 | C | T | 0.000228 | 0.00225 | 0.92 | -0.01326 | 0.002288 | 6.90E-09 |
| rs11146233 | G | A | 0.002245 | 0.001952 | 0.25 | 0.011397 | 0.001985 | 9.40E-09 |
| rs7950166 | C | T | -0.00699 | 0.002022 | 0.00054 | -0.01435 | 0.002057 | 3.00E-12 |
| rs1013402 | A | G | -0.01408 | 0.002072 | 1.10E-11 | -0.02127 | 0.002108 | 6.00E-24 |
| rs661878 | A | G | 0.018891 | 0.00285 | 3.40E-11 | 0.006176 | 0.002898 | 0.033 |
| rs11039307 | C | T | -0.01564 | 0.001964 | 1.70E-15 | -0.01543 | 0.001998 | 1.10E-14 |
| rs34292685 | C | T | 0.004459 | 0.002626 | 0.09 | 0.017997 | 0.002671 | 1.60E-11 |
| rs10896012 | T | C | -0.00869 | 0.002353 | 0.00022 | -0.01583 | 0.002394 | 3.70E-11 |
| rs678653 | C | G | -0.01224 | 0.002018 | 1.30E-09 | -0.00964 | 0.002053 | 2.70E-06 |
| rs11215403 | G | A | 0.016158 | 0.002254 | 7.60E-13 | 0.006712 | 0.002293 | 0.0034 |
| rs11218510 | G | A | 0.00291 | 0.00198 | 0.14 | 0.011176 | 0.002014 | 2.90E-08 |
| rs2512884 | C | A | -0.01008 | 0.001942 | 2.10E-07 | -0.01241 | 0.001975 | 3.30E-10 |
| rs11223204 | A | G | -0.00488 | 0.001955 | 0.013 | -0.01135 | 0.001989 | 1.10E-08 |
| rs11611246 | G | T | -0.01658 | 0.002364 | 2.30E-12 | -0.01396 | 0.002406 | 6.50E-09 |
| rs2187642 | A | C | -0.01187 | 0.001994 | 2.60E-09 | -0.00051 | 0.00203 | 0.8 |
| rs7976757 | T | C | -0.00322 | 0.00257 | 0.21 | -0.01521 | 0.002616 | 6.10E-09 |
| rs10876457 | G | A | 0.012849 | 0.002324 | 3.20E-08 | 0.006149 | 0.002365 | 0.0093 |
| rs10783302 | G | T | -0.01322 | 0.00201 | 4.70E-11 | -0.00607 | 0.002045 | 0.003 |
| rs7132908 | G | A | -0.03133 | 0.00199 | 8.00E-56 | -0.01895 | 0.002025 | 8.20E-21 |
| rs2292238 | A | C | 0.001211 | 0.00197 | 0.54 | 0.012178 | 0.002005 | 1.30E-09 |
| rs770082 | G | A | -0.00423 | 0.00196 | 0.031 | -0.01336 | 0.001995 | 2.10E-11 |
| rs2364232 | A | C | 0.012294 | 0.002213 | 2.80E-08 | 0.001021 | 0.002252 | 0.65 |
| rs11111647 | G | A | 0.013084 | 0.002394 | 4.60E-08 | 0.00548 | 0.002437 | 0.025 |

| | | | | | | | | |
|---|---|---|---|---|---|---|---|---|
| rs10849900 | T | C | 0.004161 | 0.002093 | 0.047 | 0.012032 | 0.00213 | 1.60E-08 |
| rs7305424 | A | T | -0.01206 | 0.002048 | 3.90E-09 | -0.00379 | 0.002084 | 0.069 |
| rs3803005 | T | C | -0.00278 | 0.002181 | 0.2 | -0.01681 | 0.00222 | 3.60E-14 |
| rs35202265 | C | T | -0.01159 | 0.002003 | 7.10E-09 | -0.00797 | 0.002038 | 9.10E-05 |
| rs9551428 | C | T | 0.015294 | 0.002001 | 2.10E-14 | 0.006264 | 0.002035 | 0.0021 |
| rs1336486 | T | G | -0.01472 | 0.002062 | 9.30E-13 | -0.00646 | 0.002098 | 0.0021 |
| rs9317002 | C | A | -0.01384 | 0.00195 | 1.20E-12 | -0.00876 | 0.001984 | 1.00E-05 |
| rs9540493 | A | G | 0.010968 | 0.001954 | 2.00E-08 | 0.008035 | 0.001988 | 5.30E-05 |
| rs1576655 | A | C | -0.01171 | 0.002012 | 5.80E-09 | -0.01222 | 0.002047 | 2.40E-09 |
| rs7331420 | G | A | -0.00024 | 0.002151 | 0.91 | 0.01195 | 0.002189 | 4.80E-08 |
| rs9522180 | C | T | 0.002222 | 0.001948 | 0.25 | 0.010849 | 0.001982 | 4.40E-08 |
| rs61980008 | G | A | -0.03658 | 0.005012 | 2.90E-13 | -0.02221 | 0.005099 | 1.30E-05 |
| rs8022132 | A | T | -0.00402 | 0.00211 | 0.057 | -0.01461 | 0.002147 | 1.00E-11 |
| rs6575340 | G | A | -0.00492 | 0.002016 | 0.015 | -0.01459 | 0.002051 | 1.10E-12 |
| rs12891477 | C | T | -0.00048 | 0.002016 | 0.81 | -0.01308 | 0.002051 | 1.80E-10 |
| rs1466276 | C | G | 0.004675 | 0.001947 | 0.016 | 0.011134 | 0.001981 | 1.90E-08 |
| rs8030456 | C | T | 0.023354 | 0.002306 | 4.20E-24 | 0.019398 | 0.002346 | 1.40E-16 |
| rs67962220 | T | G | -0.00814 | 0.002582 | 0.0016 | -0.01507 | 0.002627 | 9.80E-09 |
| rs4932430 | A | C | 0.011803 | 0.001953 | 1.50E-09 | 0.004887 | 0.001987 | 0.014 |
| rs939624 | C | T | -0.0003 | 0.001943 | 0.88 | -0.01231 | 0.001977 | 4.70E-10 |
| rs2531991 | G | A | -0.02012 | 0.002233 | 2.10E-19 | -0.01323 | 0.002272 | 5.90E-09 |
| rs148965598 | A | G | 0.032288 | 0.002826 | 3.10E-30 | 0.018625 | 0.002876 | 9.40E-11 |
| rs7189927 | T | C | 0.016043 | 0.002021 | 2.10E-15 | 0.017294 | 0.002056 | 4.10E-17 |
| rs34898535 | C | T | 0.006972 | 0.001995 | 0.00047 | 0.01922 | 0.00203 | 2.80E-21 |
| rs1421085 | T | C | -0.04852 | 0.001974 | 1.90E-133 | -0.04548 | 0.002008 | 1.50E-113 |
| rs34229857 | C | T | -0.04469 | 0.005779 | 1.00E-14 | -0.01802 | 0.005881 | 0.0022 |
| rs11642090 | T | C | -0.01336 | 0.002015 | 3.40E-11 | -0.00949 | 0.002051 | 3.70E-06 |
| rs4790292 | C | A | 0.009329 | 0.002689 | 0.00052 | 0.017458 | 0.002737 | 1.80E-10 |
| rs1914889 | G | A | -0.00016 | 0.00195 | 0.94 | 0.0122 | 0.001984 | 7.80E-10 |
| rs2306593 | C | T | 0.004413 | 0.001939 | 0.023 | 0.011415 | 0.001973 | 7.20E-09 |
| rs11079849 | C | T | -0.0021 | 0.002063 | 0.31 | 0.012664 | 0.002099 | 1.60E-09 |
| rs77706698 | G | A | -0.00342 | 0.002869 | 0.23 | -0.01716 | 0.002919 | 4.20E-09 |
| rs11150745 | A | G | 0.012046 | 0.002083 | 7.30E-09 | 0.014302 | 0.00212 | 1.50E-11 |
| rs1013737 | G | C | -0.01086 | 0.001939 | 2.20E-08 | -0.00575 | 0.001973 | 0.0036 |
| rs11660335 | T | C | 0.003653 | 0.002473 | 0.14 | 0.015916 | 0.002516 | 2.50E-10 |
| rs7239114 | G | A | -0.01414 | 0.001957 | 5.00E-13 | -0.00876 | 0.001991 | 1.10E-05 |
| rs66922415 | A | G | -0.03538 | 0.002285 | 4.60E-54 | -0.03858 | 0.002325 | 7.40E-62 |
| rs17066856 | T | C | 0.028217 | 0.003379 | 6.80E-17 | 0.028109 | 0.003438 | 2.90E-16 |
| rs9962947 | C | T | -0.00712 | 0.002014 | 0.00041 | -0.0112 | 0.002049 | 4.60E-08 |
| rs12974664 | G | A | -0.00127 | 0.00195 | 0.51 | -0.01228 | 0.001985 | 6.00E-10 |
| rs350832 | G | A | 0.000372 | 0.002319 | 0.87 | -0.0137 | 0.00236 | 6.50E-09 |
| rs12986231 | T | C | 0.007625 | 0.002207 | 0.00055 | 0.014685 | 0.002246 | 6.20E-11 |
| rs10404726 | C | T | 0.008036 | 0.001942 | 3.50E-05 | 0.013847 | 0.001976 | 2.50E-12 |
| rs56212061 | C | T | 0.012033 | 0.002712 | 9.10E-06 | 0.01517 | 0.00276 | 3.90E-08 |
| rs111640872 | G | C | -0.00536 | 0.002064 | 0.0094 | -0.01264 | 0.0021 | 1.80E-09 |

| | | | | | | | | |
|---|---|---|---|---|---|---|---|---|
| rs3810304 | A | G | 0.014315 | 0.002319 | 6.70E-10 | 0.00019 | 0.00236 | 0.94 |
| rs429358 | T | C | -0.00248 | 0.002682 | 0.35 | 0.017504 | 0.002729 | 1.40E-10 |
| rs12971645 | G | A | 0.000424 | 0.002177 | 0.85 | 0.012137 | 0.002215 | 4.30E-08 |
| rs1800437 | G | C | 0.011905 | 0.002442 | 1.10E-06 | 0.023047 | 0.002485 | 1.80E-20 |
| rs994308 | C | T | -0.01235 | 0.001973 | 3.80E-10 | -0.0093 | 0.002008 | 3.60E-06 |
| rs7268466 | C | T | -0.02006 | 0.002776 | 4.90E-13 | -0.00836 | 0.002825 | 0.0031 |
| rs8124896 | T | C | -0.00599 | 0.003209 | 0.062 | -0.02014 | 0.003266 | 6.90E-10 |
| rs116948922 | C | T | 0.002828 | 0.005517 | 0.61 | 0.03396 | 0.005616 | 1.50E-09 |
| rs34966255 | T | C | 0.004118 | 0.002464 | 0.095 | 0.016399 | 0.002508 | 6.20E-11 |
| rs66469746 | C | A | 0.014331 | 0.002149 | 2.60E-11 | 0.002351 | 0.002187 | 0.28 |
| rs4817973 | G | A | 0.013563 | 0.002019 | 1.80E-11 | 0.008633 | 0.002055 | 2.70E-05 |
| rs915814 | G | A | 0.001038 | 0.002272 | 0.65 | 0.013082 | 0.002313 | 1.50E-08 |
| rs400997 | T | A | -0.00231 | 0.001962 | 0.24 | -0.0143 | 0.001998 | 8.00E-13 |
| rs6001872 | A | G | 0.013892 | 0.002033 | 8.30E-12 | 0.012798 | 0.00207 | 6.30E-10 |
| rs738140 | A | G | 0.00113 | 0.002094 | 0.59 | 0.011998 | 0.002132 | 1.80E-08 |
| Abbreviations: Chr Chromosome; EA effect allele; OA other allele; se standard error | | | | | | | | |

| **Supplementary table 6: $F_{early\ life\ body\ size}$, $F_{adult\ body\ size}$ and $Q_a$ in the multivariable MR between early, adult body size and colorectal cancer** | | | | | | | |
|---|---|---|---|---|---|---|---|
| $F_{early\ life\ body\ size}$ and $F_{adult\ body\ size}$ | | | Cancer | $Q_a$ | | | |
| | $F_{early\ life\ body\ size}$ | $F_{adult\ body\ size}$ | | $Q_a$ | df | Min $Q_a$ | $Pvalue_{Q_a}$ |
| Men & women | | | | | | | |
| | 13 | 15 | Colorectal | 558 | 320 | 363 | $3\times10^{-15}$ |
| | | | Colon | 469 | 320 | 363 | $9\times10^{-8}$ |
| | | | Proximal | 387 | 320 | 363 | $6\times10^{-3}$ |
| | | | Distal | 479 | 320 | 363 | $2\times10^{-8}$ |
| | | | Rectal | 459 | 320 | 363 | $5\times10^{-7}$ |
| Men | | | | | | | |
| | 8 | 10 | Colorectal | 194 | 127 | 154 | $1\times10^{-4}$ |
| | | | Colon | 164 | 127 | 154 | 0.01 |
| | | | Proximal | 149 | 127 | 154 | 0.09 |
| | | | Distal | 150 | 127 | 154 | 0.08 |
| | | | Rectal | 142 | 127 | 154 | 0.17 |
| Women | | | | | | | |
| | 12 | 13 | Colorectal | 290 | 184 | 217 | $9\times10^{-7}$ |
| | | | Colon | 263 | 184 | 217 | $1\times10^{-4}$ |
| | | | Proximal | 238 | 184 | 217 | $5\times10^{-3}$ |
| | | | Distal | 240 | 184 | 217 | $3\times10^{-3}$ |
| | | | Rectal | 244 | 184 | 217 | $2\times10^{-3}$ |
| $F_{early\ life\ body\ size}$ and $F_{adult\ body\ size}$ measure weak-instrument bias for the two exposures in a two-sample multivariable MR framework. Qa is a measure of heterogeneity. | | | | | | | |

**Supplementary table 7: Univariable Mendelian randomization estimates between early and adult body size and colorectal cancer risk**

| | Early life body size | | | | Adult body size | | | |
|---|---|---|---|---|---|---|---|---|
| | **Estimates (OR)** | **95% CI** | **P-value** | **P-value for pleiotropy† or heterogeneity‡** | **Estimates (OR)** | **95% CI** | **P-value** | **P-value for pleiotropy† or heterogeneity‡** |
| **Men & women** | | | | | | | | |
| **Colorectal Cancer** | | | | | | | | |
| Inverse-variance weighted | 1.12 | 0.98, 1.27 | 0.09 | $5\times10^{-13}$ | 1.30 | 1.17, 1.45 | $1\times10^{-6}$ | $5\times10^{-10}$ |
| MR-Egger | 1.46 | 1.09, 1.93 | 0.01 | 0.04 | 1.32 | 0.97, 1.82 | 0.08 | 0.90 |
| Weighted median | 1.06 | 0.89, 1.27 | 0.51 | | 1.23 | 1.07, 1.43 | 0.03 | |
| **Colon Cancer** | | | | | | | | |
| Inverse-variance weighted | 1.16 | 1.00, 1.35 | 0.05 | $4\times10^{-9}$ | 1.32 | 1.19, 1.51 | $3\times10^{-6}$ | $5\times10^{-6}$ |
| MR-Egger | 1.49 | 1.08, 2.08 | 0.02 | 0.09 | 1.43 | 1.00, 2.05 | 0.05 | 0.66 |
| Weighted median | 1.21 | 0.97, 1.49 | 0.08 | | 1.31 | 1.09, 1.58 | 0.004 | |
| **Proximal Colon Cancer** | | | | | | | | |
| Inverse-variance weighted | 1.11 | 0.93, 1.32 | 0.25 | $8\times10^{-5}$ | 1.42 | 1.22, 1.63 | $2\times10^{-6}$ | $4\times10^{-3}$ |
| MR-Egger | 1.60 | 1.09, 2.34 | 0.01 | 0.03 | 1.55 | 1.01, 2.36 | 0.04 | 0.67 |
| Weighted median | 1.12 | 0.86, 1.45 | 0.41 | | 1.27 | 1.00, 1.60 | 0.05 | |
| **Distal Colon Cancer** | | | | | | | | |
| Inverse-variance weighted | 1.25 | 1.04, 1.51 | 0.02 | $2\times10^{-5}$ | 1.23 | 1.06, 1.43 | 0.008 | $4\times10^{-5}$ |
| MR-Egger | 1.42 | 0.95, 2.12 | 0.09 | 0.49 | 1.39 | 0.88, 2.18 | 0.16 | 0.60 |
| Weighted median | 1.27 | 0.97, 1.67 | 0.09 | | 1.19 | 0.92, 1.54 | 0.18 | |
| **Rectal Cancer** | | | | | | | | |
| Inverse-variance weighted | 1.14 | 0.93, 1.38 | 0.19 | $3\times10^{-8}$ | 1.27 | 1.09, 1.46 | 0.002 | $8\times10^{-3}$ |
| MR-Egger | 1.36 | 0.90, 2.08 | 0.15 | 0.33 | 1.19 | 0.76, 1.84 | 0.44 | 0.77 |
| Weighted median | 1.23 | 0.94, 1.62 | 0.13 | | 1.32 | 1.06, 1.67 | 0.02 | |
| **Men** | | | | | | | | |
| **Colorectal Cancer** | | | | | | | | |
| Inverse-variance weighted | 0.96 | 0.73, 1.26 | 0.78 | $1\times10^{-3}$ | 1.28 | 1.05, 1.58 | 0.01 | 0.003 |
| MR-Egger | 1.07 | 0.58, 1.99 | 0.82 | 0.70 | 1.08 | 0.58, 2.03 | 0.80 | 0.57 |
| Weighted median | 0.97 | 0.69, 1.35 | 0.84 | | 1.14 | 0.84, 1.52 | 0.40 | |
| **Colon Cancer** | | | | | | | | |
| Inverse-variance weighted | 1.08 | 0.80, 1.45 | 0.61 | 0.05 | 1.32 | 1.07, 1.65 | 0.01 | 0.06 |
| MR-Egger | 1.09 | 0.55, 2.16 | 0.80 | 0.97 | 1.08 | 0.53, 2.25 | 0.82 | 0.57 |
| Weighted median | 0.99 | 0.66, 1.49 | 0.98 | | 1.07 | 0.75, 1.54 | 0.7 | |
| **Proximal Colon Cancer** | | | | | | | | |
| Inverse-variance weighted | 1.31 | 0.94, 1.82 | 0.11 | 0.22 | 1.26 | 0.95, 1.65 | 0.11 | 0.64 |
| MR-Egger | 1.28 | 0.57, 2.89 | 0.55 | 0.96 | 1.22 | 0.53, 2.83 | 0.64 | 0.95 |
| Weighted median | 1.21 | 0.72, 2.03 | 0.48 | | 1.12 | 0.69, 1.80 | 0.65 | |
| **Distal Colon Cancer** | | | | | | | | |
| Inverse-variance weighted | 0.92 | 0.67, 1.28 | 0.64 | 0.14 | 1.43 | 1.09, 1.88 | 0.008 | 0.1 |

| | | | | | | | | |
|---|---|---|---|---|---|---|---|---|
| MR-Egger | 0.95 | 0.42, 2.16 | 0.91 | 0.93 | 0.98 | 0.40, 2.39 | 0.97 | 0.38 |
| Weighted median | 0.86 | 0.53, 1.42 | 0.56 | | 1.21 | 0.79, 1.86 | 0.40 | |
| **Rectal Cancer** | | | | | | | | |
| Inverse-variance weighted | 0.82 | 0.60, 1.12 | 0.21 | 0.66 | 1.21 | 0.93, 1.57 | 0.15 | 0.16 |
| MR-Egger | 1.14 | 0.55, 2.34 | 0.72 | 0.32 | 1.04 | 0.45, 2.44 | 0.92 | 0.72 |
| Weighted median | 0.94 | 0.57, 1.54 | 0.81 | | 1.06 | 0.68, 1.67 | 0.81 | |
| **Women** | | | | | | | | |
| **Colorectal Cancer** | | | | | | | | |
| Inverse-variance weighted | 1.20 | 0.97, 1.48 | 0.09 | $7\times10^{-10}$ | 1.22 | 1.03, 1.43 | 0.01 | $1.9\times10^{-3}$ |
| MR-Egger | 1.84 | 1.16, 2.92 | 0.009 | 0.04 | 1.30 | 0.79, 2.14 | 0.29 | 0.81 |
| Weighted median | 1.35 | 1.06, 1.73 | 0.02 | | 1.17 | 0.92, 1.49 | 0.19 | |
| **Colon Cancer** | | | | | | | | |
| Inverse-variance weighted | 1.20 | 0.95, 1.51 | 0.13 | $8\times10^{-7}$ | 1.22 | 1.01, 1.48 | 0.04 | 0.01 |
| MR-Egger | 1.70 | 1.02, 2.83 | 0.04 | 0.13 | 1.58 | 0.90, 2.77 | 0.11 | 0.33 |
| Weighted median | 1.19 | 0.89, 1.58 | 0.26 | | 1.22 | 0.91, 1.63 | 0.18 | |
| **Proximal Colon Cancer** | | | | | | | | |
| Inverse-variance weighted | 1.22 | 0.92, 1.62 | 0.17 | $2\times10^{-6}$ | 1.39 | 1.09, 1.75 | 0.006 | $8\times10^{-3}$ |
| MR-Egger | 1.70 | 0.91, 3.19 | 0.10 | 0.24 | 1.58 | 0.79, 3.19 | 0.20 | 0.69 |
| Weighted median | 1.04 | 0.73, 1.48 | 0.83 | | 1.21 | 0.85, 1.70 | 0.30 | |
| **Distal Colon Cancer** | | | | | | | | |
| Inverse-variance weighted | 1.14 | 0.85, 1.52 | 0.39 | $6\times10^{-4}$ | 1.06 | 0.84, 1.34 | 0.64 | 0.62 |
| MR-Egger | 1.90 | 1.00, 3.60 | 0.05 | 0.08 | 1.63 | 0.79, 3.35 | 0.18 | 0.21 |
| Weighted median | 1.48 | 0.99, 2.20 | 0.05 | | 1.22 | 0.84, 1.79 | 0.29 | |
| **Rectal Cancer** | | | | | | | | |
| Inverse-variance weighted | 1.17 | 0.86, 1.58 | 0.32 | $2\times10^{-4}$ | 1.31 | 1.00, 1.70 | 0.05 | 0.02 |
| MR-Egger | 2.01 | 1.03, 3.97 | 0.04 | 0.08 | 1.04 | 0.47, 2.29 | 0.92 | 0.56 |
| Weighted median | 1.01 | 0.67, 1.52 | 0.96 | | 1.15 | 0.77, 1.70 | 0.49 | |

Abbreviations: CI confidence intervals, MR Mendelian randomization, OR odds ratio

† P-value for pleiotropy based on MR-Egger intercept

‡ P-value for heterogeneity based on Q statistic

| Supplementary table 8: Multivariable MR Egger analysis to assess the effect of both predicted early life and adult body size on colorectal cancer | | | | | | | |
|---|---|---|---|---|---|---|---|
| | Multivariable MR Egger analysis with positive early life body size effect | | | | Multivariable MR Egger analysis with positive adult life body size effect | | |
| | Timepoint | OR | 95% CI | Pvalue | OR | 95% CI | Pvalue |
| Men & women | | | | | | | |
| Colorectal | Early life | 1.08 | 0.85, 1.39 | 0.5 | 0.97 | 0.77, 1.22 | 0.80 |
| | Adult | 1.42 | 1.13, 1.79 | 0.003 | 1.42 | 1.02, 1.97 | 0.04 |
| | Intercept | | | 0.02 | | | 0.42 |
| Colon | Early life | 1.13 | 0.86, 1.48 | 0.38 | 0.97 | 0.76, 1.25 | 0.83 |
| | Adult life | 1.52 | 1.19, 1.95 | 0.001 | 1.58 | 1.09, 2.29 | 0.01 |
| | Intercept | | | 0.005 | | | 0.21 |
| Proximal colon | Early life | 0.98 | 0.72, 1.34 | 0.91 | 0.82 | 0.61, 1.09 | 0.17 |
| | Adult | 1.88 | 1.40, 2.48 | $1.45 \times 10^{-5}$ | 1.73 | 1.14, 2.64 | 0.01 |
| | Intercept | | | 0.003 | | | 0.57 |
| Distal colon | Early life | 1.39 | 0.97, 1.99 | 0.07 | 1.27 | 0.91, 1.77 | 0.16 |
| | Adult | 1.12 | 0.80, 1.55 | 0.52 | 1.32 | 0.81, 2.14 | 0.26 |
| | Intercept | | | 0.19 | | | 0.18 |
| Rectal | Early life | 1.14 | 0.80, 1.63 | 0.46 | 1.05 | 0.76, 1.45 | 0.78 |
| | Adult | 1.22 | 0.89, 1.68 | 0.23 | 1.25 | 0.77, 1.99 | 0.37 |
| | Intercept | | | 0.21 | | | 0.61 |
| Men | | | | | | | |
| Colorectal | Early life | 0.79 | 0.51, 1.23 | 0.30 | 0.74 | 0.49, 1.12 | 0.15 |
| | Adult | 1.60 | 1.08, 2.39 | 0.02 | 1.69 | 0.88, 2.94 | 0.13 |
| | Intercept | | | 0.51 | | | 0.84 |
| Colon | Early life | 0.90 | 0.54, 1.49 | 0.69 | 0.81 | 0.51, 1.28 | 0.37 |
| | Adult | 1.65 | 1.06, 2.56 | 0.03 | 1.22 | 0.61, 2.44 | 0.57 |
| | Intercept | | | 0.34 | | | 0.42 |
| Proximal colon | Early life | 1.15 | 0.63, 2.14 | 0.65 | 0.94 | 0.53, 1.65 | 0.83 |
| | Adult | 1.57 | 0.91, 2.69 | 0.10 | 1.52 | 0.65, 3.53 | 0.33 |
| | Intercept | | | 0.11 | | | 0.72 |
| Distal colon | Early life | 0.73 | 0.39, 1.34 | 0.30 | 0.70 | 0.40, 1.21 | 0.20 |
| | Adult | 1.79 | 1.05, 3.03 | 0.03 | 1.01 | 0.44, 2.29 | 0.98 |
| | Intercept | | | 0.86 | | | 0.1 |
| Rectal | Early life | 0.80 | 0.45, 1.42 | 0.45 | 0.74 | 0.44, 1.26 | 0.27 |
| | Adult | 1.43 | 0.86, 2.36 | 0.16 | 1.80 | 0.83, 3.90 | 0.14 |
| | Intercept | | | 0.50 | | | 0.35 |
| Women | | | | | | | |
| Colorectal | Early life | 1.14 | 0.83, 1.57 | 0.43 | 1.12 | 0.84, 1.49 | 0.46 |
| | Adult | 1.11 | 0.81, 1.52 | 0.52 | 1.42 | 0.90, 2.20 | 0.13 |
| | Intercept | | | 0.54 | | | 0.10 |
| Colon | Early life | 1.06 | 0.73, 1.52 | 0.76 | 1.01 | 0.73, 1.40 | 0.94 |
| | Adult | 1.28 | 0.90, 1.82 | 0.18 | 1.62 | 0.98, 2.69 | 0.06 |
| | Intercept | | | 0.39 | | | 0.12 |
| Proximal colon | Early life | 0.86 | 0.57, 1.31 | 0.48 | 0.78 | 0.53, 1.14 | 0.20 |
| | Adult | 1.79 | 1.19, 2.72 | 0.006 | 1.68 | 0.93, 3.03 | 0.08 |
| | Intercept | | | 0.29 | | | 0.89 |
| Distal colon | Early life | 1.38 | 0.86, 2.20 | 0.18 | 1.40 | 0.92, 2.14 | 0.11 |
| | Adult | 0.85 | 0.54, 1.35 | 0.5 | 1.58 | 0.84, 3.00 | 0.16 |
| | Intercept | | | 0.73 | | | 0.009 |

| | | | | | | | |
|---|---|---|---|---|---|---|---|
| Rectal | Early life<br>Adult<br>Intercept | 0.90<br>1.15 | 0.55, 1.48<br>0.70, 1.88 | 0.69<br>0.57<br>0.56 | 0.98<br>1.54 | 0.63, 1.54<br>0.77, 3.06 | 0.94<br>0.23<br>0.41 |
| Abbreviations: CI confidence interval; MR Mendelian randomization; OR odds ratio | | | | | | | |
